# Worldwide antibiotic resistance dynamics: how different is it from one drug-bug pair to another?

**DOI:** 10.1101/2022.02.09.22270726

**Authors:** Eve Rahbe, Laurence Watier, Didier Guillemot, Philippe Glaser, Lulla Opatowski

**Author notes:** Contributed equally (Philippe Glaser, Lulla Opatowski).

## Abstract

**Background:** Antibiotic resistance (ABR) is a major concern for global health. However, factors driving its emergence and dissemination are not fully understood. Identification of such factors is crucial to explain heterogeneity in ABR rates observed across space, time and species and antibiotics.

**Methods:** We analyzed count data of clinical isolates from 51 countries over 2006-2019 for thirteen drug-bug pairs from the ATLAS surveillance system. We characterized ABR spatial and temporal patterns and used a mixed-effect negative binomial model, accounting for country-year dependences with random effects (RE), to investigate associations with potential drivers including antibiotic sales, economic and health indicators, meteorological data, population density and tourism.

**Findings:** ABR patterns were strongly country and drug-bug pair dependent. In 2019, median ABR rates ranged from 6×3% (interquartile range (IQR): 19×7%) for carbapenem-resistant (CR) *Klebsiella pneumoniae* to 80×7% (IQR: 41×8%) for fluoroquinolone-resistant (FR) *Acinetobacter baumannii*, with heterogeneity across countries. Over 2006-2019, carbapenem resistance was on the rise in >60% of investigated countries, while no global trend was observed for other resistances. Multivariable analyses identified significant associations of ABR with country-level selecting antibiotic sales, but only in FR-*Escherichia coli*, FR-*Pseudomonas aeruginosa* and CR-*A. baumannii;* with temperature in investigated Enterobacterales but not in other drug-bug pairs; and with the health system quality for all drug-bug pairs except *Enterococci* and *Streptococcus pneumoniae* pairs. Despite wide consideration of possible explanatory variables, drug-bug pairs ABR rates still showed unexplained spatial RE variance.

**Interpretation:** Our findings reflect the diversity of mechanisms driving global antibiotic resistance across pathogens and stress the need for tailored interventions to tackle bacterial resistance.

**Funding:** Independent research Pfizer Global Medical Grant; ANR Labex IBEID (ANR-10-LABX-62)

## Introduction

Antibiotic resistance (ABR) in clinically relevant bacteria is a major public health threat.^1^ Latest estimations show that 1×27 millions deaths (95% uncertainty interval: 0×911-1×71) were attributable to ABR in 2019 worldwide^2^.^3^ Although ABR is a global problem observed in all countries, resistance patterns show important geographical differences and greatly vary depending on the bacterial species and antibiotic resistance considered.^3^ Factors explaining these variations are not fully understood.

Recent studies investigating determinants of country-level ABR suggested that in addition to antibiotic consumption, socio-economic factors are correlated to ABR rates.^4,5^ Containing ABR spread, rather than only limiting antibiotic consumption, seems crucial to reduce its global burden. But the limited knowledge about mechanistic factors driving ABR spread impedes the setup of efficient control measures. Behavioral factors could play a role in ABR transmission, such as tourism.^6^ In addition, climate warming has been suggested to be associated with ABR dissemination and maintenance, hypothesizing that higher temperatures promote bacterial growth and more frequent horizontal gene transfer. ^7–9^ More complex mechanisms involving humans and their environment might thus be at stake, setting ABR in a One Health perspective.^10^

As pathogens exhibit different resistance mechanisms, thrive in different ecological niches, and can be associated with different epidemiological sources (the environment, hospitals, or the community), understanding ABR dynamics requires studying it at the bacterial species level. Global ABR determinants have been previously analyzed using data from one or two selected bacteria or aggregated resistance indices, but no study compared worldwide ABR drivers between species.^4,5,11^

Data collected by surveillance programs are needed to analyze and identify determinants of ABR dynamics globally in different pathogens. Specifically, the Antimicrobial Testing Leadership and Surveillance (ATLAS) system is a surveillance program which monitors antibiotics efficacy against a wide range of bacteria.^12^ ATLAS gathers clinical isolates from different bacterial infections using a systematic protocol and collects longitudinal data over all continents, including lower-, middle- and high-income countries. It uses a standardized and centralized procedure of determining minimal inhibitory concentrations (MICs) across multiple species and antibiotics combinations (“drug-bug pairs”).

In this study, we analyze spatial-temporal dynamics of critical drug-bug pairs from worldwide infection ABR surveillance data collected through ATLAS and identify and compare key mechanistic factors - antibiotic sales, meteorological and climatic variables, wealth and health metrics, population density, tourism - associated with such dynamics.

## Research in context

### Evidence before this study

We performed a PubMed search for articles in English published between 2000 and 2021. Search terms for titles and abstracts were: (“global” OR “worldwide” OR “country” OR “country-level”) AND (“antibiotic resistance” OR “antimicrobial resistance” OR “bacterial resistance” OR “drug resistance”) AND (“factors” OR “determinants” OR “association” OR “correlation”). We excluded studies analyzing viruses, parasites, fungi, cancers, cholera, and tuberculosis. The search yielded 100 articles.

After filtering articles referring to ecological or prevalence studies focusing on only one country, one species or one infection source, articles addressing molecular epidemiology, articles predicting country-level ABR rates in space or time, or general reviews discussing ABR, we found five relevant studies. Two articles found a link between climate warming and ABR in European countries.^8,9^ The three other articles studied multiple individual socio-economic and national associated factors, two in Europe,^5,13^ and one in a worldwide dataset.^4^ Most studies considered only resistant Enterobacterales or *Staphylococcus aureus*, or used aggregated resistance indices.^4,13^ Reviews retrieved using the search indicated that international travel and tourism could also play a role in ABR spread.^6^ No article had compared multiple bacterial species and antibiotic resistances using longitudinal data to assess differences in ABR spread factors at the worldwide level.

### Added value of this study

ATLAS represents a unique dataset, useful to compare ABR rates worldwide and question their dynamics. Here, we used longitudinal surveillance data of ABR rates from infection samples for 51 countries, not restricted to Europe, and over 14 years (2006-2019). To our knowledge, this is the first study to analyze ABR surveillance data from a large number (thirteen) of different clinically relevant drug-bug pairs and to characterize the observed heterogeneity in ABR rates across different countries and years. Our statistical analysis approach included both spatial and temporal dimensions of the ABR phenomenon, with country-year dependent outcome variable and co-variables, which were not considered in previous studies. Moreover, the choice of co-variables is not restricted to global indices, but rather detailed metrics representing putative mechanistic drivers of ABR such as meteorological factors (rainfall, relative humidity, temperature, and extreme climatic events such as flooding) or antibiotic sales described at the antibiotic class level.

### Implications of all the available evidence

Analysis of the ATLAS data showed that ABR rates are highly dependent on the country and most importantly on the drug-bug pair considered. Results from our statistical analysis suggested that factors associated with ABR rates were different across drug-bug pairs but more similar within a bacterial species, reflecting different underlying ecological behaviors. Variance between countries was only explained by the proposed factors for *Escherichia coli* where antibiotic sales, temperature, extreme climatic events, and the health system quality could explain most ABR rates differences. For other drug-bug pairs, strong spatial variance remained unexplained. Overall, results from this study suggest that ABR should be considered as a plural problem whose control should be tailored regarding the country or the drug-bug pair under consideration.

## Methods

### Antibiotic resistance data

Antibiotic resistance (ABR) data were obtained from the ATLAS database^12^. ATLAS is a surveillance system led by Pfizer, collecting bacterial isolates from hospitalized patients within a worldwide hospital network. ATLAS reports minimal inhibitory concentrations (MICs) in such clinical isolates tested against various antibiotics, and associated information on country, year, hospital ward, source of infection and bacterial species. Data represent infection isolates, with the bacteria being the putative cause of hospitalization. Isolates came from five distinct bacterial infections, associated with five main sources: blood, sputum, urine, abscess and wound. ATLAS sampling protocol is further detailed in appendix p 3.

Count data of non-resistant and resistant isolates from 51 countries (appendix p 4) over 14 years (2006-2019) were extracted. Thirteen clinically relevant antibiotic resistance-bacterium pairs (called “drug-bug pairs”) were analyzed: fluoroquinolone-resistant *Escherichia coli* (FR-*Ec*), aminopenicillin-resistant *E. coli* (APR-*Ec*), third generation cephalosporin-resistant *E. coli* (3GCR-*Ec*), fluoroquinolone-resistant *Klebsiella pneumoniae* (FR-*Kp*), 3GC-resistant *K. pneumoniae* (3GCR-*Kp*), carbapenem-resistant *K. pneumoniae* (CR-*Kp*), fluoroquinolone-resistant *Pseudomonas aeruginosa* (FR-*Pa*), carbapenem-resistant *P. aeruginosa* (CR-*Pa*), fluoroquinolone-resistant *Acinetobacter baumannii* (FR-*Ab*), carbapenem-resistant *A. baumannii* (CR-*Ab*), vancomycin-resistant *Enterococcus faecalis* and *Enterococcus faecium* (VR-*E*), penicillin-non-susceptible *Streptococcus pneumoniae* (PR-*Sp*) and macrolide-resistant *S. pneumoniae* (MLR-*Sp*). MIC values were reported in the ATLAS database and isolates were categorized into Susceptible (S), Intermediate (I) or Resistant (R) based on the 2019 v9.0 MIC breakpoints from the *European Committee on Antimicrobial Susceptibility Testing* standards.^14^ An isolate was considered resistant if it was in the R category, except for PR-*Sp* where an isolate was considered resistant if it was in the I or R categories. An isolate was considered resistant to an antibiotic class if it was resistant to at least one of the antibiotics of the class (appendix p 5).

### Co-variables data

Co-variables were selected and tested as explanatory factors in the statistical analysis based on their hypothetical mechanistic impact on the dynamics and selection of ABR. They include antibiotic sales, meteorological and climatic variables, wealth and health metrics, population density, and tourism (appendix pp 6-7).

Antibiotic sales data were obtained from the IQVIA MIDAS database (appendix p 8).^15^ Data from Denmark, Latvia, Lithuania, and the Netherlands were not available through this database, and were obtained from the ESAC-Net database by converting Defined Daily Doses into grams as presented in appendix p 8. We analyzed the five most frequently sold classes of antibiotics in IQVIA MIDAS over all countries and years (broad-spectrum penicillins, cephalosporins, macrolides, quinolones, and trimethoprim/sulfonamides). We also analyzed two other classes, carbapenems and glycopeptides, corresponding to resistances in CR-*Kp*, CR-*Pa* and CR-*Ab* and VR-*E* respectively (appendix p 9). Global sales represented the sum of these seven classes. Average temperature, minimum temperature, average rainfall and average relative humidity per country-year were obtained through the MERRA-2 dataset, using the capital city value for each country.^16^ Extreme climatic events (droughts, flooding or extreme temperatures) faced by the population in 2009 was obtained from the World Bank Data (WBD).^17^ Population size, population density, gross domestic product (GDP) per capita, and international tourism (legal arrivals and departures) data were obtained through the WBD at the country-year level.^17^ As a proxy for national health system qualities, the 2019 Global Health Security index (GHS, [0-100]) was used.^18^

### Statistical analysis

#### Descriptive statistics

ABR rate was defined as the proportion of resistant isolates *Y* over the total number of isolates tested *n* for each drug-bug-country-year observation, from all infection sources: 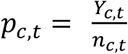 where *c* is the country and *t* the year.

Data filtering was performed to ensure ABR data reliability. Only drug-bug-country-year observations with more than 10 isolates tested were used to compute ABR rates. For each drug-bug pair, country-associated time series with less than 5 years of ABR rates data available over 2006-2019 were excluded. Data imputation was achieved when 3 or less consecutive years exhibited missing ABR rates. Imputation was performed using a moving average estimate with a time window of k = 1 year.

Median ABR rates and interquartile ranges (IQR) across all countries were estimated for the year 2019, the most recent year of the dataset. Temporal trends of ABR rates were defined as the slope of the regression equation from 2006-2019 for each country and pair. Temporal trends were estimated using linear regression minimizing weighted least squares, weights being the inverse variance of observations 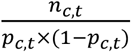. Clustering was performed on these regression slopes, using complete linkage method on Euclidean distances, and p-values associated with slope coefficients’ hypothesis test were reported.

#### Mixed-effect negative binomial model

A mixed-effect model was used to analyze ABR rates and to identify associated factors by accounting for the dependence of the data between countries and across years.^19^ The count of resistant isolates per country-year was considered as the response variable. A negative binomial (NB) response distribution was chosen to account for data overdispersion. The model included spatial and temporal random effects (RE) and fixed effects (FE) with the co-variables data. The model equation used for each drug-bug pair was the following:

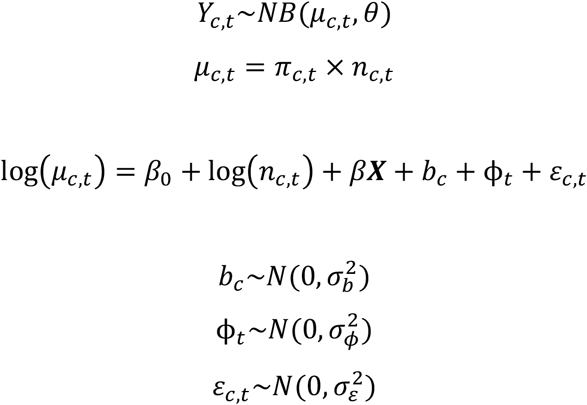

where c is the country, t the year, *Y*_*c,t*_ the number of resistant isolates per country and year; *μ*_*c,t*_ the expected number of resistant isolates; *θ* the dispersion parameter of the NB distribution; *n*_*c,t*_ the number of isolates tested (offset); *π*_*c,t*_ the proportion of resistant isolates (ABR rate); β the fixed effects parameter vector of size *p* (*p* the number of fixed effects); ***X*** the fixed effects model matrix of size C*T\**p* (with C total number of countries and T total number of years); *b*_*c*_ the country-dependent random intercept; ϕ_*t*_ the year-dependent random intercept and *ε*_*c,t*_ the residuals of the model. Random effects and residuals are assumed to be normally distributed.

Intercept-only (called M_null), univariate (M_uni) and multivariable models (M_multi) were evaluated independently for all drug-bug pairs. Co-variables data were standardized (centered with their mean and scaled with their standard deviation). Correlation analysis between each co-variable was performed using the Pearson coefficient (r), and co-variables associated with *r* > |0×7| were excluded from analyses (appendix p 10). For antibiotic sales variables, only global sales and sales of the antibiotic selecting for the specific resistance (called “antibiotic sales of interest”) were tested in the univariate analysis. Co-variables associated with a p-value < 20% in the univariate analysis were included in the multivariable analysis. Backward selection was then performed using hypothesis testing method (z-test). Every co-variable left in the M_multi final multivariable models were statistically significant with a p-value < 5%. Akaike Information Criterion (AIC) values were reported to compare M_null and M_multi models for all drug-bug pairs. The model parameters were estimated using maximum likelihood by Laplace approximation using the *lme4* R package.^20^ To test the difference between variances of RE from M_null and M_multi models, the F-test for equality of two variances was used.

Some drug-bug-country-year observations or countries had to be excluded from the statistical model analysis. For CP-*Kp* and VR-*E*, where ABR rates frequently reached zero values, countries with less than two non-zero data points over the full period were excluded. Observations for which some country-year co-variables were not available were also excluded (appendix p 11).

#### Sensitivity analyses

To assess the potential impact of the filtering threshold related to the minimum number of isolates tested per country-year, we repeated analyses with a filter on 20 isolates instead of 10. To assess whether using ESAC-Net data for Denmark, Latvia, Lithuania, and the Netherlands had any major impacts on the results, we repeated analyses by excluding these countries. To assess whether the infection source might affect ABR rates and associations with co-variables, we carried out analyses on a subset of the data where isolates only came from blood. Finally, we evaluated the potential impact of imputing missing ABR rates on given years on the resulting associations between ABR rates and co-variables by analyzing the dataset without any imputed observations.

All analyses were carried out using R, version 4.0.3.^21^

## Results

In total, 808 744 isolates from all infection sources were analyzed across the thirteen drug-bug pairs. Number of tested isolates for each drug-bug pair and each year, by regions, are described in appendix p 13 and MIC distributions in appendix p 14-15. Isolates distribution and ABR rates by infection sources or hospital wards are described in appendix pp 16-18.

Distributions of antibiotic resistance (ABR) rates across countries in 2019 highlighted strong heterogeneity across drug-bug pairs. For 2019, and study period 2006-2019, numbers of resistant and total isolates by drug-bug pairs are reported in figure 1A. The median ABR ranged from 6×3% (IQR: 19.7%) for CR-*Kp* to 80×7% (IQR: 41.8%) for FR-*Ab* (figure 1B). Interestingly, although CR-*Ab* and APR-*Ec* median rates were similarly high, 72×3% and 68×0% respectively, higher dispersion was found for CR-*Ab* compared to APR-*Ec* (IQR of 50×2% and 18×2%, respectively), revealing different ABR rates distributions across countries (figure 1C and 1D).

**Figure 1.**
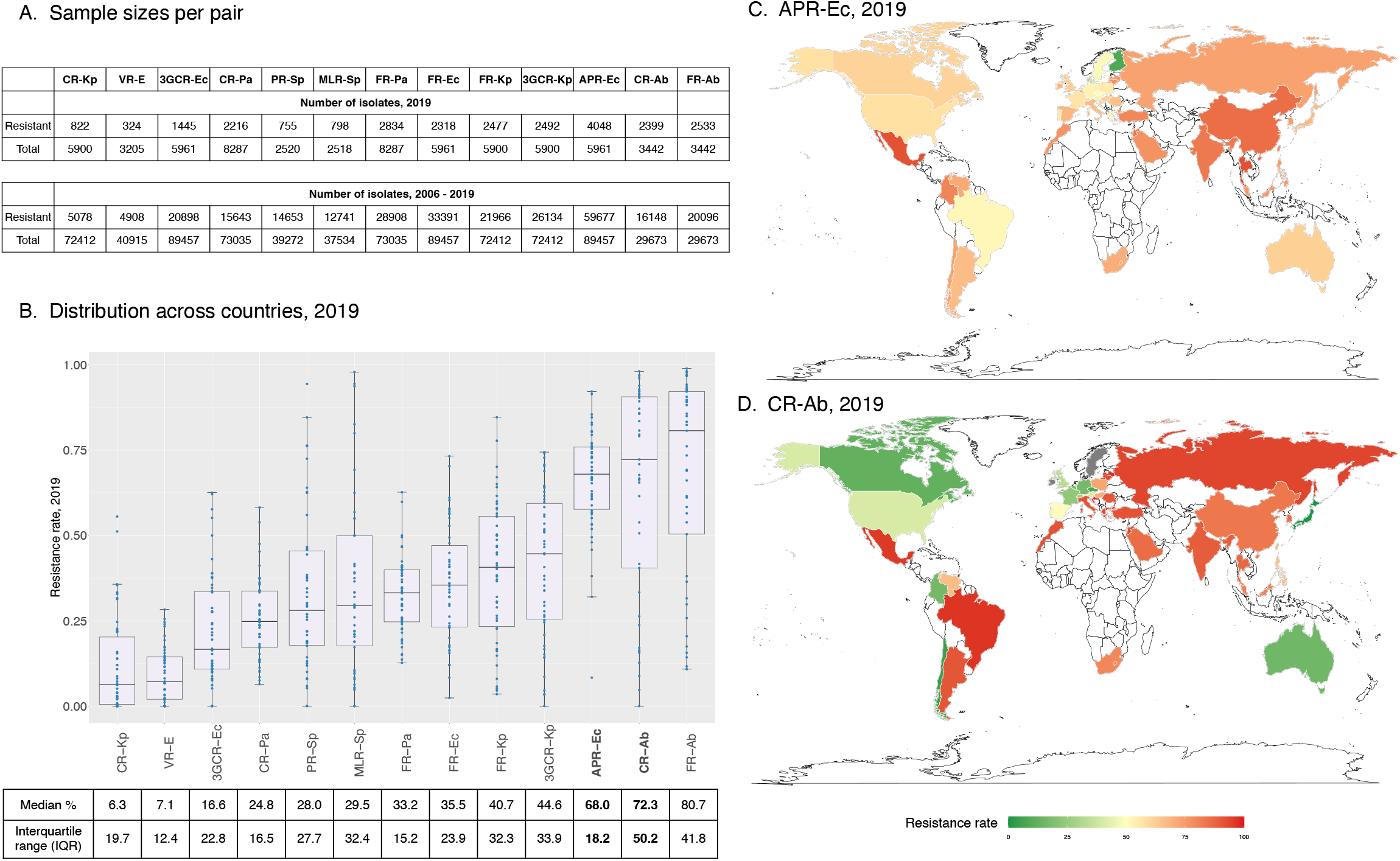
Worldwide antibiotic resistance rates distribution, 2019, ATLAS. ABR rates are reported in proportions or percentages of resistant isolates over total number of tested isolates per country for each drug-bug pair. (A) Sample sizes (number of resistant and total isolates) for each drug-bug pair, for the year 2019 and for study period 2006-2019, are shown. (B) ABR rates across countries for all drug-bug pairs are represented with boxplots, with medians (line) and lower and upper quartiles (box limits). Medians and interquartile ranges are annotated below in percentage (%) for each drug-bug pair. (C-D) Maps of worldwide ABR rates for APR-*Ec* and CR-*Ab* respectively, two pairs exhibiting high median rates in 2019. Resistance rate in % is indicated by colors in the scale below the maps. Grey countries indicate missing value for 2019. White countries are not included in the analysis. (All panels) **CR-Kp**: carbapenem-resistant *K. pneumoniae*; **VR-E**: vancomycin-resistant Enterococci; **3GCR-Ec**: third generation cephalosporin-resistant *E. coli*; **CR-Pa**: carbapenem-resistant *P. aeruginosa*; **PR-Sp**: penicillin-non-susceptible *S. pneumoniae*; **MLR-Sp**: macrolide-resistant *S. pneumoniae;* **FR-Pa**: fluoroquinolone-resistant *P. aeruginosa*; **FR-Ec**: fluoroquinolone-resistant *E. coli*; **FR-Kp**: fluoroquinolone-resistant *K. pneumoniae*; **3GCR-Kp**: third generation cephalosporin-resistant *K. pneumoniae*; **APR-Ec**: aminopenicillin-resistant *E. coli*; **CR-Ab**: carbapenem-resistant *A. baumannii*; **FR-Ab**: fluoroquinolone-resistant *A. baumannii*.

From 2006 to 2019, ABR showed different temporal trends between drug-bug pairs. Trends and sample sizes for each drug-bug-country-year observation are shown in appendix pp 19-32. The heat map in figure 2, displaying linear regression slopes, highlights that ABR temporal trends do not specifically cluster by world’s region, but rather cluster by species or resistances. Noticeably, *E. coli* and *K. pneumoniae* pairs respectively clustered together, suggesting similarities in their temporal trends, but no clear worldwide increasing or decreasing temporal pattern was observed. Two pairs showed global worldwide trends. ABR rates for FR-*Pa* showed decreasing trends over the period, with 24 out of 50 countries exhibiting significant negative slopes. On the contrary, ABR rates trends for CR-*Ab* globally increased, with 24 out of 47 countries exhibiting significant positive slopes. Drug-bug pairs associated with carbapenems resistance generally exhibited increasing trends: 61% of countries had increasing trends for CR-*Kp* (13 significant positive slopes); 76% for CR-*Pa* (14); and 83% for CR-*Ab* (24) (appendix pp 19-32).

**Figure 2.**
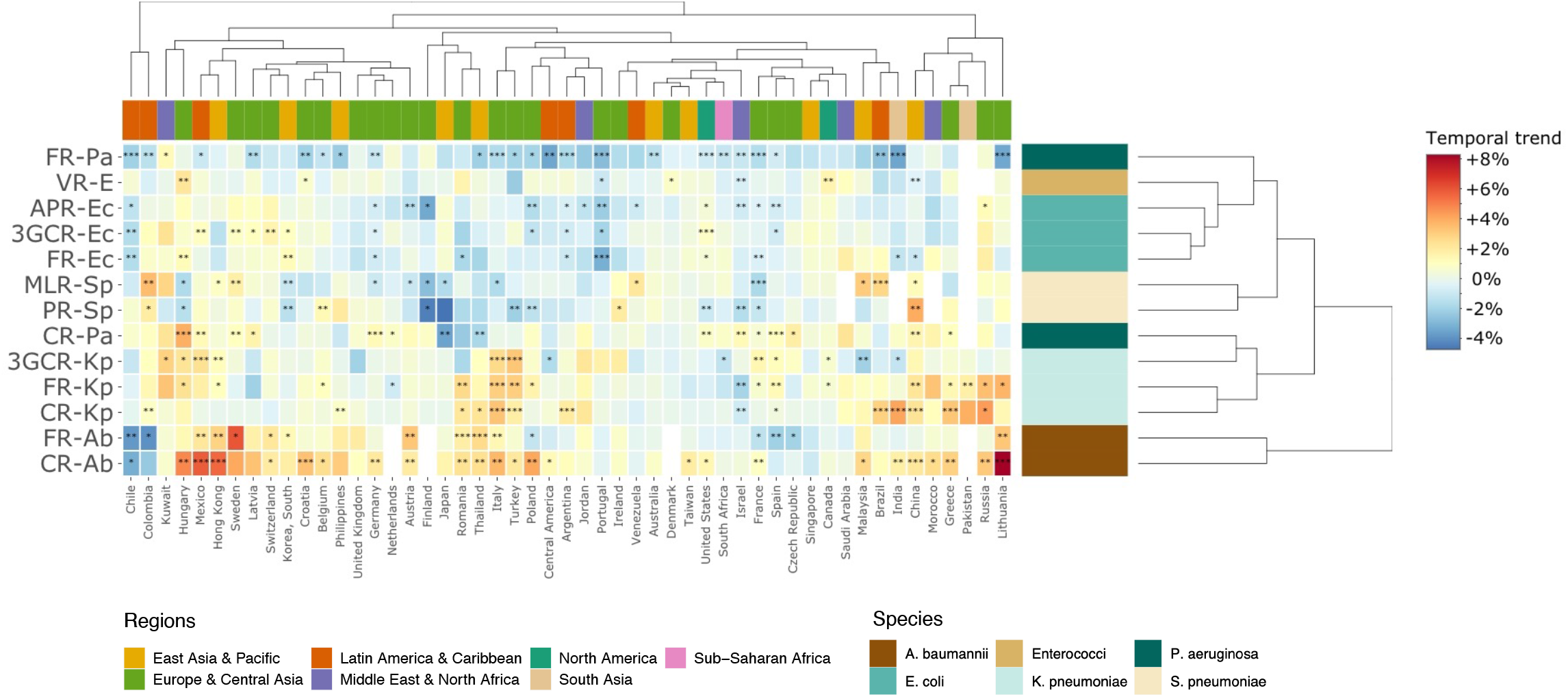
Antibiotic resistance temporal trends (2006-2019) by country and drug-bug pair. The heat map shows temporal trends, defined as slopes estimated from weighted linear temporal regressions (appendix pp 19-32 to see trends and associated p-values). Temporal trends represent the ABR rate change per year for each country and drug-bug pair. Temporal trends are colored from blue (decreasing trend) to red (increasing trend) as in the right legend. Stars represent p-values associated with hypothesis tests on regression slopes (*: p-value < 0.05; **: p-value < 0.01; ***: p-value < 0.001). A white cell represents a missing value for a specific country and drug-bug pair. Clustering was performed on both rows and columns. Possible uncertainty around clustering is investigated in appendix p 33. Countries were categorized into world’s regions based on World Bank indicators and drug-bug pairs were categorized into bacterial species, as indicated in the bottom legend. Drug-bug pairs names: **FR-Pa**: fluoroquinolones-resistant *P. aeruginosa*; **VR-E**: vancomycin-resistant Enterococci; **APR-Ec**: aminopenicillins-resistant *E. coli*; **3GCR-Ec**: third generation cephalosporins-resistant *E. coli*; **FR-Ec**: fluoroquinolones-resistant *E. coli*; **MLR-Sp**: macrolides-resistant *S. pneumoniae*; **PR-Sp**: penicillin-non-susceptible *S. pneumoniae*; **CR-Pa**: carbapenem-resistant *P. aeruginosa*; **3GCR-Kp**: third generation cephalosporins-resistant *K. pneumoniae*; **FR-Kp**: fluoroquinolones-resistant *K. pneumoniae*; **CR-Kp**: carbapenem-resistant *K. pneumoniae*; **FR-Ab**: fluoroquinolones-resistant *A. baumannii*; **CR-Ab**: carbapenem-resistant *A. baumannii*.

From the multivariable analyses, we could assess the association between ABR rates and the proposed co-variables. Results are summarized in table 1, with associated significant co-variables listed in panel A, and models’ fits presented in appendix pp 35-42.

**Table 1.**
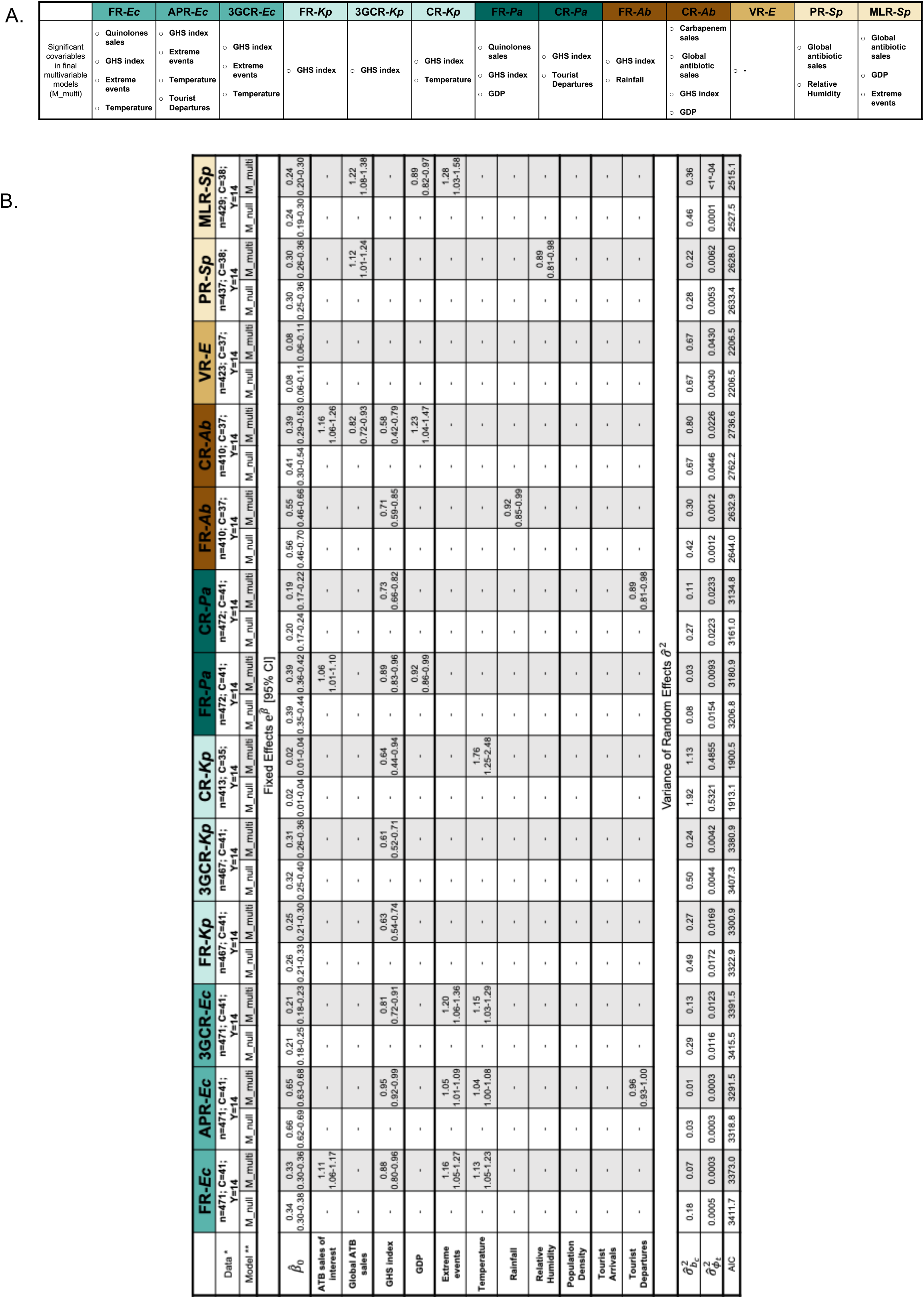
Estimated coefficients from the mixed-effect negative binomial model for each drug-bug pair. (A) Summary of significant co-variables in multivariable (M_multi) models for each drug-bug pair. (B) Results from the mixed-effect negative binomial model. * n represent the number of observations; C, the number of countries and Y, the number of years. ** M_null represent, the intercept-only model and M_multi, the final multivariable model (including only statistically significant variables after backward selection, p-value < 5%). Coefficients for the fixed effects are reported on the exponential form. Spatial and temporal random effects’ variances are reported, respectively 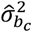 and 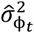. AIC are reported for all models. (All panels) **FR-Ec**: fluoroquinolones-resistant *E. coli*; **APR-Ec**: aminopenicillins-resistant *E. coli*; **3GCR-Ec**: third generation cephalosporins-resistant *E. coli*: **FR-Kp**: fluoroquinolones-resistant *K. pneumoniae*; **3GCR-Kp**: third generation cephalosporins-resistant *K. pneumoniae*; **CR-Kp**: carbapenem-resistant *K. pneumoniae*; **FR-Pa**: fluoroquinolones-resistant *P. aeruginosa*; **CR-Pa**: carbapenem-resistant *P. aeruginosa*; **FR-Ab**: fluoroquinolones-resistant *A. baumannii*; **CR-Ab**: carbapenem-resistant *A. baumannii*; **VR-E**: vancomycin-resistant Enterococci; **PR-Sp**: penicillin-non-susceptible *S. pneumoniae*; **MLR-Sp**: macrolides-resistant *S. pneumoniae*.

We found that key factors associated with ABR rates varied greatly between drug-bug pairs but were more similar within same bacterial species. First, antibiotic sales were only significantly associated with ABR rates in five drug-bug pairs: FR-*Ec* and FR-*Pa* rates were positively associated with quinolones sales; CR-*Ab* rates were also positively correlated with carbapenems sales but inversely correlated with global antibiotic sales; finally, PR-*Sp* and MLR-*Sp* rates were positively associated with global antibiotic sales. Second, meteorological factors were mostly found to be associated with Enterobacterales and *S. pneumoniae* drug-bug pairs: all *E. coli* resistances were significantly associated with average temperature and extreme climatic events; average temperature was also strongly associated with CR-*Kp*. PR-*Sp* rates were inversely associated with average relative humidity and MLR-*Sp* rates were positively associated with extreme events. Lastly, rainfall was negatively associated to FR-*Ab* rates. Third, we found that the GHS index was inversely associated with ABR rates in all *E. coli* and *K. pneumoniae* associated drug-bug pairs, as well as in *P. aeruginosa* and *A. baumannii* pairs. CR-*Ab* was the only pair positively associated with the GDP. FR-*Pa* and MLR-*Sp* were negatively associated with the GDP. This agrees with the univariate analysis, in which most drug-bug pairs were significantly inversely associated with the GDP whereas CR-*Ab* was the only pair where the relationship was positive (appendix pp 44-51). Finally, tourist departures were inversely associated with ABR rates in two pairs, APR-*Ec* and CR-*Pa*. VR-*E* was not significantly associated with any of the proposed factors in the multivariable analysis. Of interest, population density and tourism arrivals were not selected for any drug-bug pairs. Univariate analyses results are summarized in appendix pp 45-52 and full models results (before backward selection) in appendix pp 53-54.

For all drug-bug pairs, we found higher heterogeneity in ABR rates between countries than between years (table 1): in reference M_null models, spatial RE variance was high (ranging from 0×03 for APR-*Ec* to 1×92 for CR-*Kp*) in comparison with temporal RE variance (<1^e-04^ for MLR-*Sp* to 0×5321 for CR-*Kp*). For all drug-bug pairs, the M_multi model had lower AIC compared with M_null (table 1) suggesting that multivariable model was better at explaining the data.

In mixed-effect models, the introduction of explanatory factors is expected to decrease variance in RE if the co-variable explains well the observed variance between groups of interest. While temporal RE variance was reduced in final multivariable models in all pairs except for APR-*Ec* and FR-*Ab*, the low variance first observed in null models impeded strong interpretations of such reduction. On the contrary, spatial RE variance in null models was relatively high and reduction of variance in final multivariable models is presented in figure 3. As shown from the results of F-tests in figure 3A, the introduction of co-variables in the multivariable model led to a significant spatial reduction of RE variance compared to the null model in *E. coli* resistances, FR-*Kp*, 3GCR-*Kp*, FR-*Pa* and CR-*Pa*. The resulting unexplained spatial RE variances after introduction of explanatory variables in final M_multi models are shown in figure 3B. We observed that some country-drug-bug trios systematically exhibited outlying spatial RE estimates compared to the observed average ABR rate in ATLAS. For example, spatial RE estimate was systematically high for Mexico (for all *E. coli* pairs and PR-*Sp*) and South Korea (for FR-*Ec*, 3GCR-*Ec*, FR-*Kp*, FR-*Pa*, FR-*Ab*, CR-*Ab*, PR-*Sp* and MLR-*Sp*) and low for Japan (in APR-*Ec*, FR-*Kp*, 3GCR-*Kp*, FR-*Ab* and CR-*Ab*) and Czech Republic (CR-*Kp*, CR-*Ab* and PR-*Sp*). This suggests that the proposed co-variables are not sufficient to explain the observed spatial heterogeneity in ABR rates, especially in these countries (appendix pp 55-62).

**Figure 3.**
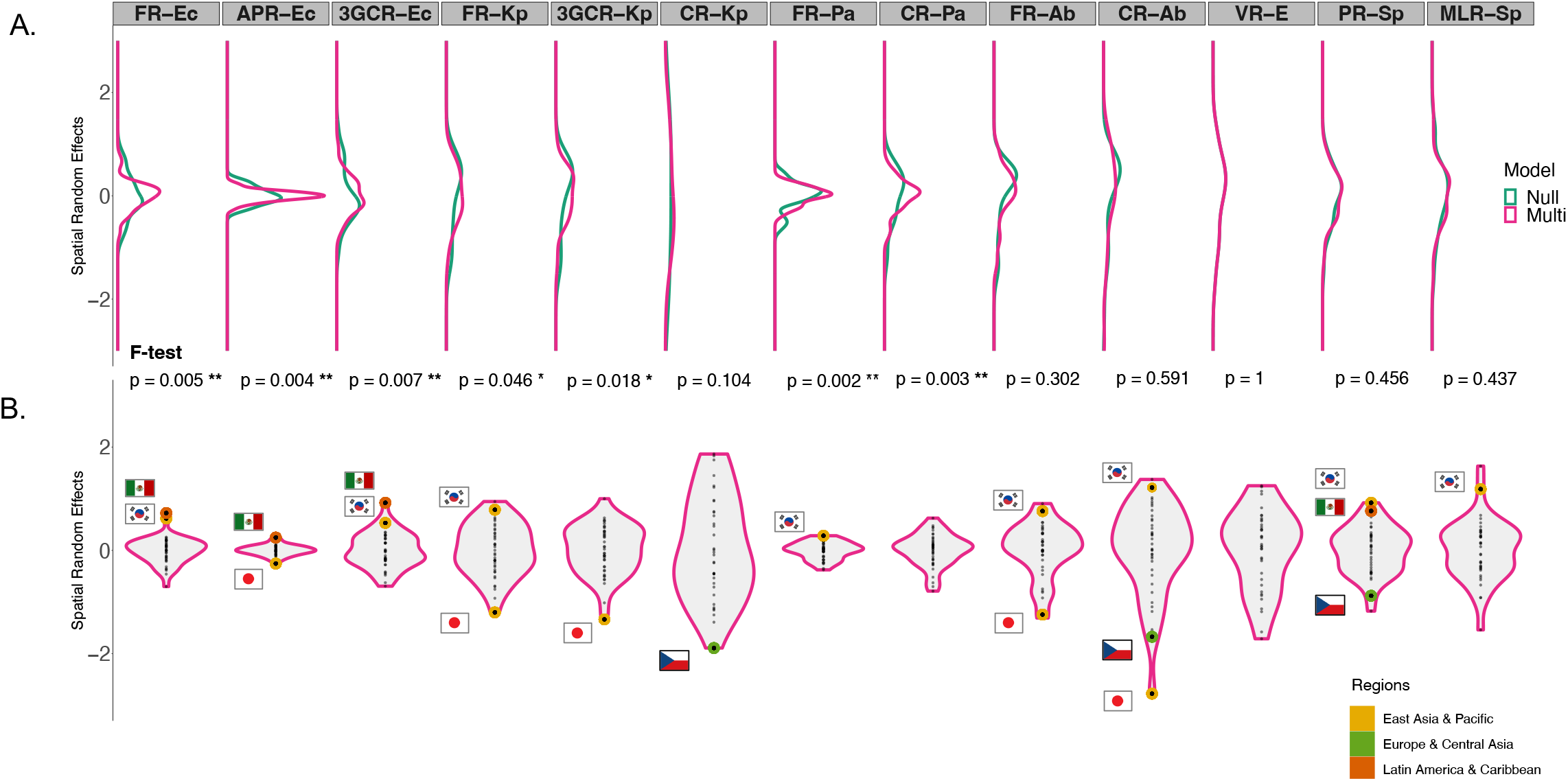
Spatial random effects distribution from final multivariable models for each drug-bug pair. (A) Distribution of spatial random effects by drug-bug pair. In blue, distributions from null models (M_null) and in pink, distributions from final multivariable models (M_multi). Significance of F-tests to test variance difference between spatial random effects of M_null and M_multi are reported below distributions (*: p-value < 0.05; **: p-value < 0.01; ***: p-value < 0.001). (B) Violin plots of spatial random effects resulting from final multivariable models (M_multi) by drug-bug pair. Random effects’ estimates from Czech Republic (Europe and Central Asia), Japan, South Korea (East Asia and Pacific) and Mexico (Latin America and Caribbean) are highlighted based on their world’s regions and corresponding flags are shown. (All panels) **FR-Ec**: fluoroquinolones-resistant *E. coli*; **APR-Ec**: aminopenicillins-resistant *E. coli*; **3GCR-Ec**: third generation cephalosporins-resistant *E. coli*: **FR-Kp**: fluoroquinolones-resistant *K. pneumoniae*; **3GCR-Kp**: third generation cephalosporins-resistant *K. pneumoniae*; **CR-Kp**: carbapenem-resistant *K. pneumoniae*; **FR-Pa**: fluoroquinolones-resistant *P. aeruginosa*; **CR-Pa**: carbapenem-resistant *P. aeruginosa*; **FR-Ab**: fluoroquinolones-resistant *A. baumannii*; **CR-Ab**: carbapenem-resistant *A. baumannii*; **VR-E**: vancomycin-resistant Enterococci; **PR-Sp**: penicillin-non-susceptible *S. pneumoniae*; **MLR-Sp**: macrolides-resistant *S. pneumoniae*.

Sensitivity analyses were carried out to evaluate the impact of some of our methodological choices: threshold on the number of isolates below which drug-bug-country-year observations were excluded, inclusion of external antibiotic sales data for countries for which IQVIA MIDAS data were not available, and finally imputation of some missing country-year ABR rates. Results leading to similar interpretations to those presented in the main analysis were obtained, suggesting no major impact of our assumptions. We also carried out a sensitivity analysis on blood isolates only. Limiting to the blood source led to lower sample sizes (appendix p 13). For example, 24% of all *E. coli* and VR-*E* isolates came from blood, with numbers down to 11% for *P. aeruginosa*. Resulting associations were overall consistent with the main analysis, with wider 95% confidence intervals but similar coefficient estimations and sign of associations with few exceptions. First, the analysis of blood samples led to negative association of GDP with CR-*Ab* and 3GCR-*Kp* ABR rates, whereas it was positively associated in the main analysis. Second, the association between temperature and CR-*Ab*, PR-*Sp* and VR-*E* showed higher estimated coefficients, suggesting stronger correlation. Lastly, some covariables associations had different signs for *S. pneumoniae* associated pairs, such as macrolides and temperature association with MLR-*Sp*. Sensitivity analyses description and results are reported in appendix pp 43-44 and pp 45-52.

## Discussion

Using longitudinal data provided by the surveillance program ATLAS, we analyzed antibiotic resistance (ABR) spatial and temporal patterns as well as ABR determinants for thirteen drug-bug pairs of clinical relevance. Our results confirmed that worldwide ABR dynamics were highly drug-bug pair dependent, both spatially and temporally. We found that key factors varied greatly between drug-bug pairs, but some similarities existed between bacteria of the same species. However, after hypothesis driven investigation of factors, high unexplained country-level variance remained in most of the drug-bug pairs.

The ATLAS data exhibited strong heterogeneities in spatial and temporal ABR patterns across drug-bug pairs. In 2019, high median ABR rates were observed for APR-*Ec* and CR-*Ab* but with different spatial heterogeneities. Aminopenicillins are broad-spectrum penicillins, the most used antibiotics class worldwide.^22^ Between 2000 and 2015, their consumption in low- and middle-income countries (LMICs) doubled, reaching levels observed in high-income countries (HICs).^22^ In contrast, consumption of carbapenems - a last resort antibiotics class - increased in all countries but levels were still much lower in LMICs compared to HICs. This could explain the large inter-country variation observed in CR-*Ab* rates compared to APR-*Ec* in 2019.^22^ Moreover, we found that carbapenem-resistant Gram-negative bacteria were on the rise globally in ATLAS in agreement with previous studies.^23,24^

The antibiotic consumption-resistance association strongly depended on the considered drug-bug pair. We found that FR-*Ec*, FR-*Pa* and CR-*Ab* ABR rates were significantly associated with sales of their selecting antibiotics (quinolones and carbapenems respectively) in the multivariable analysis (table 1). ABR rates in *S. pneumoniae* pairs were significantly associated with global antibiotic sales. Bystander selection certainly plays a role in this association, especially in commensals such as *S. pneumoniae*, where antibiotics select resistance in off-target bacteria in microbiota.^25^ Surprisingly, none of the 3GC resistances were associated to any antibiotic sales. This could be due to the fact that reported 3GC resistances trends are relatively stable whereas cephalosporins consumption is decreasing, especially in HICs.^22^ While antibiotics consumption exerts a selective pressure and favors the emergence of resistant bacteria, our results suggested that dissemination and persistence of such resistant bacteria might contribute more widely to the observed ABR heterogeneities worldwide. Significant signals were found in all drug-bug pairs for other factors including climatic and meteorological factors, the health system quality, wealth, or tourism. Thus, the spread of resistant bacteria could influence ABR rates in populations where antibiotic use is low. Indeed, it has been shown that spillover can happen at large geographical scales, weakening observed global antibiotic consumption-resistance associations.^26^

Warm temperatures were found to be significantly associated with ABR rates only for Enterobacterales (*E. coli* and *K. pneumoniae*) in the multivariable analysis (table 1). This is consistent with previous ecological studies from smaller datasets in the United States and in Europe.^7–9^ Here, we show that this association is highly drug-bug pair dependent, as the association does not hold true for other bacterial species. Extreme climatic events were only positively associated in *E. coli* and *S. pneumoniae*. This result suggests that natural disasters such as flooding, rather than rainfall itself, potentially contribute to ABR spread by disrupting infrastructures and creating sanitation failures.^27^ Examples from the literature report elevated numbers of infections with antibiotic resistant bacteria in natural disasters survivors or unprecedented levels of ABR genes in water following hurricanes.^28,29^ This association might be especially strong in orofecally-transmitted bacteria such as *E. coli*.

High GHS index, used as a proxy for health system quality, was significantly associated with decreased ABR rates in most drug-bug pairs. This result highlights the crucial role of hygiene and infection control measures for containing resistance, especially in hospitals. They are consistent with findings from Collignon et al. which showed that aggregated resistance in *E. coli, K. pneumoniae* and *S. aureus* were significantly inversely correlated with infrastructures and health expenditure.^4^ In our analysis, the GHS index was not associated in VR-*E* nor in *S. pneumoniae* pairs, the only two investigated Gram-positive bacteria. For VR-*E*, this could potentially reflect discrepancies in nosocomial infection control approaches regarding this specific pathogen across countries with similar health system quality.^30^ For *S. pneumoniae*, this could be explained by the fact that its dissemination is mostly driven by community transmission rather than nosocomial transmission. Overall, these findings highlight the importance of testing the validity of global ABR associations in different clinically relevant bacterial resistances.

Carbapenem-resistant *A. baumannii* was the only pair positively associated with the GDP per capita (table 1). From the univariate analysis, we found a large reduction in temporal RE variance indicating that the individual contribution of GDP was mostly temporal. This result is consistent with other findings that showed a fastest increase in *A. baumannii* resistances in OECD countries compared to other countries, where OECD countries have a higher GDP per capita. ^24^ On the contrary, FR-*Pa* and MLR-*Sp* showed an inverse significant association with the GDP per capita and such association also led to decreased temporal RE variance. This suggests that economic growth over time is correlated with increase of emerging resistances such as CR-*Ab* but decrease of other resistances.

After introduction of significant explanatory variables in the final models, spatial RE variance exhibited strong heterogeneity across drug-bug pairs. For Enterobacterales pairs, such as FR-*Ec*, RE variance was low suggesting that the proposed factors (quinolones sales, average temperature, extreme climatic events, GHS index) explained most ABR heterogeneities between countries. On the contrary, for VR-*E* or carbapenem resistances, RE variances remained high suggesting that the proposed factors could not explain most of ABR spatial heterogeneities. This result stresses the lack of knowledge about global factors favoring the dissemination of newly emerged resistances. In addition, specific countries such as Mexico, Japan, or South Korea, exhibited high spatial RE estimates compared to other countries in ATLAS; and this was true across different drug-bug pairs (figure 3). In South Korea, it is documented that primary care system is less established than in other OECD countries.^31^ Patients thus mostly seek care in secondary and tertiary care system (e.g. hospitals), where the per capita antibiotic exposure is higher. Additional factors, like healthcare seeking behaviors, but also antibiotic use in livestock or percentage of rural population, might then better explain ABR rates for these outliers. Due to the lack of available data, such hypotheses could not be assessed in this study.

Our findings need to be interpreted in light of the following limitations. First, as sampling protocol was based on clinical isolates from hospitals, ABR rates reported in ATLAS might not be fully representative of true resistance prevalence at the country level. Isolates from severe infections could present more resistance as going to the hospital is the last option after treatment failure in the community. This would lead to over-estimations of ABR rates globally in ATLAS. In addition, analyzed sample sizes were small for some drug-bug-country-year observations, leading to high uncertainty around ABR rates. To evaluate such potential biases, we compared ABR rates estimated from ATLAS with those reported by EARS-Net in 20 European countries over 2006-2019. While ABR rates were higher in ATLAS for some drug-bug pairs, our results suggest a global consistency for average ABR rates and trends reported across systems (appendix pp 64-79). Because our statistical analyses focused on relative average ABR rates and trends, such possible over-estimations – if stable over countries and years – should not affect the associations with co-variables found in this study. Second, while this is a common feature in most ABR surveillance programs, the number of reported isolates in ATLAS increased over years (appendix p 13). To evaluate the potential impact of sampling variations, and because blood isolates collection in ATLAS was more stable over the study period, we ran a sensitivity analysis on blood source only. Results were mainly consistent with our main analysis that included isolates from all sources. Ultimately, this highlights the need for global efforts, such as the GLASS program, to evenly collect ABR data across the globe using standardized procedures, and to focus such efforts on still under-sampled regions.^32^

Choices in analyzed co-variables data can also be discussed. First, for the meteorological factors, the capital city was used as a proxy for the country’s weather, which could be misleading for large countries. However, previous studies suggested that using either the capital city’s data or the country’s weighted temperature mean did not affect associations.^8^ Second, we used here the IQVIA MIDAS database which currently represents one of the only source of harmonized data on global antibiotic sales.^22^ However, this data might under-estimate true antibiotic consumption as it does not include over-the-counter drugs. Within a country, distribution of retail versus hospital sales were not available, nor information about heterogeneity in use in different populations. Both phenomena could contribute to resistance selection.^33^ In addition, antibiotic use in livestock could not be included here due to lack of available data. In the future, access to more detailed and finer-grained data on antibiotic use in humans and animals should improve understanding of ABR drivers. Finally, tourism was measured here by the total numbers of tourists going in and out of a country. This metric possibly missed important information such as destination country. Yet, travel to certain African or Asian countries, rather than travel itself, has been shown to be a significant risk factor for resistant bacteria colonization.^34,35^ Such lack of precision in the available data could explain the unexpected inverse association with tourism-associated factors obtained in our statistical analysis.

Overall, our findings show that antibiotic resistance is a plural threat. Associations between antibiotic resistance rates and global factors, found in Enterobacterales for example, do not necessarily hold true in other bacterial species. Moreover, these known global factors cannot explain all heterogeneities observed worldwide. Our work highlights the importance to study determinants of worldwide antibiotic resistance in all drug-bug pairs of clinical relevance. Despite differences across pathogens, resistances are driven by both individual behaviors and global environmental mechanisms setting ABR in a One Health framework. Strategies to tackle worldwide antibiotic resistance should thus be tailored accounting for the species, the resistance, and the epidemiological settings at stake.

## Data Availability

The antibiotic resistance data can be visualized through the ATLAS website (https://atlas-surveillance.com). Co-variables data are publicly available through the World Bank Data website (https://data.worldbank.org); the Global Health Security index website (https://www.ghsindex.org); the Solar Radiation and Meteorological Data services website (http://www.soda-pro.com) for the MERRA-2 data; the ECDC website (https://www.ecdc.europa.eu/en/antimicrobial-consumption/surveillance-and-disease-data/database) for the ESAC-Net data. IQVIA MIDAS data are not publicly available.
R codes to describe country-year longitudinal resistance data for an antibiotic-bacterium pair of interest and quantify its association with candidate factors are available here: https://github.com/EveRah/drug_bug_pair_NBMM.

## Contributors

ER, PG and LO participated in the initial statistical design and results interpretation. ER analyzed the data and wrote the manuscript. PG and LO were major contributors in the writing of the manuscript. LW contributed to the statistical design and to the manuscript review. DG participated in the manuscript review.

## Data sharing

The antibiotic resistance data can be visualized through the ATLAS website (https://atlas-surveillance.com). Co-variables data are publicly available through the World Bank Data website (https://data.worldbank.org); the Global Health Security index website (https://www.ghsindex.org); the Solar Radiation and Meteorological Data services website (http://www.soda-pro.com) for the MERRA-2 data; the ECDC websites (https://www.ecdc.europa.eu/en/all-topics/related-topics) for the EARS-Net and ESAC-Net data. IQVIA MIDAS data are not publicly available.

R codes to describe country-year longitudinal resistance data for an antibiotic-bacterium pair of interest and quantify its association with candidate factors are available here: https://github.com/EveRah/drug_bug_pair_NBMM.

## Acknowledgments

We thank the data providers for sharing helpful material with us: IQVIA for the antibiotic sales data and Pfizer for the isolates data from the ATLAS database.

