## Appendix for "Worldwide antibiotic resistance dynamics: how different is it from one drug-bug pair to another?"

** Contributed equally*

#### Supplementary methods

***Supplementary Text S1. Description of the ATLAS database***

ATLAS is a surveillance system led by Pfizer, collecting antibiotic resistance (ABR) information on bacterial isolates from hospitalized patients, within a network of participating hospitals around the world. The network includes a limited number of hospitals in each country, and within each hospital a sub-sample of patients is selected. The primary goal is to monitor the activity of antibiotic molecules through longitudinal *in vitro* activity monitoring.

ATLAS collects information on hospitalized patient and associated bacterial isolate, including country, year, hospital ward, source of infection and bacterial species. Data represent infection isolates, with the bacteria being the putative cause of disease of the hospitalized patient. Sample selection is independent from the resistance status of the isolate but is based on the following criteria. A fixed number of the clinical isolates need to come from both Gram-positive and Gram-negative bacteria; and from five different infections (*and associated infection sources*):

- complicated intra-abdominal infections (*abscess* as most frequent source);
- complicated urinary tract infections (*urine*);
- complicated skin and skin structure infections (*wound*);
- lower respiratory tract infections (*sputum and endotracheal aspirate*);
- and blood infections (*blood*).

The origin of the infection, hospital or community-acquired, is not documented. Thus, it could represent a mix of both.

All bacterial isolates from participating hospitals are sent to a central laboratory. Minimal inhibition concentration (MIC) values against a vast number of antibiotic molecules are determined using broth micro-dilution and reported in the database.

From the MIC values, we interpreted the clinical status of isolates using the v9.0 2019 EUCAST standards by categorizing it into: susceptible, intermediary, or resistant.

Data from ATLAS analyzed in the present study represent 344 764 unique bacterial isolates (from all sources), and 808 744 non-unique isolates (different antibiotic tested across same bacterial species, from all sources). We included data from thirteen drug-bug pairs across 6 bacterial species, and from 51 countries (appendix p 4) across 14 years (2006-2019).

ATLAS antibiotic resistance surveillance data can be accessed and visualize through https://atlas-surveillance.com.

***Supplementary Figure S1. Countries included in the analysis (C=51 over 2006-2019).***

The following countries, by world's regions (from the World's Bank indicators), were included in the analysis:

- North America: United States, Canada.
- Latin America & Caribbean: Argentina, Brazil, Chile, Colombia, México, Venezuela and Central America (grouping Panamá, Guatemala, Honduras, Nicaragua, Costa Rica, El Salvador);
- Europe & Central Asia: Austria, Belgium, Croatia, Czech Republic, Denmark, Finland, France, Germany, Greece, Hungary, Ireland, Italy, Latvia, Lithuania, Netherlands, Poland, Portugal, Romania, Russia, Spain, Sweden, Switzerland, Turkey, United Kingdom.
- East Asia & Pacific: Australia, China, Hong Kong, Japan, South Korea, Malaysia, Philippines, Singapore, Taiwan, Thailand.
- South Asia: India, Pakistan.
- Sub-Saharan Africa: South Africa.
- Middle East & North Africa: Israel, Jordan, Kuwait, Morocco, Saudi Arabia.

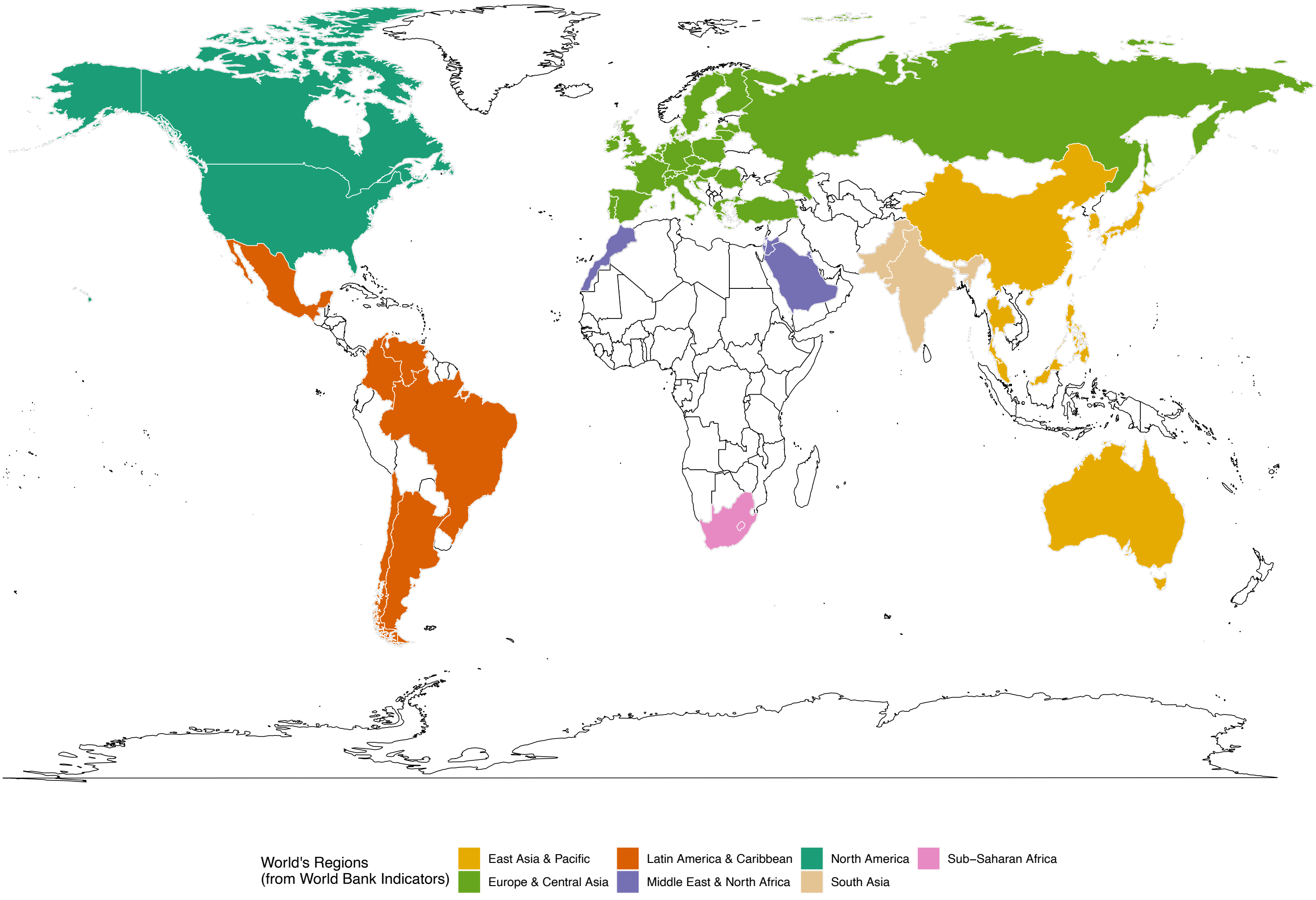

***Supplementary Table S1. Antibiotic molecules tested to determine antibiotic resistance susceptibility for isolates in the ATLAS database.***

| Resistance | Molecules tested |
|---|---|
| Fluoroquinolones | Ciprofloxacin and levofloxacin |
| Aminopenicillins | Ampicillin |
| Third generation cephalosporins | Ceftriaxone and ceftazidime |
| Carbapenems | Imipenem and meropenem |
| Penicillin | Penicillin G |
| Vancomycin | Vancomycin |
| Macrolides | Erythromycin |

***Supplementary Table S2. Co-variables data summary used in the statistical analysis.***

Co-variables, unit, source, number of countries and years for which data are available are summarized in the following data.
Average (and standard deviation SD), minimum and maximum values are calculated over all countries and years available.

| | Co-variables | Unit | Source | Available countries (C=51) | Available years | Average (SD) | Minimum | Maximum |
|---|---|---|---|---|---|---|---|---|
| Country-year observations | **Population density** | People/km$^2$ | World Bank Data | **50** (Taiwan not available) | **2006-2019** | 417.6 (1 380.3) | 2.7 | 7 990.3 |
| | **GDP per capita, PPP** | Current international dollar ($) | World Bank Data | **50** (Taiwan not available) | **2006-2019** | 31 530.7 (18 506.8) | 3237 | 101 376 |
| | **Tourism Arrivals** | Annual number of tourists arriving over national population (%) | World Bank Data | **50** (Taiwan not available) | **2006-2019** | 141.5 (215.3) | 0.38 | 1475.6 |
| | **Tourism Departures** | Annual number of tourists departing over national population (%) | World Bank Data | **48** (Pakistan, South Africa, Malaysia and Taiwan not available) | **2006-2019** | 99.5 (180.6) | 0.71 | 1261.6 |
| | **Broad spectrum penicillins sales** | Grams/1000 inhabitants/day | IQVIA MIDAS Data or ESAC-Net Data | **47**<br>43 via IQVIA MIDAS and 4 via ESAC-Net (Denmark, Netherland, Latvia, Lithuania)<br>(Israel, Jordan, Taiwan and Venezuela not available) | **2006-2019** | 11.3 (6.8) | 0.6 | 33.3 |
| | **Cephalosporins sales** | | | | | 2.7 (2) | 0.05 | 13.3 |
| | **Carbapenems sales** | | | | | 0.1 (0.09) | 0 | 0.4 |
| | **Quinolones sales** | | | | | 1.5 (0.8) | 0.18 | 4.2 |
| | **Macrolides sales** | | | | | 1.4 (0.8) | 0.21 | 6.2 |
| | **Trimethoprim and combinations sales** | | | | | 1.2 (1.5) | 0 | 24.7 |
| | **Glycopeptides sales** | | | | | 0.04 (0.05) | 0 | 0.4 |
| | **Average annual Temperature** | Kelvin | MERRA-2 Data | **51** | **2006-2019** | 288.38 (6.9) | 276.6 | 301.9 |
| | **Average annual Rainfall** | Rain depth in millimeters | MERRA-2 Data | **51** | **2006-2019** | 96.5 (95.5) | 3 | 512.4 |
| | **Average annual Relative Humidity** | Normalized pseudorelative humidity (water vapor mixing ratio scaled by its saturation value, %) | MERRA-2 Data | **51** | **2006-2019** | 70.7 (15.2) | 19.6 | 90 |
| Country observations | **GHS index** | | Global Health Security index Data | **49** (Hong Kong and Taiwan not available) | **2019** | 57.5 (12) | 23 | 83.5 |
| | **Population facing extreme climatic events** | Annual number of people facing extreme climatic events over national population (%) | World Bank Data | **49** (Singapore and Taiwan not available) | **2019** | 0.6 (1.4) | 0 | 7.9 |

***Supplementary Text S2. Description of the IQVIA MIDAS database.***

IQVIA MIDAS is a database collecting antibiotic sales from both retailers and hospitals at the country-year level around the world. It represents data from both community and hospital use. Data are global official sales of antibiotics aggregated at the country-year level, in kilograms.

---

***Supplementary Table S3. DDD to gram conversion coefficients A by antibiotic class.***

Coefficients are calculated based on consumption data in the community and hospitals from available countries in both ESAC-Net and IQVIA MIDAS. It is constructed as follow:
*Antibiotic consumption in grams/1000inh/day* $_{IQVIA}$ *= A * antibiotic consumption in DDD/1000inh/day* $_{ECDC}$

| Antibiotic class | A (DDD to gram conversion) |
|---|---|
| Broad spectrum Penicillin | 2 |
| Carbapenem | 3.5 |
| Cephalosporin | 1.4 |
| Quinolone | 0.95 |
| Macrolide | 0.6 |
| Trimethoprim/sulfonamide | 1.8 |
| Glycopeptide | 1.45 |

***Supplementary Table S4. Molecules included in each class for antibiotic sales categories.***

Classification is from *ResistanceMap*: https://resistancemap.cddep.org/MethodologyAU.php
WHO/ATC indexing system for antimicrobials: https://www.whocc.no/atc_ddd_index/

| Class | Molecules | Corresponding group in WHO/ATC indexing system |
|---|---|---|
| Broad spectrum Penicillins and combinations | Amoxicillin, Ampicillin, Amoxicillin-clavulanate, Ampicillin-sulbactam, Carbenicillin, Carfecillin, Carindacillin, Piperacillin-tazobactam, Ticarcillin, Sultamicillin, Piperacillin, Temocillin | J01CA<br>J01CR |
| Cephalosporins | Cefazolin, Cefaclor, Ceftibuten, Cefadroxil, Cefdinir, Cefditoren pivoxil, Cefepime, Ceftizoxime, Cefotetan, Cefotaxime, Cefoxitin, Cefpodoxime proxetil, Cefprozil, Ceftazidime, Cefuroxime axetil, Ceftriaxone, Cefuroxime, Cefalexin, Cefradine, Loracarbef, Cefixime, Ceftaroline fosamil, Faropenem | J01DB<br>J01DC<br>J01DD<br>J01DE |
| Carbapenems | Doripenem, Imipenem-cilastatin, Ertapenem, Meropenem | J01DH |
| Quinolones | Moxifloxacin, Ciprofloxacin, Gemifloxacin, Ofloxacin, Levofloxacin, Lomefloxacin, Norfloxacin, Enoxacin, Gatifloxacin, Trovafloxacin, Sparfloxacin | J01M |
| Macrolides | Erythromycin, Azithromycin, Clarithromycin, Dirithromycin, Telithromycin, Clindamycin, Fidaxomicin, Lincomycin, Troleandomycin, Dalfopristin-quinupristin | J01F |
| Trimethoprim and combinations of sulfonamides | Sulfamethoxazole-trimethoprim, Sulfamethoxazole, Sulfisoxazole, Sulfadiazine, Trimethoprim | J01EA<br>J01EC<br>J01EE |
| Glycopeptides | Vancomycin, Telavancin | J01XA |

***Supplementary Figure S2. Correlogram for covariables data used in the mixed-effect negative binomial model (for all 51 countries and 14 years).***

> *GDP: Gross Domestic product; GHS: Global Health Security Index; D: population density; P: population number; Q_C: quinolone consumption; BSP_C: broad spectrum penicillin consumption; CPH_C: cephalosporin consumption; M_C: macrolide consumption; T_C: trimethoprim consumption; CPM_C: carbapenem consumption; G_C: glycopeptide consumption; R: average rainfall; AV_T: average temperature; MIN_T: minimum temperature; H: average relative humidity; E_E: extreme climatic events, T_A: tourism arrivals; T_D: tourism departures; ATB_G: global consumption.*

Shown here are the Pearson correlation coefficient for each pair of variables (r). Blue colors represent positive correlation between variables. Red colors represent negative correlation. Only significant correlation coefficients (p-value < 5%) are shown in the correlogram. Co-variables associated with $r > |0{\cdot}7|$ were excluded from the model. Population number and Minimum Temperature were excluded (because considered less biologically relevant or less correlated to ABR rates). Global antibiotic consumption was not included at the same time as broad-spectrum penicillin consumption in any model.

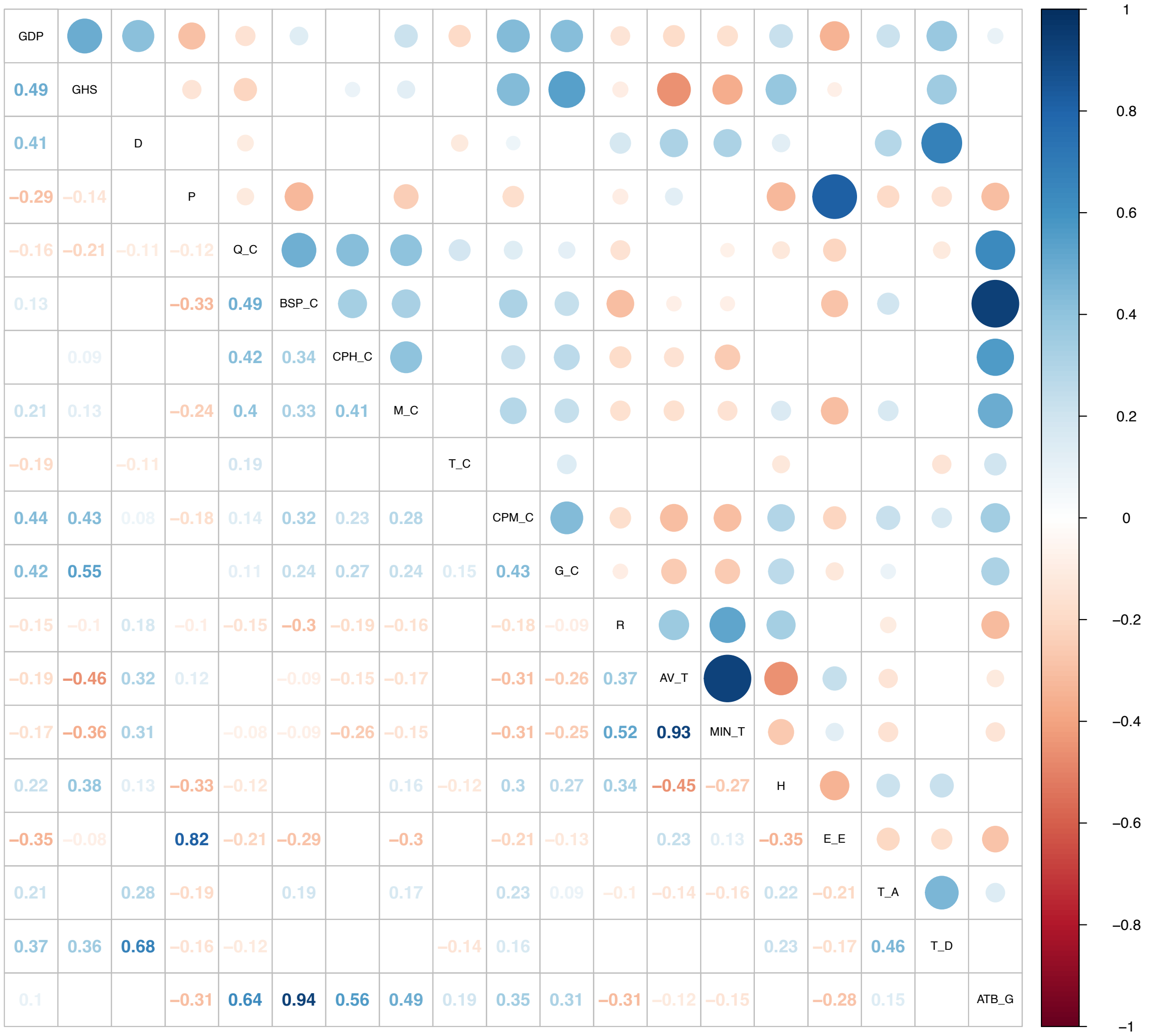

***Supplementary Table S5. Number of countries and observations included by drug-bug pair, at each stage of analyses.***

In red, the number of countries included. In blue, the number of observations included.
An observation is an ABR rate datapoint for a drug-bug-country-year combination.

| | Number of Countries & Observations included at each stage (2006-2019) | | | | | |
|---|---|---|---|---|---|---|
| | First stage: **Descriptive statistics** | | Intermediary stage: **Exclusion of countries with many 0% resistance** | | Final stage: **Final model analysis** (exclusion of observations with no covariables) | |
| **FR-Ec** | 50 | 643 | - | - | 41 | 471 |
| **APR-Ec** | 50 | 643 | - | - | 41 | 471 |
| **3GCR-Ec** | 50 | 643 | - | - | 41 | 471 |
| **FR-Kp** | 51 | 641 | - | - | 41 | 467 |
| **3GCR-Kp** | 51 | 641 | - | - | 41 | 467 |
| **CR-Kp** | 51 | 641 | 44 | 563 | 35 | 413 |
| **FR-Pa** | 50 | 640 | - | - | 41 | 472 |
| **CR-Pa** | 50 | 640 | - | - | 41 | 472 |
| **FR-Ab** | 47 | 577 | - | - | 37 | 410 |
| **CR-Ab** | 47 | 577 | - | - | 37 | 410 |
| **VR-E** | 49 | 621 | 45 | 473 | 37 | 423 |
| **PR-Sp** | 47 | 602 | - | - | 38 | 437 |
| **MLR_Sp** | 47 | 595 | - | - | 38 | 429 |

#### Supplementary data description

***Supplementary Figure S3. Number of isolates tested in ATLAS, by world's region for all sources isolates and blood isolates.***

(A) Number of total isolates tested in ATLAS from all infection sources per drug-bug pair (upper panel) and per year (lower panel).

(B) Number of total isolates tested in ATLAS from blood source only (*sensitivity analysis 3*) per drug-bug pair (upper panel) and per year (lower panel).

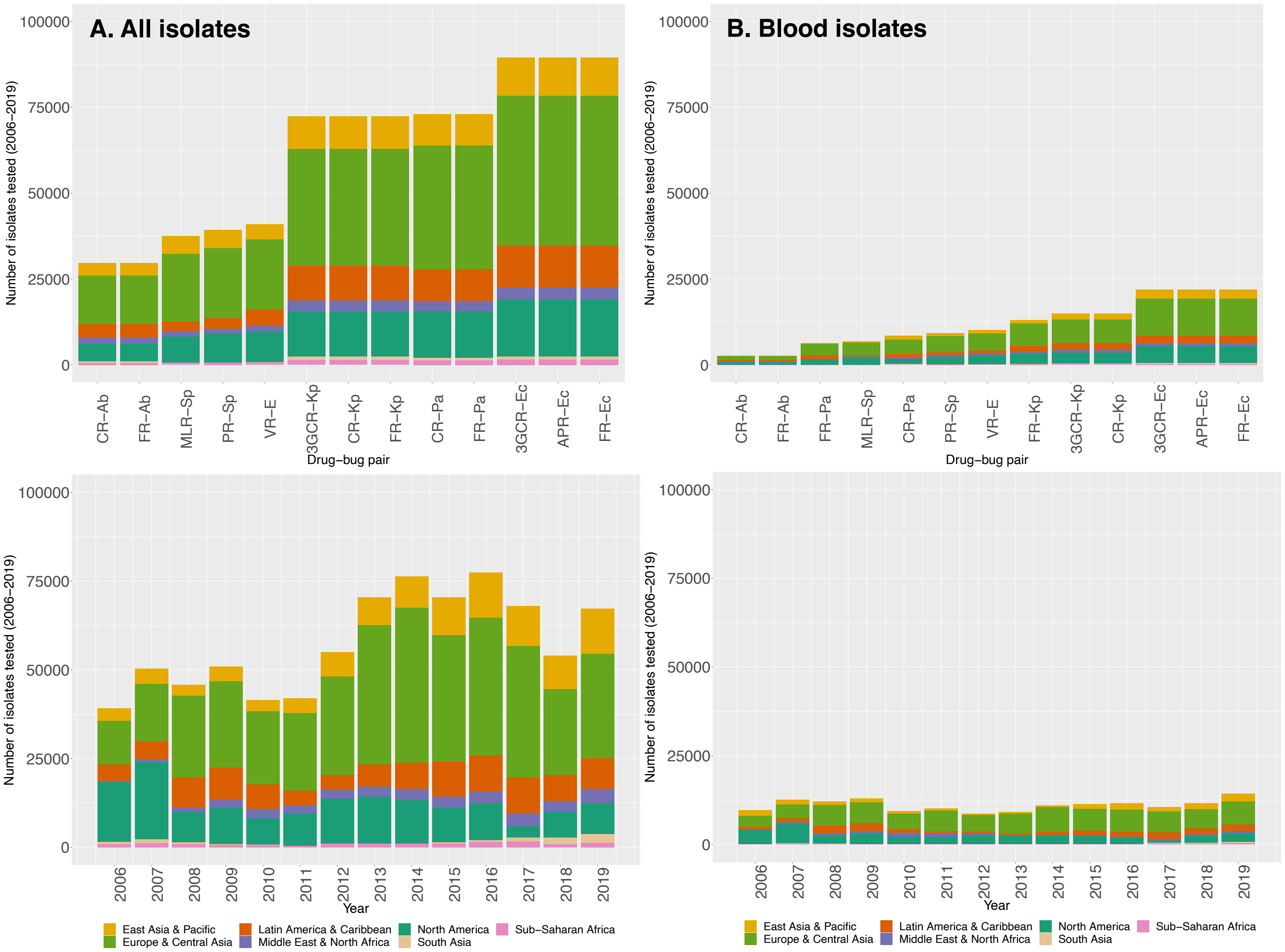

***Supplementary Figure S4. Distribution of MIC values for 3 years (2006, 2012 and 2019), by drug-bug pair from all infection sources.***

Represented here are the distribution of MIC values for all bacterial isolates tested against a specific class of antibiotics in ATLAS. If two molecules are considered in a class, highest MIC value between the 2 molecules is considered. MIC values are shown in log scale base 2.

The MIC breakpoints from the v9.0 2019 *European Committee on Antimicrobial Susceptibility Testing* (EUCAST*)* standards are depicted with the dashed lines.

Grouping into Resistant and Non-Resistant (or Susceptible and Non-Susceptible for PR-*Sp*) for each drug-bug pair was done based on the EARS-Net (ECDC) protocol.

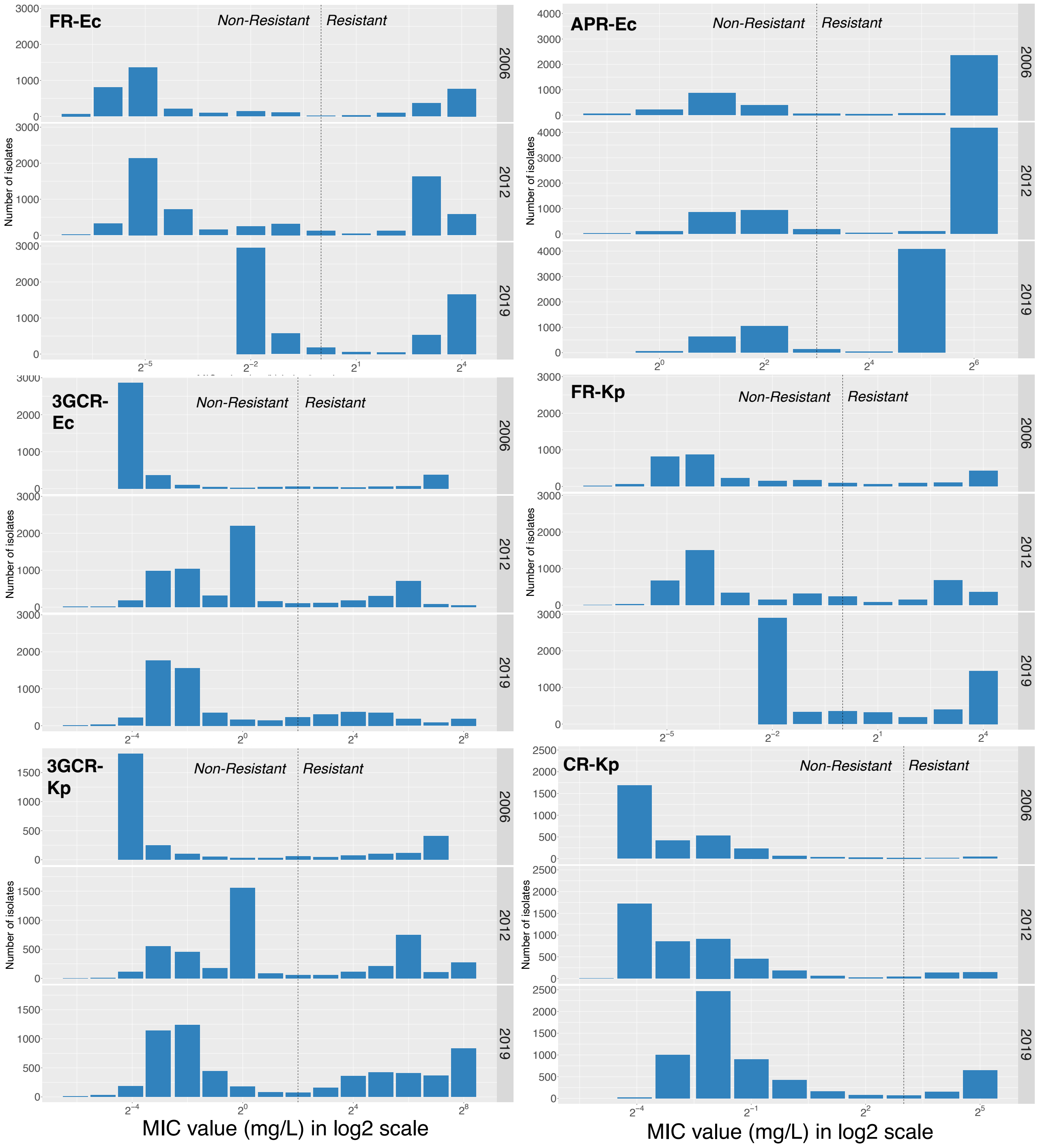

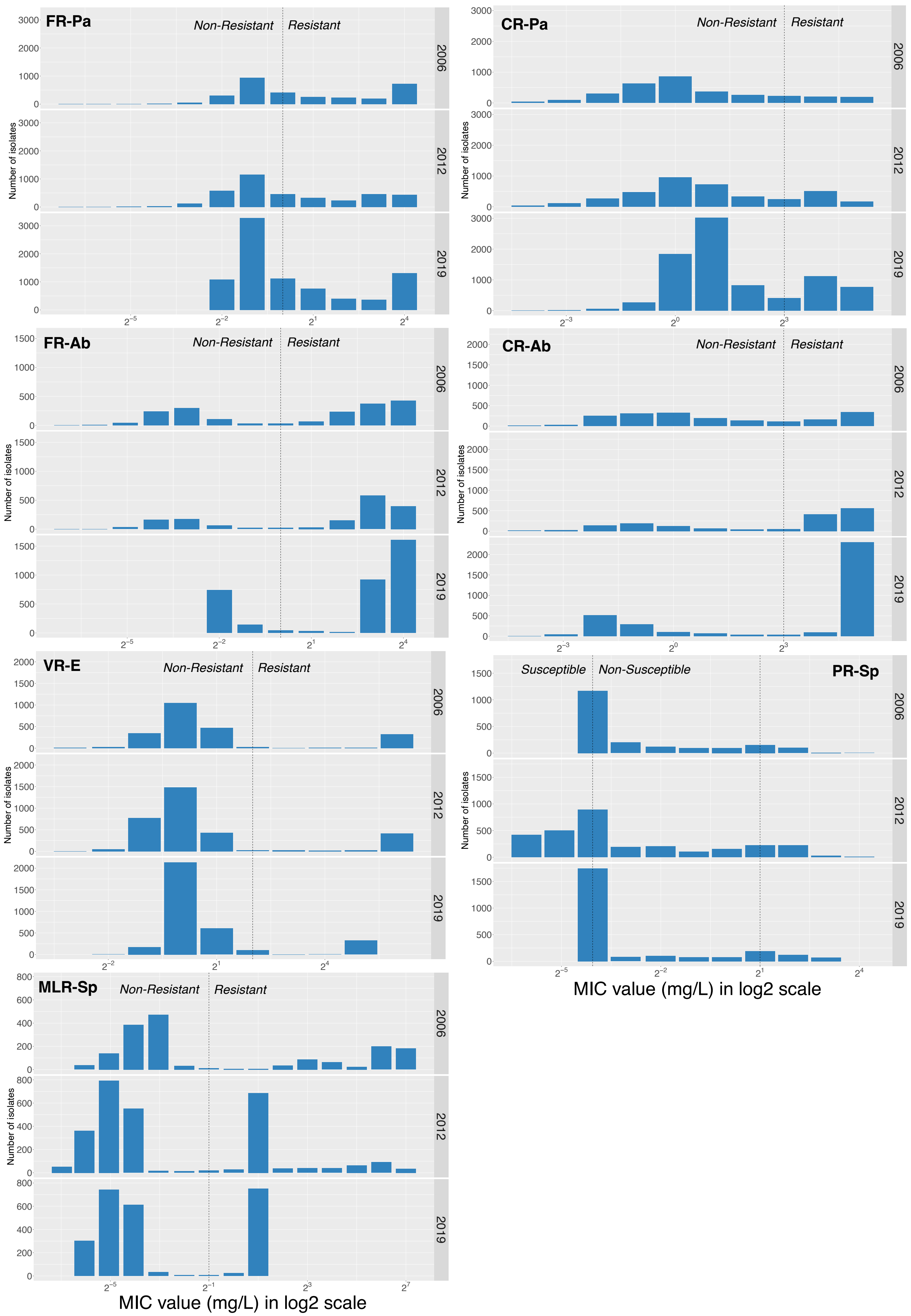
FR-Pa
CR-Pa
FR-Ab
CR-Ab
VR-E
PR-Sp
MLR-Sp
Non-Resistant
Resistant
Susceptible
Non-Susceptible
2006
2012
2019
Number of isolates
MIC value (mg/L) in log2 scale

***Supplementary Figure S5.*** **Number of isolates by bacterial species, infection sources and hospital wards from ATLAS.**

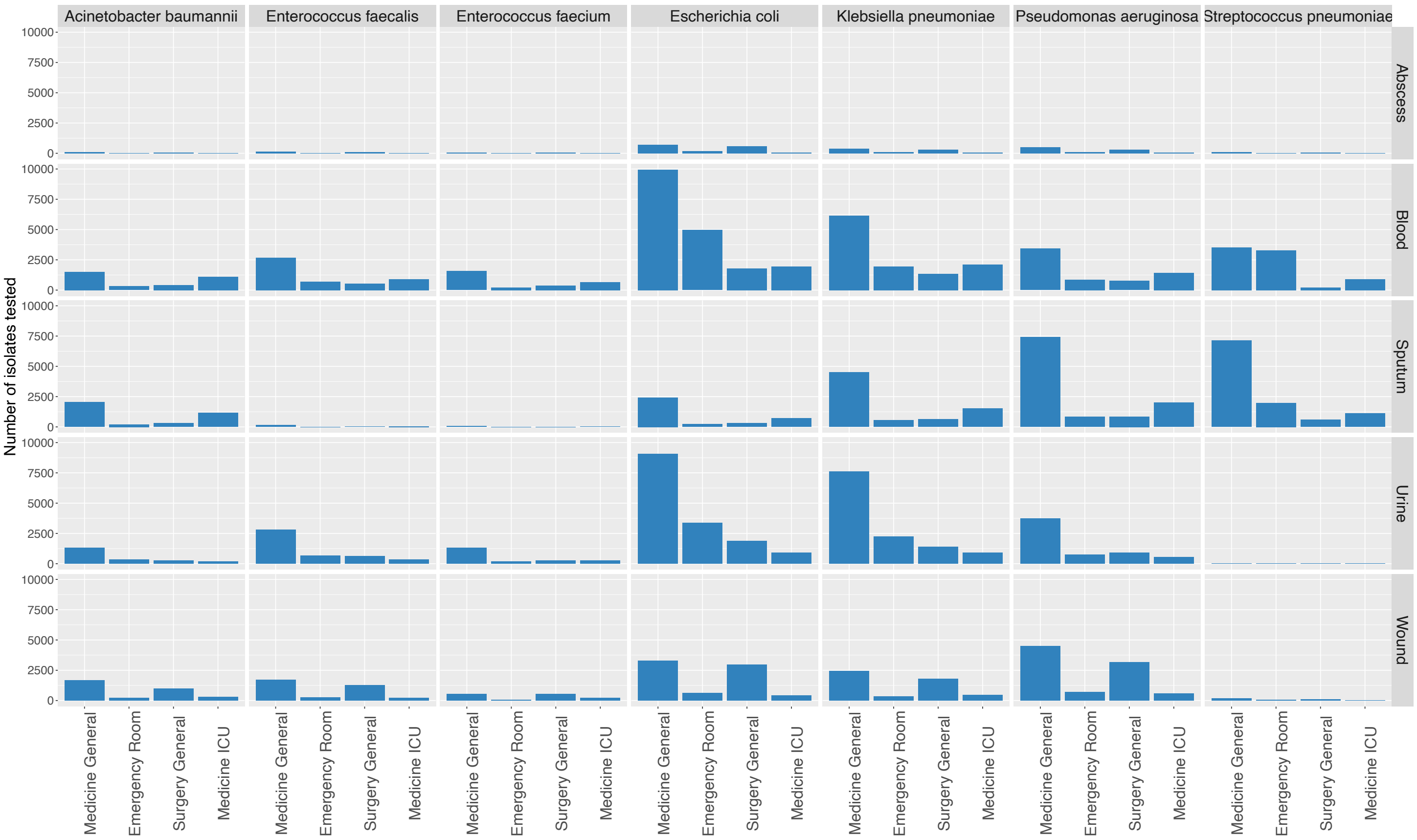

***Supplementary Figure S6.* Number of isolates by infection sources and countries from ATLAS.**

23 countries have **blood** as the most frequent source tested.
13 countries have **urine** as the most frequent source tested.
10 countries have **sputum** as the most frequent source tested.
4 countries have **wound** as the most frequent source tested.

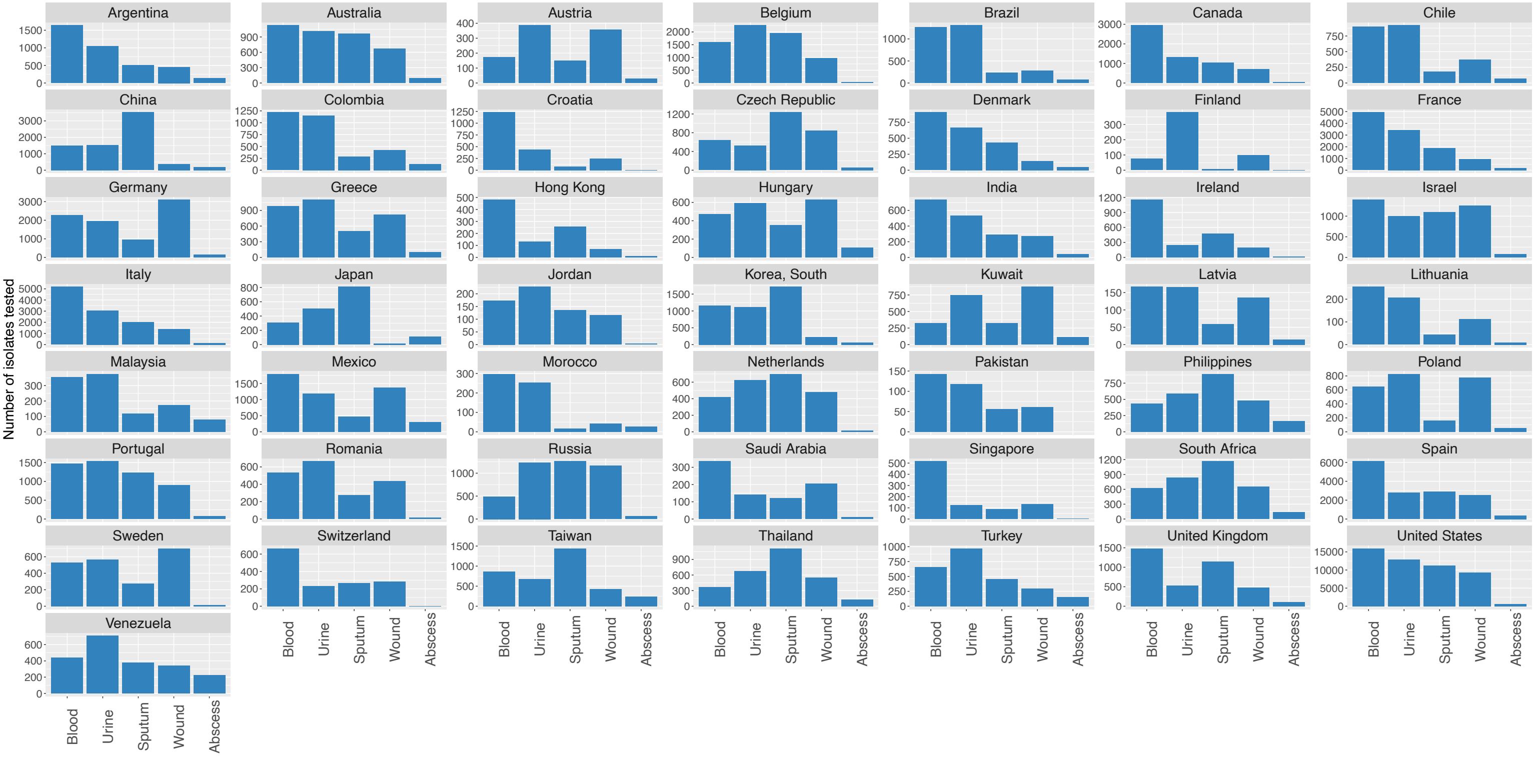

***Supplementary Figure S7. Distribution of ABR rates across countries, 2019: comparison between infection sources, by drug-bug pair.***

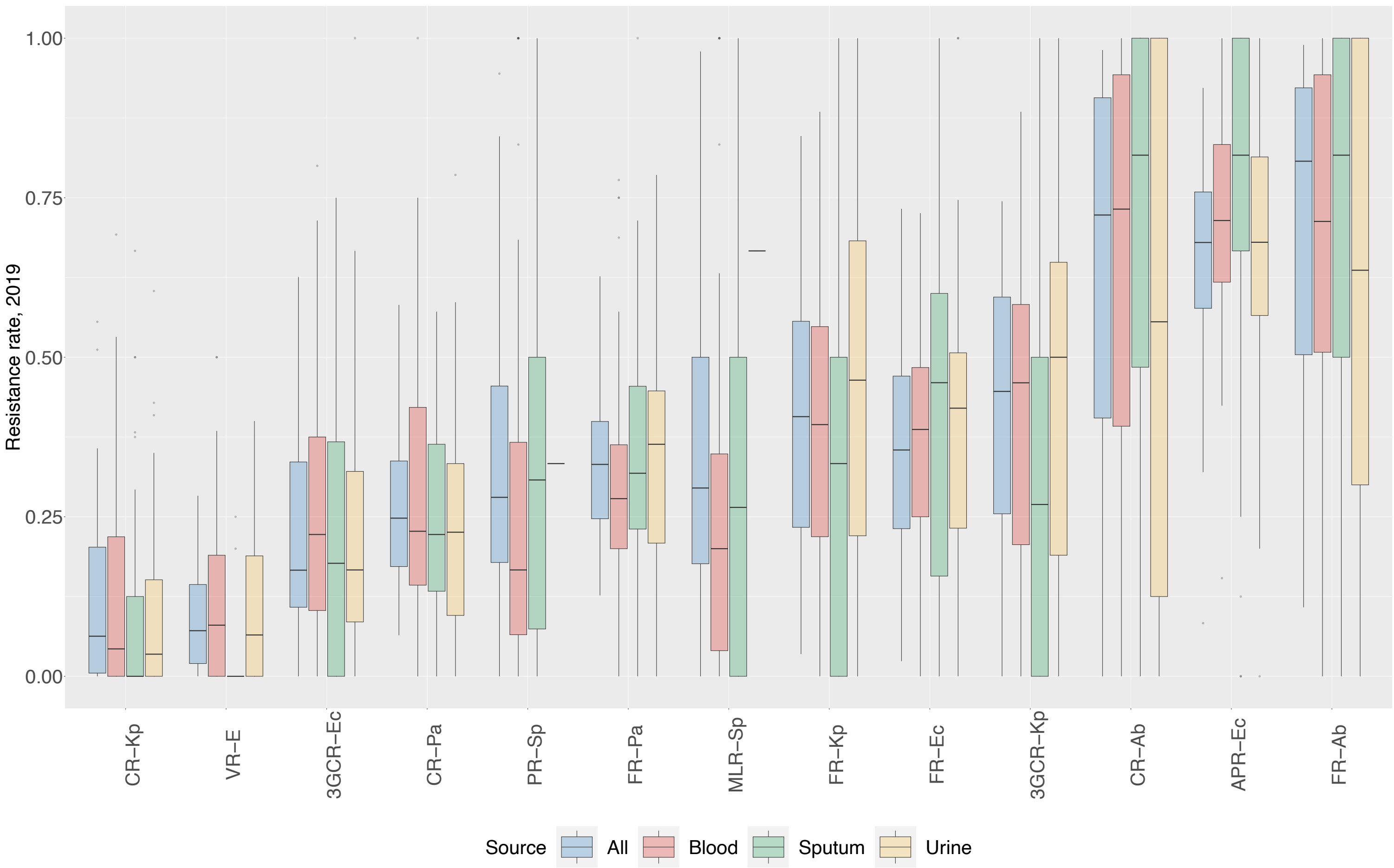

***Supplementary Figure S8. Temporal trends and sample sizes (2006-2019) per country, by drug-bug pair.***

Temporal trends and sample sizes for:

- **A. FR-Ec**: fluoroquinolone-resistant *E. coli*
- **B. APR-Ec**: aminopenicillin-resistant *E. coli*
- **C. 3GCR-Ec**: third generation cephalosporin-resistant *E. coli*
- **D. FR-Kp**: fluoroquinolone-resistant *K. pneumoniae*
- **E. 3GCR-Kp**: third generation cephalosporin-resistant *K. pneumoniae*
- **F. CR-Kp**: carbapenem-resistant *K. pneumoniae*
- **G. FR-Pa**: fluoroquinolone-resistant *P. aeruginosa*
- **H. CR-Pa**: carbapenem-resistant *P. aeruginosa*
- **I. FR-Ab**: fluoroquinolone-resistant *A. baumannii*
- **J. CR-Ab**: carbapenem-resistant *A. baumannii*
- **K. VR-E**: vancomycin-resistant Enterococci
- **L. PR-Sp**: penicillin-non-susceptible *S. pneumoniae*
- **M. MLR-Sp**: macrolide-resistant *S. pneumoniae*

*Temporal trends*

Estimation of the weighted linear regression temporal trends are shown in yellow. P-values associated with the temporal trend (regression slopes) are included on the top right. Blue color represents a decreasing trend. Red color represents an increasing trend.

95% confidence intervals (CI) around ABR rates (data) were calculated using Bayesian probability interval estimates with Jeffrey's prior.

*Sample sizes*

Sample sizes represent number of resistant isolates (upper number) and total number of isolates tested (lower number), per drug-bug-country-year combination.

A. **FR-*Ec***

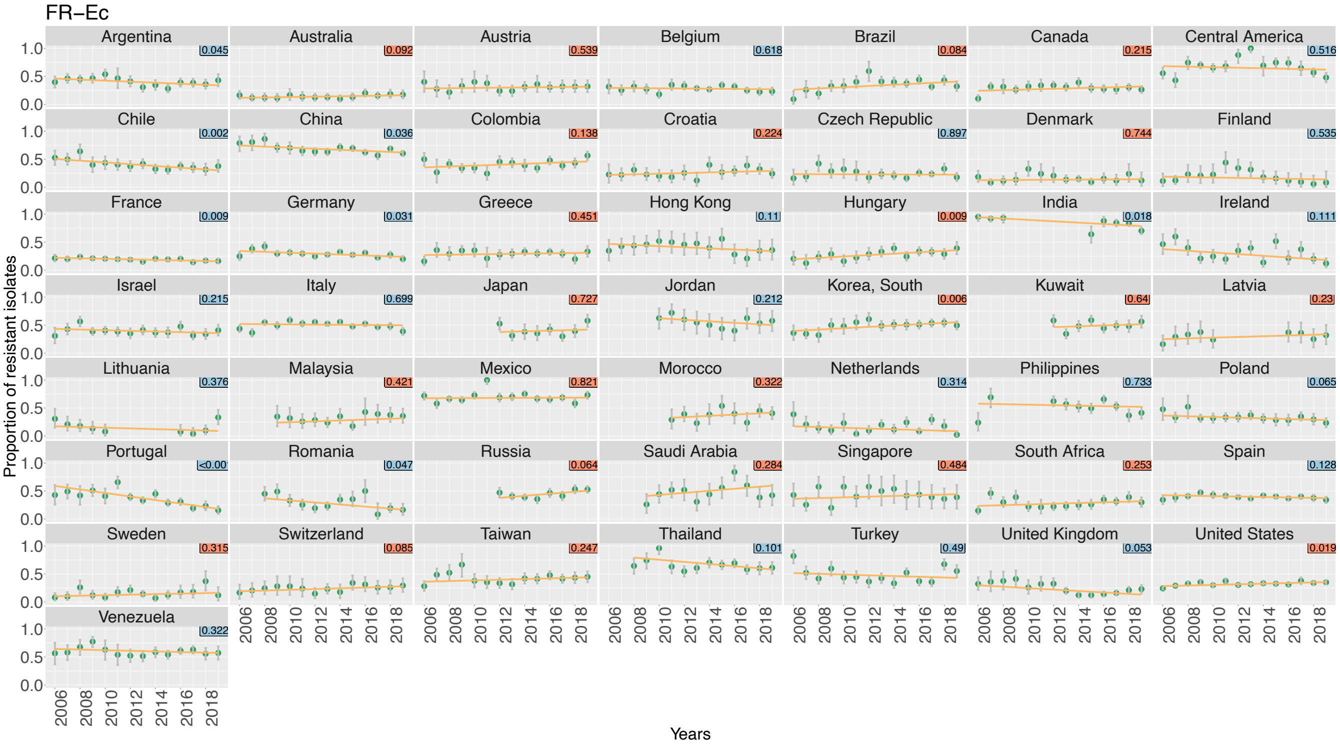
FR–Ec
Argentina
Australia
Austria
Belgium
Brazil
Canada
Central America
Chile
China
Colombia
Croatia
Czech Republic
Denmark
Finland
France
Germany
Greece
Hong Kong
Hungary
India
Ireland
Israel
Italy
Japan
Jordan
Korea, South
Kuwait
Latvia
Lithuania
Malaysia
Mexico
Morocco
Netherlands
Philippines
Poland
Portugal
Romania
Russia
Saudi Arabia
Singapore
South Africa
Spain
Sweden
Switzerland
Taiwan
Thailand
Turkey
United Kingdom
United States
Venezuela
Proportion of resistant isolates
Years

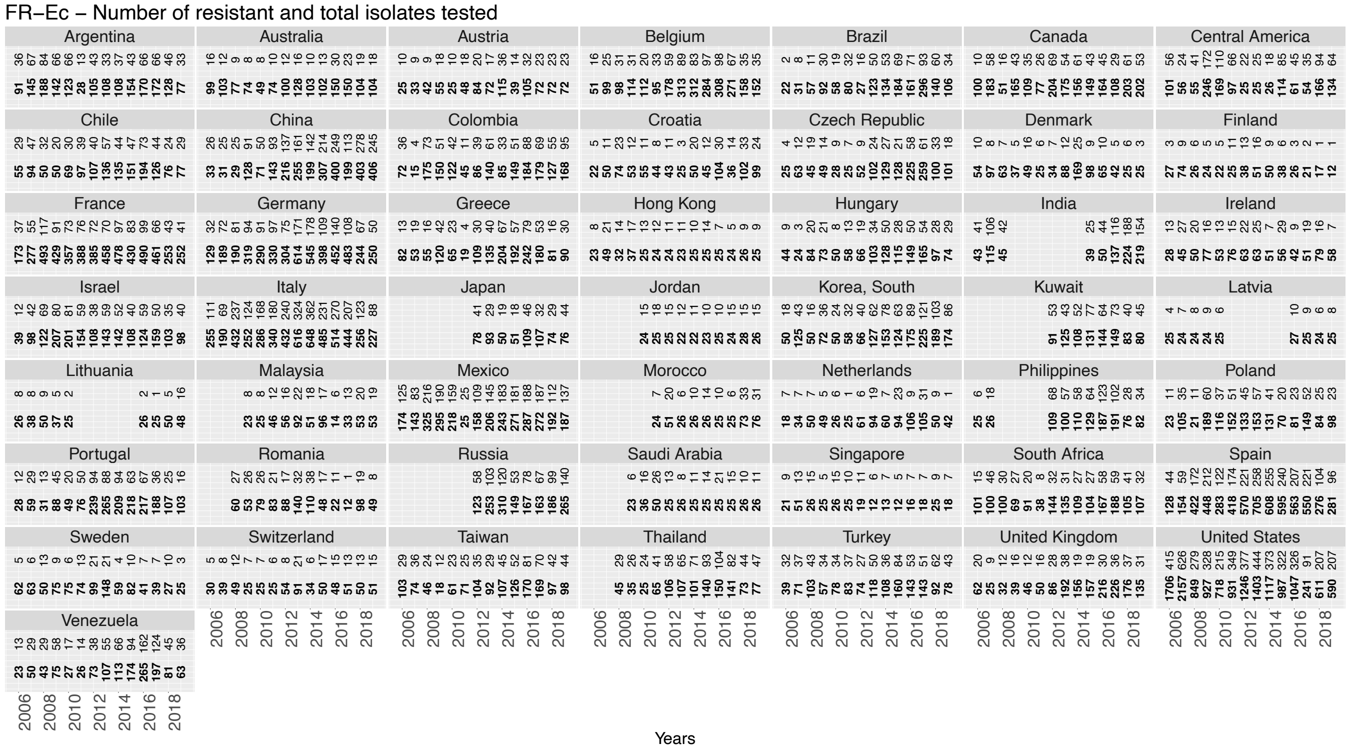
FR–Ec – Number of resistant and total isolates tested
Years

#### B. **APR-*Ec***

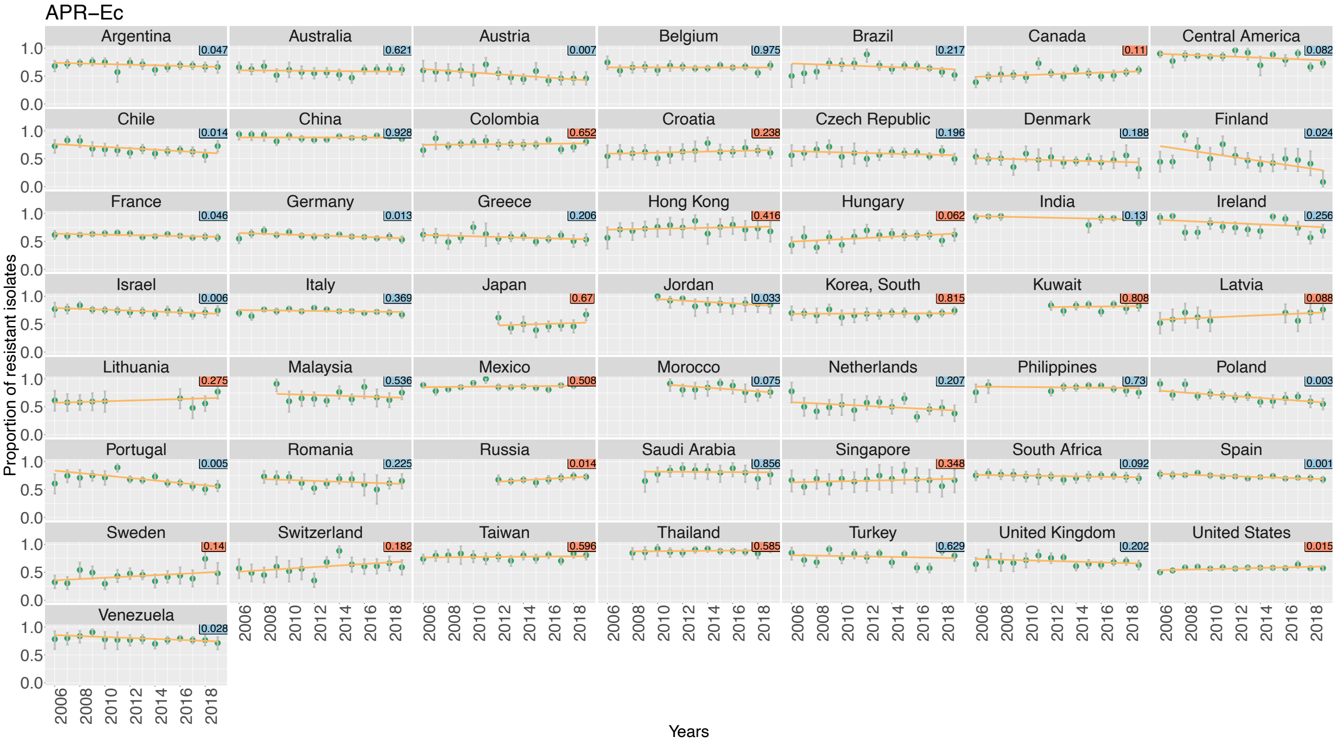
APR–Ec
Proportion of resistant isolates
Years
Argentina
Australia
Austria
Belgium
Brazil
Canada
Central America
Chile
China
Colombia
Croatia
Czech Republic
Denmark
Finland
France
Germany
Greece
Hong Kong
Hungary
India
Ireland
Israel
Italy
Japan
Jordan
Korea, South
Kuwait
Latvia
Lithuania
Malaysia
Mexico
Morocco
Netherlands
Philippines
Poland
Portugal
Romania
Russia
Saudi Arabia
Singapore
South Africa
Spain
Sweden
Switzerland
Taiwan
Thailand
Turkey
United Kingdom
United States
Venezuela

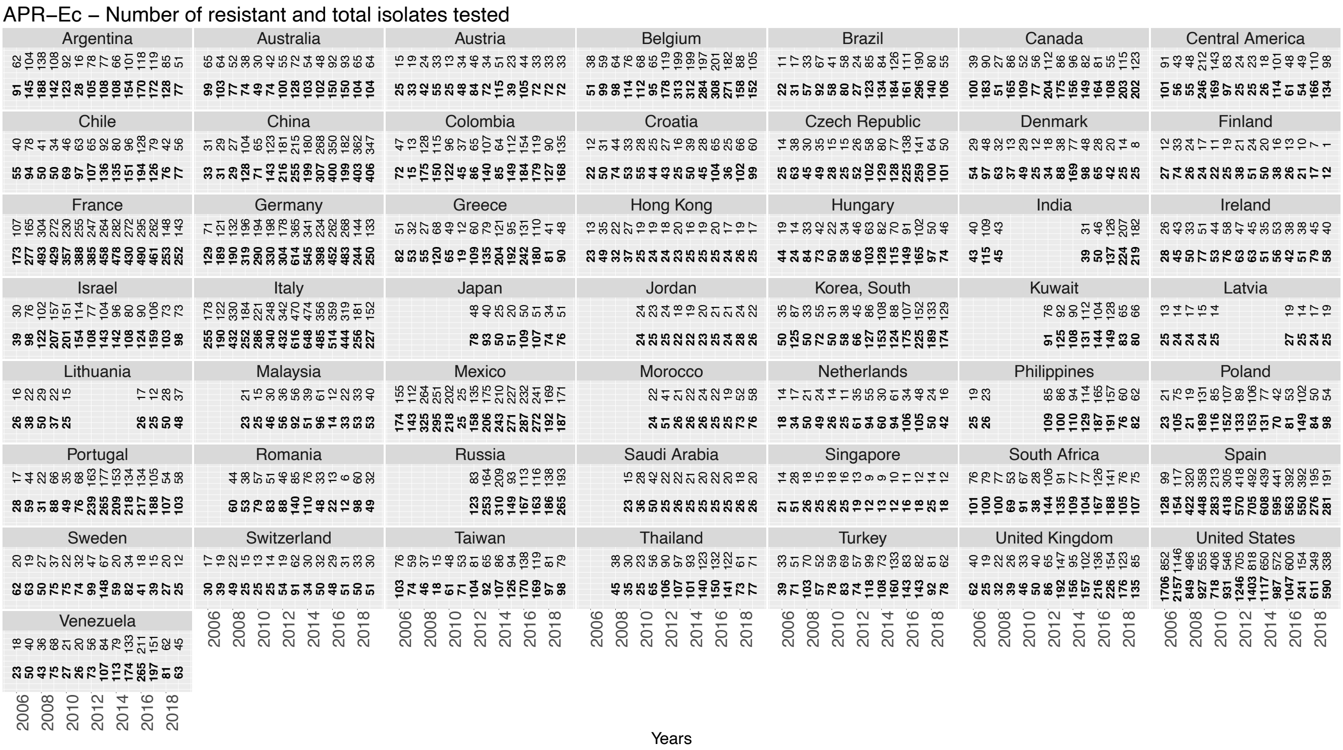
APR–Ec – Number of resistant and total isolates tested
Years

C. **3GCR-*Ec***

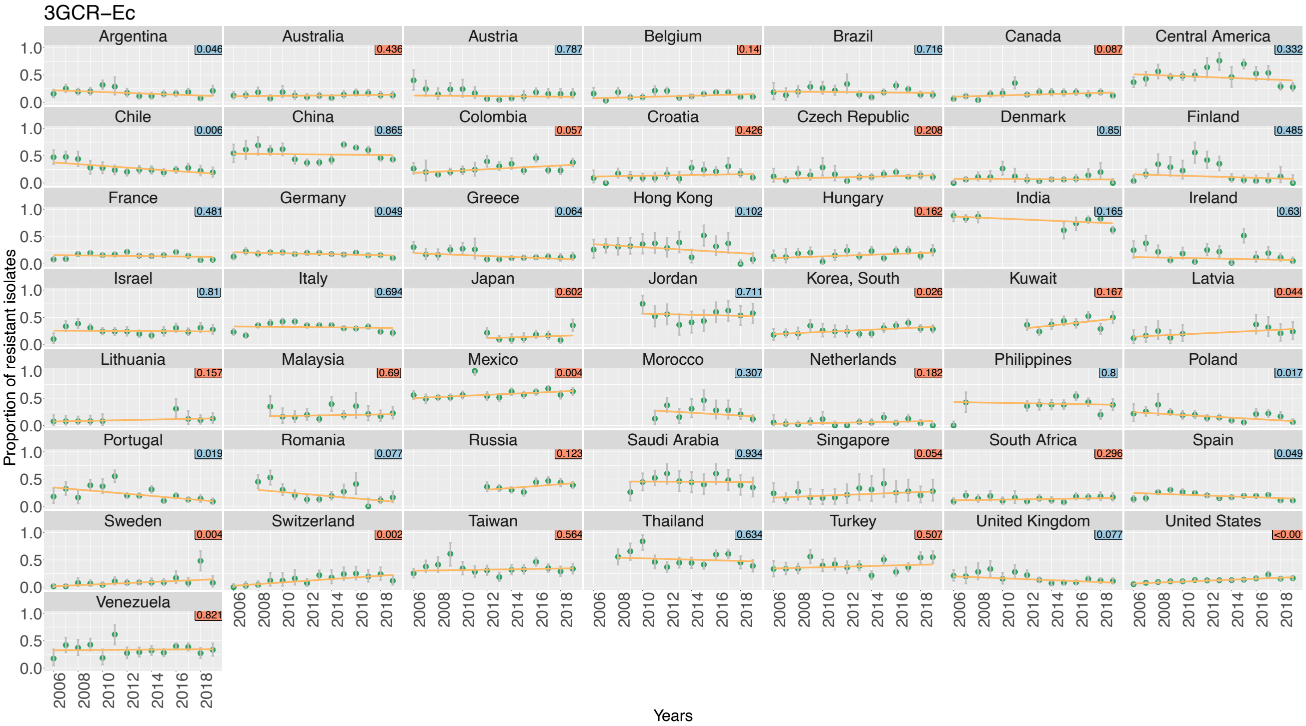
3GCR−Ec
Argentina
Australia
Austria
Belgium
Brazil
Canada
Central America
Chile
China
Colombia
Croatia
Czech Republic
Denmark
Finland
France
Germany
Greece
Hong Kong
Hungary
India
Ireland
Israel
Italy
Japan
Jordan
Korea, South
Kuwait
Latvia
Lithuania
Malaysia
Mexico
Morocco
Netherlands
Philippines
Poland
Portugal
Romania
Russia
Saudi Arabia
Singapore
South Africa
Spain
Sweden
Switzerland
Taiwan
Thailand
Turkey
United Kingdom
United States
Venezuela
Proportion of resistant isolates
Years

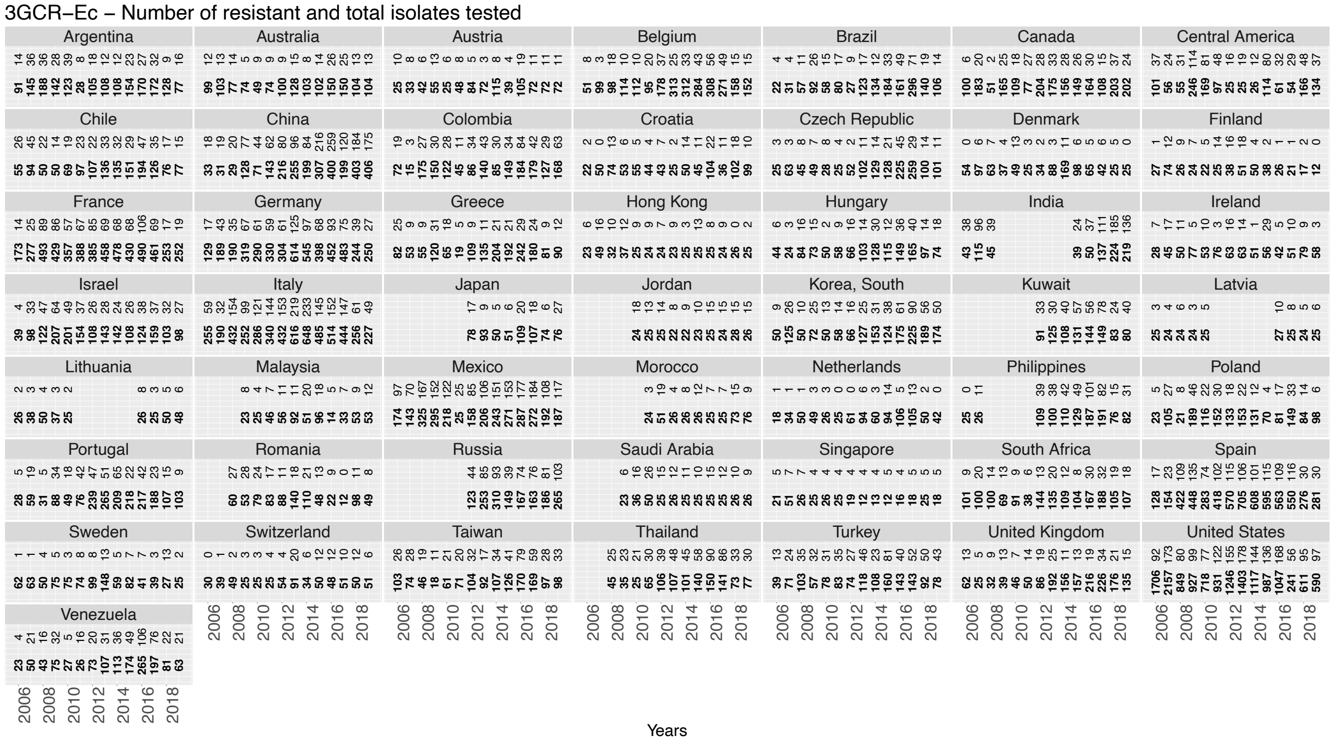
3GCR−Ec – Number of resistant and total isolates tested
Argentina
Australia
Austria
Belgium
Brazil
Canada
Central America
Chile
China
Colombia
Croatia
Czech Republic
Denmark
Finland
France
Germany
Greece
Hong Kong
Hungary
India
Ireland
Israel
Italy
Japan
Jordan
Korea, South
Kuwait
Latvia
Lithuania
Malaysia
Mexico
Morocco
Netherlands
Philippines
Poland
Portugal
Romania
Russia
Saudi Arabia
Singapore
South Africa
Spain
Sweden
Switzerland
Taiwan
Thailand
Turkey
United Kingdom
United States
Venezuela
Years

## D. **FR-*Kp***

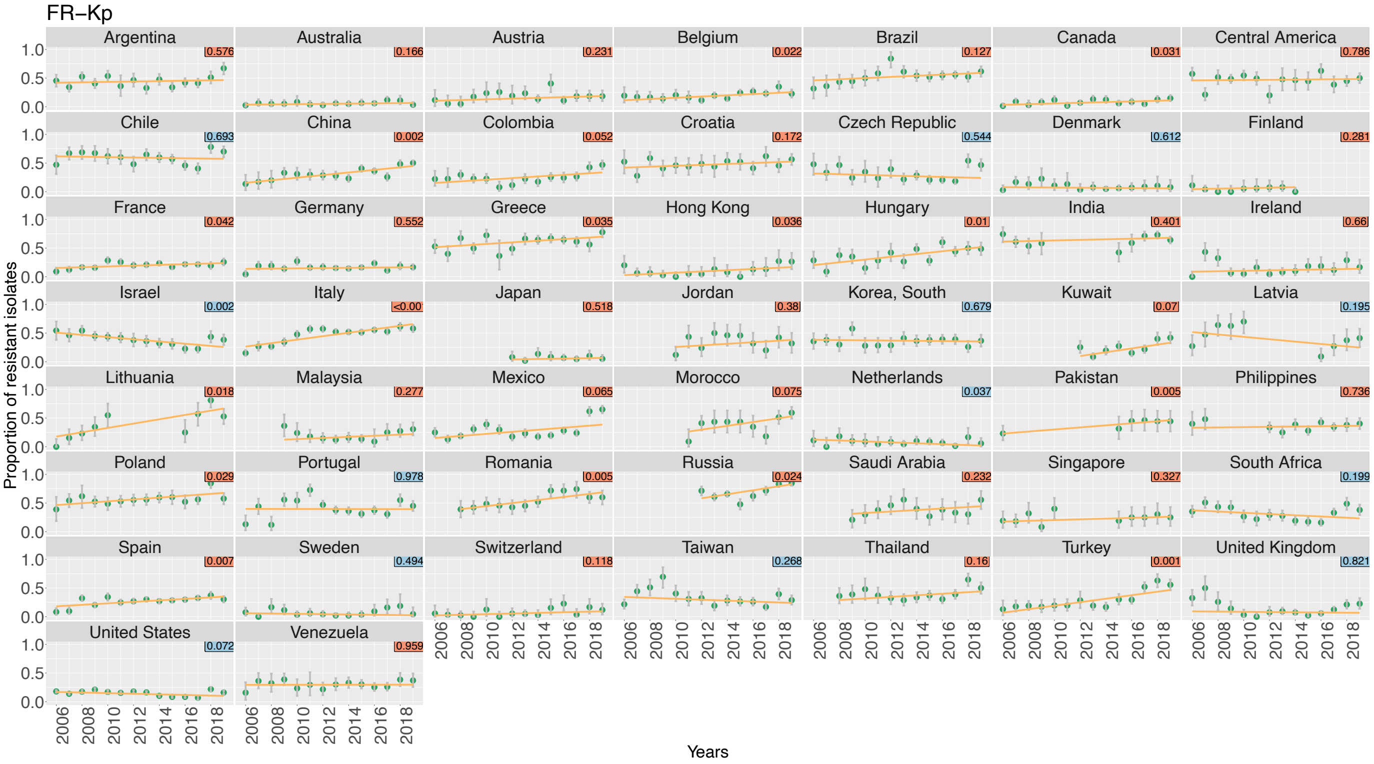
FR−Kp
Argentina
Australia
Austria
Belgium
Brazil
Canada
Central America
Chile
China
Colombia
Croatia
Czech Republic
Denmark
Finland
France
Germany
Greece
Hong Kong
Hungary
India
Ireland
Israel
Italy
Japan
Jordan
Korea, South
Kuwait
Latvia
Lithuania
Malaysia
Mexico
Morocco
Netherlands
Pakistan
Philippines
Poland
Portugal
Romania
Russia
Saudi Arabia
Singapore
South Africa
Spain
Sweden
Switzerland
Taiwan
Thailand
Turkey
United Kingdom
United States
Venezuela
Proportion of resistant isolates
Years

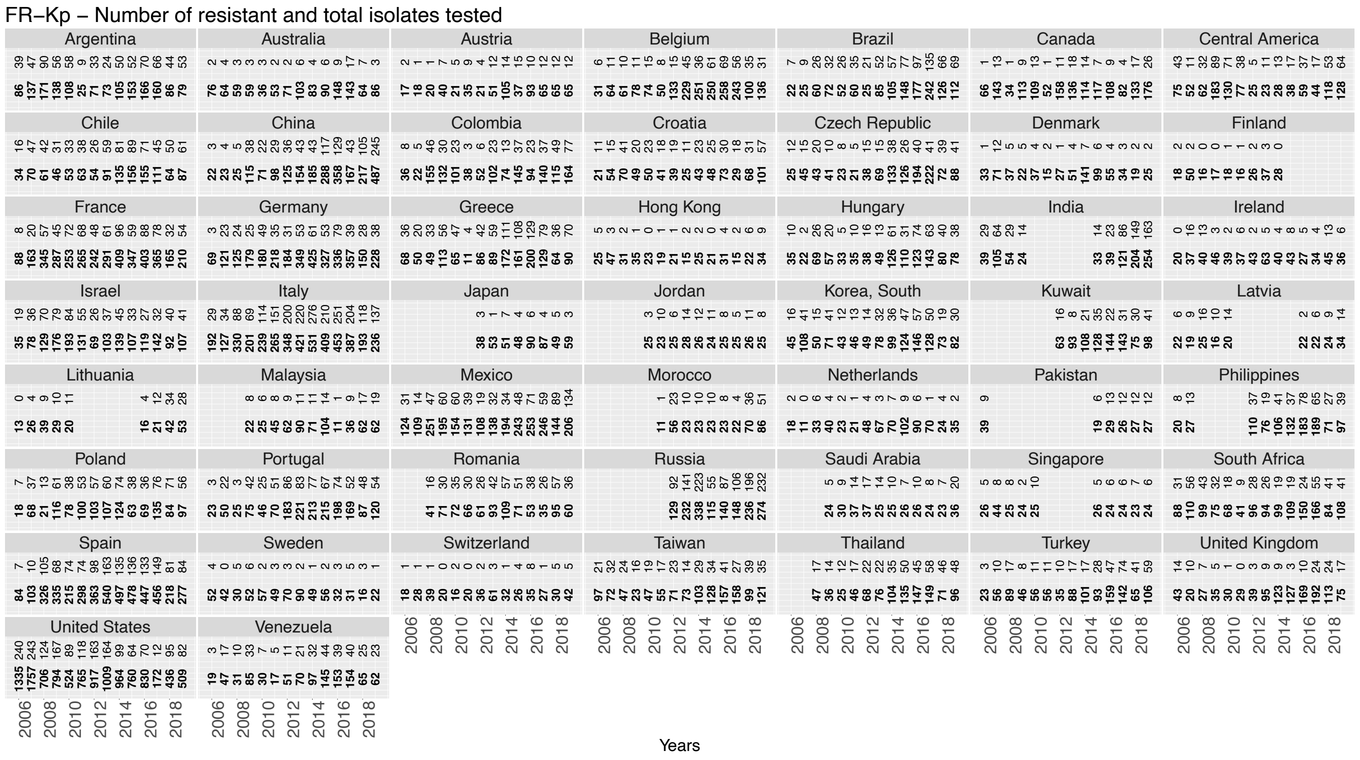
FR−Kp – Number of resistant and total isolates tested
Years

#### E. **3GCR-*Kp***

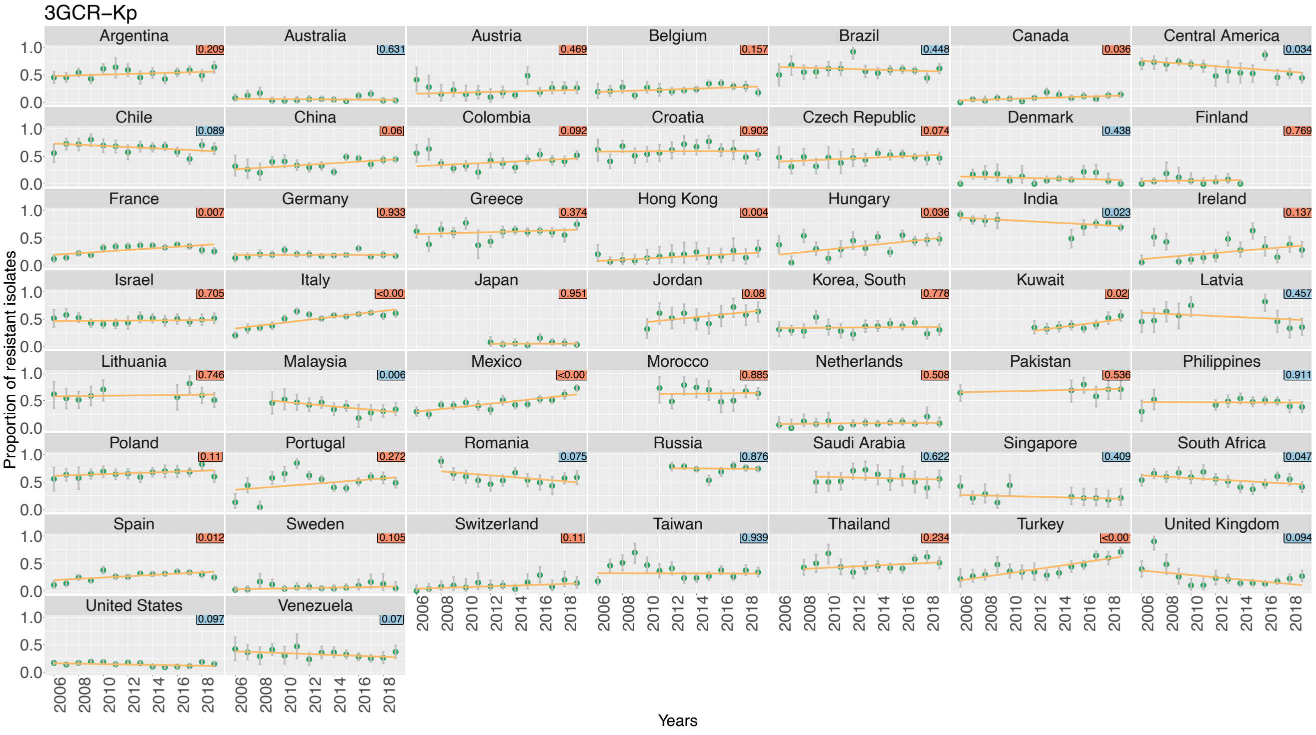
3GCR−Kp
Argentina
Australia
Austria
Belgium
Brazil
Canada
Central America
Chile
China
Colombia
Croatia
Czech Republic
Denmark
Finland
France
Germany
Greece
Hong Kong
Hungary
India
Ireland
Israel
Italy
Japan
Jordan
Korea, South
Kuwait
Latvia
Lithuania
Malaysia
Mexico
Morocco
Netherlands
Pakistan
Philippines
Poland
Portugal
Romania
Russia
Saudi Arabia
Singapore
South Africa
Spain
Sweden
Switzerland
Taiwan
Thailand
Turkey
United Kingdom
United States
Venezuela
Proportion of resistant isolates
Years

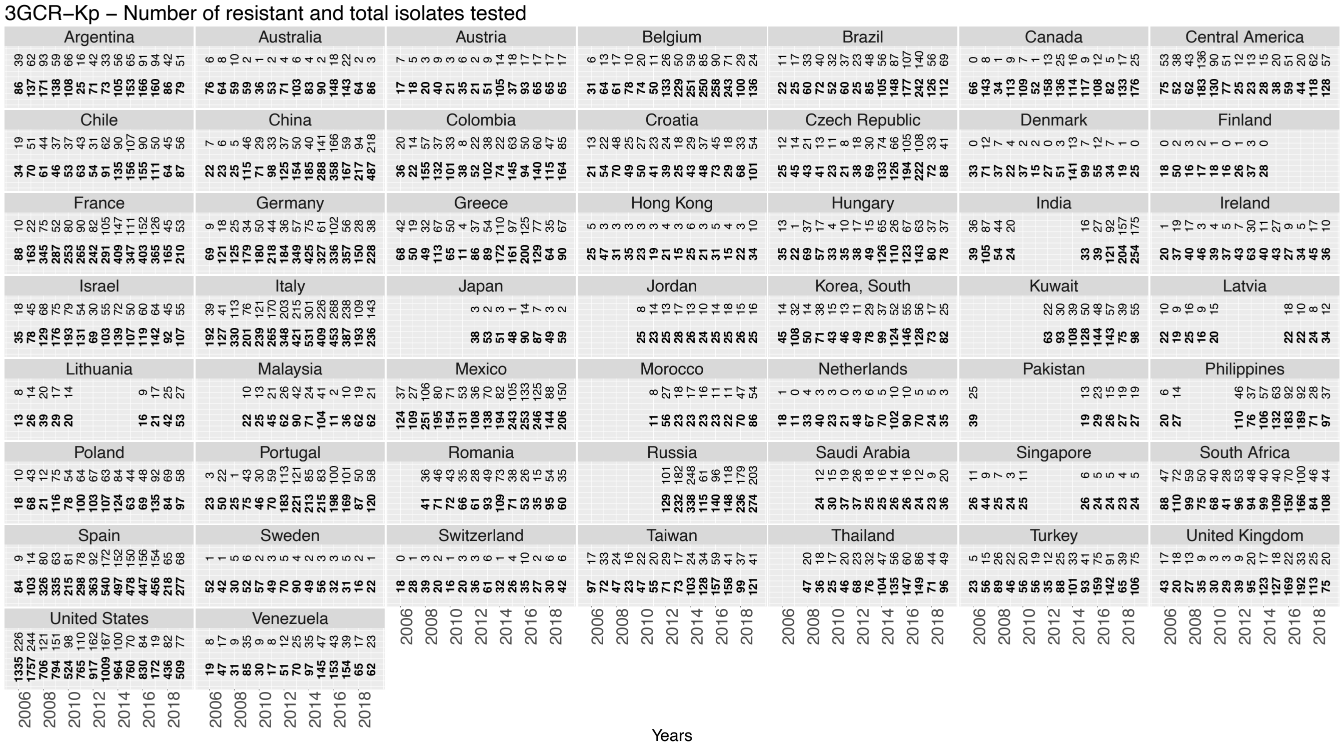
3GCR−Kp − Number of resistant and total isolates tested
Years

# F. CR-*Kp*

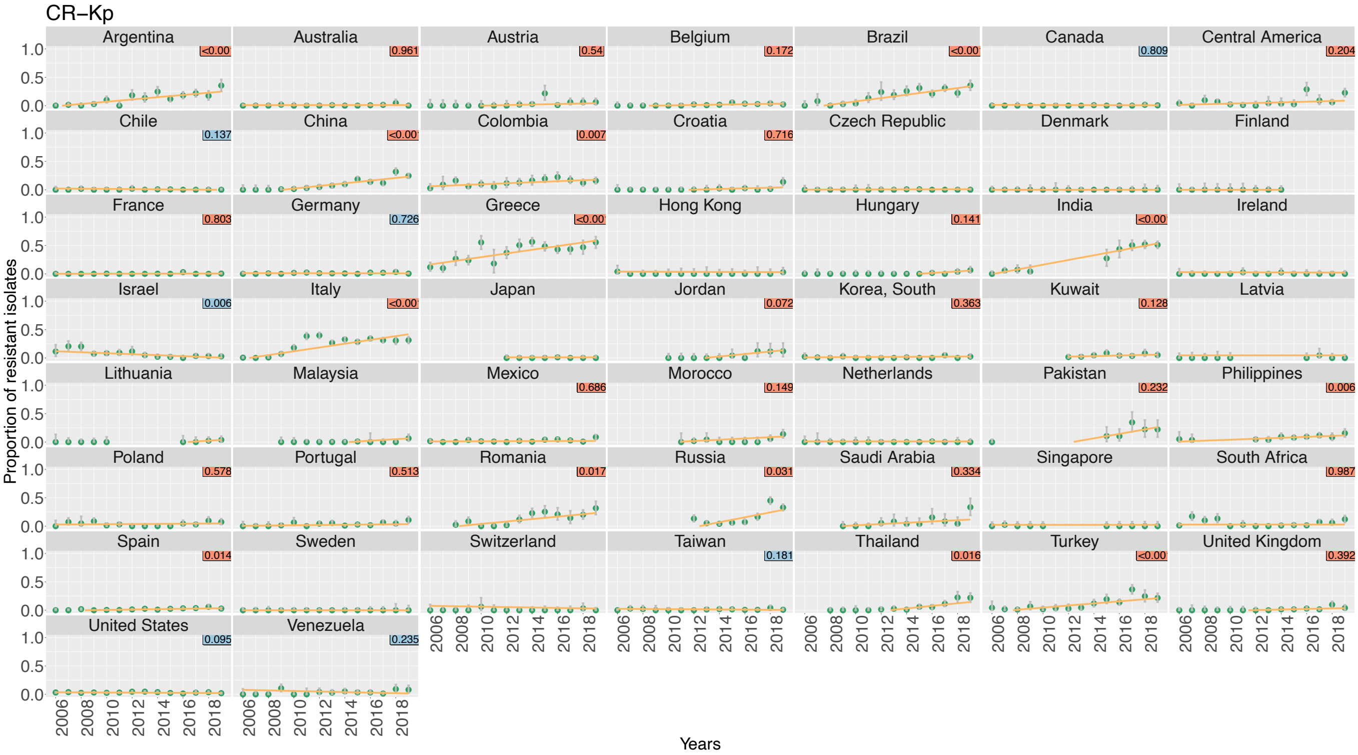
CR–Kp
Proportion of resistant isolates
Years
Argentina
Australia
Austria
Belgium
Brazil
Canada
Central America
Chile
China
Colombia
Croatia
Czech Republic
Denmark
Finland
France
Germany
Greece
Hong Kong
Hungary
India
Ireland
Israel
Italy
Japan
Jordan
Korea, South
Kuwait
Latvia
Lithuania
Malaysia
Mexico
Morocco
Netherlands
Pakistan
Philippines
Poland
Portugal
Romania
Russia
Saudi Arabia
Singapore
South Africa
Spain
Sweden
Switzerland
Taiwan
Thailand
Turkey
United Kingdom
United States
Venezuela

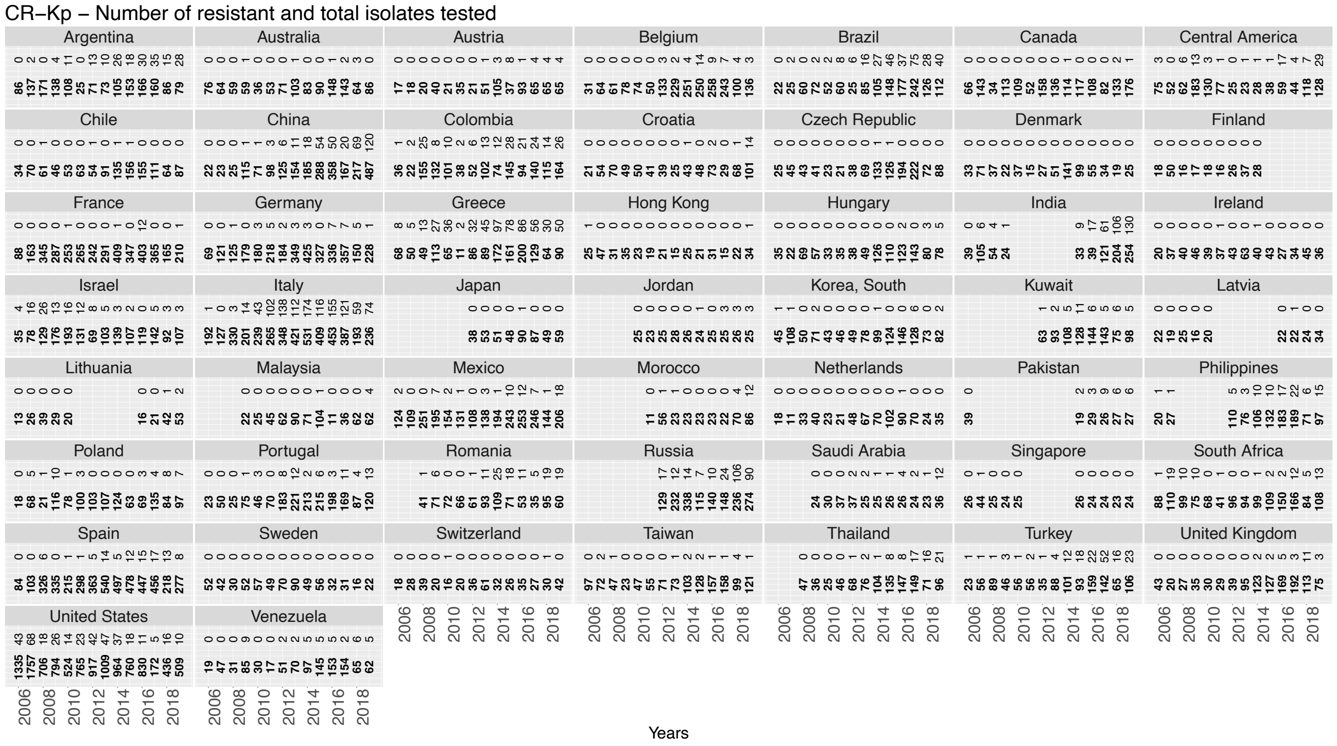
CR–Kp – Number of resistant and total isolates tested
Years

## G. **FR-*Pa***

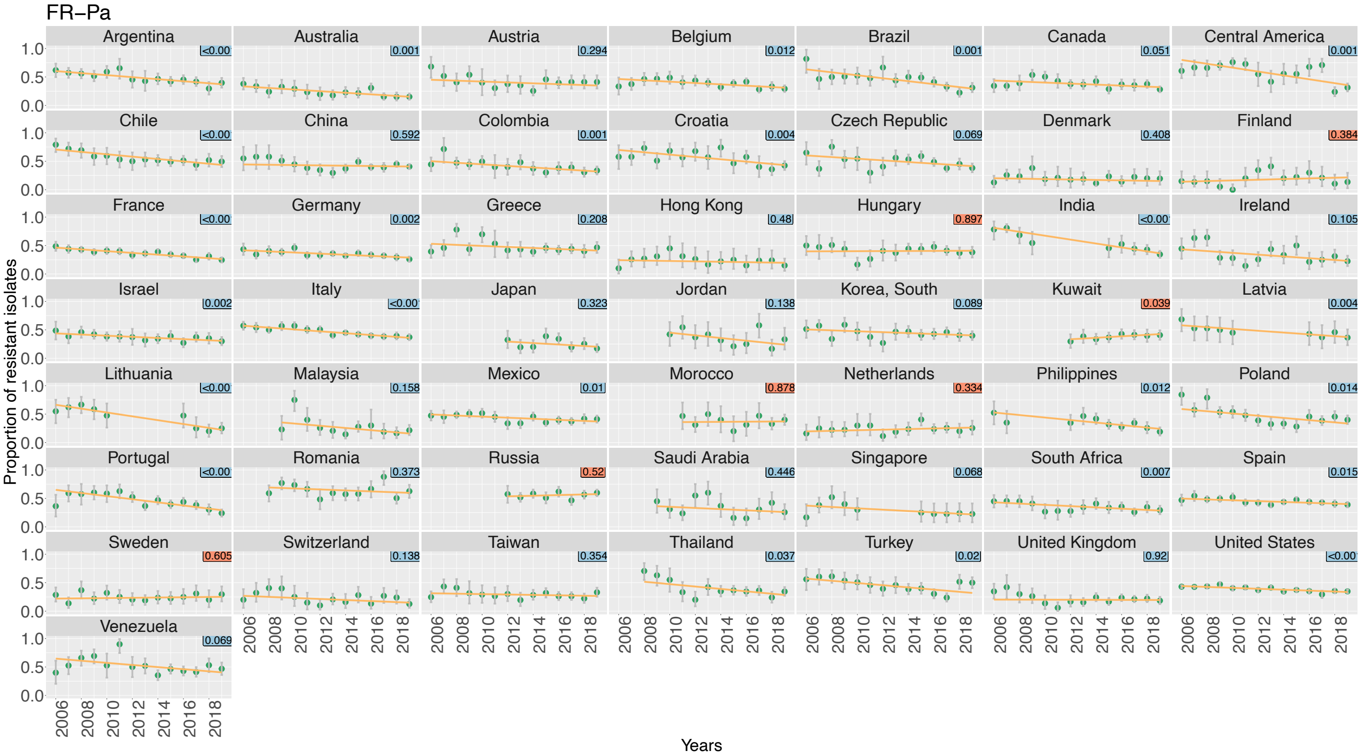
FR–Pa
Argentina
Australia
Austria
Belgium
Brazil
Canada
Central America
Chile
China
Colombia
Croatia
Czech Republic
Denmark
Finland
France
Germany
Greece
Hong Kong
Hungary
India
Ireland
Israel
Italy
Japan
Jordan
Korea, South
Kuwait
Latvia
Lithuania
Malaysia
Mexico
Morocco
Netherlands
Philippines
Poland
Portugal
Romania
Russia
Saudi Arabia
Singapore
South Africa
Spain
Sweden
Switzerland
Taiwan
Thailand
Turkey
United Kingdom
United States
Venezuela
Proportion of resistant isolates
Years

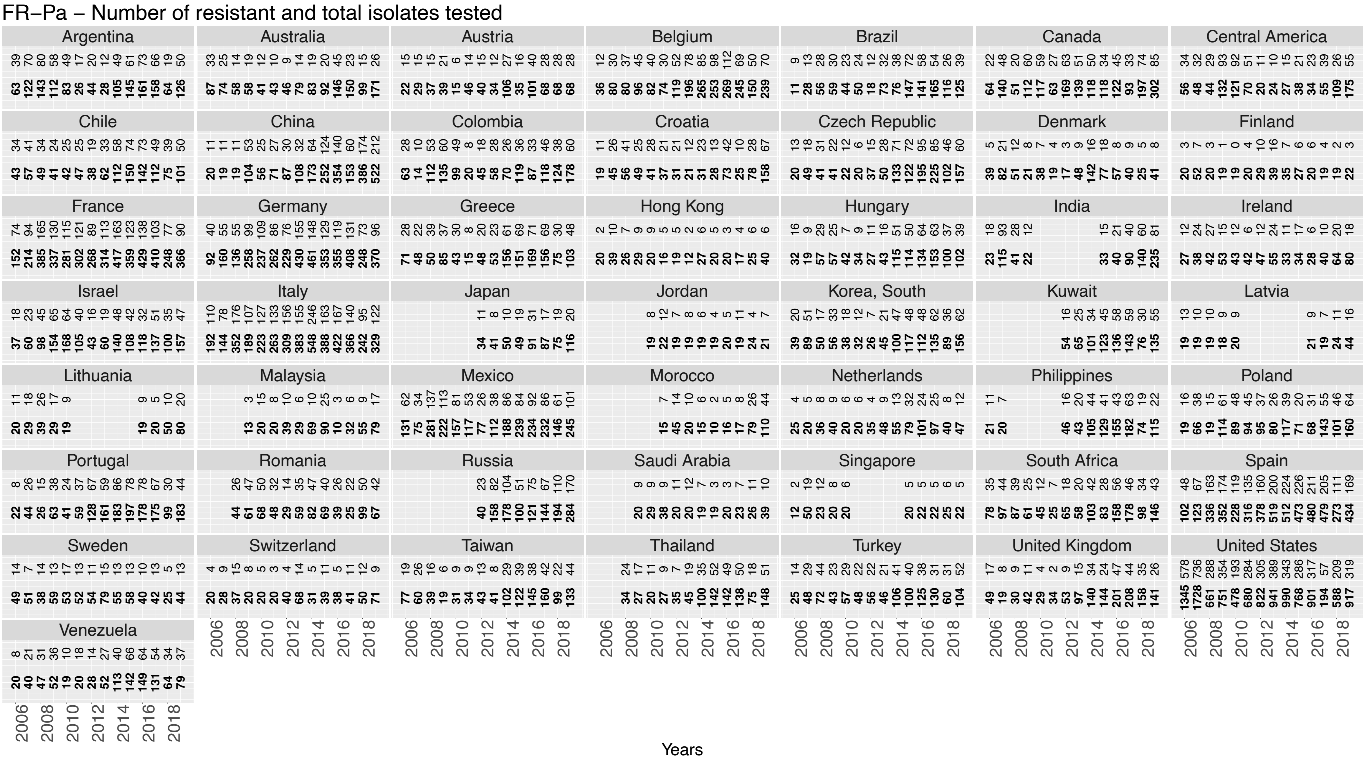
FR–Pa – Number of resistant and total isolates tested
Argentina
Australia
Austria
Belgium
Brazil
Canada
Central America
Chile
China
Colombia
Croatia
Czech Republic
Denmark
Finland
France
Germany
Greece
Hong Kong
Hungary
India
Ireland
Israel
Italy
Japan
Jordan
Korea, South
Kuwait
Latvia
Lithuania
Malaysia
Mexico
Morocco
Netherlands
Philippines
Poland
Portugal
Romania
Russia
Saudi Arabia
Singapore
South Africa
Spain
Sweden
Switzerland
Taiwan
Thailand
Turkey
United Kingdom
United States
Venezuela
Years

# H. **CR-*Pa***

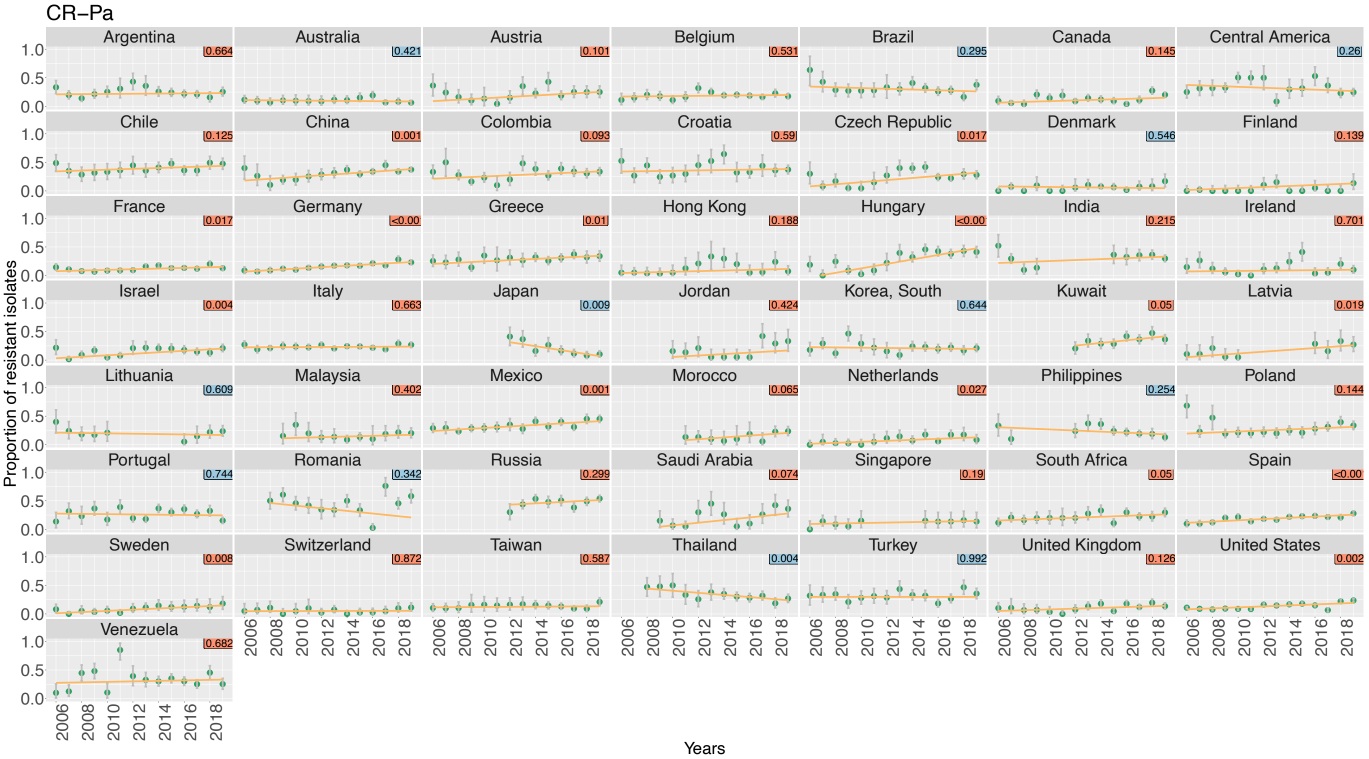
CR−Pa
Proportion of resistant isolates
Years
Argentina
Australia
Austria
Belgium
Brazil
Canada
Central America
Chile
China
Colombia
Croatia
Czech Republic
Denmark
Finland
France
Germany
Greece
Hong Kong
Hungary
India
Ireland
Israel
Italy
Japan
Jordan
Korea, South
Kuwait
Latvia
Lithuania
Malaysia
Mexico
Morocco
Netherlands
Philippines
Poland
Portugal
Romania
Russia
Saudi Arabia
Singapore
South Africa
Spain
Sweden
Switzerland
Taiwan
Thailand
Turkey
United Kingdom
United States
Venezuela

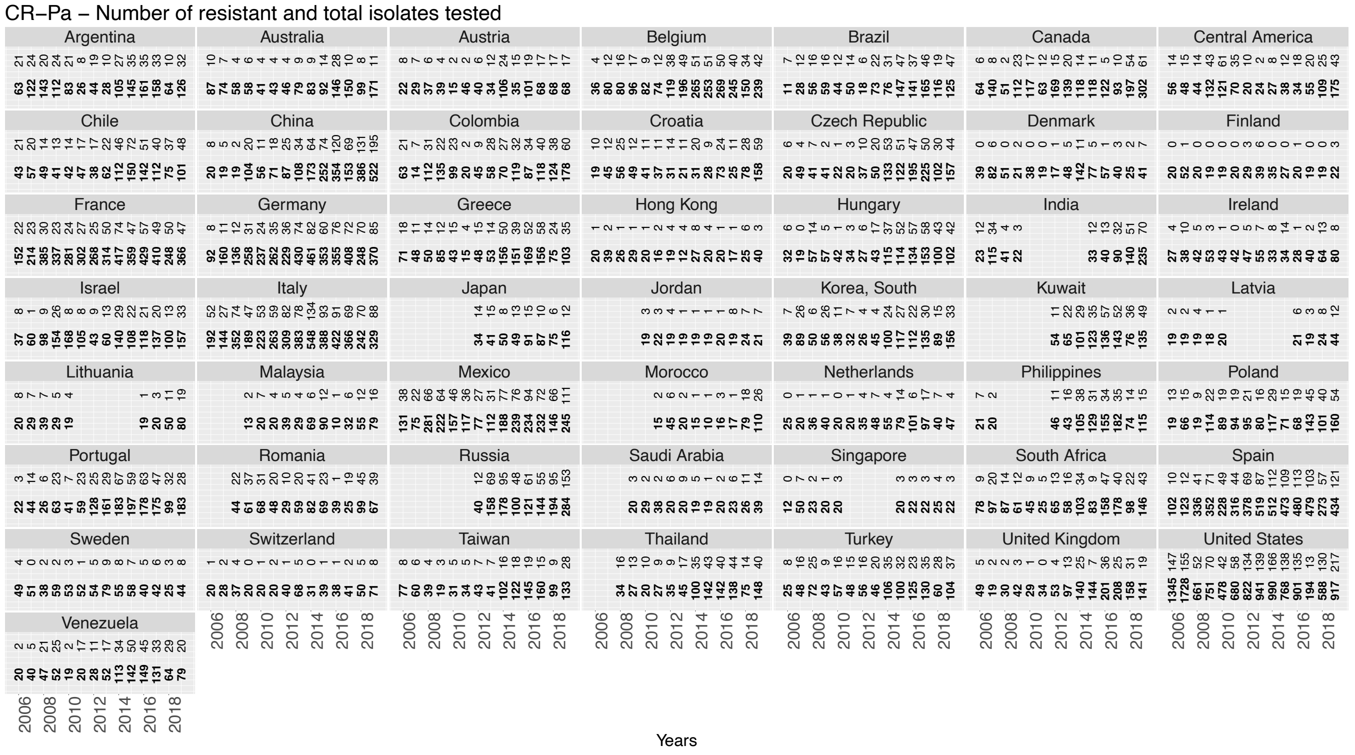
CR−Pa – Number of resistant and total isolates tested
Years

## I. FR-*Ab*

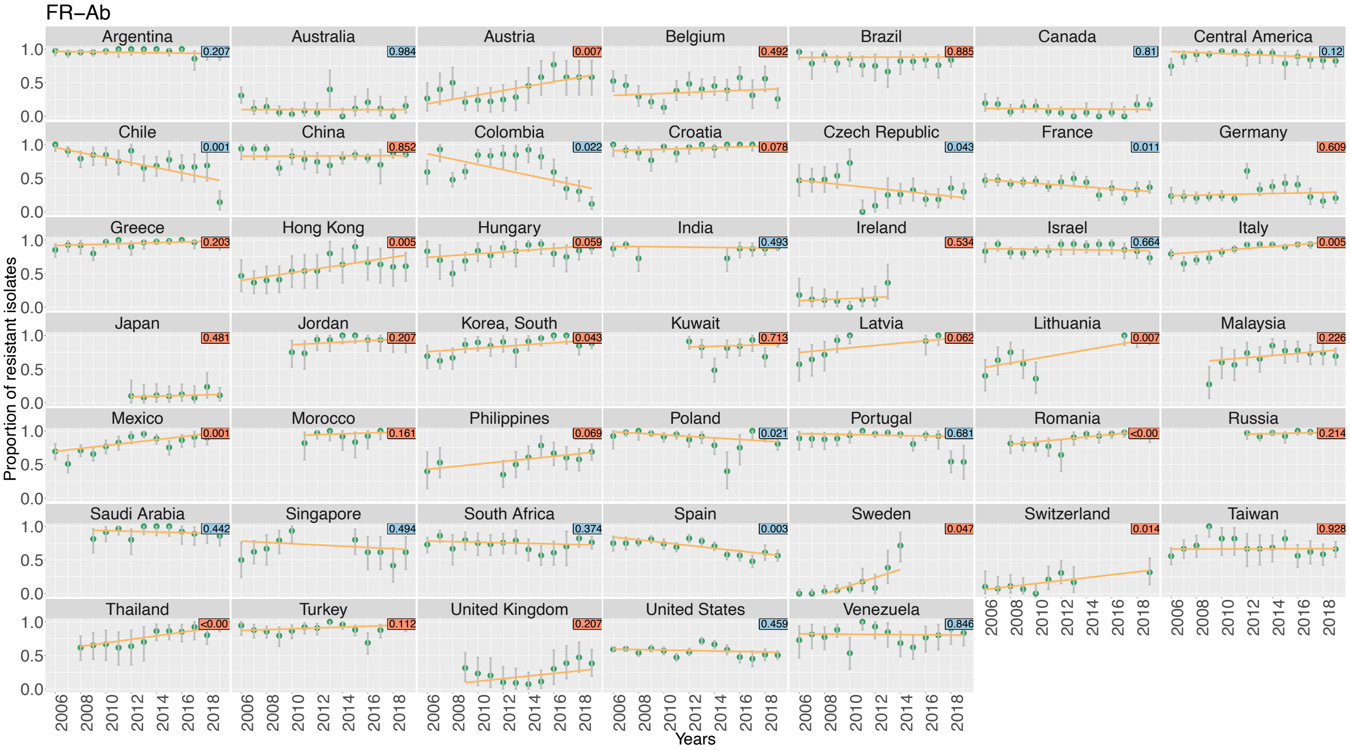
FR–Ab
Argentina
Australia
Austria
Belgium
Brazil
Canada
Central America
Chile
China
Colombia
Croatia
Czech Republic
France
Germany
Greece
Hong Kong
Hungary
India
Ireland
Israel
Italy
Japan
Jordan
Korea, South
Kuwait
Latvia
Lithuania
Malaysia
Mexico
Morocco
Philippines
Poland
Portugal
Romania
Russia
Saudi Arabia
Singapore
South Africa
Spain
Sweden
Switzerland
Taiwan
Thailand
Turkey
United Kingdom
United States
Venezuela
Proportion of resistant isolates
Years

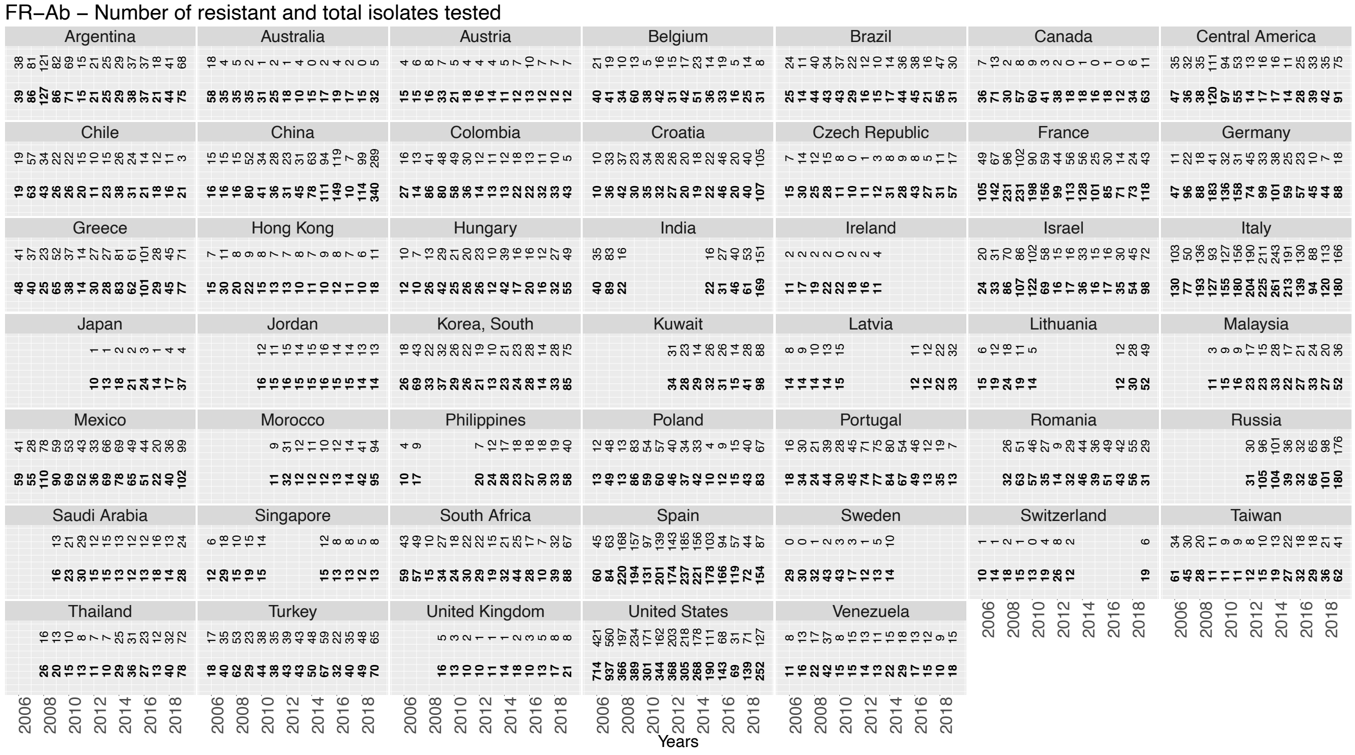
FR–Ab – Number of resistant and total isolates tested
Argentina
Australia
Austria
Belgium
Brazil
Canada
Central America
Chile
China
Colombia
Croatia
Czech Republic
France
Germany
Greece
Hong Kong
Hungary
India
Ireland
Israel
Italy
Japan
Jordan
Korea, South
Kuwait
Latvia
Lithuania
Malaysia
Mexico
Morocco
Philippines
Poland
Portugal
Romania
Russia
Saudi Arabia
Singapore
South Africa
Spain
Sweden
Switzerland
Taiwan
Thailand
Turkey
United Kingdom
United States
Venezuela
Years

## J. CR-*Ab*

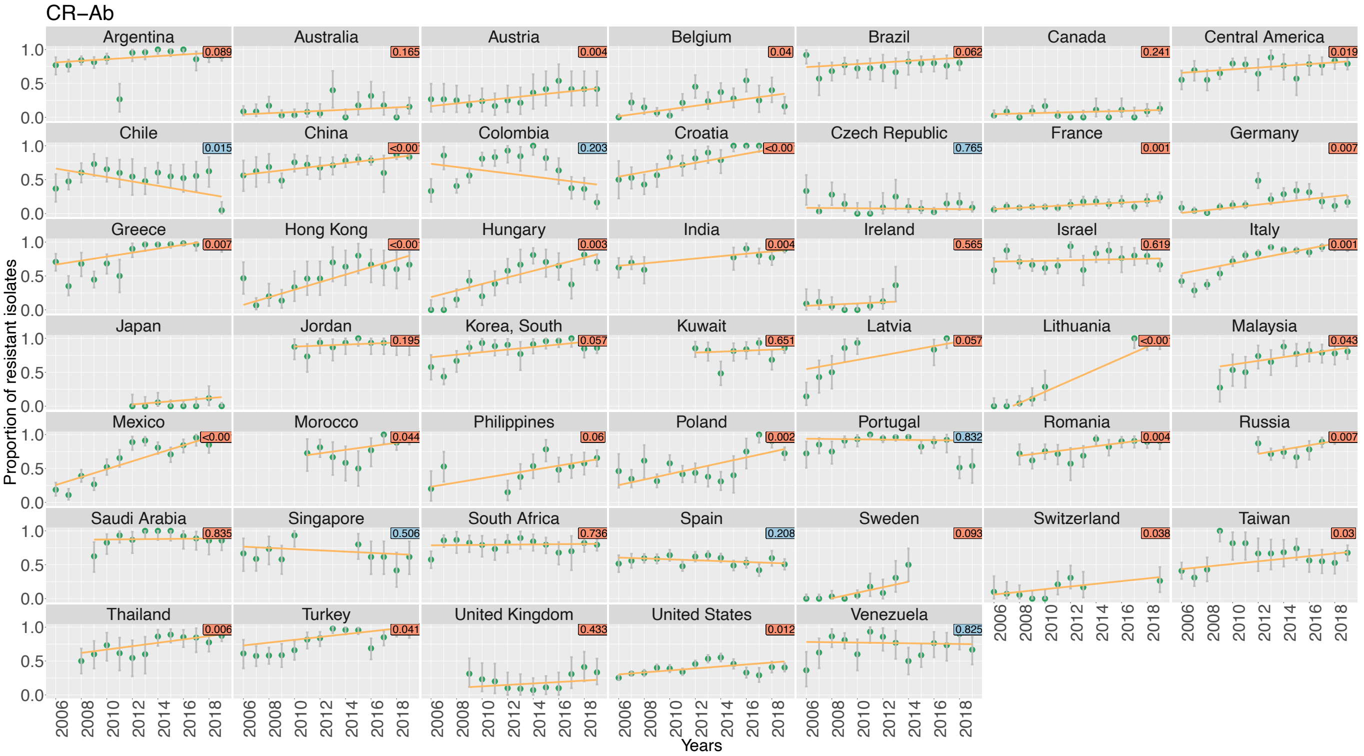
CR–Ab
Argentina
Australia
Austria
Belgium
Brazil
Canada
Central America
Chile
China
Colombia
Croatia
Czech Republic
France
Germany
Greece
Hong Kong
Hungary
India
Ireland
Israel
Italy
Japan
Jordan
Korea, South
Kuwait
Latvia
Lithuania
Malaysia
Mexico
Morocco
Philippines
Poland
Portugal
Romania
Russia
Saudi Arabia
Singapore
South Africa
Spain
Sweden
Switzerland
Taiwan
Thailand
Turkey
United Kingdom
United States
Venezuela
Proportion of resistant isolates
Years

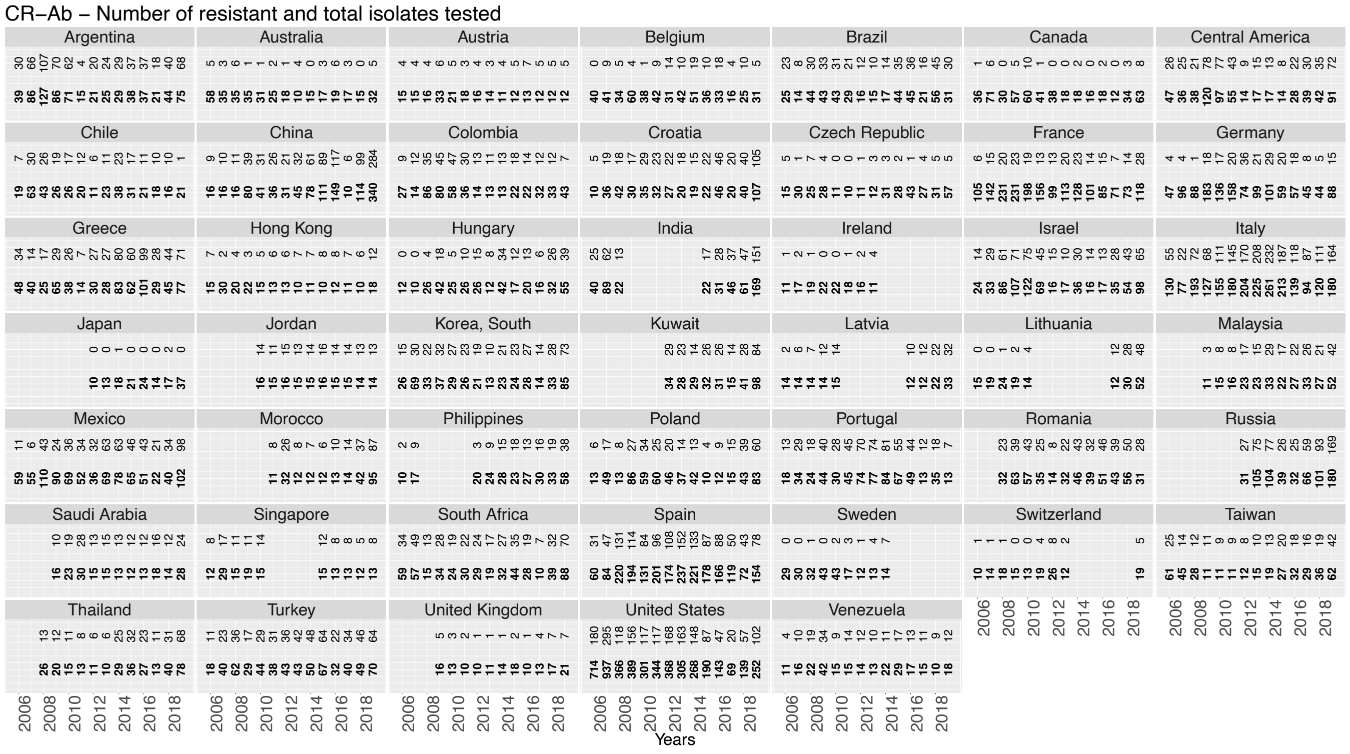
CR–Ab – Number of resistant and total isolates tested
Argentina
Australia
Austria
Belgium
Brazil
Canada
Central America
Chile
China
Colombia
Croatia
Czech Republic
France
Germany
Greece
Hong Kong
Hungary
India
Ireland
Israel
Italy
Japan
Jordan
Korea, South
Kuwait
Latvia
Lithuania
Malaysia
Mexico
Morocco
Philippines
Poland
Portugal
Romania
Russia
Saudi Arabia
Singapore
South Africa
Spain
Sweden
Switzerland
Taiwan
Thailand
Turkey
United Kingdom
United States
Venezuela
Years

## K. **VR-*E***

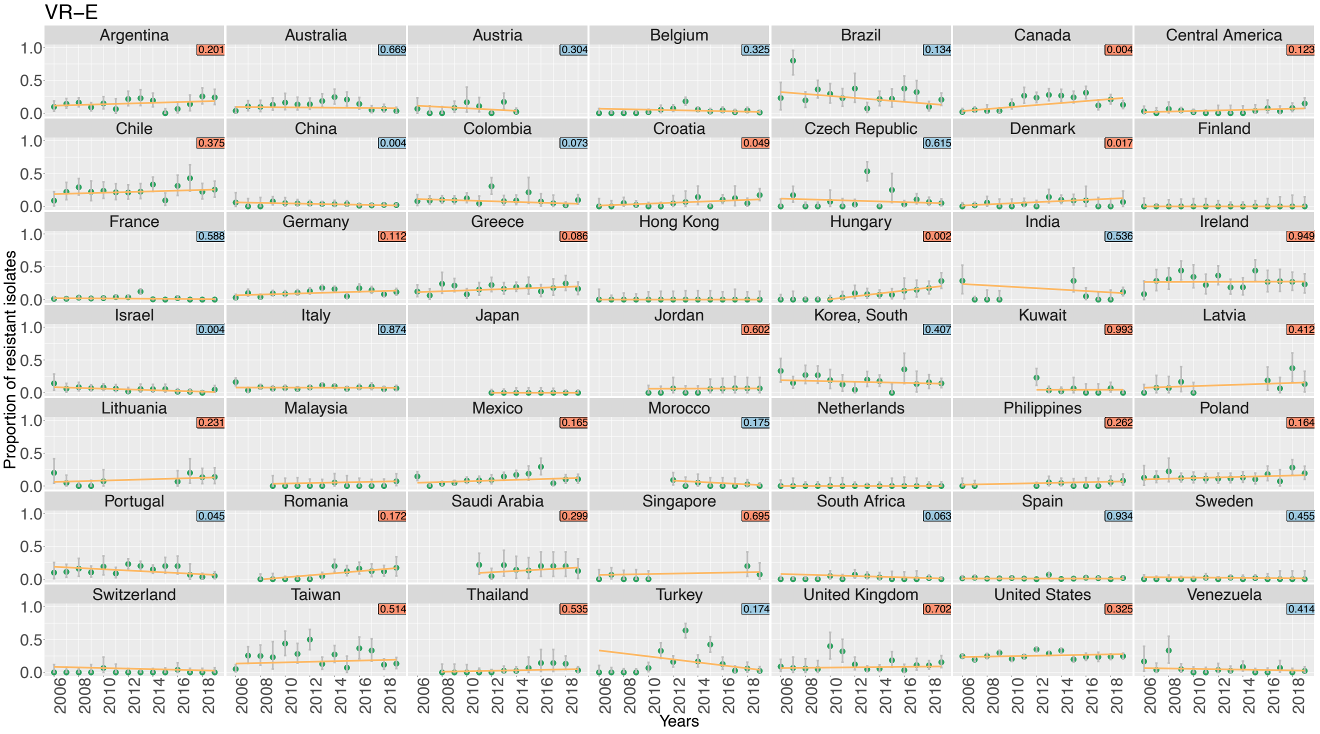
VR–E
Argentina
Australia
Austria
Belgium
Brazil
Canada
Central America
Chile
China
Colombia
Croatia
Czech Republic
Denmark
Finland
France
Germany
Greece
Hong Kong
Hungary
India
Ireland
Israel
Italy
Japan
Jordan
Korea, South
Kuwait
Latvia
Lithuania
Malaysia
Mexico
Morocco
Netherlands
Philippines
Poland
Portugal
Romania
Saudi Arabia
Singapore
South Africa
Spain
Sweden
Switzerland
Taiwan
Thailand
Turkey
United Kingdom
United States
Venezuela
Proportion of resistant isolates
Years

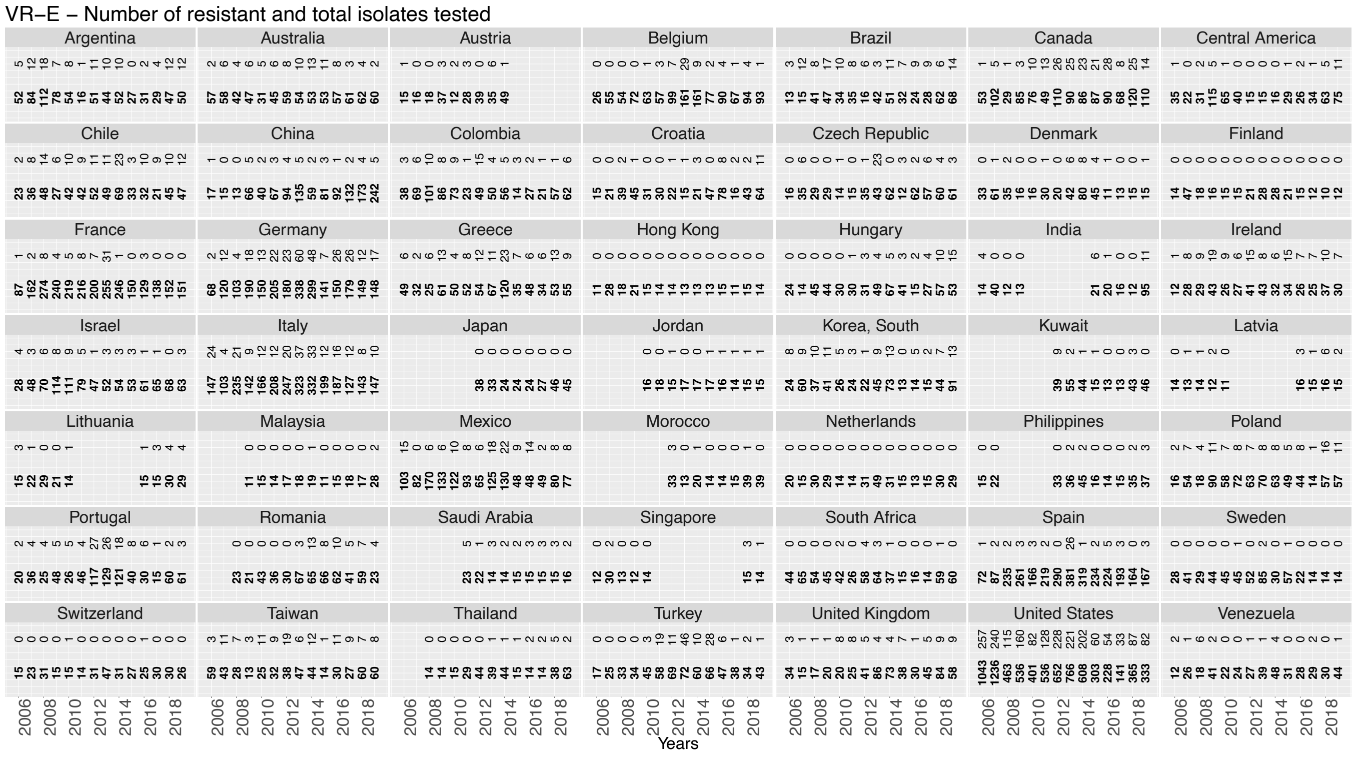
VR–E – Number of resistant and total isolates tested
Argentina
Australia
Austria
Belgium
Brazil
Canada
Central America
Chile
China
Colombia
Croatia
Czech Republic
Denmark
Finland
France
Germany
Greece
Hong Kong
Hungary
India
Ireland
Israel
Italy
Japan
Jordan
Korea, South
Kuwait
Latvia
Lithuania
Malaysia
Mexico
Morocco
Netherlands
Philippines
Poland
Portugal
Romania
Saudi Arabia
Singapore
South Africa
Spain
Sweden
Switzerland
Taiwan
Thailand
Turkey
United Kingdom
United States
Venezuela
Years

## L. **PR-*Sp***

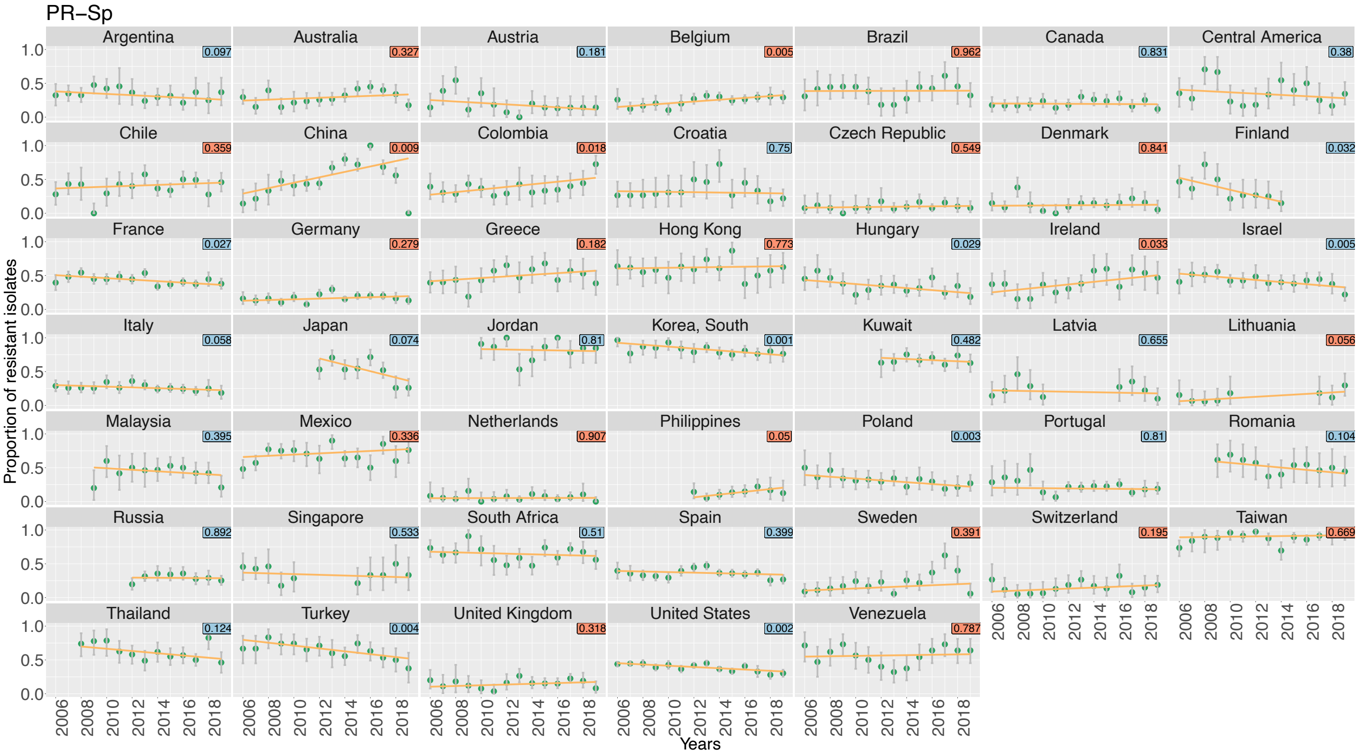
PR−Sp
Argentina 0.097
Australia 0.327
Austria 0.181
Belgium 0.005
Brazil 0.962
Canada 0.831
Central America 0.38
Chile 0.359
China 0.009
Colombia 0.018
Croatia 0.75
Czech Republic 0.549
Denmark 0.841
Finland 0.032
France 0.027
Germany 0.279
Greece 0.182
Hong Kong 0.773
Hungary 0.029
Ireland 0.033
Israel 0.005
Italy 0.058
Japan 0.074
Jordan 0.81
Korea, South 0.001
Kuwait 0.482
Latvia 0.655
Lithuania 0.056
Malaysia 0.395
Mexico 0.336
Netherlands 0.907
Philippines 0.05
Poland 0.003
Portugal 0.81
Romania 0.104
Russia 0.892
Singapore 0.533
South Africa 0.51
Spain 0.399
Sweden 0.391
Switzerland 0.195
Taiwan 0.669
Thailand 0.124
Turkey 0.004
United Kingdom 0.318
United States 0.002
Venezuela 0.787
Proportion of resistant isolates
Years

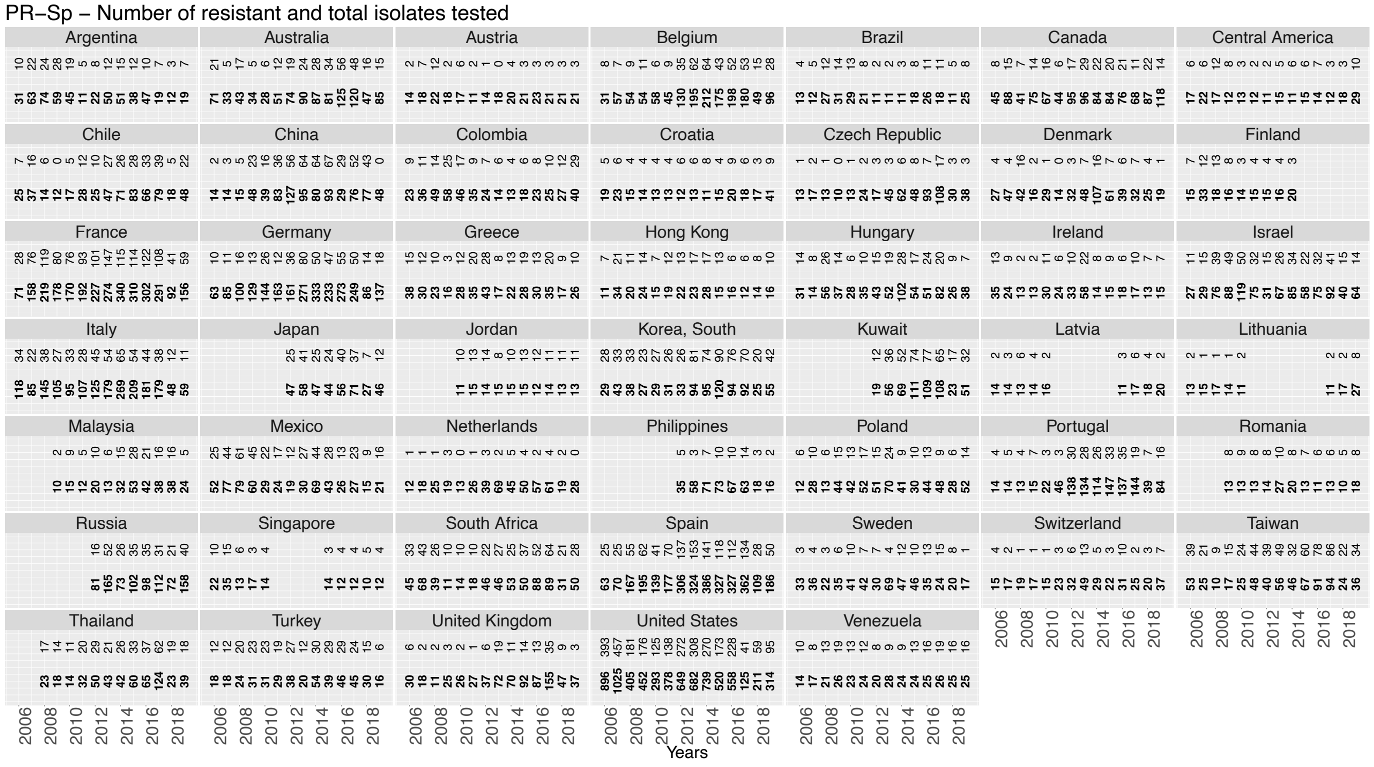
PR−Sp – Number of resistant and total isolates tested
Argentina
Australia
Austria
Belgium
Brazil
Canada
Central America
Chile
China
Colombia
Croatia
Czech Republic
Denmark
Finland
France
Germany
Greece
Hong Kong
Hungary
Ireland
Israel
Italy
Japan
Jordan
Korea, South
Kuwait
Latvia
Lithuania
Malaysia
Mexico
Netherlands
Philippines
Poland
Portugal
Romania
Russia
Singapore
South Africa
Spain
Sweden
Switzerland
Taiwan
Thailand
Turkey
United Kingdom
United States
Venezuela
Years

### M. **MLR-*Sp***

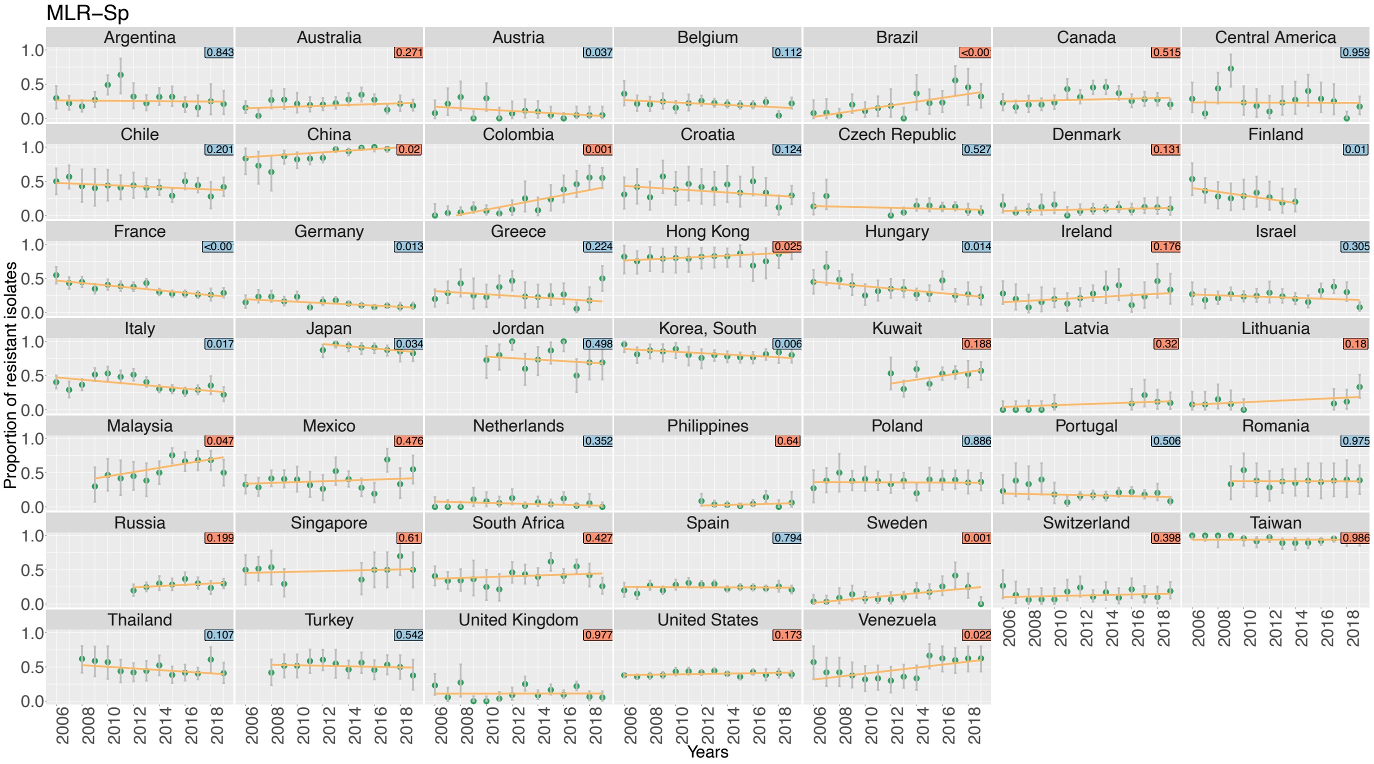
MLR–Sp
Proportion of resistant isolates
Years
Argentina
Australia
Austria
Belgium
Brazil
Canada
Central America
Chile
China
Colombia
Croatia
Czech Republic
Denmark
Finland
France
Germany
Greece
Hong Kong
Hungary
Ireland
Israel
Italy
Japan
Jordan
Korea, South
Kuwait
Latvia
Lithuania
Malaysia
Mexico
Netherlands
Philippines
Poland
Portugal
Romania
Russia
Singapore
South Africa
Spain
Sweden
Switzerland
Taiwan
Thailand
Turkey
United Kingdom
United States
Venezuela

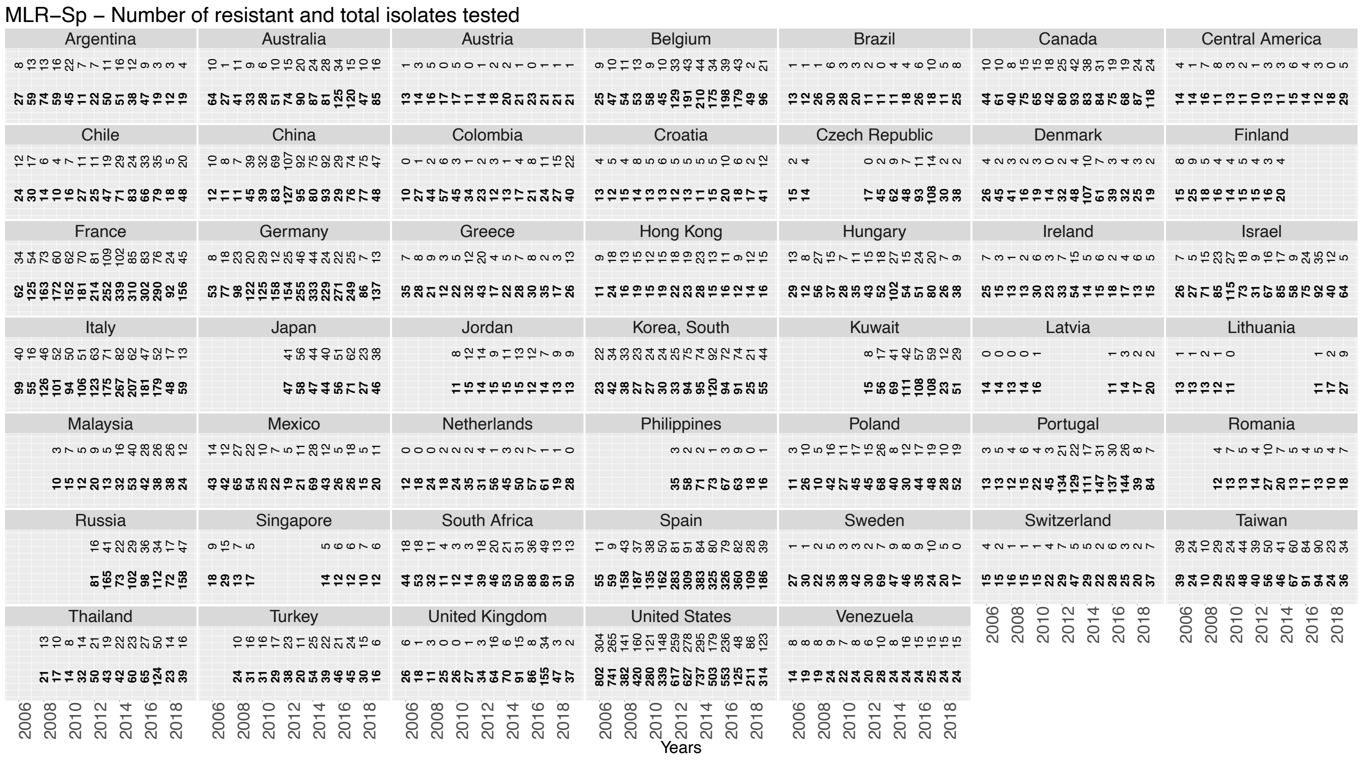
MLR–Sp – Number of resistant and total isolates tested
Years

***Supplementary Figure S9. Comparison of dendrograms for country regression slopes of ABR rates (temporal trends) by drug-bug pair, using different methods for distance computation and clustering algorithm.***

To explore possible uncertainty in the clustering, we assessed different distance metric (Euclidean, Maximum, Manhattan) and clustering algorithms (Complete, Ward, Single) and see how robust our clustering patterns was depending on the methods chosen. We show that row clustering (drug-bug pairs) is robust to methodological choices: *E. coli* pairs always clustered together (blue), and *S. pneumoniae* and *A. baumnannii* pairs clustered 5 out of 6 times and 4 out 6 times respectively (beige and brown respectively).

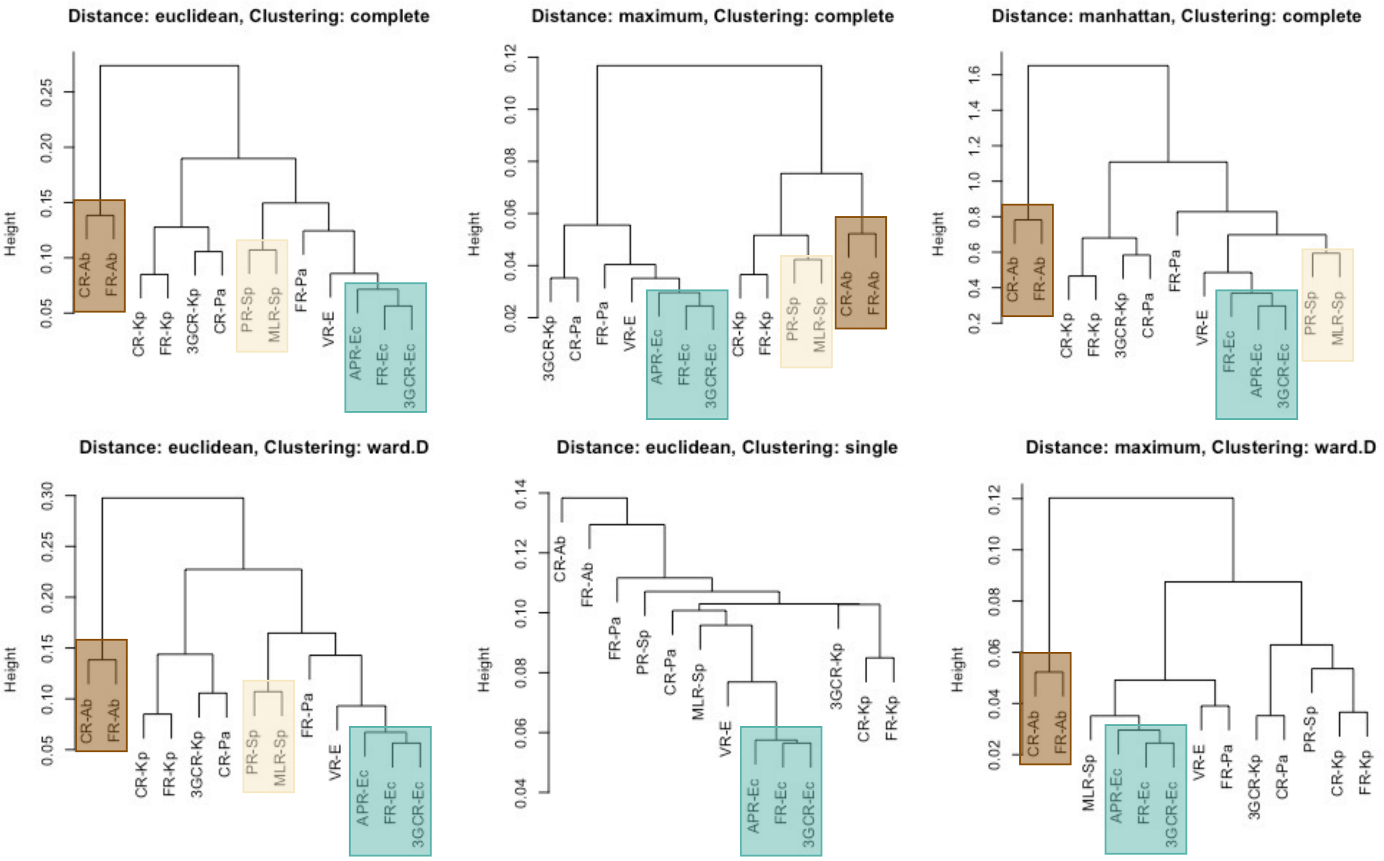

#### Supplementary model’s results

***Supplementary Figure S10. Observed vs fitted values by final multivariable models (M_multi), by drug-bug pair.***

Model fits for:

- **A. FR-Ec**: fluoroquinolone-resistant *E. coli*
- **B. APR-Ec**: aminopenicillin-resistant *E. coli*
- **C. 3GCR-Ec**: third generation cephalosporin-resistant *E. coli*
- **D. FR-Kp**: fluoroquinolone-resistant *K. pneumoniae*
- **E. 3GCR-Kp**: third generation cephalosporin-resistant *K. pneumoniae*
- **F. CR-Kp**: carbapenem-resistant *K. pneumoniae*
- **G. FR-Pa**: fluoroquinolone-resistant *P. aeruginosa*
- **H. CR-Pa**: carbapenem-resistant *P. aeruginosa*
- **I. FR-Ab**: fluoroquinolone-resistant *A. baumannii*
- **J. CR-Ab**: carbapenem-resistant *A. baumannii*
- **K. VR-E**: vancomycin-resistant Enterococci
- **L. PR-Sp**: penicillin-non-susceptible *S. pneumoniae*
- **M. MLR-Sp**: macrolide-resistant *S. pneumoniae*

Red points are the observed ABR rates by country-year with corresponding 95% confidence intervals around proportions. Black lines are the fitted values by the final multivariable models M_multi specific to each drug-bug pair.

A. **FR-*Ec***

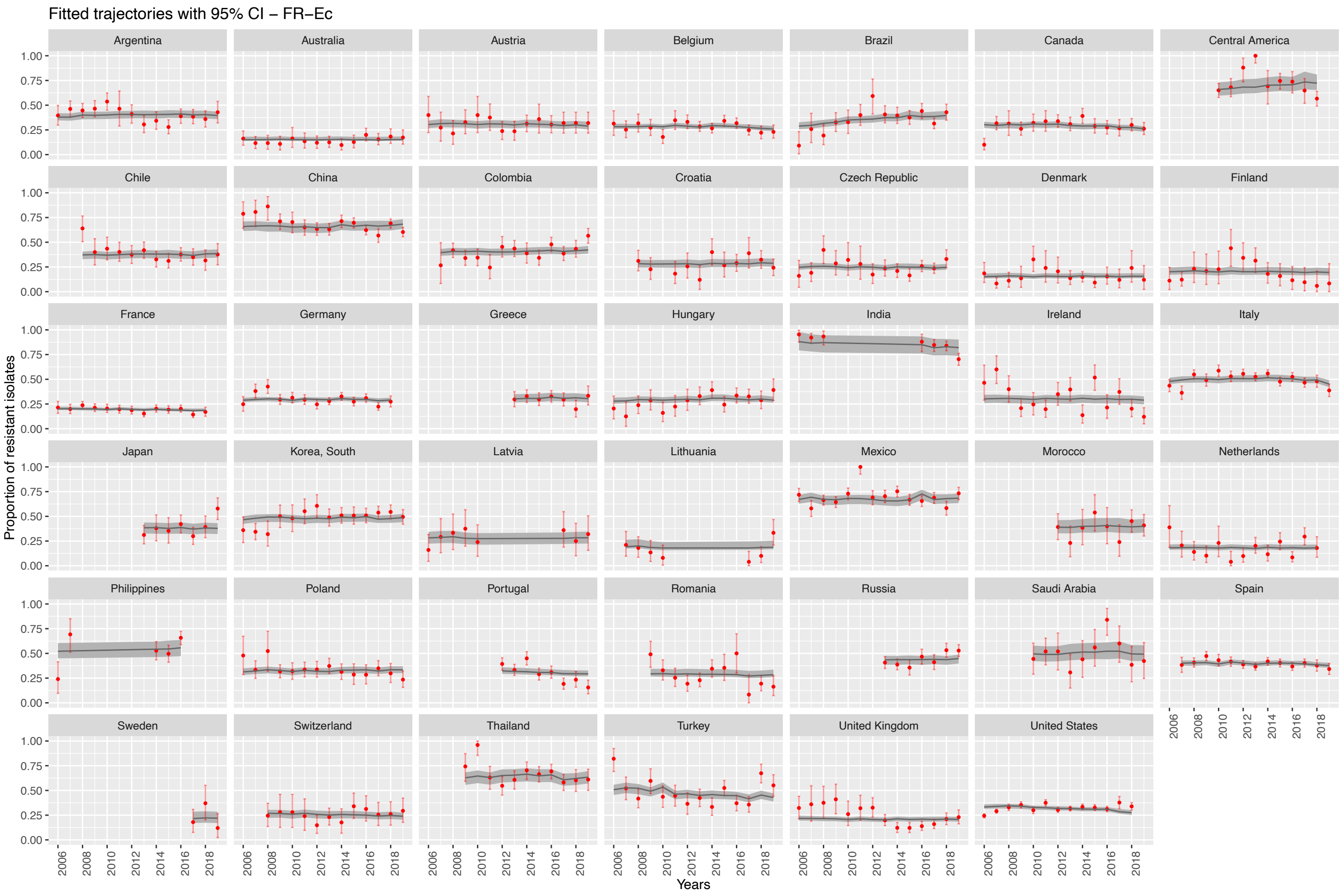

B. **APR-*Ec***

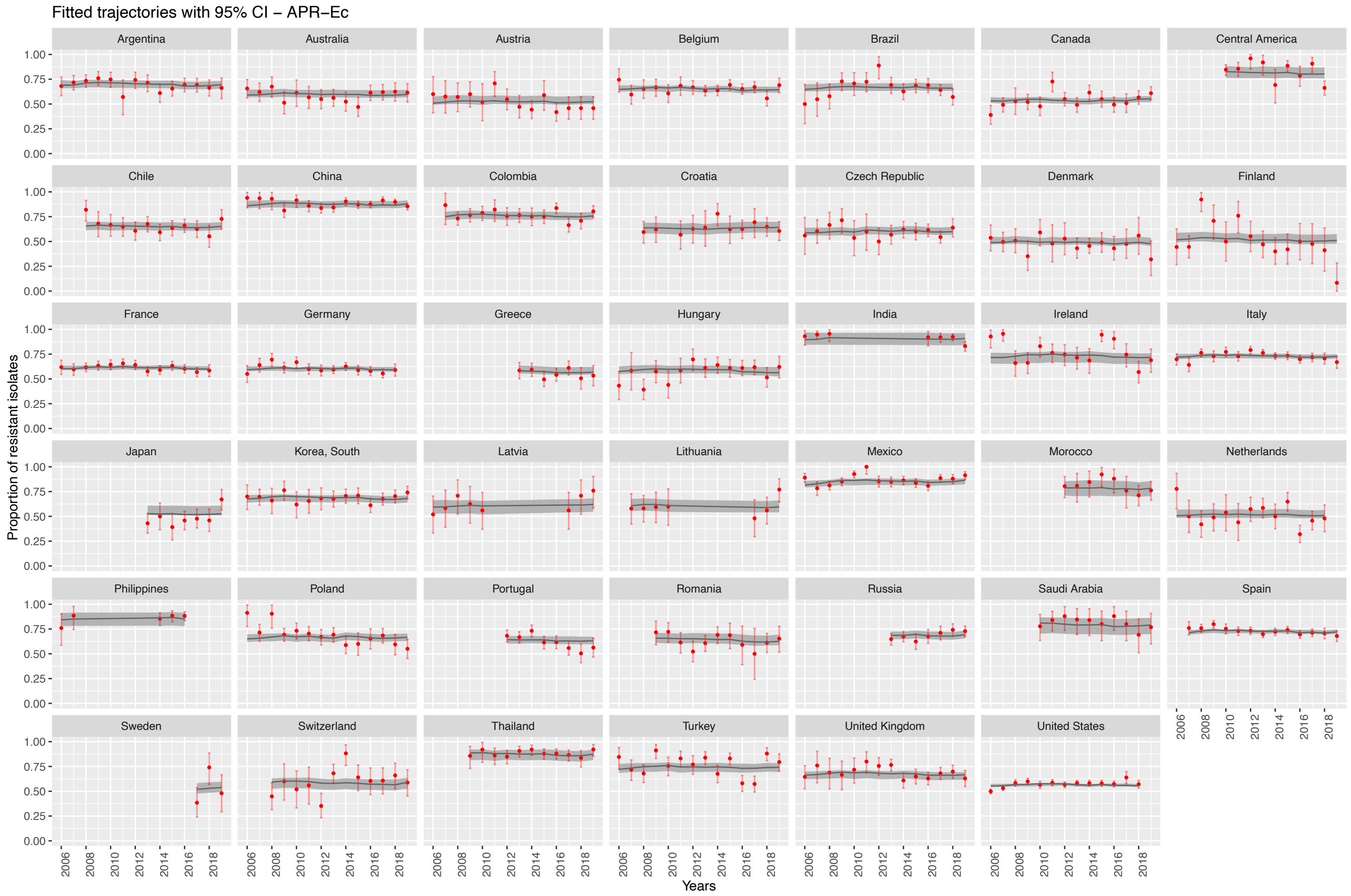

#### C. **3GCR-*Ec***

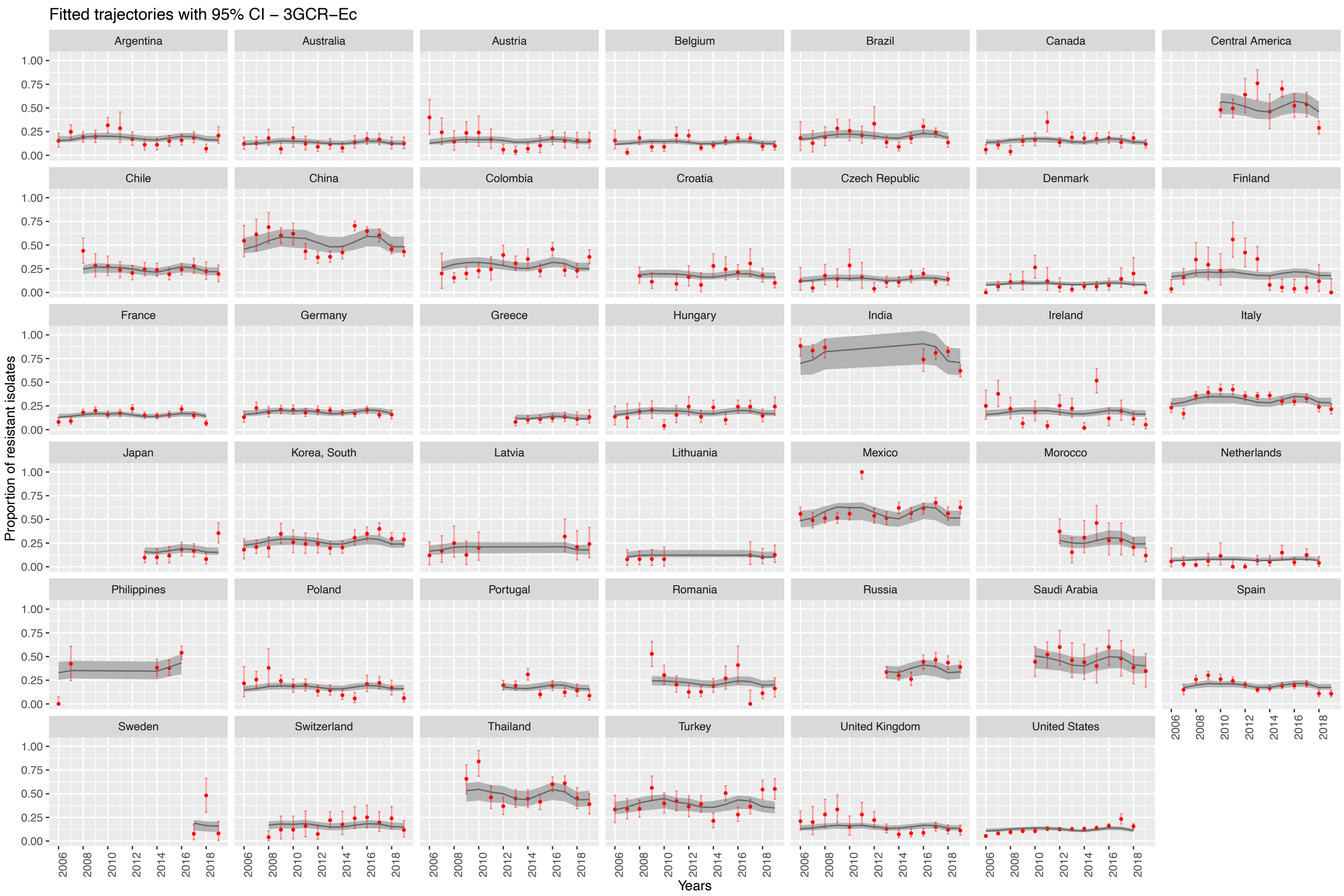

## D. **FR-*Kp***

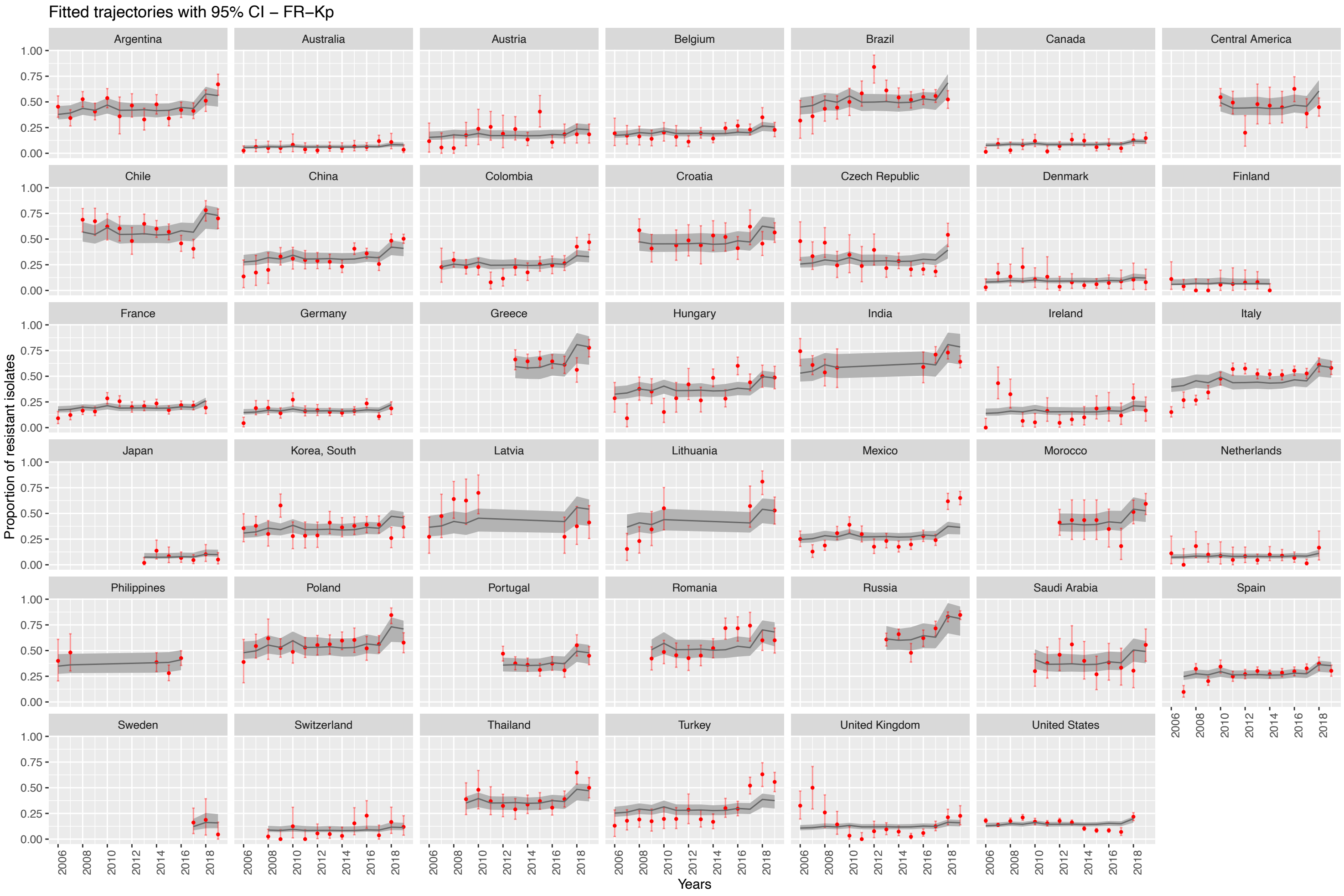

E. **3GCR-*Kp***

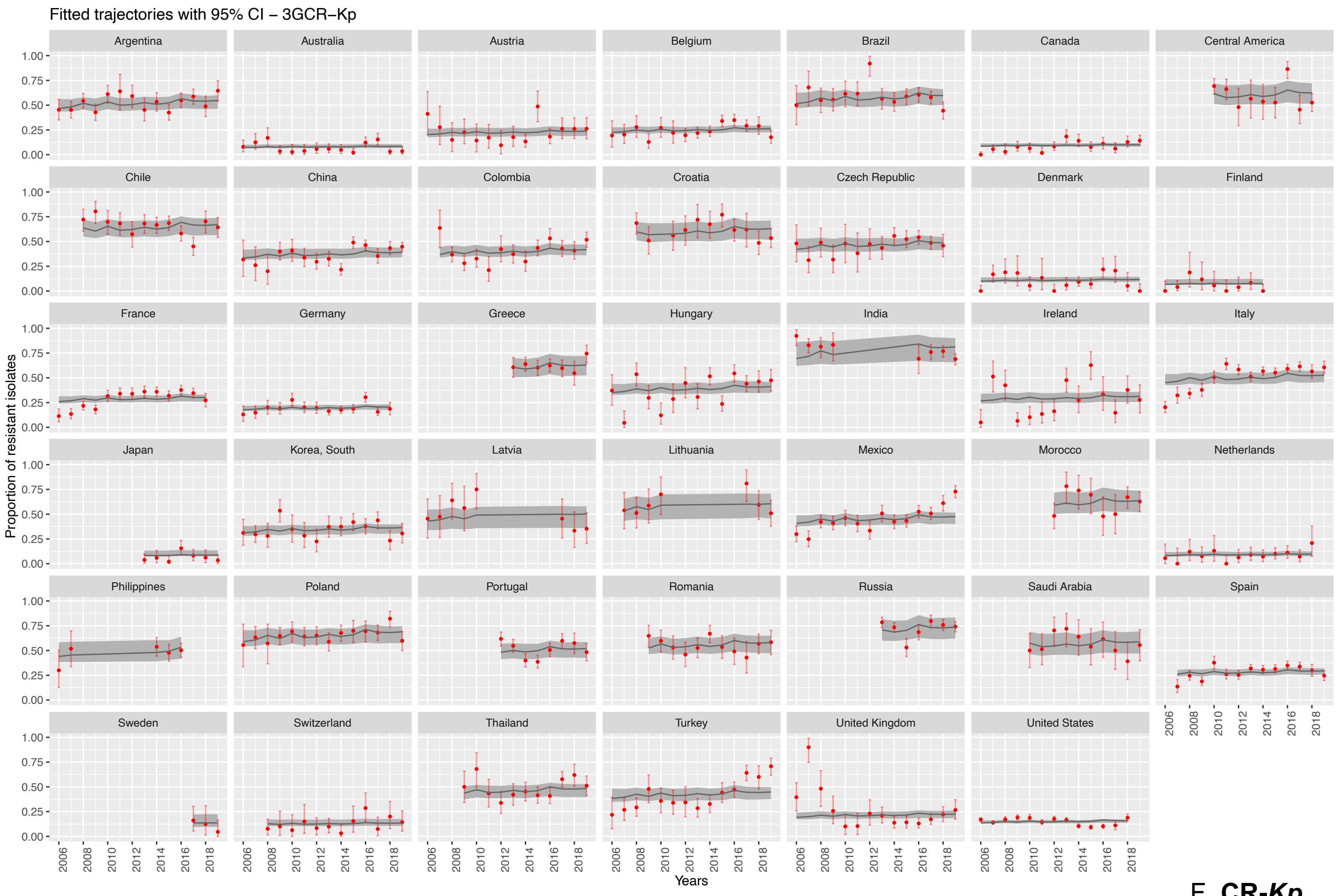

F. **CR-*Kp***

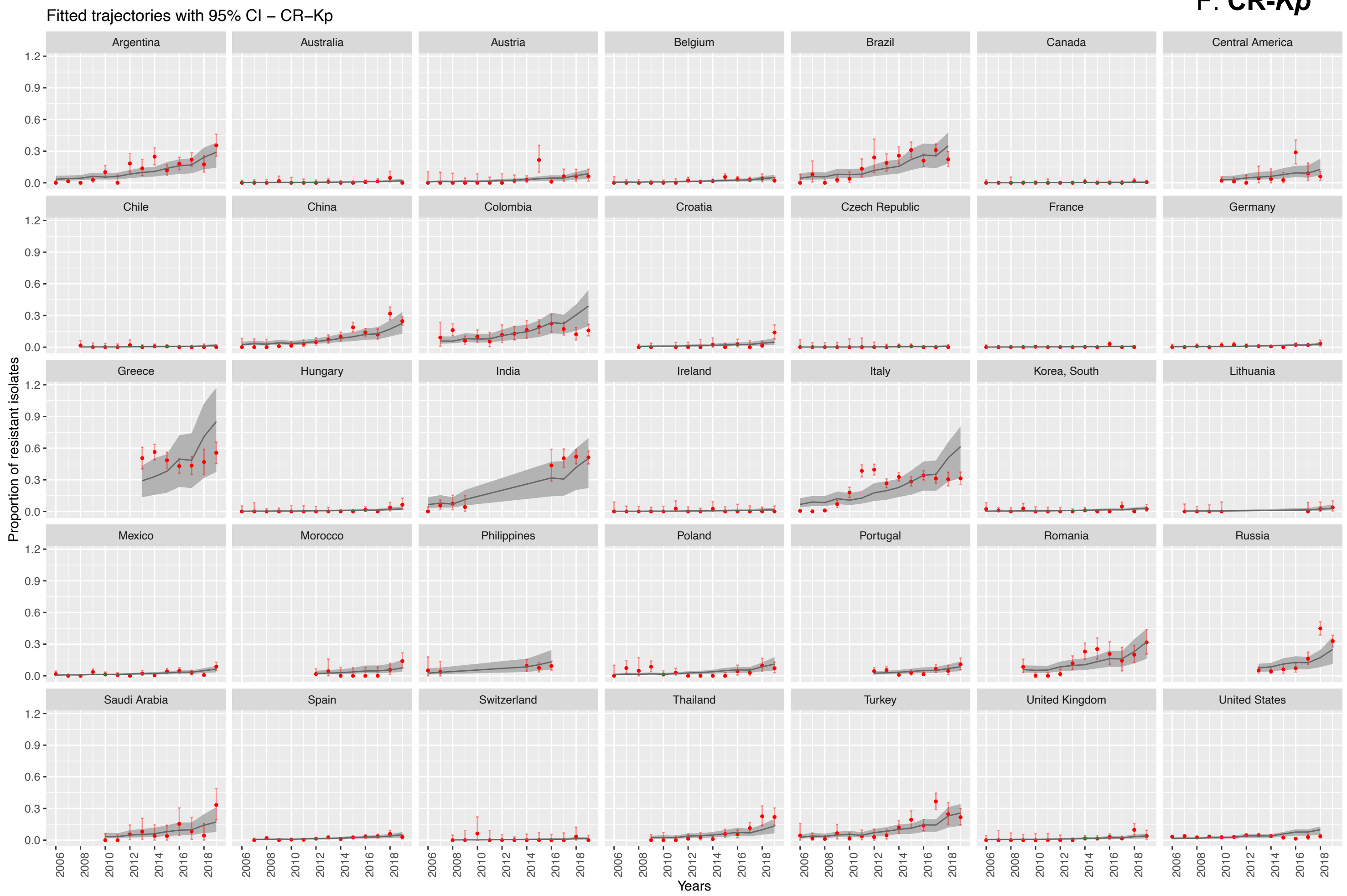

G. **FR-*Pa***

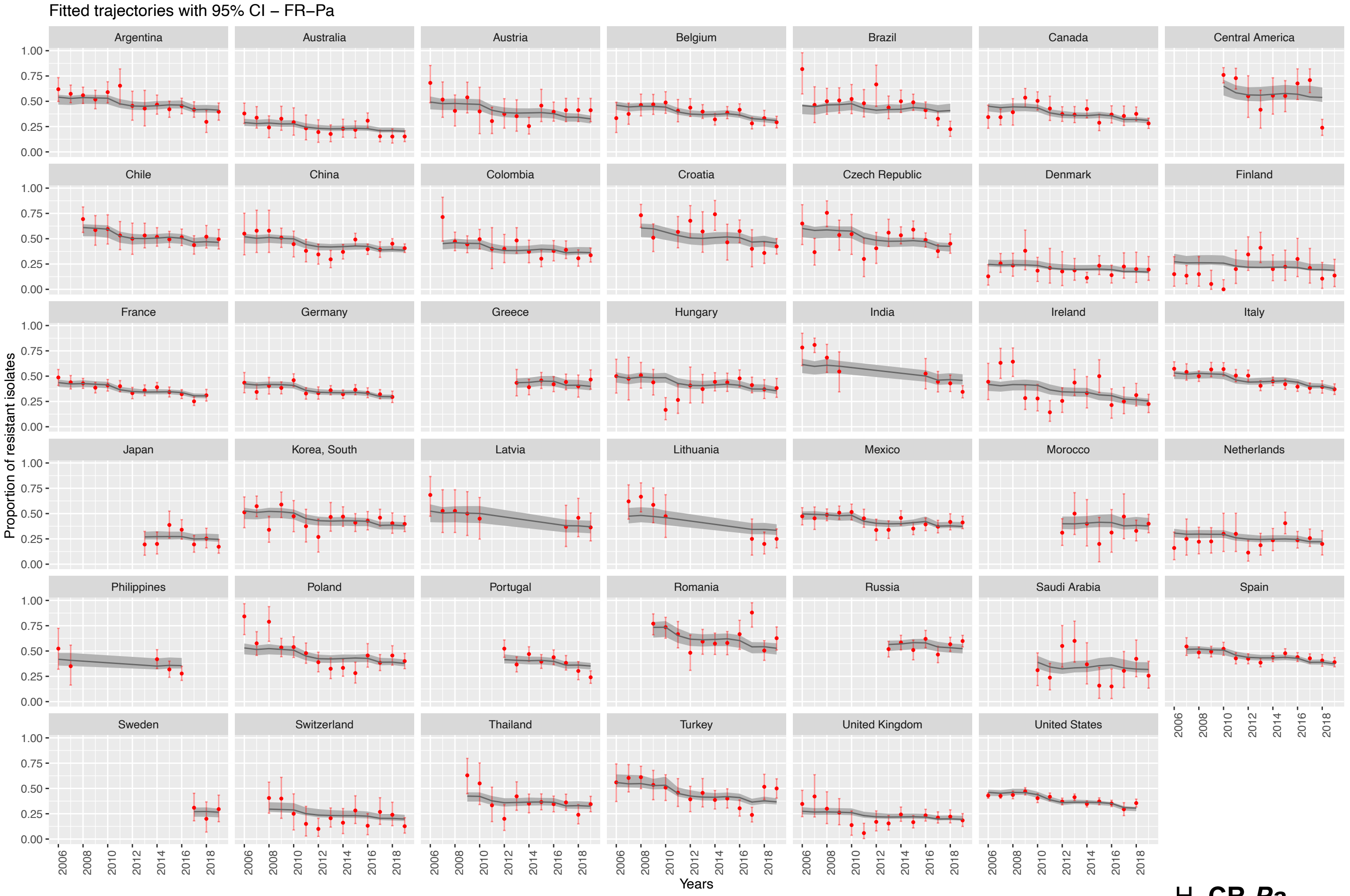
Fitted trajectories with 95% CI – FR–Pa
Argentina
Australia
Austria
Belgium
Brazil
Canada
Central America
Chile
China
Colombia
Croatia
Czech Republic
Denmark
Finland
France
Germany
Greece
Hungary
India
Ireland
Italy
Japan
Korea, South
Latvia
Lithuania
Mexico
Morocco
Netherlands
Philippines
Poland
Portugal
Romania
Russia
Saudi Arabia
Spain
Sweden
Switzerland
Thailand
Turkey
United Kingdom
United States
Proportion of resistant isolates
Years

H. **CR-*Pa***

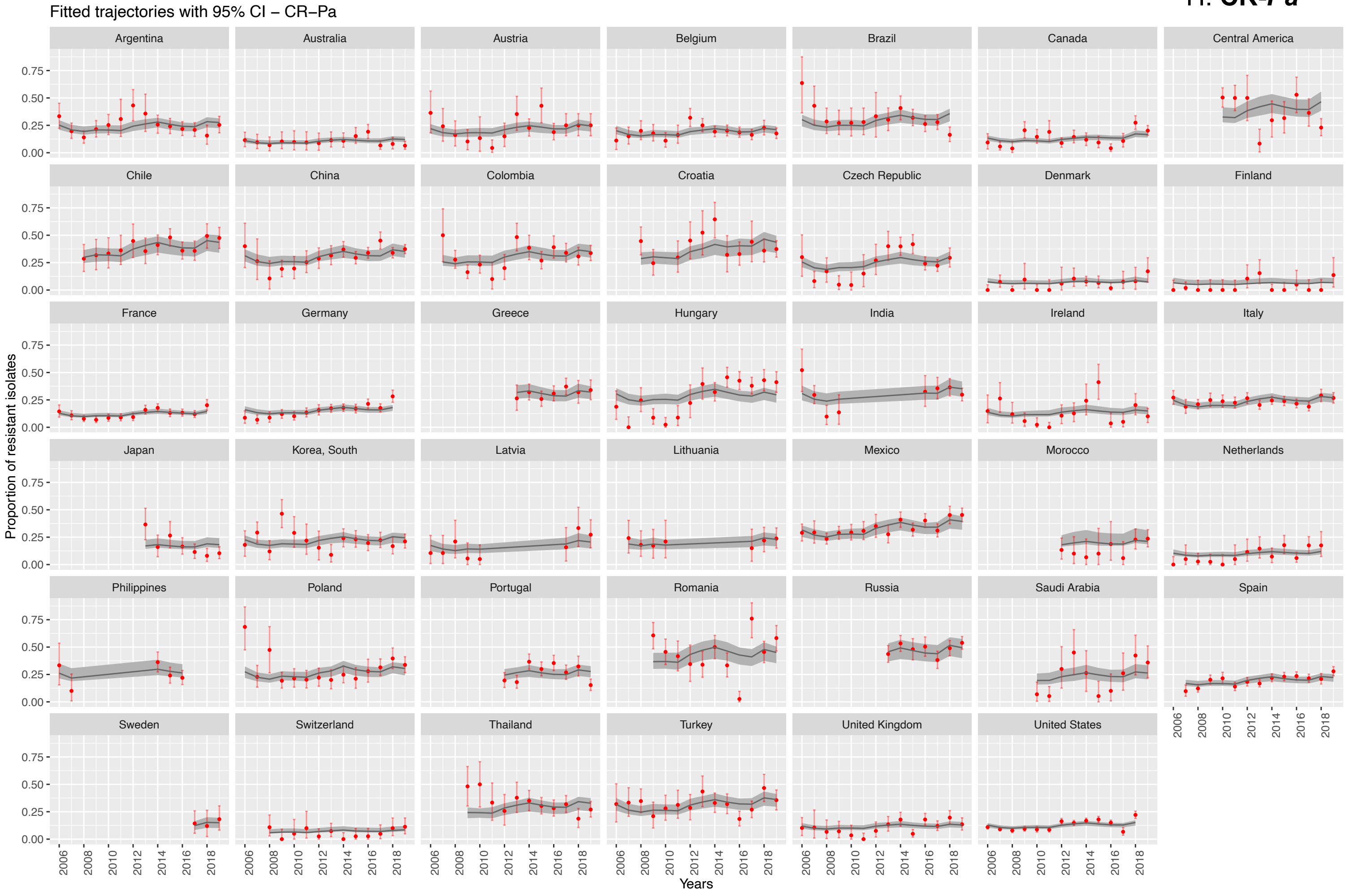
Fitted trajectories with 95% CI – CR–Pa
Argentina
Australia
Austria
Belgium
Brazil
Canada
Central America
Chile
China
Colombia
Croatia
Czech Republic
Denmark
Finland
France
Germany
Greece
Hungary
India
Ireland
Italy
Japan
Korea, South
Latvia
Lithuania
Mexico
Morocco
Netherlands
Philippines
Poland
Portugal
Romania
Russia
Saudi Arabia
Spain
Sweden
Switzerland
Thailand
Turkey
United Kingdom
United States
Proportion of resistant isolates
Years

I. **FR-*Ab***

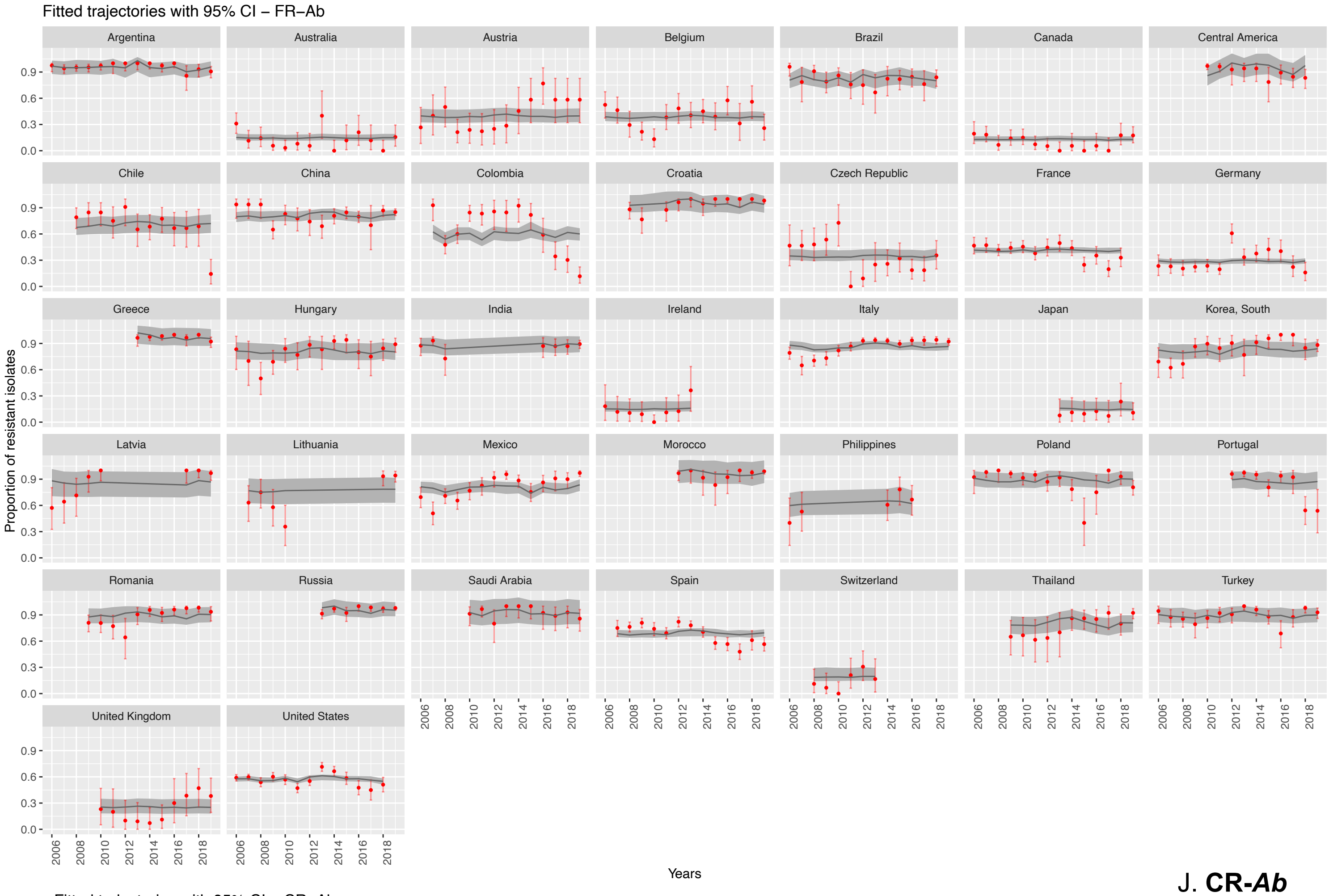
Fitted trajectories with 95% CI – FR–Ab
Argentina
Australia
Austria
Belgium
Brazil
Canada
Central America
Chile
China
Colombia
Croatia
Czech Republic
France
Germany
Greece
Hungary
India
Ireland
Italy
Japan
Korea, South
Latvia
Lithuania
Mexico
Morocco
Philippines
Poland
Portugal
Romania
Russia
Saudi Arabia
Spain
Switzerland
Thailand
Turkey
United Kingdom
United States
Proportion of resistant isolates
0.0
0.3
0.6
0.9
2006
2008
2010
2012
2014
2016
2018
Years

J. **CR-*Ab***

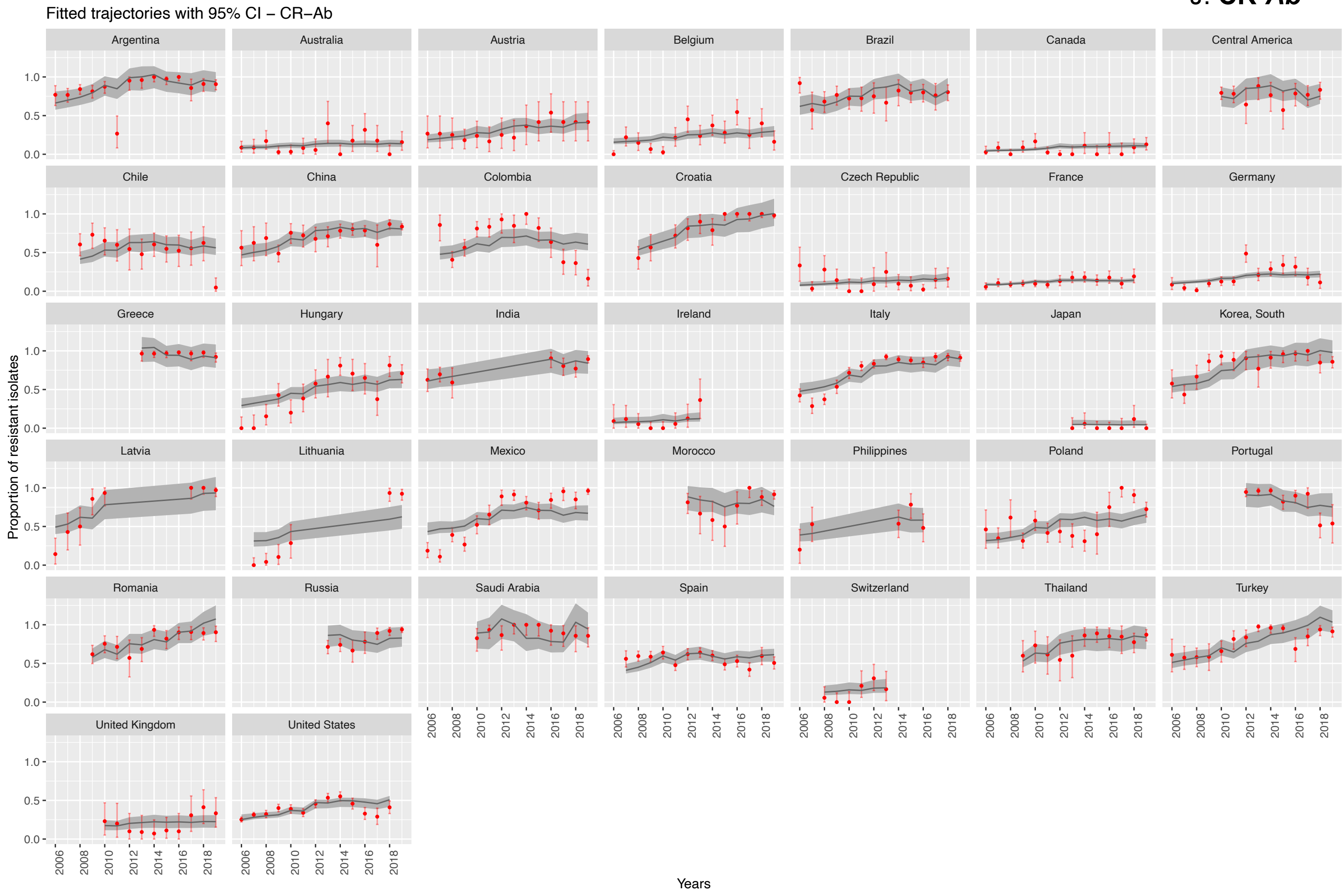
Fitted trajectories with 95% CI – CR–Ab
Argentina
Australia
Austria
Belgium
Brazil
Canada
Central America
Chile
China
Colombia
Croatia
Czech Republic
France
Germany
Greece
Hungary
India
Ireland
Italy
Japan
Korea, South
Latvia
Lithuania
Mexico
Morocco
Philippines
Poland
Portugal
Romania
Russia
Saudi Arabia
Spain
Switzerland
Thailand
Turkey
United Kingdom
United States
Proportion of resistant isolates
0.0
0.5
1.0
2006
2008
2010
2012
2014
2016
2018
Years

K. **VR-*E***

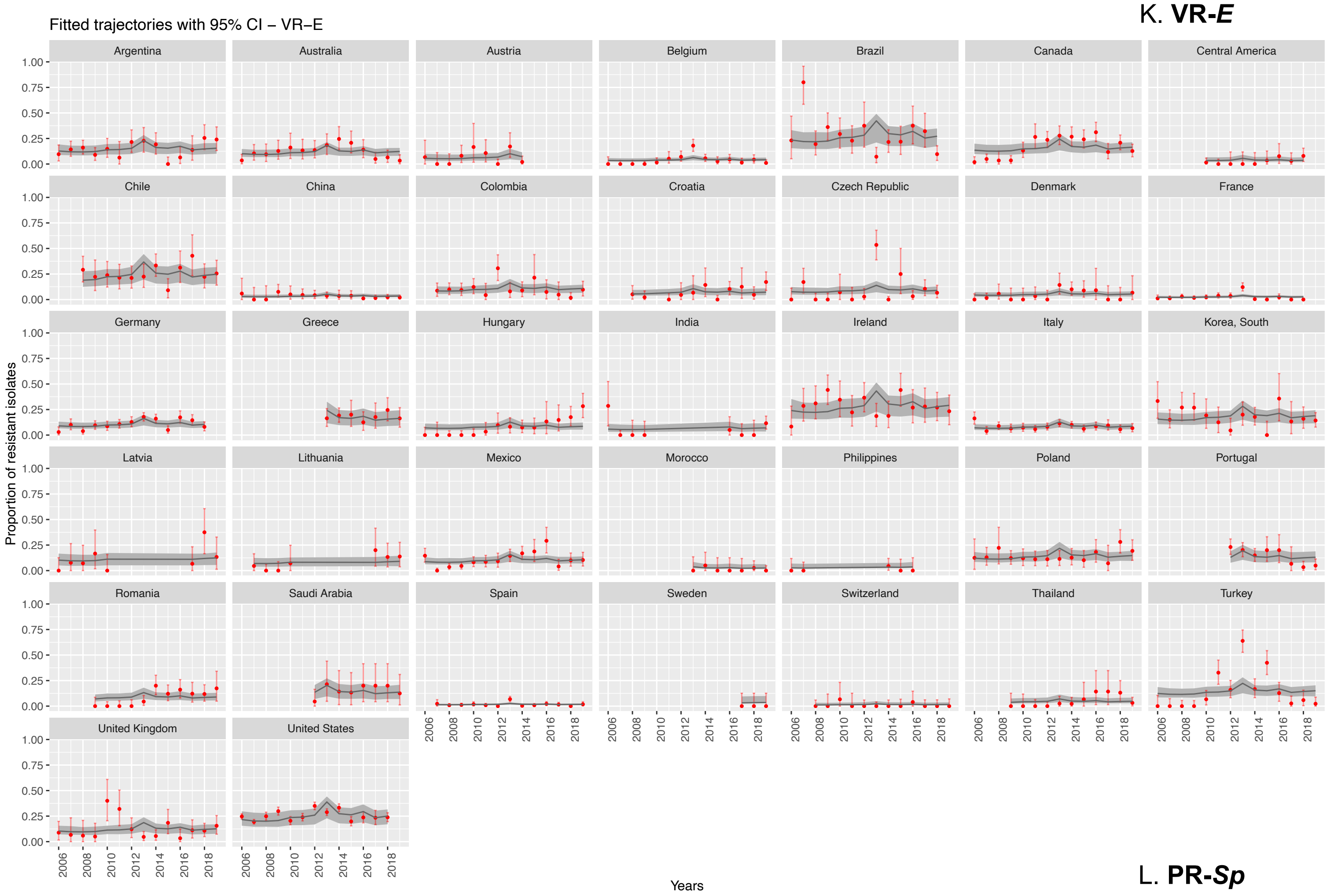

Fitted trajectories with 95% CI – VR–E
Argentina
Australia
Austria
Belgium
Brazil
Canada
Central America
Chile
China
Colombia
Croatia
Czech Republic
Denmark
France
Germany
Greece
Hungary
India
Ireland
Italy
Korea, South
Latvia
Lithuania
Mexico
Morocco
Philippines
Poland
Portugal
Romania
Saudi Arabia
Spain
Sweden
Switzerland
Thailand
Turkey
United Kingdom
United States
Proportion of resistant isolates
Years

L. **PR-*Sp***

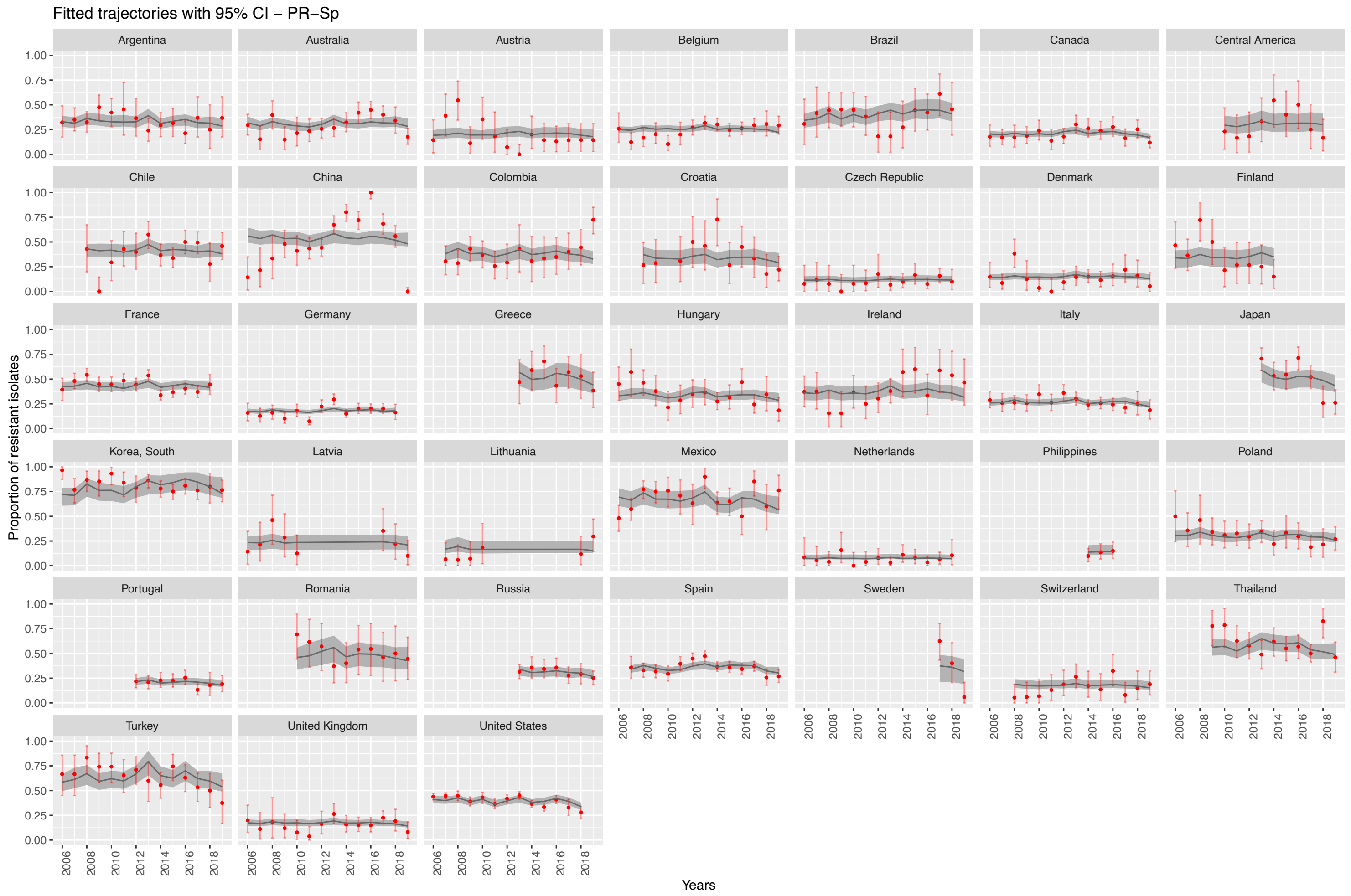

Fitted trajectories with 95% CI – PR–Sp
Argentina
Australia
Austria
Belgium
Brazil
Canada
Central America
Chile
China
Colombia
Croatia
Czech Republic
Denmark
Finland
France
Germany
Greece
Hungary
Ireland
Italy
Japan
Korea, South
Latvia
Lithuania
Mexico
Netherlands
Philippines
Poland
Portugal
Romania
Russia
Spain
Sweden
Switzerland
Thailand
Turkey
United Kingdom
United States
Proportion of resistant isolates
Years

M. **MLR-*Sp***

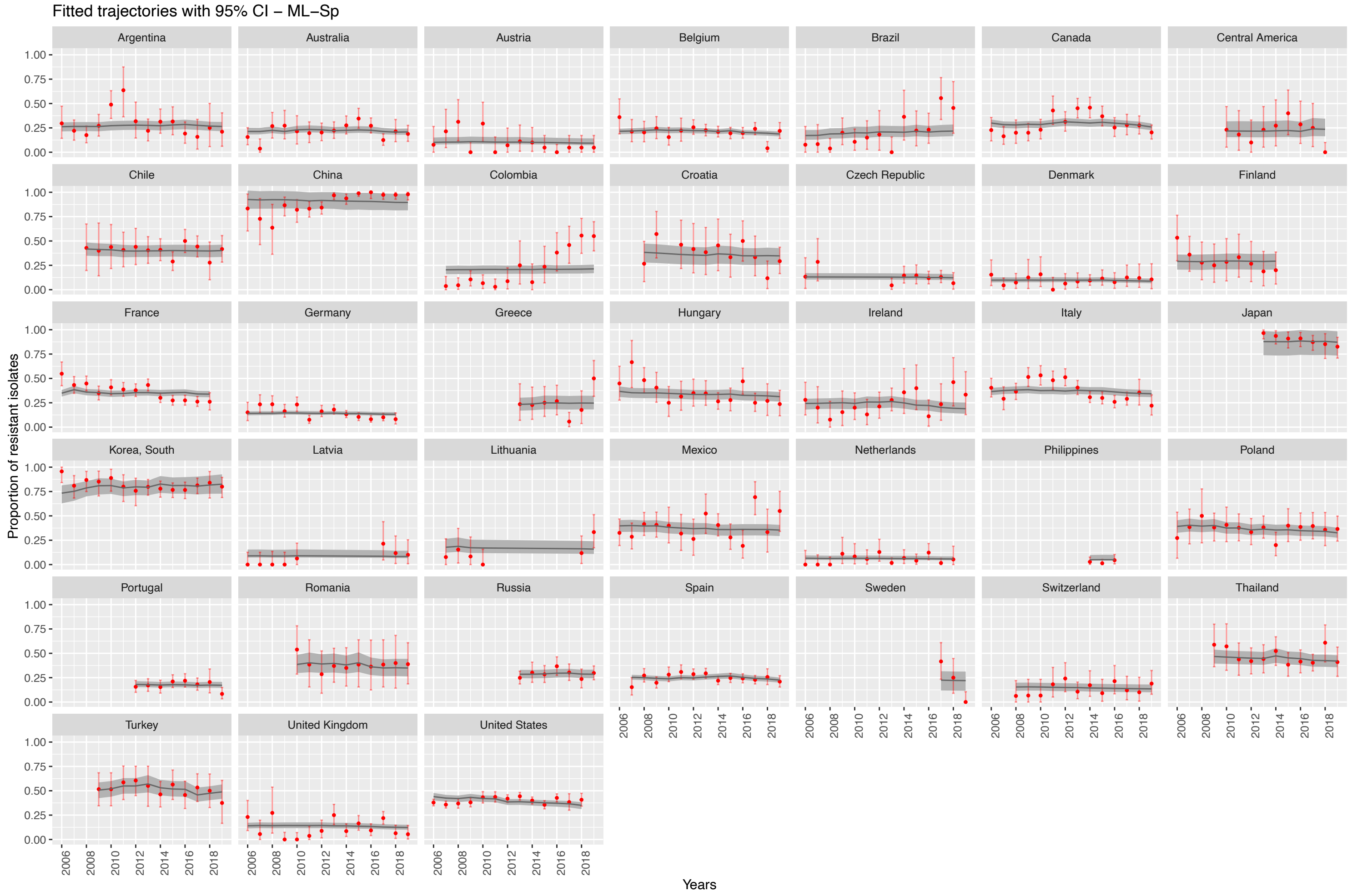
Fitted trajectories with 95% CI – ML–Sp
Argentina
Australia
Austria
Belgium
Brazil
Canada
Central America
Chile
China
Colombia
Croatia
Czech Republic
Denmark
Finland
France
Germany
Greece
Hungary
Ireland
Italy
Japan
Korea, South
Latvia
Lithuania
Mexico
Netherlands
Philippines
Poland
Portugal
Romania
Russia
Spain
Sweden
Switzerland
Thailand
Turkey
United Kingdom
United States
Proportion of resistant isolates
0.00
0.25
0.50
0.75
1.00
Years
2006
2008
2010
2012
2014
2016
2018

***Supplementary Text S3. Sensitivity analyses description.***

*Description of analyses*

**Sensitivity analysis 1** was performed to assess the potential impact of defining a threshold at 10 on the minimum number of isolates tested by country-year to be included in the study. We evaluated model's estimations by running an additional analysis in which the threshold was set to 20 isolates instead of 10.

**Sensitivity analysis 2** was performed to assess the potential impact on the model's estimates of using ESAC-Net data instead of IQVIA MIDAS data for antibiotic sales for Denmark, Latvia, Lithuania, and the Netherlands. We ran an additional analysis where we excluded these four countries from the analysis.

**Sensitivity analysis 3** was performed to assess whether mixing isolates sources could impact the association between antibiotic resistance and covariables. We ran an additional analysis on a subset of isolates originating only from blood source, representing bloodstream infections.

**Sensitivity analysis 4** was performed to assess whether imputation of missing ABR rates for some drug-bug-country-year combination could impact the resulting association between antibiotic resistance and covariables. We ran an additional analysis on a dataset where we excluded all missing datapoints.

*Description of results*

**Sensitivity analyses 1, 2 and 4** assessed the impacts of methodological choices on the model's results. Despite sample sizes reduction, resulting in different 95% confidence interval widths, sensitivity analysis 1, 2 and 4 showed similar signs of association and coefficients estimations for all covariables in the univariate analysis, compared to the main analysis.

Choosing an isolate threshold of 10 to ensure data reliability was adequate, as sample size is high, but estimations are still consistent when increasing this threshold to 20. Similarly, results concerning association of antibiotic sales and antibiotic resistance were thus not affected by using EARS-Net data. Moreover, ABR rates imputation did not significantly alter the model results.

Overall, sensitivity analyses 1, 2 and 4 showed that there were no major changes in interpretation in the statistical analyses.

Results from **sensitivity analysis 3** were expected to be sensibly different. First, bloodstream infections are a particular infections' subset, usually associated with worse medical outcomes and hospitalization. Consequently, patients presenting to the hospital with bacterial bloodstream infections are possibly more representative of overall bloodstream infections than other infection types (urinary tract infections, respiratory infections, …). Second, blood isolates represent more stable surveillance data across different countries and maybe can lead to a better representation of true prevalence. It was then expected that including only blood samples would strongly reduce sample sizes and possibly modify ABR rates (appendix p 18 figure S7). Number of blood isolates included in the analysis are depicted in appendix p 13 figure S3 B.

Overall, analysis of blood only samples showed consistent coefficients estimations and signs of associations in univariate results compared to the main analysis; except for some drug-bug pairs and covariables.

First, GDP was negatively associated with ABR rates for two drug-bug pairs (3GCR-*Kp* and CR-*Ab*), whereas it was mostly positively associated with ABR rates when looking at all samples' sources. For example, we observed that CR-*Ab* coefficients for GDP showed different estimations: 1.25 [1.02-1.54] in the main analysis versus 0.82 [0.60-1.12] in the blood-only analysis. However, as 95% confidence intervals are overlapping, no significant conclusion is possible. Second, temperature coefficients suggested a stronger association between temperature and ABR rates in CR-*Ab*, PR-*Sp* and VR-*E* from blood samples than in the main analysis. Third, some co-variables association coefficients had different signs for *S. pneumoniae* associated pairs: this was the case for macrolides and temperature for MLR-*Sp* for example. This last point could be explained by the fact that ABR rates in blood for *S. pneumoniae* were overall lower than ABR rates in all infection samples – as seen in the boxplots in appendix Figure S7 p 18 – thus changing the association between ABR rates and some co-variables.

***Supplementary Table S6. Univariate results for the main analysis and sensitivity analyses 1-4, by drug-bug pair.***

Estimated coefficients from univariate analyses, using the mixed-effect negative binomial, for:

- **A. FR-Ec**: fluoroquinolone-resistant *E. coli*
- **B. APR-Ec**: aminopenicillin-resistant *E. coli*
- **C. 3GCR-Ec**: third generation cephalosporin-resistant *E. coli*
- **D. FR-Kp**: fluoroquinolone-resistant *K. pneumoniae*
- **E. 3GCR-Kp**: third generation cephalosporin-resistant *K. pneumoniae*
- **F. CR-Kp**: carbapenem-resistant *K. pneumoniae*
- **G. FR-Pa**: fluoroquinolone-resistant *P. aeruginosa*
- **H. CR-Pa**: carbapenem-resistant *P. aeruginosa*
- **I. FR-Ab**: fluoroquinolone-resistant *A. baumannii*
- **J. CR-Ab**: carbapenem-resistant *A. baumannii*
- **K. VR-E**: vancomycin-resistant Enterococci
- **L. PR-Sp**: penicillin-non-susceptible *S. pneumoniae*
- **M. MLR-Sp**: macrolide-resistant *S. pneumoniae*

Data represent n, the number of observations; C, the number of countries and Y, the number of years. Models represent M_null, the intercept-only model and M_uni, the univariate model. Fixed effects (FE) are reported in the exponential form. Random effects (RE) are reported: spatial (top) and temporal (bottom). Coefficients associated with a p-value < 5% are highlighted in bold.

Antibiotic sales of interest for each drug-bug pair were the following:

quinolones (FR-*Ec*); broad-spectrum penicillins (APR-*Ec* and PR-*Sp*); third generation cephalosporins (3GCR-*Ec* and 3GCR-*Kp*); carbapenems (CR-*Kp*; CR-*Pa* and CR-*Ab*); glycopeptides (VR-*E*) and macrolides (MLR-*Sp*).

Sensitivity analyses are the following:

- **Sensitivity analysis 1:** impact of changing the 10 isolates threshold for excluding country-year to 20 isolates threshold.
- **Sensitivity analysis 2:** impact of excluding countries for which we used ESAC-Net data for antibiotic use as no data was available in IQVIA MIDAS: only countries with available IQVIA MIDAS data tested.
- **Sensitivity analysis 3:** analysis on blood samples only.
- **Sensitivity analysis 4:** analysis when no imputation of missing ABR rates is performed: only non-imputed data included.

A.

**FR-*Ec***

| | Main analysis | | Sensitivity analysis 1 | | Sensitivity analysis 2 | | Sensitivity analysis 3 | | Sensitivity analysis 4 | |
|---|---|---|---|---|---|---|---|---|---|---|
| Data | (n=471; C=41; Y=14) | | (n=471; C=41; Y=14) | | (n=429; C=37; Y=14) | | (n=415; C=38; Y=14) | | (n=385; C=41; Y=14) | |
| **Variable** | **FE** | **RE *** | **FE** | **RE *** | **FE** | **RE *** | **FE** | **RE *** | **FE** | **RE *** |
| | **M_null** | | | | | | | | | |
| $\widehat{\beta}_0$ | 0.34<br>0.30-0.38 | 0.18<br>0.0005 | 0.34<br>0.29-0.38 | 0.18<br>0.0005 | 0.36<br>0.32-0.41 | 0.15<br>0.0008 | 0.34<br>0.29-0.39 | 0.19<br>0.0000 | 0.34<br>0.30-0.39 | 0.17<br>0.0007 |
| | **M_uni** | | | | | | | | | |
| ATB sales of interest | **1.11<br>1.06-1.17** | **0.17<br>0.0002** | **1.11<br>1.05-1.17** | **0.17<br>0.0002** | **1.10<br>1.05-1.16** | **0.15<br>0.0005** | **1.08<br>1.00-1.16** | **0.18<br>0.0000** | **1.13<br>1.07-1.19** | **0.16<br>0.0004** |
| Global ATB sales | 1.00<br>0.92-1.08 | 0.18<br>0.0005 | 1.00<br>0.92-1.08 | 0.18<br>0.0005 | 0.98<br>0.91-1.06 | 0.15<br>0.0008 | 0.98<br>0.88-1.09 | 0.19<br>0.0000 | 0.99<br>0.91-1.08 | 0.17<br>0.0007 |
| GHS index | **0.80<br>0.72-0.90** | **0.13<br>0.0005** | **0.80<br>0.72-0.90** | **0.13<br>0.0005** | **0.82<br>0.74-0.92** | **0.11<br>0.0008** | **0.77<br>0.68-0.87** | **0.12<br>0.0000** | **0.80<br>0.72-0.86** | **0.12<br>0.0007** |
| GDP | **0.89<br>0.83-0.94** | **0.13<br>0.0002** | **0.89<br>0.84-0.94** | **0.13<br>0.0002** | **0.88<br>0.82-0.93** | **0.11<br>0.0005** | **0.86<br>0.80-0.94** | **0.13<br>0.0000** | **0.87<br>0.81-0.93** | **0.12<br>0.0006** |
| Extreme events | **1.20<br>1.06-1.35** | **0.15<br>0.0005** | **1.20<br>1.06-1.35** | **0.15<br>0.0005** | **1.18<br>1.05-1.33** | **0.13<br>0.0008** | **1.19<br>1.03-1.36** | **0.16<br>0.0000** | **1.18<br>1.05-1.32** | **0.14<br>0.0007** |
| Temperature | **1.21<br>1.11-1.32** | **0.11<br>0.0009** | **1.21<br>1.11-1.32** | **0.11<br>0.0009** | **1.20<br>1.10-1.31** | **0.09<br>0.0012** | **1.15<br>1.03-1.27** | **0.15<br>0.0000** | **1.20<br>1.10-1.30** | **0.11<br>0.0011** |
| Rainfall | 1.05<br>0.98-1.12 | 0.17<br>0.0004 | 1.05<br>0.98-1.11 | 0.17<br>0.0004 | 1.05<br>0.98-1.12 | 0.14<br>0.0007 | 1.05<br>0.95-1.15 | 0.18<br>0.0000 | 1.06<br>0.98-1.13 | 0.16<br>0.0006 |
| Relative Humidity | **0.93<br>0.86-1.00** | **0.15<br>0.0006** | **0.93<br>0.86-1.00** | **0.15<br>0.0006** | 0.95<br>0.87-1.02 | 0.13<br>0.0009 | **0.87<br>0.79-0.96** | **0.15<br>0.0006** | **0.92<br>0.86-1.00** | **0.14<br>0.0008** |
| Population Density | 1.04<br>0.92-1.18 | 0.17<br>0.0005 | 1.04<br>0.92-1.18 | 0.17<br>0.0005 | 1.09<br>0.97-1.22 | 0.14<br>0.0009 | 1.05<br>0.91-1.21 | 0.19<br>0.0000 | 1.04<br>0.92-1.18 | 0.17<br>0.0007 |
| Tourist Arrivals | 0.91<br>0.83-1.01 | 0.16<br>0.0003 | 0.91<br>0.83-1.01 | 0.16<br>0.0003 | 0.92<br>0.84-1.02 | 0.14<br>0.0006 | **0.88<br>0.78-0.99** | **0.18<br>0.0000** | **0.88<br>0.80-0.98** | **0.16<br>0.0004** |
| Tourist Departures | **0.90<br>0.83-0.97** | **0.14<br>0.0002** | **0.90<br>0.83-0.97** | **0.14<br>0.0002** | **0.91<br>0.84-0.98** | **0.12<br>0.0005** | **0.85<br>0.77-0.93** | **0.15<br>0.0000** | **0.89<br>0.83-0.97** | **0.13<br>0.0004** |

*spatial (top) and temporal (bottom) random effect

B.

**APR-*Ec***

| | Main analysis | | Sensitivity analysis 1 | | Sensitivity analysis 2 | | Sensitivity analysis 3 | | Sensitivity analysis 4 | |
|---|---|---|---|---|---|---|---|---|---|---|
| Data | (n=471; C=41; Y=14) | | (n=471; C=41; Y=14) | | (n=429; C=37; Y=14) | | (n=415; C=38; Y=14) | | (n=385; C=41; Y=14) | |
| **Variable** | **FE** | **RE *** | **FE** | **RE *** | **FE** | **RE *** | **FE** | **RE *** | **FE** | **RE *** |
| | **M_null** | | | | | | | | | |
| $\widehat{\beta}_0$ | 0.66<br>0.62-0.69 | 0.03<br>0.0003 | 0.66<br>0.62-0.69 | 0.03<br>0.0003 | 0.67<br>0.63-0.71 | 0.03<br>0.0003 | 0.66<br>0.63-0.70 | 0.02<br>0.0000 | 0.66<br>0.63-0.70 | 0.03<br>0.0004 |
| | **M_uni** | | | | | | | | | |
| ATB sales of interest | 0.98<br>0.94-1.02 | 0.03<br>0.0003 | 0.98<br>0.95-1.02 | 0.03<br>0.0004 | 0.98<br>0.94-1.02 | 0.03<br>0.0003 | 0.98<br>0.93-1.03 | 0.02<br>0.0000 | 0.98<br>0.94-1.02 | 0.03<br>0.0004 |
| Global ATB sales | 0.98<br>0.94-1.01 | 0.03<br>0.0003 | 0.98<br>0.94-1.01 | 0.03<br>0.0003 | **0.97<br>0.93-1.00** | **0.02<br>0.0003** | 0.97<br>0.93-1.02 | 0.02<br>0.0000 | 0.97<br>0.93-1.01 | 0.02<br>0.0004 |
| GHS index | **0.92<br>0.88-0.96** | **0.02<br>0.0003** | **0.92<br>0.88-0.96** | **0.02<br>0.0003** | **0.93<br>0.89-0.97** | **0.02<br>0.0003** | **0.90<br>0.86-0.95** | **0.01<br>0.0000** | **0.92<br>0.88-0.96** | **0.02<br>0.0004** |
| GDP | **0.94<br>0.91-0.97** | **0.02<br>0.0003** | **0.94<br>0.91-0.97** | **0.02<br>0.0003** | **0.95<br>0.91-0.98** | **0.02<br>0.0003** | **0.93<br>0.89-0.97** | **0.01<br>0.0000** | **0.94<br>0.91-0.97** | **0.02<br>0.0005** |
| Extreme events | **1.09<br>1.04-1.14** | **0.02<br>0.0003** | **1.09<br>1.04-1.14** | **0.02<br>0.0003** | **1.08<br>1.03-1.14** | **0.02<br>0.0003** | **1.08<br>1.03-1.14** | **0.02<br>0.0000** | **1.08<br>1.03-1.13** | **0.02<br>0.0004** |
| Temperature | **1.09<br>1.05-1.13** | **0.02<br>0.0005** | **1.09<br>1.05-1.13** | **0.02<br>0.0005** | **1.08<br>1.04-1.12** | **0.02<br>0.0005** | **1.08<br>1.04-1.13** | **0.02<br>0.0000** | **1.08<br>1.04-1.11** | **0.02<br>0.0006** |
| Rainfall | 1.02<br>0.98-1.05 | 0.03<br>0.0003 | 1.02<br>0.99-1.05 | 0.03<br>0.0003 | 1.02<br>0.98-1.05 | 0.02<br>0.0003 | 1.02<br>0.97-1.07 | 0.02<br>0.0000 | 1.02<br>0.99-1.06 | 0.02<br>0.0004 |
| Relative Humidity | **0.95<br>0.92-0.99** | **0.02<br>0.0004** | **0.95<br>0.92-0.99** | **0.02<br>0.0004** | **0.96<br>0.92-1.00** | **0.02<br>0.0004** | **0.93<br>0.89-0.98** | **0.02<br>0.0000** | **0.96<br>0.92-0.99** | **0.02<br>0.0005** |
| Population Density | 1.00<br>0.96-1.06 | 0.03<br>0.0003 | 1.01<br>0.96-1.06 | 0.03<br>0.0003 | 1.03<br>0.98-1.08 | 0.02<br>0.0003 | 1.02<br>0.97-1.08 | 0.02<br>0.0000 | 1.00<br>0.95-1.05 | 0.03<br>0.0004 |
| Tourist Arrivals | **0.95<br>0.91-0.99** | **0.03<br>0.0002** | **0.95<br>0.91-0.99** | **0.03<br>0.0002** | **0.95<br>0.91-1.00** | **0.02<br>0.0002** | **0.95<br>0.90-1.00** | **0.02<br>0.0000** | **0.95<br>0.91-0.99** | **0.02<br>0.0003** |
| Tourist Departures | **0.92<br>0.89-0.95** | **0.02<br>0.0001** | **0.92<br>0.89-0.95** | **0.02<br>0.0001** | **0.92<br>0.89-0.96** | **0.02<br>0.0001** | **0.92<br>0.88-0.96** | **0.02<br>0.0000** | **0.92<br>0.89-0.96** | **0.02<br>0.0002** |

*spatial (top) and temporal (bottom) random effect

C.

**3GCR-*Ec***

| | Main analysis | | Sensitivity analysis 1 | | Sensitivity analysis 2 | | Sensitivity analysis 3 | | Sensitivity analysis 4 | |
|---|---|---|---|---|---|---|---|---|---|---|
| Data | (n=471; C=41; Y=14) | | (n=471; C=41; Y=14) | | (n=429; C=37; Y=14) | | (n=415; C=38; Y=14) | | (n=385; C=41; Y=14) | |
| **Variable** | **FE** | **RE *** | **FE** | **RE *** | **FE** | **RE *** | **FE** | **RE *** | **FE** | **RE *** |
| | **M_null** | | | | | | | | | |
| $\hat{\beta}_0$ | 0.34<br>0.30-0.38 | 0.18<br>0.0005 | 0.21<br>0.18-0.25 | 0.29<br>0.0116 | 0.23<br>0.19-0.27 | 0.24<br>0.0106 | 0.19<br>0.15-0.24 | 0.46<br>0.0095 | 0.22<br>0.18-0.26 | 0.29<br>0.0178 |
| | **M_uni** | | | | | | | | | |
| **ATB sales of interest** | 1.02<br>0.92-1.13 | 0.29<br>0.0116 | 1.02<br>0.92-1.13 | 0.29<br>0.012 | 0.99<br>0.90-1.09 | 0.24<br>0.0106 | 1.03<br>0.90-1.19 | 0.46<br>0.0090 | 0.99<br>0.89-1.10 | 0.29<br>0.0179 |
| **Global ATB sales** | 0.98<br>0.87-1.11 | 0.29<br>0.0117 | 0.98<br>0.87-1.11 | 0.29<br>0.0117 | 0.94<br>0.84-1.06 | 0.23<br>0.0108 | 0.97<br>0.82-1.15 | 0.45<br>0.0099 | 0.92<br>0.81-1.05 | 0.28<br>0.0186 |
| **GHS index** | **0.75<br>0.65-0.86** | **0.20<br>0.0115** | **0.75<br>0.65-0.86** | **0.20<br>0.0115** | **0.78<br>0.68-0.90** | **0.18<br>0.0105** | **0.69<br>0.57-0.83** | **0.31<br>0.0092** | **0.75<br>0.65-0.87** | **0.21<br>0.0175** |
| **GDP** | **0.85<br>0.76-0.96** | **0.21<br>0.0123** | **0.85<br>0.76-0.96** | **0.21<br>0.0123** | **0.85<br>0.76-0.95** | **0.17<br>0.0105** | **0.83<br>0.67-0.99** | **0.32<br>0.0166** | **0.85<br>0.75-0.95** | **0.21<br>0.0186** |
| **Extreme events** | **1.30<br>1.12-1.52** | **0.22<br>0.0116** | **1.30<br>1.12-1.52** | **0.22<br>0.0116** | **1.28<br>1.11-1.48** | **0.18<br>0.0105** | **1.36<br>1.10-1.67** | **0.37<br>0.0094** | **1.28<br>1.11-1.48** | **0.22<br>0.0178** |
| **Temperature** | **1.28<br>1.13-1.44** | **0.19<br>0.0131** | **1.28<br>1.13-1.44** | **0.20<br>0.0131** | **1.26<br>1.12-1.42** | **0.17<br>0.0118** | **1.27<br>1.08-1.49** | **0.35<br>0.0088** | **1.22<br>1.08-1.38** | **0.21<br>0.0188** |
| **Rainfall** | 1.07<br>0.96-1.19 | 0.27<br>0.0114 | 1.07<br>0.96-1.20 | 0.27<br>0.0114 | 1.07<br>0.96-1.20 | 0.23<br>0.0104 | 1.04<br>0.90-1.21 | 0.45<br>0.0094 | 1.10<br>0.98-1.23 | 0.27<br>0.0174 |
| **Relative Humidity** | **0.86<br>0.77-0.97** | **0.22<br>0.0119** | **0.86<br>0.77-0.97** | **0.22<br>0.0119** | **0.89<br>0.79-1.00** | **0.19<br>0.0107** | **0.81<br>0.70-0.95** | **0.33<br>0.0092** | 0.89<br>0.79-1.00 | 0.23<br>0.0178 |
| **Population Density** | 1.01<br>0.86-1.19 | 0.29<br>0.0116 | 1.01<br>0.86-1.19 | 0.29<br>0.0116 | 1.09<br>0.93-1.27 | 0.24<br>0.0106 | 1.08<br>0.87-1.35 | 0.46<br>0.0090 | 0.99<br>0.84-1.17 | 0.29<br>0.0178 |
| **Tourist Arrivals** | 0.91<br>0.79-1.04 | 0.26<br>0.0113 | 0.91<br>0.79-1.04 | 0.27<br>0.0113 | 0.90<br>0.79-1.03 | 0.22<br>0.0102 | 0.84<br>0.70-1.01 | 0.42<br>0.0097 | 0.88<br>0.77-1.02 | 0.26<br>0.0172 |
| **Tourist Departures** | **0.85<br>0.76-0.95** | **0.23<br>0.0111** | **0.85<br>0.76-0.95** | **0.23<br>0.0111** | **0.87<br>0.77-0.97** | **0.21<br>0.0100** | **0.77<br>0.66-0.89** | **0.34<br>0.0104** | **0.85<br>0.76-0.96** | **0.24<br>0.0171** |

*spatial (top) and temporal (bottom) random effect

D.

**FR-*Kp***

| | Main analysis | | Sensitivity analysis 1 | | Sensitivity analysis 2 | | Sensitivity analysis 3 | | Sensitivity analysis 4 | |
|---|---|---|---|---|---|---|---|---|---|---|
| Data | (n=467; C=41; Y=14) | | (n=443; C=39; Y=14) | | (n=425; C=37; Y=14) | | (n=429; C=39; Y=14) | | (n=382; C=41; Y=14) | |
| **Variable** | **FE** | **RE *** | **FE** | **RE *** | **FE** | **RE *** | **FE** | **RE *** | **FE** | **RE *** |
| | **M_null** | | | | | | | | | |
| $\hat{\beta}_0$ | 0.26<br>0.21-0.33 | 0.49<br>0.0172 | 0.27<br>0.21-0.33 | 0.47<br>0.0154 | 0.27<br>0.21-0.34 | 0.46<br>0.0171 | 0.26<br>0.20-0.33 | 0.47<br>0.0200 | 0.26<br>0.20-0.33 | 0.52<br>0.0212 |
| | **M_uni** | | | | | | | | | |
| **ATB sales of interest** | 1.06<br>0.97-1.16 | 0.46<br>0.0188 | 1.07<br>0.98-1.17 | 0.44<br>0.0171 | 1.07<br>0.97-1.17 | 0.44<br>0.0188 | 1.07<br>0.95-1.19 | 0.43<br>0.0220 | **1.12<br>1.01-1.23** | **0.46<br>0.0241** |
| **Global ATB sales** | 1.00<br>0.88-1.13 | 0.49<br>0.0172 | 1.00<br>0.87-1.14 | 0.47<br>0.0154 | 0.99<br>0.87-1.13 | 0.47<br>0.0171 | 1.11<br>0.95-1.29 | 0.46<br>0.0192 | 0.98<br>0.85-1.13 | 0.52<br>0.0214 |
| **GHS index** | **0.63<br>0.53-0.74** | **0.27<br>0.0169** | **0.64<br>0.54-0.75** | **0.26<br>0.0151** | **0.65<br>0.55-0.77** | **0.26<br>0.0168** | **0.62<br>0.53-0.72** | **0.21<br>0.0193** | **0.61<br>0.52-0.72** | **0.26<br>0.0208** |
| **GDP** | **0.84<br>0.72-0.99** | **0.34<br>0.0287** | **0.82<br>0.70-0.96** | **0.31<br>0.0277** | 0.87<br>0.74-1.03 | 0.35<br>0.0258 | **0.69<br>0.59-0.81** | **0.23<br>0.0471** | **0.79<br>0.67-0.91** | **0.32<br>0.0393** |
| **Extreme events** | 1.07<br>0.86-1.34 | 0.49<br>0.0172 | 1.07<br>0.85-1.34 | 0.47<br>0.0154 | 1.06<br>0.84-1.34 | 0.46<br>0.0171 | 1.07<br>0.85-1.34 | 0.46<br>0.0200 | 1.07<br>0.86-1.33 | 0.51<br>0.0212 |
| **Temperature** | **1.24<br>1.06-1.46** | **0.41<br>0.0161** | **1.24<br>1.05-1.45** | **0.40<br>0.0143** | **1.28<br>1.08-1.50** | **0.37<br>0.0158** | **1.30<br>1.09-1.54** | **0.38<br>0.0180** | **1.23<br>1.05-1.44** | **0.44<br>0.0207** |
| **Rainfall** | 0.99<br>0.89-1.11 | 0.49<br>0.0172 | 0.99<br>0.88-1.11 | 0.47<br>0.0154 | 0.99<br>0.88-1.10 | 0.46<br>0.0171 | 0.94<br>0.82-1.09 | 0.46<br>0.0202 | 0.98<br>0.86-1.10 | 0.52<br>0.0213 |
| **Relative Humidity** | 0.90<br>0.79-1.02 | 0.44<br>0.0170 | 0.90<br>0.79-1.02 | 0.42<br>0.0152 | 0.89<br>0.78-1.02 | 0.41<br>0.0170 | **0.83<br>0.71-0.96** | **0.39<br>0.0202** | 0.89<br>0.79-1.01 | 0.45<br>0.0212 |
| **Population Density** | 0.92<br>0.74-1.13 | 0.48<br>0.0176 | 0.90<br>0.74-1.11 | 0.46<br>0.0159 | 1.01<br>0.82-1.24 | 0.46<br>0.0170 | 0.93<br>0.74-1.16 | 0.46<br>0.0204 | 0.94<br>0.75-1.16 | 0.50<br>0.0215 |
| **Tourist Arrivals** | 1.01<br>0.87-1.18 | 0.49<br>0.0170 | 0.99<br>0.85-1.16 | 0.47<br>0.0155 | 1.05<br>0.90-1.23 | 0.46<br>0.0163 | 0.97<br>0.81-1.16 | 0.47<br>0.0206 | 0.99<br>0.84-1.17 | 0.52<br>0.0213 |
| **Tourist Departures** | 0.99<br>0.87-1.13 | 0.49<br>0.0173 | 0.98<br>0.86-1.11 | 0.46<br>0.0157 | 0.95<br>0.83-1.09 | 0.43<br>0.0178 | **0.84<br>0.72-0.98** | **0.38<br>0.0230** | 0.92<br>0.80-1.05 | 0.47<br>0.0225 |

*spatial (top) and temporal (bottom) random effect

E.

**3GCR-*Kp***

| | Main analysis | | Sensitivity analysis 1 | | Sensitivity analysis 2 | | Sensitivity analysis 3 | | Sensitivity analysis 4 | |
|---|---|---|---|---|---|---|---|---|---|---|
| Data | (n=467; C=41; Y=14) | | (n=443; C=39; Y=14) | | (n=425; C=37; Y=14) | | (n=406; C=37; Y=14) | | (n=382; C=41; Y=14) | |
| **Variable** | **FE** | **RE *** | **FE** | **RE *** | **FE** | **RE *** | **FE** | **RE *** | **FE** | **RE *** |
| | **M_null** | | | | | | | | | |
| $\hat{\beta}_0$ | 0.32 | 0.50 | 0.33 | 0.46 | 0.33 | 0.46 | 0.32 | 0.54 | 0.31 | 0.51 |
| | 0.25-0.40 | 0.0044 | 0.26-0.40 | 0.0043 | 0.26-0.41 | 0.0045 | 0.25-0.41 | 0.0044 | 0.25-0.39 | 0.0075 |
| | **M_uni** | | | | | | | | | |
| ATB sales of interest | 0.95 | 0.50 | 0.97 | 0.46 | 0.93 | 0.45 | 1.09 | 0.56 | 0.90 | 0.51 |
| | 0.87-1.04 | 0.0046 | 0.88-1.07 | 0.0045 | 0.85-1.03 | 0.0048 | 0.95-1.26 | 0.0041 | 0.82-1.00 | 0.0083 |
| Global ATB sales | 1.05 | 0.50 | 1.06 | 0.45 | 1.04 | 0.46 | **1.17** | **0.56** | 1.01 | 0.51 |
| | 0.93-1.17 | 0.0040 | 0.94-1.19 | 0.0038 | 0.92-1.17 | 0.0041 | **1.00-1.37** | **0.0031** | 0.88-1.15 | 0.0073 |
| GHS index | **0.61** | **0.24** | **0.62** | **0.22** | **0.63** | **0.24** | **0.59** | **0.21** | **0.60** | **0.23** |
| | **0.52-0.71** | **0.0042** | **0.54-0.72** | **0.0041** | **0.54-0.74** | **0.0043** | **0.50-0.69** | **0.0043** | **0.52-0.70** | **0.0072** |
| GDP | 1.02 | 0.52 | 1.02 | 0.48 | 1.08 | 0.55 | **0.76** | **0.29** | 0.91 | 0.41 |
| | 0.86-1.20 | 0.0038 | 0.86-1.22 | 0.0035 | 0.92-1.28 | 0.0017 | **0.62-0.93** | **0.0187** | 0.77-1.07 | 0.0132 |
| Extreme events | 1.07 | 0.50 | 1.06 | 0.46 | 1.06 | 0.46 | 1.07 | 0.54 | 1.07 | 0.50 |
| | 0.85-1.34 | 0.0044 | 0.84-1.32 | 0.0043 | 0.84-1.33 | 0.0045 | 0.83-1.37 | 0.0044 | 0.86-1.32 | 0.0075 |
| Temperature | **1.21** | **0.42** | **1.21** | **0.38** | **1.24** | **0.37** | **1.32** | **0.42** | **1.18** | **0.43** |
| | **1.03-1.41** | **0.0040** | **1.04-1.41** | **0.0038** | **1.07-1.45** | **0.0039** | **1.11-1.57** | **0.0030** | **1.01-1.37** | **0.0072** |
| Rainfall | 1.01 | 0.50 | 1.00 | 0.46 | 1.01 | 0.46 | 1.06 | 0.54 | 1.01 | 0.51 |
| | 0.92-1.11 | 0.0044 | 0.91-1.11 | 0.0043 | 0.91-1.11 | 0.0045 | 0.93-1.22 | 0.0044 | 0.91-1.12 | 0.0074 |
| Relative Humidity | 0.97 | 0.49 | 0.96 | 0.44 | 0.96 | 0.44 | 0.92 | 0.50 | 0.96 | 0.48 |
| | 0.86-1.09 | 0.0044 | 0.85-1.08 | 0.0043 | 0.85-1.08 | 0.0044 | 0.79-1.06 | 0.0043 | 0.86-1.08 | 0.0075 |
| Population Density | 0.92 | 0.49 | 0.90 | 0.44 | 1.00 | 0.46 | 0.96 | 0.54 | 0.93 | 0.49 |
| | 0.75-1.13 | 0.0047 | 0.73-1.09 | 0.0048 | 0.81-1.23 | 0.0045 | 0.76-1.22 | 0.0046 | 0.75-1.15 | 0.0078 |
| Tourist Arrivals | 1.04 | 0.50 | 1.03 | 0.46 | 1.09 | 0.46 | 0.99 | 0.54 | 1.03 | 0.50 |
| | 0.90-1.20 | 0.0040 | 0.89-1.20 | 0.0040 | 0.94-1.27 | 0.0036 | 0.82-1.19 | 0.0045 | 0.88-1.20 | 0.0072 |
| Tourist Departures | 0.93 | 0.47 | 0.94 | 0.43 | 0.91 | 0.41 | **0.82** | **0.43** | 0.90 | 0.45 |
| | 0.83-1.05 | 0.0050 | 0.84-1.06 | 0.0049 | 0.81-1.03 | 0.0054 | **0.71-0.96** | **0.0065** | 0.79-1.01 | 0.0086 |

*spatial (top) and temporal (bottom) random effect

F.

**CR-*Kp***

| | Main analysis | | Sensitivity analysis 1 | | Sensitivity analysis 2 | | Sensitivity analysis 3 | | Sensitivity analysis 4 | |
|---|---|---|---|---|---|---|---|---|---|---|
| Data | (n=413; C=35; Y=14) | | (n=390; C=33; Y=14) | | (n=406; C=34; Y=14) | | (n=295; C=27; Y=14) | | (n=340; C=35; Y=14) | |
| **Variable** | **FE** | **RE *** | **FE** | **RE *** | **FE** | **RE *** | **FE** | **RE *** | **FE** | **RE *** |
| | **M_null** | | | | | | | | | |
| $\hat{\beta}_0$ | 0.02 | 1.92 | 0.02 | 1.89 | 0.02 | 1.93 | 0.04 | 1.22 | 0.03 | 1.93 |
| | 0.01-0.04 | 0.5321 | 0.01-0.05 | 0.5751 | 0.01-0.04 | 0.5278 | 0.02-0.08 | 0.4539 | 0.01-0.05 | 0.4163 |
| | **M_uni** | | | | | | | | | |
| ATB sales of interest | 1.20 | 2.1 | 1.18 | 2.06 | 1.19 | 2.11 | 0.97 | 1.20 | 1.07 | 1.99 |
| | 0.90-1.60 | 0.4722 | 0.88-1.59 | 0.5195 | 0.89-1.60 | 0.4703 | 0.69-1.36 | 0.4631 | 0.79-1.45 | 0.3981 |
| Global ATB sales | 1.20 | 1.94 | 1.13 | 1.91 | 1.20 | 1.95 | 1.27 | 1.33 | 1.25 | 1.96 |
| | 0.84-1.71 | 0.5181 | 0.79-1.62 | 0.5643 | 0.84-1.71 | 0.5141 | 0.84-1.92 | 0.4296 | 0.86-1.81 | 0.3997 |
| GHS index | **0.54** | **1.51** | **0.55** | **1.50** | **0.53** | **1.49** | **0.56** | **0.87** | **0.52** | **1.48** |
| | **0.36-0.82** | **0.5202** | **0.36-0.84** | **0.5622** | **0.35-0.81** | **0.5159** | **0.38-0.82** | **0.4332** | **0.35-0.80** | **0.4044** |
| GDP | **0.56** | **1.43** | **0.61** | **1.48** | **0.55** | **1.43** | **0.60** | **0.92** | **0.56** | **1.43** |
| | **0.39-0.80** | **0.7466** | **0.42-0.88** | **0.7627** | **0.38-0.79** | **0.7443** | **0.41-0.88** | **0.6010** | **0.38-0.81** | **0.6113** |
| Extreme events | 1.36 | 1.83 | 1.33 | 1.81 | 1.35 | 1.84 | 1.25 | 1.17 | 1.35 | 1.84 |
| | 0.85-2.19 | 0.5308 | 0.82-2.16 | 0.5740 | 0.84-2.18 | 0.5266 | 0.80-1.95 | 0.4527 | 0.87-2.11 | 0.4156 |
| Temperature | **1.95** | **1.33** | **1.84** | **1.37** | **1.92** | **1.36** | 1.40 | 1.09 | **1.91** | **1.30** |
| | **1.37-2.78** | **0.4900** | **1.28-2.65** | **0.5343** | **1.34-2.75** | **0.4866** | 0.97-2.01 | 0.4381 | **1.37-2.67** | **0.3797** |
| Rainfall | **1.35** | **1.83** | **1.38** | **1.80** | 1.35 | 1.85 | 1.05 | 1.22 | 1.28 | 1.85 |
| | **1.00-1.83** | **0.5547** | **1.01-1.88** | **0.5991** | 0.88-1.84 | 0.5500 | 0.72-1.51 | 0.4563 | 0.93-1.76 | 0.4317 |
| Relative Humidity | 0.87 | 1.83 | 0.90 | 1.83 | 0.89 | 1.86 | **0.66** | **1.09** | 0.83 | 1.81 |
| | 0.61-1.24 | 0.5296 | 0.63-1.29 | 0.5735 | 0.62-1.27 | 0.5257 | **0.47-0.95** | **0.4626** | 0.60-1.15 | 0.4147 |
| Population Density | 1.12 | 1.92 | 1.12 | 1.90 | 1.10 | 1.93 | 0.96 | 1.22 | 1.07 | 1.94 |
| | 0.71-1.77 | 0.5269 | 0.70-1.79 | 0.5693 | 0.59-1.74 | 0.5233 | 0.61-1.50 | 0.4555 | 0.67-1.70 | 0.4141 |
| Tourist Arrivals | 0.86 | 1.83 | 0.85 | 1.79 | 0.86 | 1.83 | 1.01 | 1.23 | 0.85 | 1.84 |
| | 0.58-1.27 | 0.5439 | 0.57-1.27 | 0.5870 | 0.58-1.27 | 0.5393 | 0.67-1.51 | 0.4533 | 0.58-1.25 | 0.4271 |
| Tourist Departures | **0.67** | **1.52** | 0.73 | 1.60 | **0.68** | **1.55** | 0.74 | 1.05 | **0.67** | **1.54** |
| | **0.46-0.97** | **0.5667** | 0.51-1.05 | 0.6072 | **0.46-0.99** | **0.5602** | 0.51-1.08 | 0.4849 | **0.45-0.97** | **0.4480** |

*spatial (top) and temporal (bottom) random effect

G.

**FR-*Pa***

| | Main analysis | | Sensitivity analysis 1 | | Sensitivity analysis 2 | | Sensitivity analysis 3 | | Sensitivity analysis 4 | |
|---|---|---|---|---|---|---|---|---|---|---|
| Data | (n=472; C=41; Y=14) | | (n=444; C=39; Y=14) | | (n=430; C=37; Y=14) | | (n=412; C=37; Y=14) | | (n=385; C=41; Y=14) | |
| **Variable** | **FE** | **RE *** | **FE** | **RE *** | **FE** | **RE *** | **FE** | **RE *** | **FE** | **RE *** |
| | **M_null** | | | | | | | | | |
| $\widehat{\beta}_0$ | 0.39<br>0.35-0.44 | 0.08<br>0.0154 | 0.39<br>0.35-0.44 | 0.08<br>0.0136 | 0.40<br>0.36-0.45 | 0.07<br>0.0152 | 0.34<br>0.30-0.39 | 0.10<br>0.0072 | 0.39<br>0.35-0.43 | 0.09<br>0.0136 |
| | **M_uni** | | | | | | | | | |
| ATB sales of interest | **1.06<br>1.01-1.11** | **0.07<br>0.0137** | **1.06<br>1.02-1.11** | **0.07<br>0.0118** | **1.05<br>1.00-1.10** | **0.06<br>0.0135** | **1.13<br>1.05-1.22** | **0.07<br>0.0044** | **1.05<br>1.01-1.10** | **0.07<br>0.0120** |
| Global ATB sales | 1.00<br>0.94-1.07 | 0.08<br>0.0155 | 1.00<br>0.94-1.07 | 0.08<br>0.0136 | 0.99<br>0.93-1.06 | 0.07<br>0.0151 | 1.08<br>0.98-1.19 | 0.09<br>0.0077 | 1.01<br>0.94-1.07 | 0.08<br>0.0136 |
| GHS index | **0.84<br>0.78-0.90** | **0.05<br>0.0154** | **0.83<br>0.77-0.89** | **0.05<br>0.0135** | **0.85<br>0.80-0.91** | **0.04<br>0.0152** | **0.85<br>0.77-0.94** | **0.07<br>0.0081** | **0.84<br>0.78-0.90** | **0.05<br>0.0136** |
| GDP | **0.86<br>0.81-0.92** | **0.05<br>0.0073** | **0.87<br>0.81-0.92** | **0.05<br>0.0063** | **0.88<br>0.83-0.94** | **0.05<br>0.0081** | **0.79<br>0.72-0.86** | **0.06<br><1e-04** | **0.87<br>0.81-0.93** | **0.05<br>0.0062** |
| Extreme events | 1.02<br>0.93-1.12 | 0.08<br>0.0154 | 1.02<br>0.93-1.13 | 0.08<br>0.0135 | 1.01<br>0.93-1.11 | 0.07<br>0.0152 | 0.99<br>0.88-1.11 | 0.09<br>0.0072 | 1.03<br>0.94-1.12 | 0.08<br>0.0136 |
| Temperature | 1.06<br>0.99-1.14 | 0.08<br>0.0160 | 1.07<br>0.99-1.15 | 0.08<br>0.0140 | 1.06<br>0.98-1.14 | 0.07<br>0.0156 | 1.06<br>0.96-1.18 | 0.09<br>0.0076 | 1.07<br>1.00-1.15 | 0.08<br>0.0141 |
| Rainfall | 1.03<br>0.97-1.09 | 0.08<br>0.0153 | 1.03<br>0.98-1.09 | 0.08<br>0.0135 | 1.02<br>0.97-1.08 | 0.07<br>0.0151 | 0.96<br>0.87-1.07 | 0.09<br>0.0072 | 1.04<br>0.98-1.10 | 0.09<br>0.0135 |
| Relative Humidity | 0.95<br>0.90-1.01 | 0.07<br>0.0156 | 0.95<br>0.89-1.01 | 0.08<br>0.0137 | 0.96<br>0.90-1.02 | 0.06<br>0.0153 | **0.89<br>0.81-0.98** | **0.08<br>0.0070** | 0.95<br>0.90-1.01 | 0.08<br>0.0138 |
| Population Density | 0.93<br>0.86-1.02 | 0.08<br>0.0151 | 0.92<br>0.84-1.01 | 0.08<br>0.0131 | 0.95<br>0.88-1.04 | 0.07<br>0.0149 | 0.98<br>0.87-1.10 | 0.10<br>0.0071 | 0.94<br>0.86-1.03 | 0.08<br>0.0131 |
| Tourist Arrivals | 1.01<br>0.94-1.09 | 0.08<br>0.0156 | 1.02<br>0.94-1.10 | 0.08<br>0.0138 | 1.03<br>0.96-1.11 | 0.07<br>0.0156 | 1.03<br>0.93-1.14 | 0.09<br>0.0075 | 1.02<br>0.95-1.10 | 0.08<br>0.0138 |
| Tourist Departures | 0.96<br>0.90-1.02 | 0.07<br>0.0150 | 0.97<br>0.91-1.03 | 0.08<br>0.0132 | 0.97<br>0.91-1.04 | 0.06<br>0.0148 | 0.90<br>0.82-1.00 | 0.08<br>0.0064 | 0.94<br>0.88-1.01 | 0.07<br>0.0129 |

*spatial (top) and temporal (bottom) random effect

H.

**CR-*Pa***

| | Main analysis | | Sensitivity analysis 1 | | Sensitivity analysis 2 | | Sensitivity analysis 3 | | Sensitivity analysis 4 | |
|---|---|---|---|---|---|---|---|---|---|---|
| Data | (n=472; C=41; Y=14) | | (n=444; C=39; Y=14) | | (n=430; C=37; Y=14) | | (n=356; C=32; Y=14) | | (n=385; C=41; Y=14) | |
| **Variable** | **FE** | **RE *** | **FE** | **RE *** | **FE** | **RE *** | **FE** | **RE *** | **FE** | **RE *** |
| | **M_null** | | | | | | | | | |
| $\widehat{\beta}_0$ | 0.20<br>0.17-0.24 | 0.27<br>0.0223 | 0.20<br>0.17-0.24 | 0.27<br>0.0229 | 0.21<br>0.18-0.25 | 0.24<br>0.0206 | 0.19<br>0.15-0.23 | 0.30<br>0.0231 | 0.20<br>0.16-0.24 | 0.25<br>0.0279 |
| | **M_uni** | | | | | | | | | |
| ATB sales of interest | 0.97<br>0.90-1.04 | 0.26<br>0.0247 | 0.96<br>0.89-1.04 | 0.26<br>0.0257 | 0.97<br>0.89-1.04 | 0.23<br>0.0228 | 0.96<br>0.82-1.11 | 0.29<br>0.0262 | 0.96<br>0.89-1.04 | 0.24<br>0.0305 |
| Global ATB sales | 0.91<br>0.80-1.03 | 0.29<br>0.0238 | 0.91<br>0.80-1.03 | 0.29<br>0.0246 | **0.89<br>0.79-1.00** | **0.25<br>0.0222** | 1.02<br>0.85-1.22 | 0.30<br>0.0229 | 0.92<br>0.81-1.05 | 0.26<br>0.0292 |
| GHS index | **0.71<br>0.63-0.79** | **0.13<br>0.0218** | **0.69<br>0.62-0.77** | **0.12<br>0.0227** | **0.73<br>0.65-0.82** | **0.13<br>0.0201** | **0.67<br>0.57-0.77** | **0.13<br>0.0191** | **0.71<br>0.63-0.79** | **0.12<br>0.0273** |
| GDP | 0.89<br>0.77-1.04 | 0.20<br>0.0303 | 0.88<br>0.74-1.05 | 0.18<br>0.0323 | 0.89<br>0.77-1.04 | 0.17<br>0.0276 | 0.85<br>0.69-1.06 | 0.20<br>0.0382 | 0.90<br>0.77-1.05 | 0.18<br>0.0365 |
| Extreme events | 1.11<br>0.94-1.31 | 0.26<br>0.0223 | 1.10<br>0.93-1.31 | 0.26<br>0.0230 | 1.09<br>0.92-1.28 | 0.24<br>0.0206 | 0.99<br>0.79-1.23 | 0.30<br>0.0231 | 1.08<br>0.93-1.26 | 0.24<br>0.0279 |
| Temperature | **1.24<br>1.09-1.41** | **0.23<br>0.0199** | **1.24<br>1.09-1.42** | **0.23<br>0.0206** | **1.21<br>1.06-1.39** | **0.21<br>0.0185** | 1.18<br>0.98-1.42 | 0.28<br>0.0196 | **1.22<br>1.08-1.38** | **0.21<br>0.0256** |
| Rainfall | 1.10<br>0.99-1.22 | 0.26<br>0.0229 | 1.10<br>0.99-1.22 | 0.26<br>0.0235 | 1.09<br>0.98-1.21 | 0.24<br>0.0210 | 0.96<br>0.80-1.15 | 0.30<br>0.0229 | 1.11<br>1.00-1.24 | 0.24<br>0.0291 |
| Relative Humidity | **0.89<br>0.80-1.00** | **0.23<br>0.0220** | **0.89<br>0.80-1.00** | **0.23<br>0.0227** | 0.91<br>0.81-1.03 | 0.22<br>0.0204 | **0.76<br>0.65-0.89** | **0.24<br>0.0200** | **0.89<br>0.80-0.98** | **0.21<br>0.0275** |
| Population Density | 0.94<br>0.80-1.10 | 0.27<br>0.0227 | 0.92<br>0.78-1.08 | 0.26<br>0.0234 | 0.97<br>0.83-1.13 | 0.24<br>0.0207 | 1.03<br>0.84-1.26 | 0.30<br>0.0229 | 0.93<br>0.80-1.08 | 0.24<br>0.0283 |
| Tourist Arrivals | 1.10<br>0.96-1.26 | 0.28<br>0.0209 | 1.09<br>0.95-1.25 | 0.27<br>0.0217 | 1.12<br>0.98-1.28 | 0.24<br>0.0190 | 1.12<br>0.93-1.35 | 0.29<br>0.0203 | 1.09<br>0.95-1.25 | 0.25<br>0.0265 |
| Tourist Departures | **0.86<br>0.77-0.96** | **0.21<br>0.0244** | **0.86<br>0.77-0.96** | **0.20<br>0.0251** | **0.87<br>0.78-0.97** | **0.19<br>0.0222** | 0.89<br>0.74-1.07 | 0.27<br>0.0261 | **0.83<br>0.74-0.92** | **0.18<br>0.0308** |

*spatial (top) and temporal (bottom) random effect

I.

| | **Main analysis** | | **Sensitivity analysis 1** | | **Sensitivity analysis 2** | | **Sensitivity analysis 3** | | **Sensitivity analysis 4** | |
|---|---|---|---|---|---|---|---|---|---|---|
| Data | (n=410; C=37; Y=14) | | (n=272; C=28; Y=14) | | (n=396; C=35; Y=14) | | (n=369; C=33; Y=14) | | (n=277; C=37; Y=14) | |
| **Variable** | **FE** | **RE *** | **FE** | **RE *** | **FE** | **RE *** | **FE** | **RE *** | **FE** | **RE *** |
| | **M_null** | | | | | | | | | |
| $\hat{\beta}_0$ | 0.56<br>0.46-0.70 | 0.42<br>0.0012 | 0.65<br>0.54-0.78 | 0.25<br>0.0015 | 0.55<br>0.44-0.69 | 0.43<br>0.0011 | 0.52<br>0.41-0.66 | 0.44<br>0.0016 | 0.57<br>0.46-0.71 | 0.43<br>0.0023 |
| | **M_uni** | | | | | | | | | |
| **ATB sales of interest** | 1.05<br>1.00-1.11 | 0.40<br>0.0014 | **1.05<br>1.00-1.11** | **0.24<br>0.0016** | **1.05<br>1.01-1.11** | **0.41<br>0.0013** | 1.07<br>0.96-1.19 | 0.40<br>0.0017 | **1.08<br>1.02-1.14** | **0.40<br>0.0028** |
| **Global ATB sales** | 0.95<br>0.87-1.03 | 0.43<br>0.0012 | 0.92<br>0.84-1.02 | 0.26<br>0.0015 | 0.94<br>0.86-1.02 | 0.45<br>0.0011 | 0.99<br>0.84-1.17 | 0.44<br>0.0016 | 0.95<br>0.86-1.05 | 0.44<br>0.0023 |
| **GHS index** | **0.72<br>0.60-0.86** | **0.30<br>0.0011** | **0.78<br>0.67-0.91** | **0.18<br>0.0015** | **0.71<br>0.59-0.86** | **0.31<br>0.0011** | **0.64<br>0.53-0.78** | **0.25<br>0.0014** | **0.73<br>0.60-0.88** | **0.32<br>0.0023** |
| **GDP** | 0.97<br>0.89-1.05 | 0.39<br>0.0014 | 0.94<br>0.87-1.02 | 0.22<br>0.0016 | 0.94<br>0.86-1.03 | 0.39<br>0.0014 | 0.85<br>0.71-1.02 | 0.33<br>0.0045 | **0.89<br>0.80-0.99** | **0.33<br>0.0035** |
| **Extreme events** | 1.06<br>0.85-1.32 | 0.41<br>0.0012 | 1.02<br>0.85-1.22 | 0.25<br>0.0015 | 1.07<br>0.85-1.35 | 0.43<br>0.0011 | 1.00<br>0.78-1.28 | 0.43<br>0.0016 | 1.04<br>0.84-1.30 | 0.43<br>0.0023 |
| **Temperature** | 1.07<br>0.93-1.22 | 0.039<br>0.0011 | 1.02<br>0.91-1.15 | 0.24<br>0.0016 | 1.07<br>0.93-1.23 | 0.40<br>0.0011 | **1.22<br>1.02-1.46** | **0.34<br>0.0020** | 1.05<br>0.91-1.22 | 0.40<br>0.0024 |
| **Rainfall** | 0.93<br>0.86-1.01 | 0.42<br>0.0012 | 0.93<br>0.86-1.00 | 0.26<br>0.0016 | 0.93<br>0.86-1.01 | 0.44<br>0.0012 | 0.93<br>0.80-1.10 | 0.43<br>0.0018 | 0.92<br>0.85-1.00 | 0.44<br>0.0025 |
| **Relative Humidity** | 0.93<br>0.85-1.01 | 0.39<br>0.0014 | 0.94<br>0.87-1.02 | 0.24<br>0.0017 | 0.93<br>0.85-1.02 | 0.40<br>0.0014 | 0.85<br>0.72-1.01 | 0.37<br>0.0032 | 0.95<br>0.87-1.04 | 0.41<br>0.0026 |
| **Population Density** | 0.99<br>0.83-1.20 | 0.42<br>0.0012 | 1.09<br>0.92-1.31 | 0.25<br>0.0015 | 1.02<br>0.84-1.24 | 0.44<br>0.0011 | 1.00<br>0.80-1.26 | 0.44<br>0.0016 | 1.03<br>0.83-1.27 | 0.43<br>0.0023 |
| **Tourist Arrivals** | 1.06<br>0.95-1.19 | 0.41<br>0.0011 | 1.01<br>0.91-1.13 | 0.25<br>0.0015 | 1.03<br>0.92-1.16 | 0.43<br>0.0012 | 1.03<br>0.87-1.22 | 0.43<br>0.0016 | 1.03<br>0.90-1.18 | 0.42<br>0.0024 |
| **Tourist Departures** | 0.93<br>0.86-1.02 | 0.39<br>0.0011 | 0.93<br>0.86-1.01 | 0.23<br>0.0013 | 0.92<br>0.84-1.001 | 0.39<br>0.0011 | **0.83<br>0.71-0.98** | **0.37<br>0.0013** | **0.90<br>0.82-0.99** | **0.39<br>0.0020** |

**FR-*Ab***

*spatial (top) and temporal (bottom) random effect

J.

| | **Main analysis** | | **Sensitivity analysis 1** | | **Sensitivity analysis 2** | | **Sensitivity analysis 3** | | **Sensitivity analysis 4** | |
|---|---|---|---|---|---|---|---|---|---|---|
| Data | (n=410; C=37; Y=14) | | (n=272; C=28; Y=14) | | (n=396; C=35; Y=14) | | (n=292; C=29; Y=14) | | (n=277; C=37; Y=14) | |
| **Variable** | **FE** | **RE *** | **FE** | **RE *** | **FE** | **RE *** | **FE** | **RE *** | **FE** | **RE *** |
| | **M_null** | | | | | | | | | |
| $\hat{\beta}_0$ | 0.41<br>0.30-0.54 | 0.67<br>0.0446 | 0.47<br>0.36-0.63 | 0.47<br>0.0394 | 0.40<br>0.29-0.54 | 0.71<br>0.0387 | 0.38<br>0.27-0.54 | 0.72<br>0.0366 | 0.42<br>0.31-0.56 | 0.67<br>0.0460 |
| | **M_uni** | | | | | | | | | |
| **ATB sales of interest** | **1.13<br>1.04-1.24** | **0.77<br>0.0328** | **1.17<br>1.07-1.27** | **0.15<br>0.0450** | **1.12<br>1.03-1.22** | **0.79<br>0.0293** | 1.14<br>0.97-1.33 | 0.80<br>0.0258 | **1.13<br>1.01-1.26** | **0.75<br>0.0351** |
| **Global ATB sales** | **0.84<br>0.74-0.96** | **0.74<br>0.0509** | **0.81<br>0.69-0.94** | **0.51<br>0.0468** | **0.83<br>0.73-0.95** | **0.78<br>0.0448** | 0.82<br>0.65-1.03 | 0.77<br>0.0422 | **0.83<br>0.71-0.97** | **0.72<br>0.0527** |
| **GHS index** | **0.69<br>0.55-0.88** | **0.52<br>0.0442** | **0.73<br>0.59-0.91** | **0.37<br>0.0390** | **0.68<br>0.53-0.87** | **0.54<br>0.0384** | **0.57<br>0.44-0.73** | **0.39<br>0.0348** | **0.69<br>0.54-0.88** | **0.52<br>0.0455** |
| **GDP** | **1.25<br>1.02-1.54** | **0.98<br>0.0258** | 1.21<br>0.98-1.49 | 0.68<br>0.0246 | 1.15<br>0.93-1.41 | 0.89<br>0.0279 | 0.82<br>0.60-1.12 | 0.49<br>0.0545 | 0.95<br>0.76-1.19 | 0.61<br>0.0509 |
| **Extreme events** | 1.13<br>0.86-1.50 | 0.66<br>0.0446 | 1.08<br>0.84-1.37 | 0.47<br>0.0394 | 1.15<br>0.86-1.54 | 0.69<br>0.0387 | 1.09<br>0.81-1.47 | 0.71<br>0.0366 | 1.11<br>0.84-1.46 | 0.66<br>0.0460 |
| **Temperature** | **1.21<br>1.00-1.46** | **0.57<br>0.0429** | 1.17<br>0.99-1.38 | 0.39<br>0.0382 | **1.22<br>1.01-1.49** | **0.58<br>0.0371** | **1.39<br>1.12-1.72** | **0.54<br>0.0342** | **1.22<br>1.01-1.48** | **0.57<br>0.0441** |
| **Rainfall** | 1.00<br>0.89-1.12 | 0.67<br>0.0446 | 0.99<br>0.88-1.11 | 0.47<br>0.0394 | 1.01<br>0.90-1.13 | 0.71<br>0.0387 | 1.01<br>0.83-1.23 | 0.71<br>0.0366 | 0.99<br>0.87-1.12 | 0.67<br>0.0459 |
| **Relative Humidity** | 0.94<br>0.82-1.08 | 0.64<br>0.0447 | 0.97<br>0.85-1.11 | 0.46<br>0.0395 | 0.95<br>0.83-1.09 | 0.67<br>0.0388 | **0.81<br>0.67-0.99** | **0.58<br>0.0404** | 0.96<br>0.83-1.10 | 0.64<br>0.0458 |
| **Population Density** | 0.86<br>0.67-1.10 | 0.67<br>0.0464 | 1.06<br>0.83-1.37 | 0.47<br>0.0386 | 0.91<br>0.70-1.18 | 0.70<br>0.0398 | 1.03<br>0.78-1.37 | 0.72<br>0.0363 | 0.94<br>0.72-1.22 | 0.66<br>0.0466 |
| **Tourist Arrivals** | **1.22<br>1.04-1.45** | **0.71<br>0.0410** | 1.12<br>0.95-1.33 | 0.49<br>0.0379 | 1.18<br>0.99-1.40 | 0.73<br>0.0365 | 1.08<br>0.86-1.36 | 0.72<br>0.0355 | 1.15<br>0.95-1.40 | 0.68<br>0.0439 |
| **Tourist Departures** | 0.95<br>0.84-1.08 | 0.64<br>0.0454 | 0.89<br>0.79-1.01 | 0.42<br>0.0410 | 0.89<br>0.78-1.01 | 0.63<br>0.0406 | **0.81<br>0.67-0.98** | **0.59<br>0.0377** | 0.90<br>0.79-1.04 | 0.62<br>1.04 |

**CR-*Ab***

*spatial (top) and temporal (bottom) random effect

K.

**VR-*E***

| | Main analysis | | Sensitivity analysis 1 | | Sensitivity analysis 2 | | Sensitivity analysis 3 | | Sensitivity analysis 4 | |
|---|---|---|---|---|---|---|---|---|---|---|
| Data | (n=423; C=37; Y=14) | | (n=325; C=28; Y=14) | | (n=394; C=34; Y=14) | | (n=333; C=30; Y=14) | | (n=326; C=37; Y=14) | |
| **Variable** | **FE** | **RE *** | **FE** | **RE *** | **FE** | **RE *** | **FE** | **RE *** | **FE** | **RE *** |
| | **M_null** | | | | | | | | | |
| $\widehat{\beta}_0$ | 0.08<br>0.06-0.11 | 0.67<br>0.0430 | 0.10<br>0.07-0.13 | 0.53<br>0.0310 | 0.08<br>0.06-0.11 | 0.73<br>0.0364 | 0.09<br>0.06-0.13 | 0.77<br>0.0116 | 0.08<br>0.06-0.11 | 0.63<br>0.0403 |
| | **M_uni** | | | | | | | | | |
| **ATB sales of interest** | 1.20<br>0.96-1.48 | 0.62<br>0.0361 | 1.13<br>0.91-1.41 | 0.50<br>0.0360 | 1.19<br>0.95-1.49 | 0.67<br>0.0313 | 1.03<br>0.89-1.19 | 0.76<br>0.0110 | 1.19<br>0.94-1.51 | 0.59<br>0.0349 |
| **Global ATB sales** | 1.14<br>0.90-1.42 | 0.68<br>0.0387 | 1.10<br>0.87-1.40 | 0.54<br>0.0367 | 1.14<br>0.90-1.45 | 0.74<br>0.0328 | 0.94<br>0.75-1.19 | 0.75<br>0.0116 | 1.03<br>0.82-1.29 | 0.63<br>0.0393 |
| **GHS index** | 1.13<br>0.86-1.49 | 0.65<br>0.0431 | 1.17<br>0.90-1.53 | 0.50<br>0.0400 | 1.15<br>0.86-1.55 | 0.70<br>0.0365 | 1.06<br>0.75-1.49 | 0.77<br>0.0117 | 1.15<br>0.88-1.51 | 0.61<br>0.0406 |
| **GDP** | 1.14<br>0.93-1.39 | 0.67<br>0.0363 | 1.14<br>0.95-1.37 | 0.51<br>0.0358 | 1.12<br>0.91-1.38 | 0.71<br>0.0322 | 1.09<br>0.92-1.29 | 0.77<br>0.0101 | 1.08<br>0.88-1.33 | 0.63<br>0.0372 |
| **Extreme events** | 0.82<br>0.62-1.08 | 0.62<br>0.0427 | 0.79<br>0.61-1.02 | 0.47<br>0.0398 | 0.81<br>0.60-1.10 | 0.67<br>0.0361 | 0.92<br>0.68-1.24 | 0.76<br>0.0117 | 0.83<br>0.65-1.07 | 0.58<br>0.0396 |
| **Temperature** | 0.87<br>0.68-1.12 | 0.67<br>0.0430 | 0.86<br>0.68-1.08 | 0.52<br>0.0390 | 0.85<br>0.64-1.11 | 0.73<br>0.0359 | 1.25<br>0.96-1.63 | 0.71<br>0.0103 | 0.85<br>0.66-1.09 | 0.63<br>0.0398 |
| **Rainfall** | 0.94<br>0.76-1.17 | 0.67<br>0.0429 | 0.97<br>0.78-1.20 | 0.52<br>0.0399 | 0.95<br>0.76-1.20 | 0.72<br>0.0364 | 1.03<br>0.81-1.32 | 0.77<br>0.116 | 0.93<br>0.74-1.17 | 0.63<br>0.0402 |
| **Relative Humidity** | 1.13<br>0.89-1.42 | 0.71<br>0.0428 | 1.19<br>0.95-1.49 | 0.55<br>0.0390 | 1.16<br>0.91-1.48 | 0.78<br>0.0358 | 0.88<br>0.69-1.13 | 0.74<br>0.0113 | 1.09<br>0.88-1.37 | 0.64<br>0.0402 |
| **Population Density** | 0.81<br>0.62-1.06 | 0.64<br>0.0439 | 0.82<br>0.62-1.09 | 0.50<br>0.0407 | 0.81<br>0.61-1.09 | 0.68<br>0.0370 | 0.86<br>0.62-1.20 | 0.76<br>0.0121 | 0.81<br>0.62-1.06 | 0.58<br>0.0405 |
| **Tourist Arrivals** | 1.18<br>0.89-1.56 | 0.75<br>0.0422 | 1.02<br>0.79-1.33 | 0.54<br>0.0400 | 1.18<br>0.88-1.57 | 0.80<br>0.0361 | 1.05<br>0.74-1.48 | 0.80<br>0.0114 | 1.08<br>0.84-1.40 | 0.65<br>0.0403 |
| **Tourist Departures** | 1.09<br>0.88-1.36 | 0.69<br>0.0430 | 1.15<br>0.94-1.40 | 0.52<br>0.0403 | 1.08<br>0.86-1.37 | 0.74<br>0.0365 | 1.00<br>0.79-1.26 | 0.77<br>0.0116 | 1.05<br>0.84-1.32 | 0.63<br>0.0404 |

*spatial (top) and temporal (bottom) random effect

L.

**PR-*Sp***

| | Main analysis | | Sensitivity analysis 1 | | Sensitivity analysis 2 | | Sensitivity analysis 3 | | Sensitivity analysis 4 | |
|---|---|---|---|---|---|---|---|---|---|---|
| Data | (n=437; C=38; Y=14) | | (n=310; C=30; Y=14) | | (n=396; C=34; Y=14) | | (n=340; C=31; Y=14) | | (n=310; C=37; Y=14) | |
| **Variable** | **FE** | **RE *** | **FE** | **RE *** | **FE** | **RE *** | **FE** | **RE *** | **FE** | **RE *** |
| | **M_null** | | | | | | | | | |
| $\widehat{\beta}_0$ | 0.30<br>0.25-0.36 | 0.28<br>0.0053 | 0.31<br>0.25-0.38 | 0.32<br>0.0055 | 0.33<br>0.28-0.29 | 0.21<br>0.0053 | 0.23<br>0.18-0.29 | 0.39<br>0.0102 | 0.31<br>0.26-0.37 | 0.26<br>0.0071 |
| | **M_uni** | | | | | | | | | |
| **ATB sales of interest** | 1.03<br>0.92-1.16 | 0.27<br>0.0054 | 1.04<br>0.91-1.18 | 0.32<br>0.0056 | 1.02<br>0.91-1.14 | 0.21<br>0.0053 | 1.00<br>0.84-1.19 | 0.39<br>0.0102 | 1.05<br>0.93-1.19 | 0.26<br>0.0072 |
| **Global ATB sales** | **1.12**<br>**1.01-1.25** | **0.26**<br>**0.0053** | 1.12<br>0.99-1.26 | 0.31<br>0.0055 | 1.09<br>0.98-1.21 | 0.21<br>0.0052 | 1.11<br>0.94-1.31 | 0.40<br>0.0102 | **1.12**<br>**1.00-1.25** | **0.25**<br>**0.0071** |
| **GHS index** | 0.93<br>0.78-1.09 | 0.27<br>0.0053 | 0.91<br>0.75-1.11 | 0.31<br>0.0055 | 0.99<br>0.85-1.16 | 0.21<br>0.0053 | 0.84<br>0.67-1.05 | 0.35<br>0.0105 | 0.91<br>0.77-1.07 | 0.25<br>0.0071 |
| **GDP** | 0.93<br>0.84-1.03 | 0.26<br>0.0047 | **0.87**<br>**0.78-0.96** | **0.28**<br>**0.0046** | 0.93<br>0.85-1.03 | 0.19<br>0.0046 | **0.72**<br>**0.62-0.82** | **0.26**<br>**0.0027** | 0.94<br>0.85-1.05 | 0.24<br>0.0064 |
| **Extreme events** | 1.15<br>0.97-1.37 | 0.26<br>0.0053 | 1.16<br>0.95-1.41 | 0.30<br>0.0055 | 1.12<br>0.95-1.32 | 0.20<br>0.0053 | 1.22<br>0.99-1.49 | 0.34<br>0.0104 | 1.16<br>0.98-1.38 | 0.24<br>0.0071 |
| **Temperature** | 1.11<br>0.97-1.26 | 0.26<br>0.0060 | 1.08<br>0.94-1.25 | 0.31<br>0.0061 | 1.06<br>0.93-1.20 | 0.20<br>0.0056 | **1.33**<br>**1.12-1.58** | **0.29**<br>**0.0120** | 1.09<br>0.95-1.24 | 0.25<br>0.0077 |
| **Rainfall** | 0.97<br>0.88-1.07 | 0.28<br>0.0054 | 0.95<br>0.86-1.05 | 0.33<br>0.0056 | 0.96<br>0.87-1.07 | 0.21<br>0.0053 | 1.02<br>0.85-1.22 | 0.38<br>0.0103 | 1.01<br>0.92-1.10 | 0.26<br>0.0071 |
| **Relative Humidity** | **0.89**<br>**0.81-0.98** | **0.23**<br>**0.0062** | **0.88**<br>**0.80-0.97** | **0.27**<br>**0.0066** | **0.91**<br>**0.83-0.99** | **0.18**<br>**0.0060** | **0.80**<br>**0.69-0.92** | **0.29**<br>**0.0110** | **0.89**<br>**0.80-0.99** | **0.22**<br>**0.0080** |
| **Population Density** | 0.90<br>0.76-1.06 | 0.26<br>0.0053 | 0.86<br>0.71-1.05 | 0.30<br>0.0054 | 0.98<br>0.84-1.15 | 0.21<br>0.0053 | 0.80<br>0.63-1.02 | 0.36<br>0.0098 | 0.88<br>0.74-1.04 | 0.25<br>0.0071 |
| **Tourist Arrivals** | 0.88<br>0.76-1.02 | 0.28<br>0.0048 | **0.84**<br>**0.71-0.98** | **0.31**<br>**0.0046** | 0.90<br>0.78-1.03 | 0.21<br>0.0048 | 0.84<br>0.69-1.01 | 0.36<br>0.0085 | 0.93<br>0.81-1.06 | 0.25<br>0.0066 |
| **Tourist Departures** | 0.92<br>0.82-1.02 | 0.25<br>0.0049 | 0.91<br>0.81-1.03 | 0.29<br>0.0050 | 0.93<br>0.83-1.04 | 0.19<br>0.0049 | 0.85<br>0.71-1.03 | 0.30<br>0.0095 | **0.89**<br>**0.79-1.00** | **0.22**<br>**0.0062** |

*spatial (top) and temporal (bottom) random effect

M.

**MLR-*Sp***

| | Main analysis | | Sensitivity analysis 1 | | Sensitivity analysis 2 | | Sensitivity analysis 3 | | Sensitivity analysis 4 | |
|---|---|---|---|---|---|---|---|---|---|---|
| Data | (n=429; C=38; Y=14) | | (n=297; C=29; Y=14) | | (n=388; C=34; Y=14) | | (n=385; C=34; Y=14) | | (n=307; C=37; Y=14) | |
| **Variable** | **FE** | **RE *** | **FE** | **RE *** | **FE** | **RE *** | **FE** | **RE *** | **FE** | **RE *** |
| | **M_null** | | | | | | | | | |
| $\hat{\beta}_0$ | 0.24 | 0.46 | 0.26 | 0.51 | 0.27 | 0.36 | 0.17 | 0.67 | 0.26 | 0.42 |
| | 0.19-0.30 | 0.0001 | 0.20-0.34 | 0.0012 | 0.22-0.33 | 0.0003 | 0.13-0.23 | <1e-04 | 0.21-0.32 | 0.0025 |
| | **M_uni** | | | | | | | | | |
| ATB sales of interest | **1.13** | **0.47** | **1.16** | **0.52** | **1.11** | **0.38** | 0.98 | 0.66 | 1.08 | 0.43 |
| | **1.04-1.23** | **<1e-04** | **1.06-1.26** | **0.0008** | **1.02-1.21** | **0.0002** | 0.86-1.13 | <1e-04 | 0.99-1.18 | 0.0015 |
| Global ATB sales | **1.16** | **0.44** | 1.10 | 0.49 | 1.12 | 0.36 | 1.13 | 0.67 | 1.12 | 0.41 |
| | **1.02-1.32** | **0.0003** | 0.96-1.27 | 0.0014 | 0.99-1.27 | 0.0005 | 0.92-1.38 | <1e-04 | 0.98-1.27 | 0.0029 |
| GHS index | 0.95 | 0.45 | 0.93 | 0.50 | 1.03 | 0.36 | 0.91 | 0.65 | 0.94 | 0.41 |
| | 0.76-1.18 | 0.0001 | 0.72-1.19 | 0.0012 | 0.84-1.26 | 0.0003 | 0.68-1.22 | <1e-04 | 0.76-1.15 | 0.0025 |
| GDP | **0.90** | **0.44** | **0.86** | **0.48** | **0.89** | **0.35** | **0.81** | **0.56** | **0.87** | **0.39** |
| | **0.83-0.98** | **<1e-04** | **0.79-0.94** | **0.0002** | **0.81-0.96** | **<1e-04** | **0.68-0.96** | **<1e-04** | **0.79-0.95** | **0.0005** |
| Extreme events | **1.25** | **0.41** | 1.25 | 0.45 | 1.21 | 0.32 | **1.37** | **0.55** | **1.24** | **0.37** |
| | **1.00-1.57** | **0.0001** | 0.97-1.61 | 0.0012 | 0.98-1.49 | 0.0003 | **1.04-1.81** | **<1e-04** | **1.01-1.54** | **0.0026** |
| Temperature | 0.90 | 0.48 | 0.86 | 0.54 | **0.86** | **0.37** | 1.06 | 0.65 | 0.88 | 0.44 |
| | 0.77-1.05 | <1e-04 | 0.72-1.03 | 0.003 | **0.74-1.00** | **<1e-04** | 0.83-1.36 | <1e-04 | 0.75-1.04 | 0.0017 |
| Rainfall | 0.98 | 0.46 | 1.00 | 0.51 | 0.96 | 0.35 | 0.90 | 0.68 | 1.02 | 0.42 |
| | 0.87-1.10 | 0.0001 | 0.90-1.11 | 0.0012 | 0.86-1.09 | 0.0003 | 0.73-1.12 | <1e-04 | 0.92-1.13 | 0.0025 |
| Relative Humidity | 1.01 | 0.47 | 1.02 | 0.53 | 1.03 | 0.37 | 0.83 | 0.54 | 1.00 | 0.42 |
| | 0.91-1.13 | <1e-04 | 0.91-1.14 | 0.0011 | 0.92-1.15 | 0.0002 | 0.67-1.01 | <1e-04 | 0.89-1.12 | 0.0025 |
| Population Density | 0.91 | 0.46 | 0.83 | 0.49 | 0.99 | 0.36 | 0.81 | 0.64 | 0.87 | 0.41 |
| | 0.73-1.13 | <1e-04 | 0.64-1.08 | 0.0011 | 0.81-1.22 | 0.0003 | 0.60-1.10 | <1e-04 | 0.70-1.09 | 0.0024 |
| Tourist Arrivals | **0.83** | **0.47** | **0.77** | **0.49** | **0.83** | **0.40** | 0.87 | 0.68 | 0.88 | 0.43 |
| | **0.70-0.98** | **<1e-04** | **0.64-0.93** | **0.0004** | **0.70-0.99** | **<1e-04** | 0.70-1.09 | <1e-04 | 0.75-1.04 | 0.0019 |
| Tourist Departures | 0.93 | 0.43 | 0.93 | 0.48 | 0.95 | 0.35 | 0.97 | 0.64 | 0.92 | 0.39 |
| | 0.82-1.05 | <1e-04 | 0.81-1.06 | 0.0009 | 0.84-1.07 | 0.0002 | 0.78-1.20 | <1e-04 | 0.80-1.04 | 0.0019 |

*spatial (top) and temporal (bottom) random effect

***Supplementary Table S7. Multivariable full model results (before backward selection) for the main analysis, by drug-bug pair.***

Estimated coefficients from multivariable **full** analyses (before backward selection), compared to multivariable **final** analyses (after backward selection) using the mixed-effect negative binomial model, for:

**FR-Ec**: fluoroquinolone-resistant *E. coli*
**APR-Ec**: aminopenicillin-resistant *E. coli*
**3GCR-Ec**: third generation cephalosporin-resistant *E. coli*
**FR-Kp**: fluoroquinolone-resistant *K. pneumoniae*
**3GCR-Kp**: third generation cephalosporin-resistant *K. pneumoniae*
**CR-Kp**: carbapenem-resistant *K. pneumoniae*
**FR-Pa**: fluoroquinolone-resistant *P. aeruginosa*
**CR-Pa**: carbapenem-resistant *P. aeruginosa*
**FR-Ab**: fluoroquinolone-resistant *A. baumannii*
**CR-Ab**: carbapenem-resistant *A. baumannii*
**VR-E**: vancomycin-resistant Enterococci
**PR-Sp**: penicillin-non-susceptible *S. pneumoniae*
**MLR-Sp**: macrolide-resistant *S. pneumoniae*

Data represent n, the number of observations; C, the number of countries and Y, the number of years. Fixed effects (FE) are reported in the exponential form. Random effects (RE) are reported as variances: spatial (top) and temporal (bottom).

Antibiotic sales of interest for each drug-bug pair were the following:

quinolones (FR-*Ec*); broad-spectrum penicillins (APR-*Ec* and PR-*Sp*); third generation cephalosporins (3GCR-*Ec* and 3GCR-*Kp*); carbapenems (CR-*Kp*; CR-*Pa* and CR-*Ab*); glycopeptides (VR-*E*) and macrolides (MLR-*Sp*).

Covariables that entered the full model were selected from covariables associated with a p-value <20% in the univariate model.

Covariables that are statistically significant (p-value < 5%) are highlighted in bold.

| | FR-*Ec* | | APR-*Ec* | | 3GCR-*Ec* | | FR-*Kp* | | 3GCR-*Kp* | | CR-*Kp* | | FR-*Pa* | | CR-*Pa* | | FR-*Ab* | | CR-*Ab* | | VR-*E* | | PR-*Sp* | | MLR-*Sp* | |
|---|---|---|---|---|---|---|---|---|---|---|---|---|---|---|---|---|---|---|---|---|---|---|---|---|---|---|
| Data * | n=471; C=41; Y=14 | | n=471; C=41; Y=14 | | n=471; C=41; Y=14 | | n=467; C=41; Y=14 | | n=467; C=41; Y=14 | | n=413; C=35; Y=14 | | n=472; C=41; Y=14 | | n=472; C=41; Y=14 | | n=410; C=37; Y=14 | | n=410; C=37; Y=14 | | n=423; C=37; Y=14 | | n=437; C=38; Y=14 | | n=429; C=38; Y=14 | |
| Model ** | **M_full** | M_multi | **M_full** | M_multi | **M_full** | M_multi | **M_full** | M_multi | **M_full** | M_multi | **M_full** | M_multi | **M_full** | M_multi | **M_full** | M_multi | **M_full** | M_multi | **M_full** | M_multi | **M_full** | M_multi | **M_full** | M_multi | **M_full** | M_multi |
| Fixed Effects $e^{\beta}$ [95% CI] | | | | | | | | | | | | | | | | | | | | | | | | | | |
| $\hat{\beta}_0$ | 0.33 0.30-0.36 | 0.33 0.30-0.36 | 0.65 0.63-0.67 | 0.65 0.63-0.68 | 0.21 0.18-0.23 | 0.21 0.18-0.23 | 0.25 0.21-0.30 | 0.25 0.21-0.30 | 0.31 0.26-0.36 | 0.31 0.26-0.36 | 0.02 0.01-0.04 | 0.02 0.01-0.04 | 0.39 0.36-0.42 | 0.39 0.36-0.42 | 0.19 0.17-0.22 | 0.19 0.17-0.22 | 0.55 0.47-0.65 | 0.55 0.46-0.66 | 0.39 0.29-0.52 | 0.39 0.29-0.53 | 0.08 0.06-0.11 | 0.08 0.06-0.11 | 0.30 0.26-0.35 | 0.30 0.26-0.36 | 0.25 0.20-0.30 | 0.24 0.20-0.30 |
| **ATB sales of interest** | **1.10 1.05-1.16** | 1.11 1.06-1.17 | - | - | - | - | 1.06 0.97-1.15 | - | - | - | - | - | **1.06 1.01-1.10** | 1.06 1.01-1.10 | - | - | 1.05 0.99-1.10 | - | **1.14 1.05-1.24** | 1.16 1.06-1.26 | 1.17 0.95-1.45 | - | - | - | 1.08 0.98-1.18 | - |
| **Global ATB sales** | - | - | - | - | - | - | - | - | - | - | - | - | - | - | 0.93 0.84-1.04 | - | - | - | **0.82 0.72-0.93** | 0.82 0.72-0.93 | 1.06 0.84-1.33 | - | **1.15 1.04-1.28** | 1.12 1.01-1.24 | **1.18 1.03-1.35** | 1.22 1.08-1.38 |
| **GHS index** | **0.91 0.83-0.99** | 0.88 0.80-0.96 | **0.96 0.92-1.00** | 0.95 0.92-0.99 | **0.83 0.73-0.95** | 0.81 0.72-0.91 | **0.68 0.57-0.81** | 0.63 0.54-0.74 | **0.63 0.54-0.74** | 0.61 0.52-0.71 | 0.71 0.45-1.10 | 0.64 0.44-0.94 | **0.90 0.84-0.96** | 0.89 0.83-0.96 | **0.73 0.63-0.84** | 0.73 0.66-0.82 | **0.73 0.62-0.87** | 0.71 0.59-0.85 | **0.62 0.45-0.84** | 0.58 0.42-0.79 | - | - | - | - | - | - |
| **GDP** | 0.96 0.90-1.03 | - | 0.98 0.95-1.01 | - | 0.97 0.86-1.10 | - | 0.96 0.83-1.12 | - | - | - | 0.88 0.54-1.43 | - | **0.92 0.85-0.99** | 0.92 0.86-0.99 | 1.11 0.96-1.30 | - | - | - | **1.19 1.00-1.42** | 1.23 1.04-1.47 | - | - | 0.96 0.86-1.06 | - | 0.93 0.85-1.02 | 0.89 0.82-0.97 |
| **Extreme events** | **1.12 1.02-1.23** | 1.16 1.05-1.27 | **1.04 1.00-1.08** | 1.05 1.01-1.09 | **1.16 1.03-1.31** | 1.20 1.06-1.36 | - | - | - | - | 1.05 0.69-1.58 | - | - | - | - | - | - | - | - | - | 0.88 0.66-1.17 | - | 1.09 0.92-1.29 | - | **1.34 1.08-1.68** | 1.28 1.03-1.58 |
| **Temperature** | 1.08 0.99-1.19 | 1.13 1.05-1.23 | 1.03 0.99-1.08 | 1.04 1.00-1.08 | 1.08 0.94-1.23 | 1.15 1.03-1.29 | 1.09 0.93-1.27 | - | 1.10 0.97-1.25 | - | **1.54 1.04-2.28** | 1.76 1.25-2.48 | 0.96 0.90-1.03 | - | 1.06 0.91-1.24 | - | - | - | 1.11 0.91-1.37 | - | - | - | 1.02 0.89-1.16 | - | 0.87 0.76-1.01 | - |
| **Rainfall** | **1.07 1.00-1.14** | - | - | - | 1.06 0.95-1.18 | - | - | - | - | - | 1.14 0.86-1.52 | - | - | - | 1.05 0.94-1.17 | - | 0.94 0.86-1.02 | 0.92 0.85-0.99 | - | - | - | - | - | - | - | - |
| **Relative Humidity** | 0.95 0.88-1.04 | - | 0.99 0.96-1.03 | - | 0.92 0.81-1.05 | - | 0.96 0.85-1.09 | - | - | - | - | - | 0.97 0.92-1.03 | - | 0.94 0.82-1.07 | - | 0.97 0.88-1.07 | - | - | - | - | - | 0.93 0.84-1.02 | 0.89 0.81-0.98 | - | - |
| **Population Density** | - | - | - | - | - | - | - | - | - | - | - | - | 0.97 0.92-1.03 | - | - | - | - | - | - | - | 0.83 0.64-1.08 | - | 0.91 0.78-1.05 | - | - | - |
| **Tourist Arrivals** | 0.96 0.89-1.03 | - | 0.98 0.94-1.01 | - | 0.94 0.84-1.04 | - | - | - | - | - | - | - | - | - | **1.13 1.01-1.26** | - | - | - | 1.14 0.97-1.35 | - | - | - | 0.90 0.79-1.03 | - | 0.88 0.75-1.03 | - |
| **Tourist Departures** | 0.99 0.92-1.06 | - | 0.97 0.94-1.01 | 0.96 0.93-1.00 | 0.99 0.88-1.11 | - | - | - | - | - | 0.90 0.60-1.36 | - | 0.99 0.93-1.05 | - | **0.87 0.78-0.98** | 0.89 0.81-0.98 | 0.94 0.87-1.02 | - | - | - | - | - | 0.96 0.86-1.08 | - | - | - |
| Variance of Random Effects $\hat{\sigma}^2$ | | | | | | | | | | | | | | | | | | | | | | | | | | |
| $\hat{\sigma}^2_{b_c}$ | 0.06 | 0.07 | 0.01 | 0.01 | 0.11 | 0.13 | 0.22 | 0.27 | 0.23 | 0.24 | 1.13 | 1.13 | 0.03 | 0.03 | 0.13 | 0.11 | 0.26 | 0.30 | 0.71 | 0.80 | 0.57 | 0.67 | 0.19 | 0.22 | 0.36 | 0.36 |
| $\hat{\sigma}^2_{\phi_t}$ | 0.0001 | 0.0003 | 0.0002 | 0.0003 | 0.0113 | 0.0123 | 0.0199 | 0.0169 | 0.0040 | 0.0042 | 0.5570 | 0.4855 | 0.0085 | 0.0093 | 0.0161 | 0.0233 | 0.0014 | 0.0012 | 0.0243 | 0.0226 | 0.0357 | 0.0430 | 0.0047 | 0.0062 | $<1^{e}$-04 | $<1^{e}$-04 |
| AIC | 3374.6 | 3373.0 | 3294.1 | 3291.5 | 3397.8 | 3391.5 | 3304.0 | 3300.9 | 3380.8 | 3380.9 | 1906.5 | 1900.5 | 3186.0 | 3180.9 | 3136.2 | 3134.8 | 2633.5 | 2632.9 | 2737.2 | 2736.6 | 2208.2 | 2206.5 | 2630.7 | 2628.0 | 2512.9 | 2515.1 |

***Supplementary Figure S11.*** **Spatial Random Effects distribution, by drug-bug pair.**

Spatial Random Effects distribution for:

A. **FR-Ec**: fluoroquinolone-resistant *E. coli*
B. **APR-Ec**: aminopenicillin-resistant *E. coli*
C. **3GCR-Ec**: third generation cephalosporin-resistant *E. coli*
D. **FR-Kp**: fluoroquinolone-resistant *K. pneumoniae*
E. **3GCR-Kp**: third generation cephalosporin-resistant *K. pneumoniae*
F. **CR-Kp**: carbapenem-resistant *K. pneumoniae*
G. **FR-Pa**: fluoroquinolone-resistant *P. aeruginosa*
H. **CR-Pa**: carbapenem-resistant *P. aeruginosa*
I. **FR-Ab**: fluoroquinolone-resistant *A. baumannii*
J. **CR-Ab**: carbapenem-resistant *A. baumannii*
K. **VR-E**: vancomycin-resistant Enterococci
L. **PR-Sp**: penicillin-non-susceptible *S. pneumoniae*
M. **MLR-Sp**: macrolide-resistant *S. pneumoniae*

Main plot shows the spatial Random Effects distribution per country from the final model M_multi. Small right bottom plot represents the density of the spatial Random Effects for both M_null (blue) and M_multi (red).

Countries are colored based on world’s regions (from World Bank indicators):

East Asia & Pacific
Europe & Central Asia
Latin America & Caribbean
Middle East & North Africa
North America
South Asia
Sub–Saharan Africa

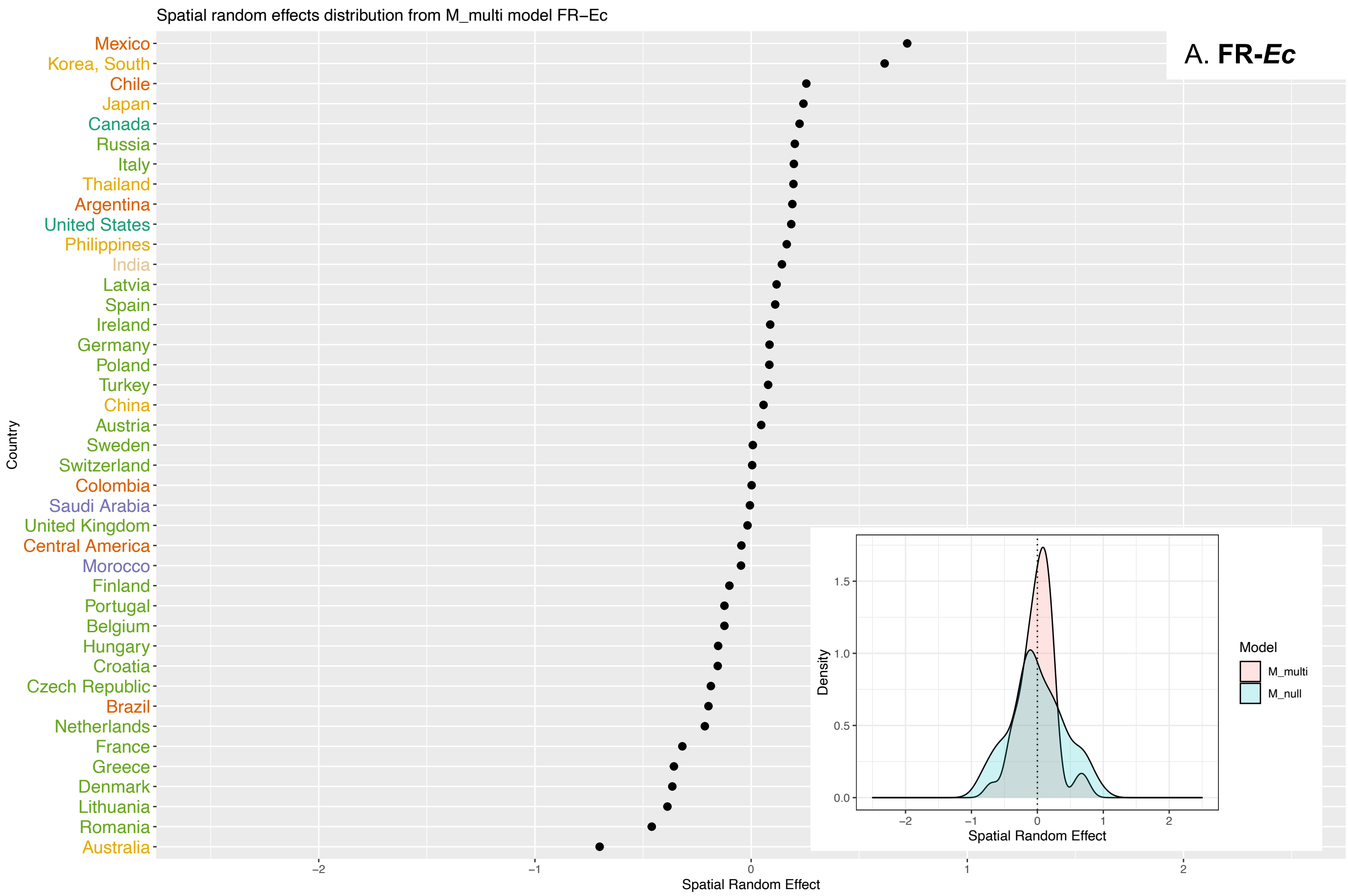
Spatial random effects distribution from M_multi model FR−Ec
A. FR-*Ec*
Mexico
Korea, South
Chile
Japan
Canada
Russia
Italy
Thailand
Argentina
United States
Philippines
India
Latvia
Spain
Ireland
Germany
Poland
Turkey
China
Austria
Sweden
Switzerland
Colombia
Saudi Arabia
United Kingdom
Central America
Morocco
Finland
Portugal
Belgium
Hungary
Croatia
Czech Republic
Brazil
Netherlands
France
Greece
Denmark
Lithuania
Romania
Australia
Country
Spatial Random Effect
Density
Model
M_multi
M_null

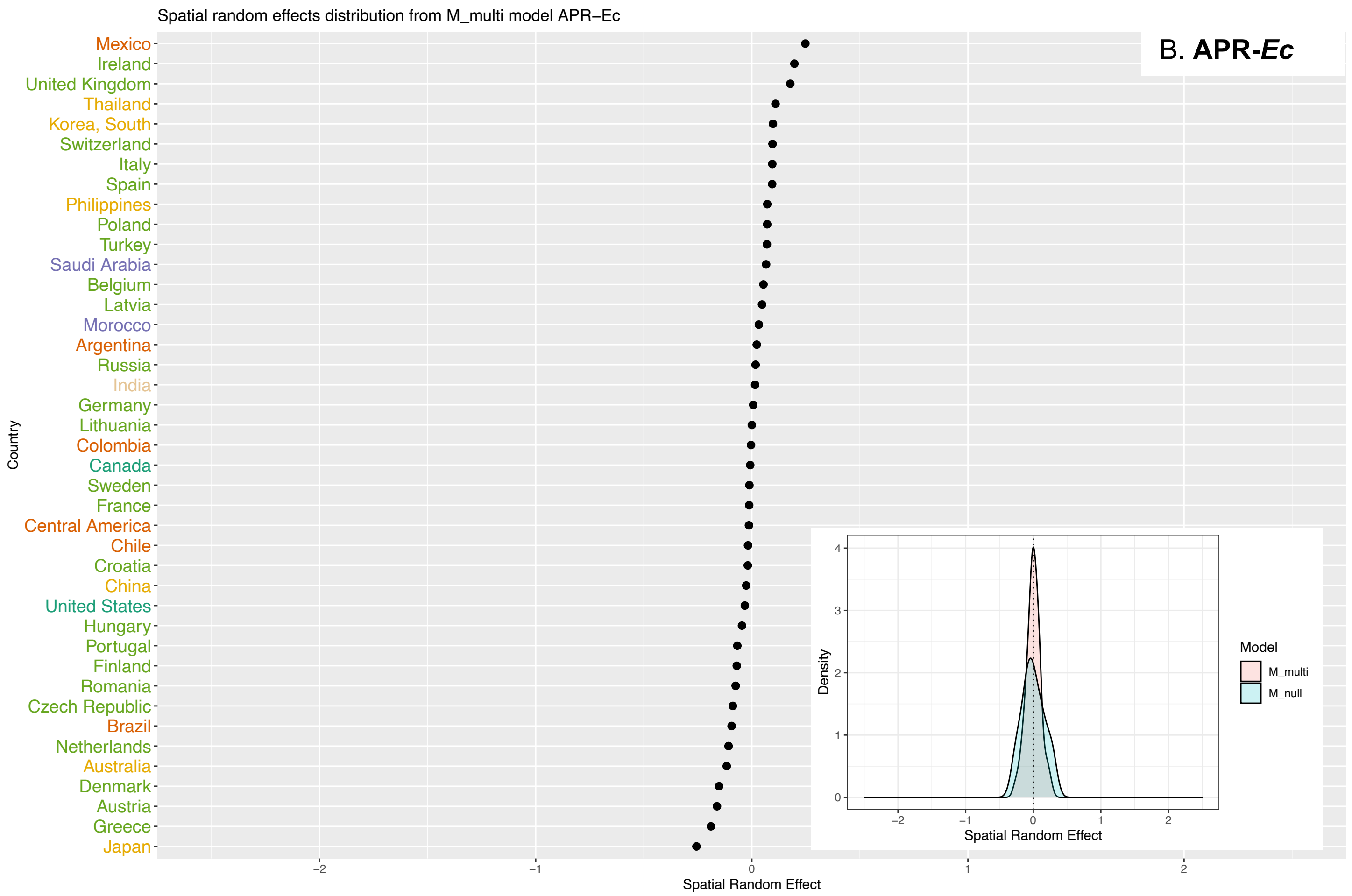
Spatial random effects distribution from M_multi model APR−Ec
B. APR-*Ec*
Mexico
Ireland
United Kingdom
Thailand
Korea, South
Switzerland
Italy
Spain
Philippines
Poland
Turkey
Saudi Arabia
Belgium
Latvia
Morocco
Argentina
Russia
India
Germany
Lithuania
Colombia
Canada
Sweden
France
Central America
Chile
Croatia
China
United States
Hungary
Portugal
Finland
Romania
Czech Republic
Brazil
Netherlands
Australia
Denmark
Austria
Greece
Japan
Country
Spatial Random Effect
Density
Model
M_multi
M_null

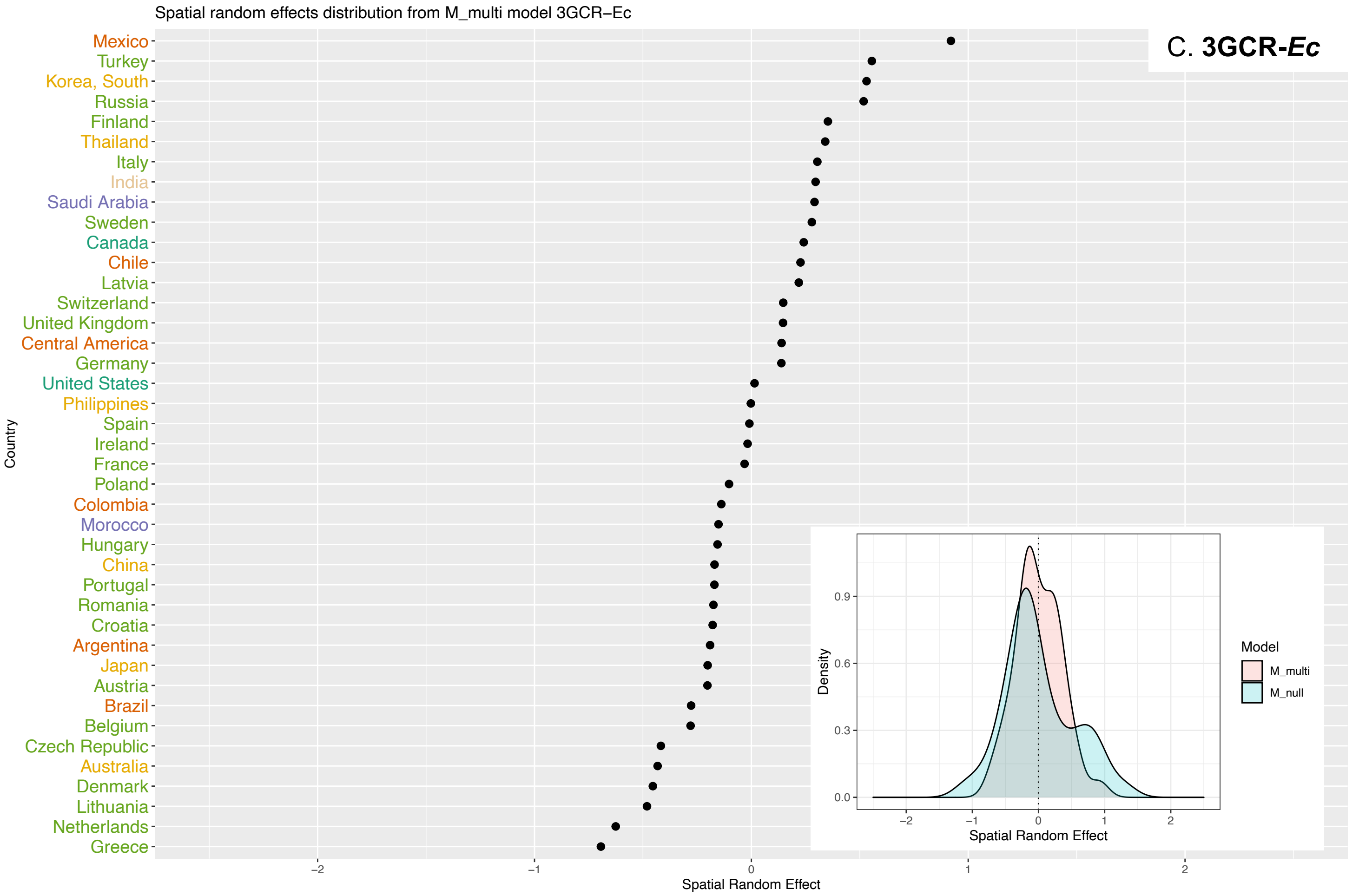
Spatial random effects distribution from M_multi model 3GCR−Ec
C. 3GCR-Ec
Mexico
Turkey
Korea, South
Russia
Finland
Thailand
Italy
India
Saudi Arabia
Sweden
Canada
Chile
Latvia
Switzerland
United Kingdom
Central America
Germany
United States
Philippines
Spain
Ireland
France
Poland
Colombia
Morocco
Hungary
China
Portugal
Romania
Croatia
Argentina
Japan
Austria
Brazil
Belgium
Czech Republic
Australia
Denmark
Lithuania
Netherlands
Greece
Country
Spatial Random Effect
Density
Model
M_multi
M_null

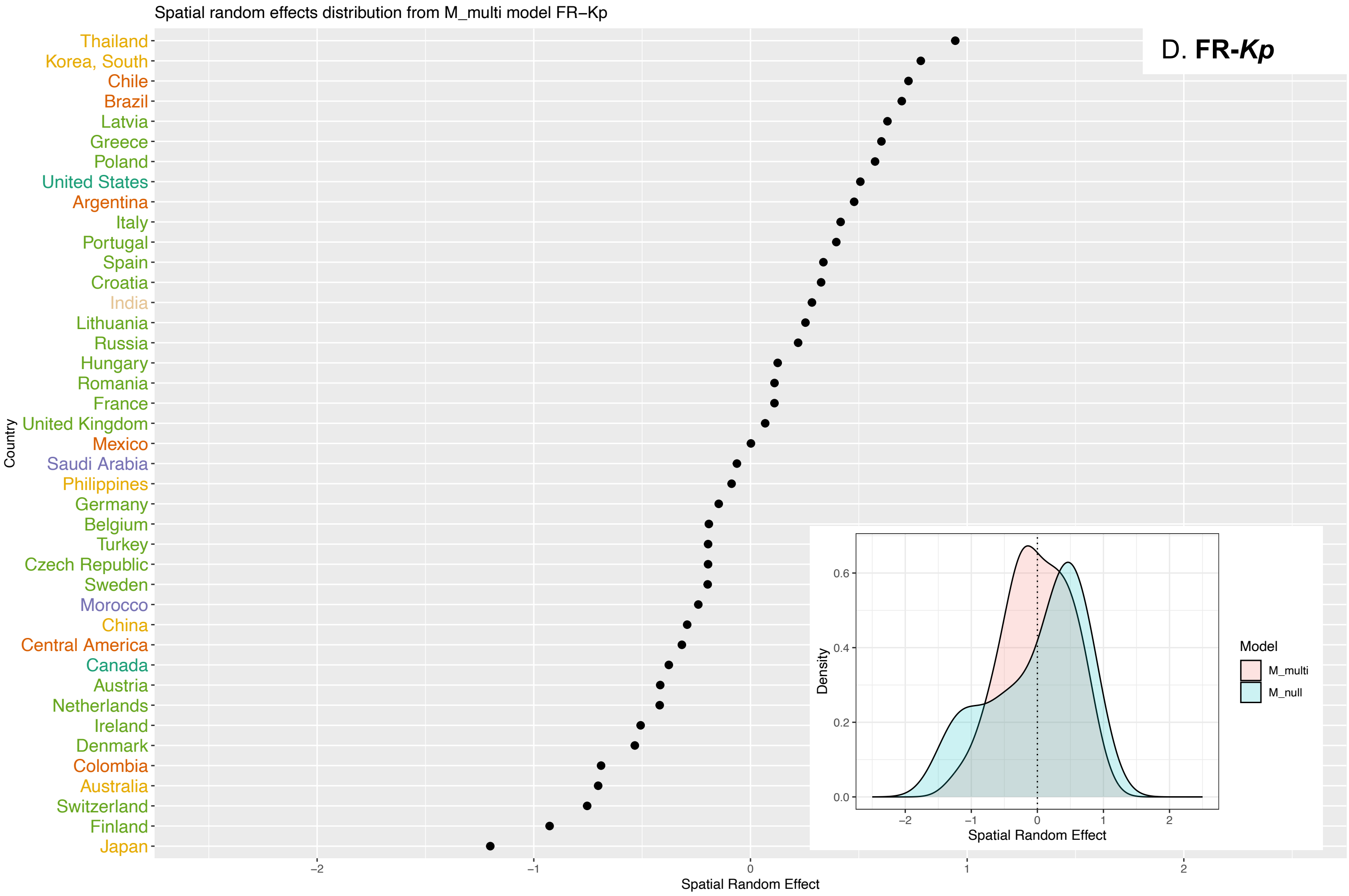
Spatial random effects distribution from M_multi model FR−Kp
D. FR-Kp
Thailand
Korea, South
Chile
Brazil
Latvia
Greece
Poland
United States
Argentina
Italy
Portugal
Spain
Croatia
India
Lithuania
Russia
Hungary
Romania
France
United Kingdom
Mexico
Saudi Arabia
Philippines
Germany
Belgium
Turkey
Czech Republic
Sweden
Morocco
China
Central America
Canada
Austria
Netherlands
Ireland
Denmark
Colombia
Australia
Switzerland
Finland
Japan
Country
Spatial Random Effect
Density
Model
M_multi
M_null

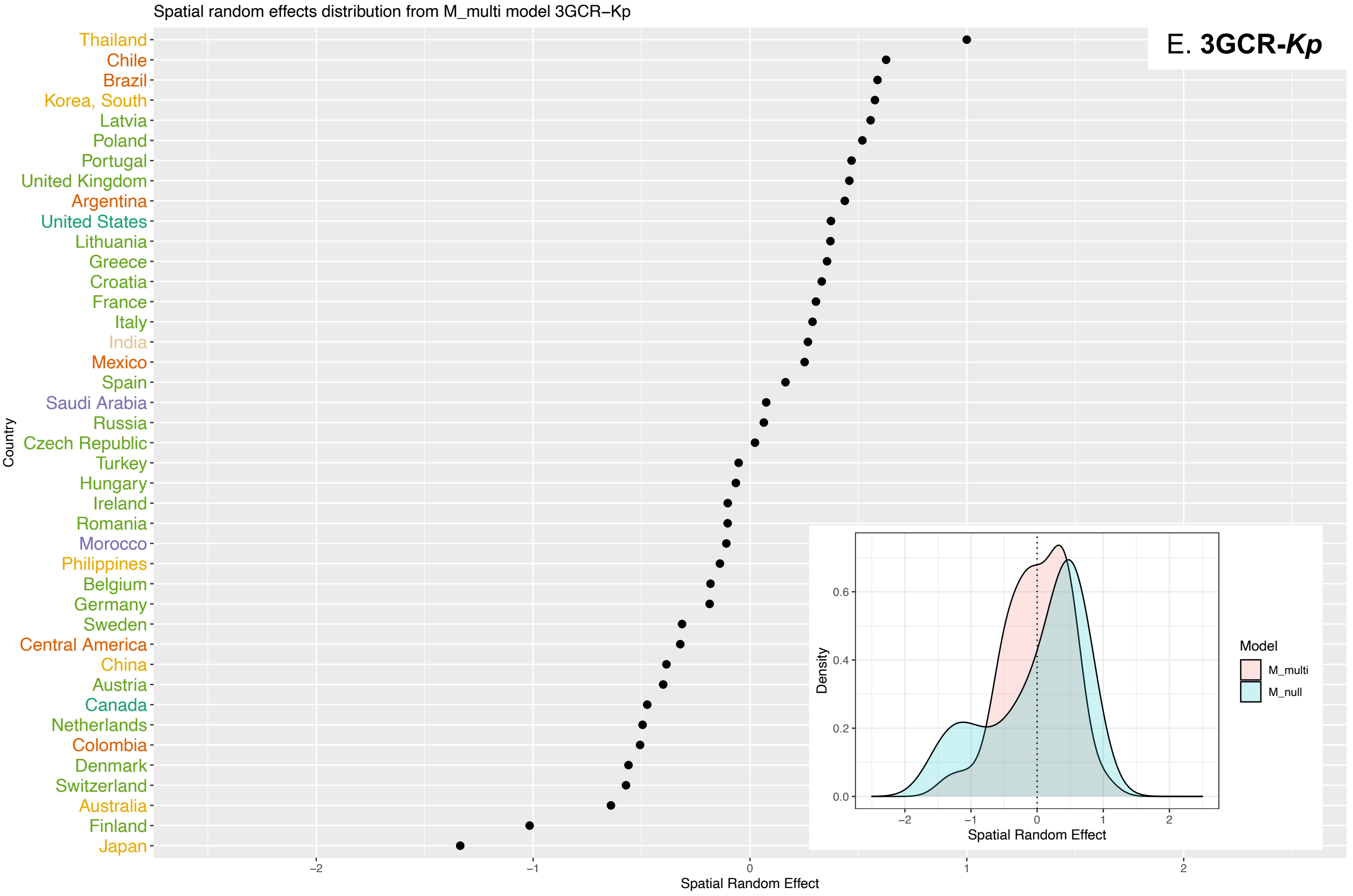

Spatial random effects distribution from M_multi model 3GCR−Kp
E. 3GCR-Kp
Thailand
Chile
Brazil
Korea, South
Latvia
Poland
Portugal
United Kingdom
Argentina
United States
Lithuania
Greece
Croatia
France
Italy
India
Mexico
Spain
Saudi Arabia
Russia
Czech Republic
Turkey
Hungary
Ireland
Romania
Morocco
Philippines
Belgium
Germany
Sweden
Central America
China
Austria
Canada
Netherlands
Colombia
Denmark
Switzerland
Australia
Finland
Japan
Country
Spatial Random Effect
Density
Model
M_multi
M_null

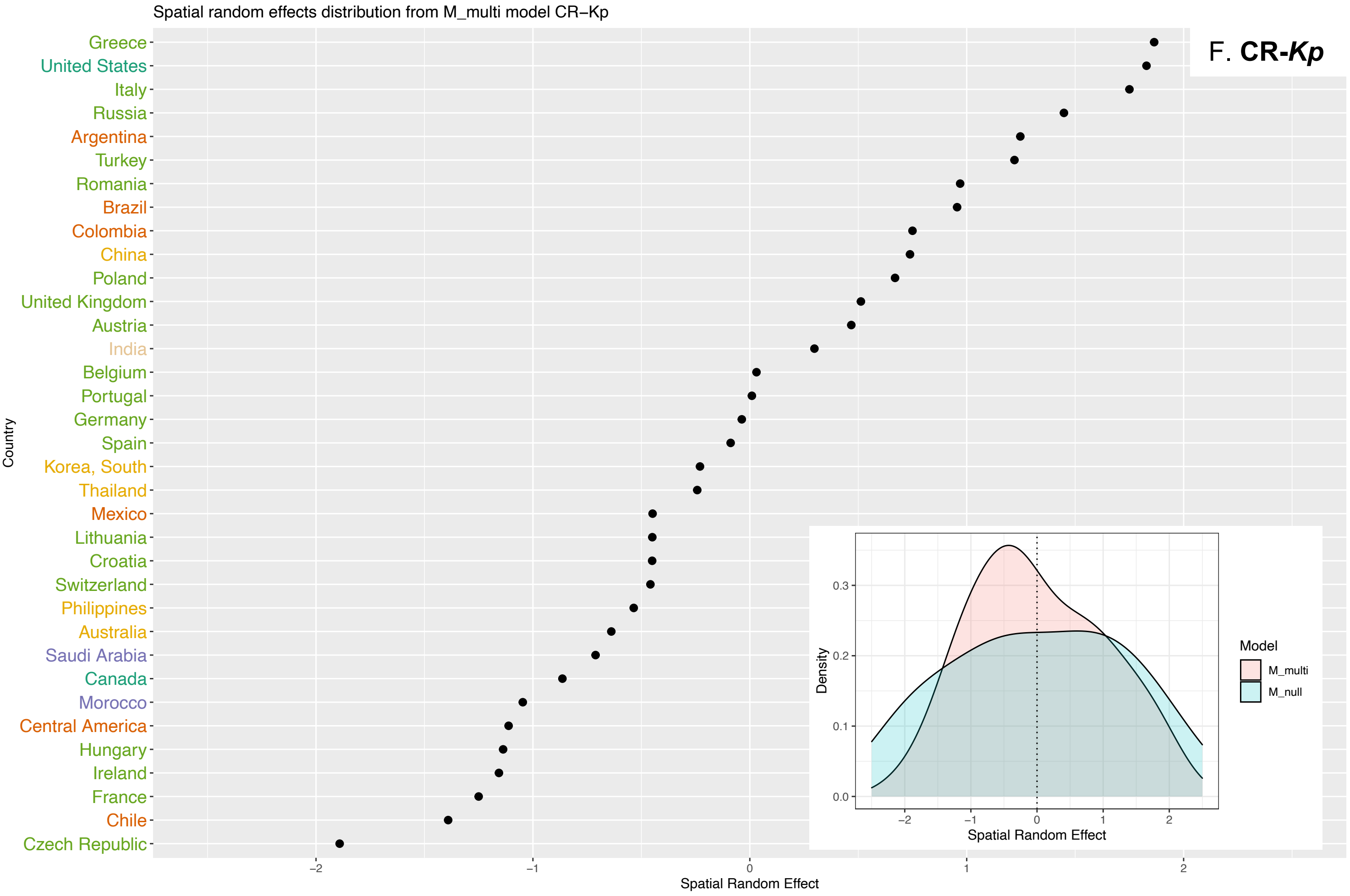

Spatial random effects distribution from M_multi model CR−Kp
F. CR-Kp
Greece
United States
Italy
Russia
Argentina
Turkey
Romania
Brazil
Colombia
China
Poland
United Kingdom
Austria
India
Belgium
Portugal
Germany
Spain
Korea, South
Thailand
Mexico
Lithuania
Croatia
Switzerland
Philippines
Australia
Saudi Arabia
Canada
Morocco
Central America
Hungary
Ireland
France
Chile
Czech Republic
Country
Spatial Random Effect
Density
Model
M_multi
M_null

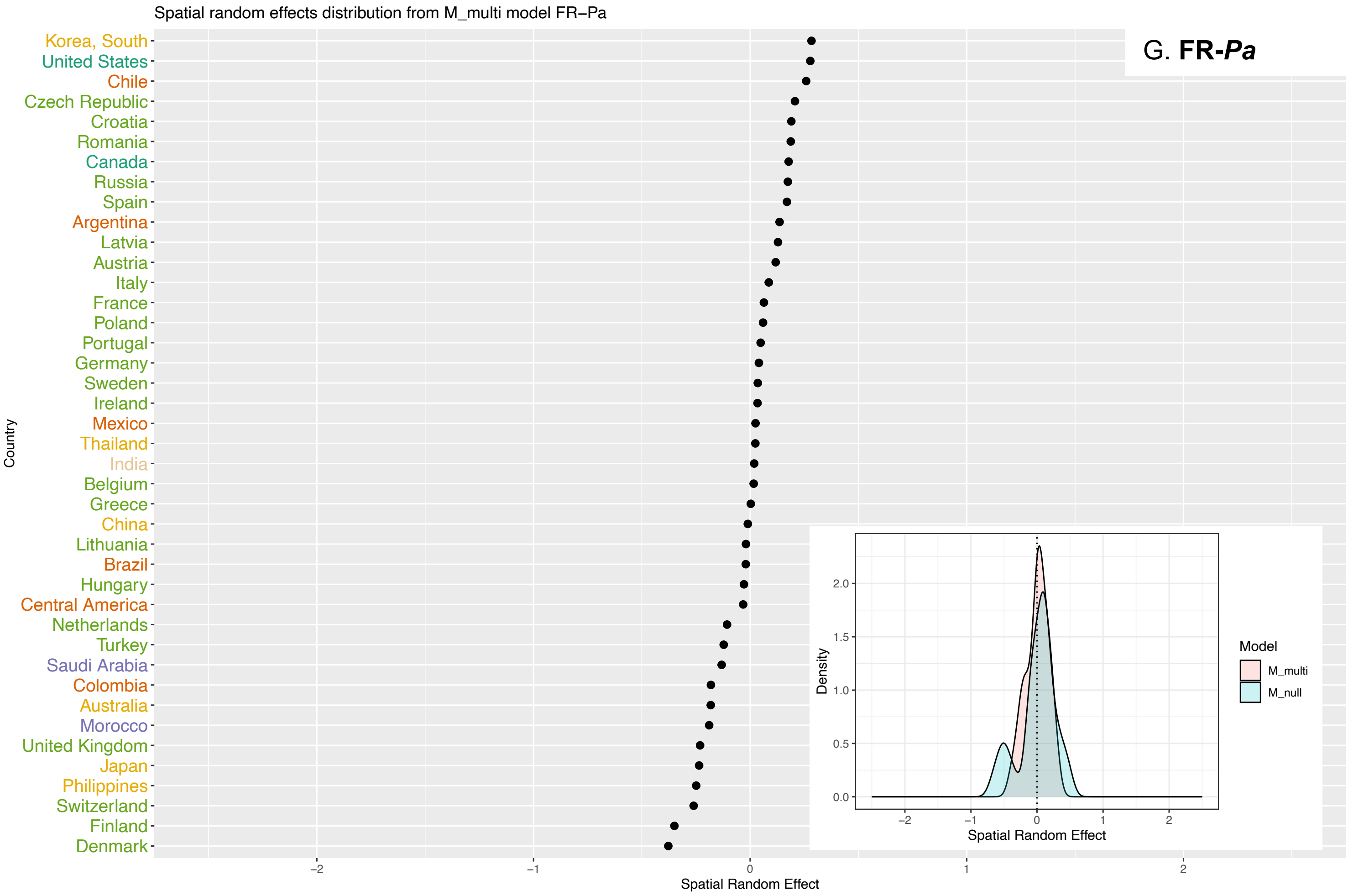
Spatial random effects distribution from M_multi model FR–Pa
G. FR-Pa
Korea, South
United States
Chile
Czech Republic
Croatia
Romania
Canada
Russia
Spain
Argentina
Latvia
Austria
Italy
France
Poland
Portugal
Germany
Sweden
Ireland
Mexico
Thailand
India
Belgium
Greece
China
Lithuania
Brazil
Hungary
Central America
Netherlands
Turkey
Saudi Arabia
Colombia
Australia
Morocco
United Kingdom
Japan
Philippines
Switzerland
Finland
Denmark
Country
Spatial Random Effect
Density
Model
M_multi
M_null

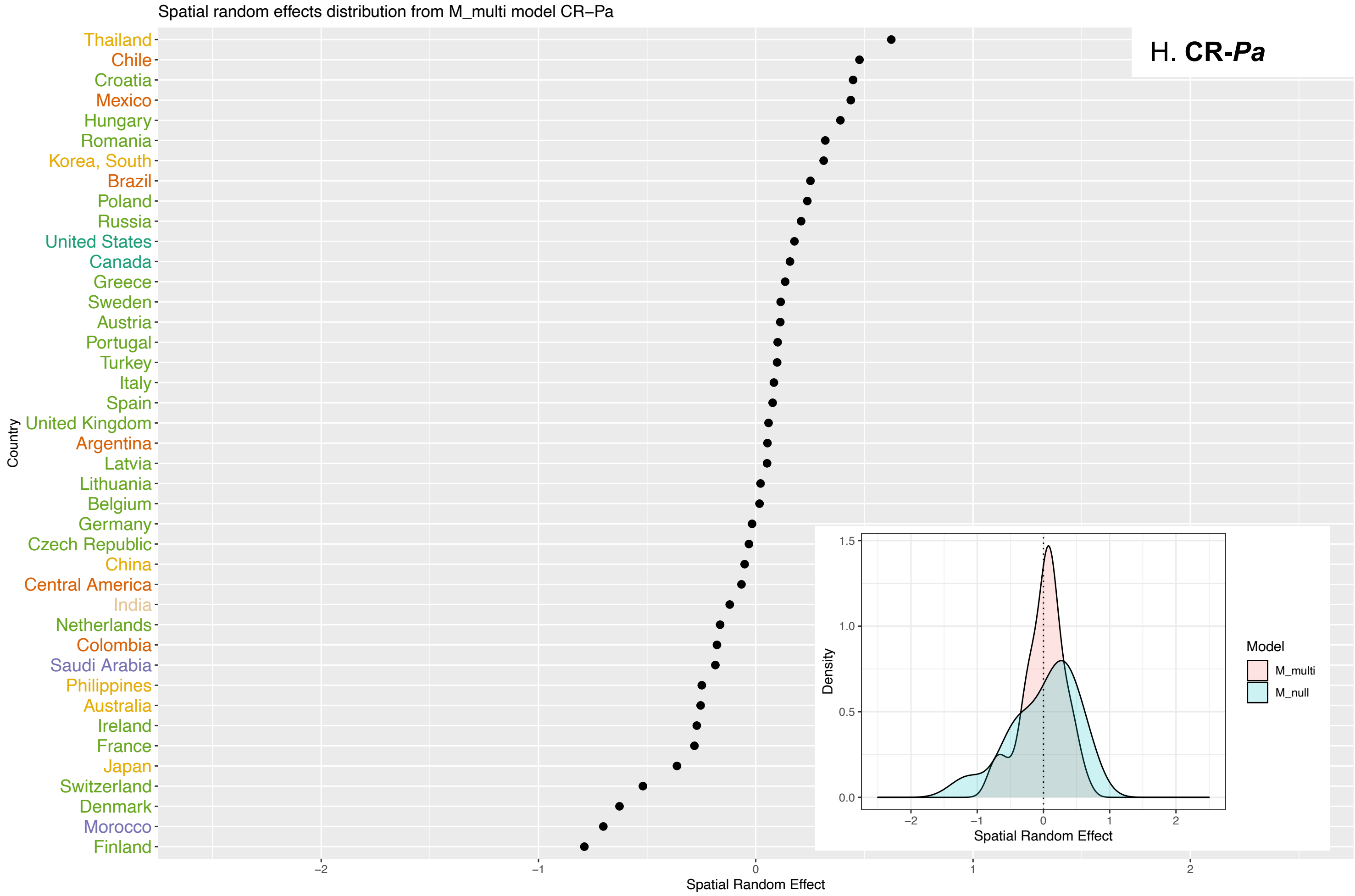
Spatial random effects distribution from M_multi model CR–Pa
H. CR-Pa
Thailand
Chile
Croatia
Mexico
Hungary
Romania
Korea, South
Brazil
Poland
Russia
United States
Canada
Greece
Sweden
Austria
Portugal
Turkey
Italy
Spain
United Kingdom
Argentina
Latvia
Lithuania
Belgium
Germany
Czech Republic
China
Central America
India
Netherlands
Colombia
Saudi Arabia
Philippines
Australia
Ireland
France
Japan
Switzerland
Denmark
Morocco
Finland
Country
Spatial Random Effect
Density
Model
M_multi
M_null

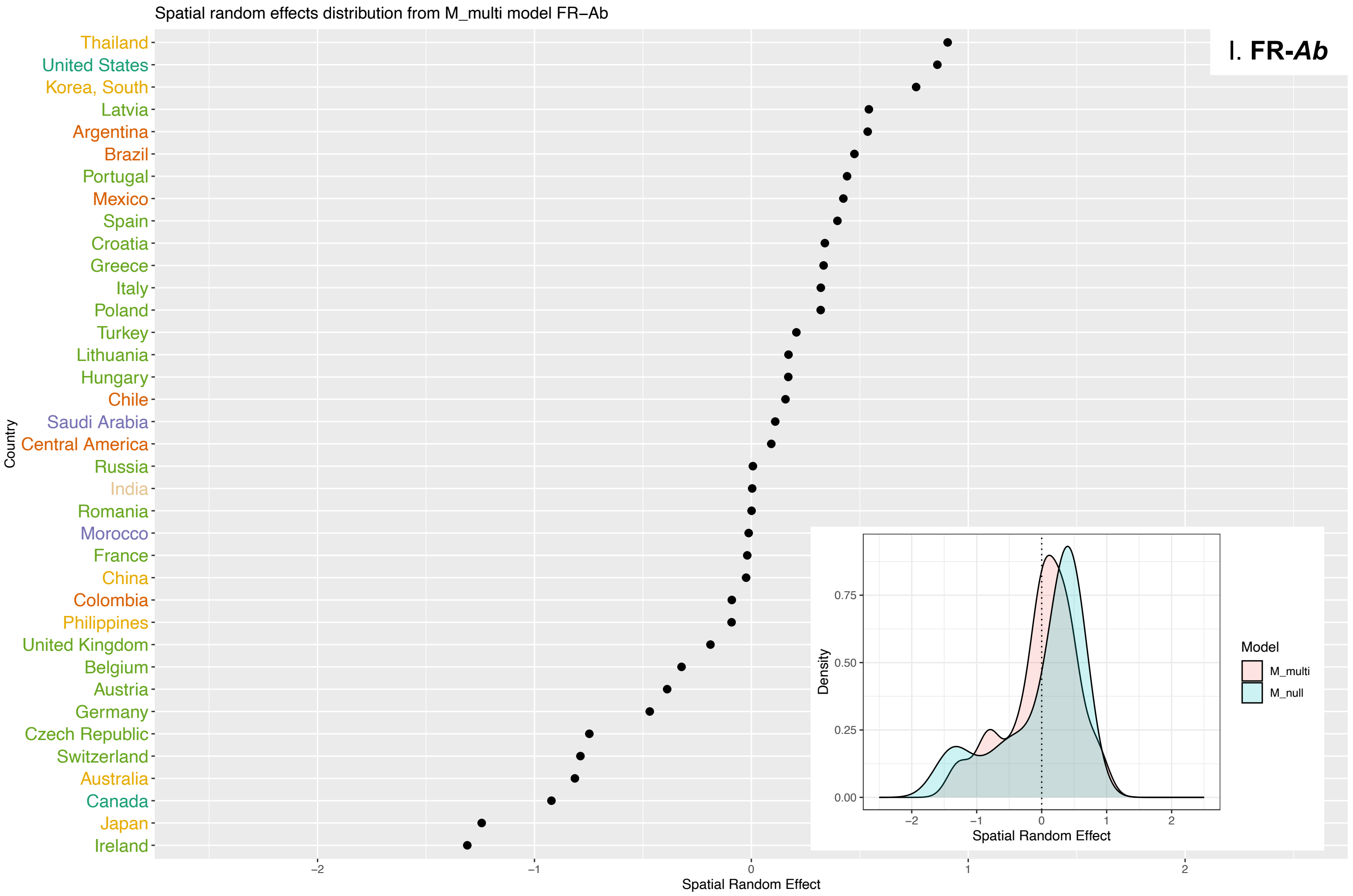
Spatial random effects distribution from M_multi model FR−Ab
I. FR-Ab
Thailand
United States
Korea, South
Latvia
Argentina
Brazil
Portugal
Mexico
Spain
Croatia
Greece
Italy
Poland
Turkey
Lithuania
Hungary
Chile
Saudi Arabia
Central America
Russia
India
Romania
Morocco
France
China
Colombia
Philippines
United Kingdom
Belgium
Austria
Germany
Czech Republic
Switzerland
Australia
Canada
Japan
Ireland
Country
Spatial Random Effect
Density
Model
M_multi
M_null

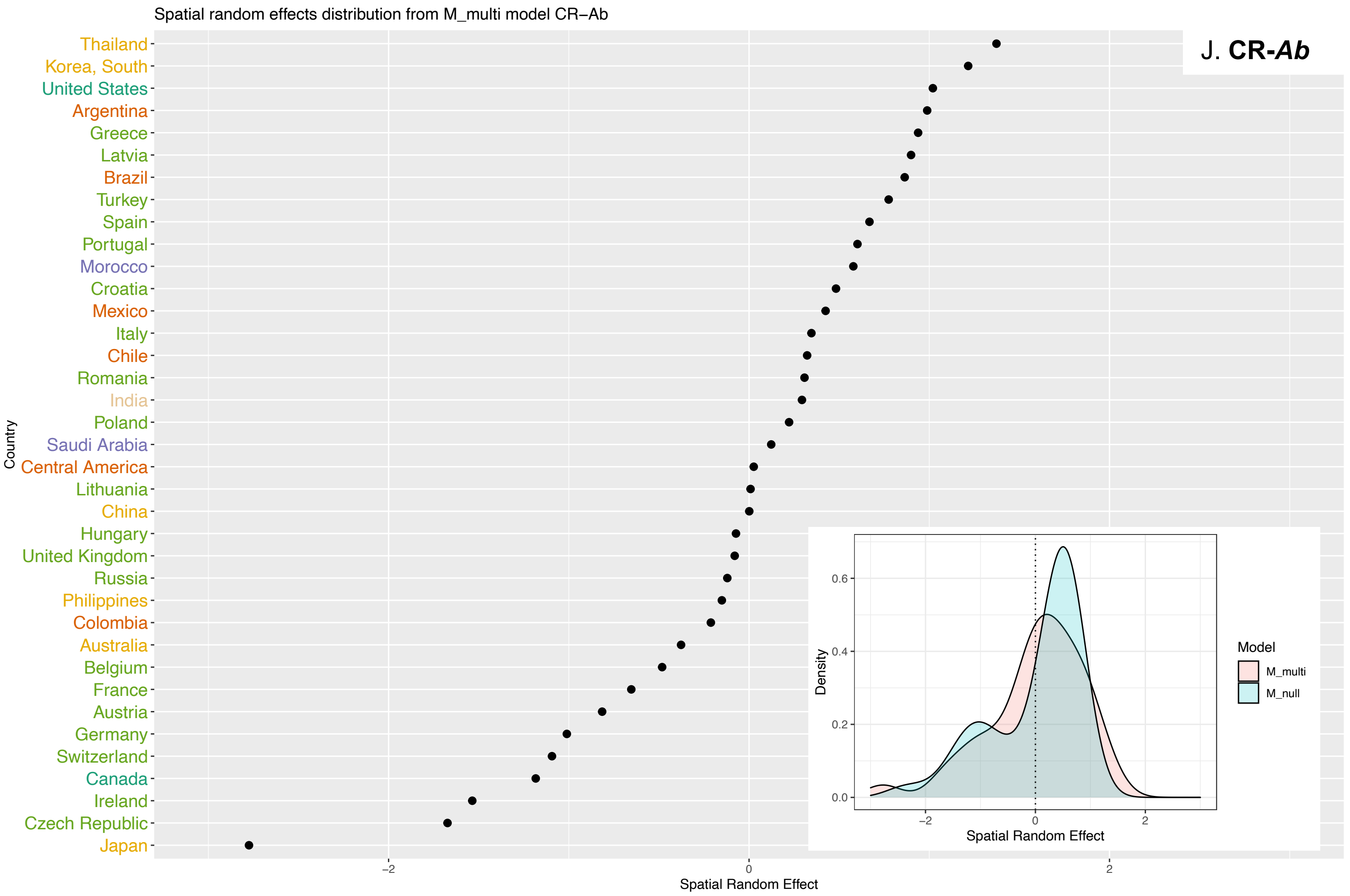
Spatial random effects distribution from M_multi model CR−Ab
J. CR-Ab
Thailand
Korea, South
United States
Argentina
Greece
Latvia
Brazil
Turkey
Spain
Portugal
Morocco
Croatia
Mexico
Italy
Chile
Romania
India
Poland
Saudi Arabia
Central America
Lithuania
China
Hungary
United Kingdom
Russia
Philippines
Colombia
Australia
Belgium
France
Austria
Germany
Switzerland
Canada
Ireland
Czech Republic
Japan
Country
Spatial Random Effect
Density
Model
M_multi
M_null

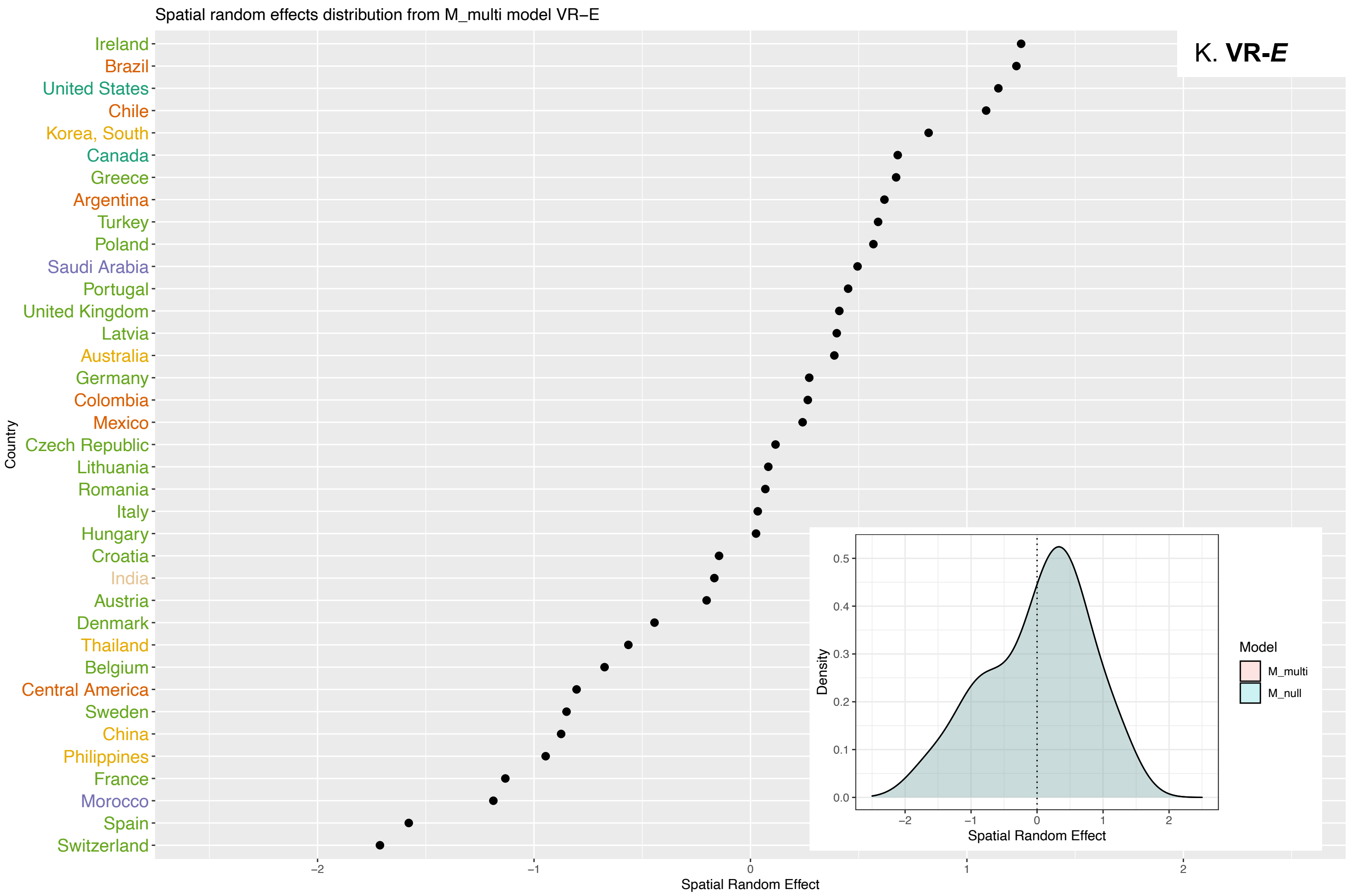
Spatial random effects distribution from M_multi model VR−E
K. **VR-*E***
Ireland
Brazil
United States
Chile
Korea, South
Canada
Greece
Argentina
Turkey
Poland
Saudi Arabia
Portugal
United Kingdom
Latvia
Australia
Germany
Colombia
Mexico
Czech Republic
Lithuania
Romania
Italy
Hungary
Croatia
India
Austria
Denmark
Thailand
Belgium
Central America
Sweden
China
Philippines
France
Morocco
Spain
Switzerland
Country
Spatial Random Effect
Density
Model
M_multi
M_null

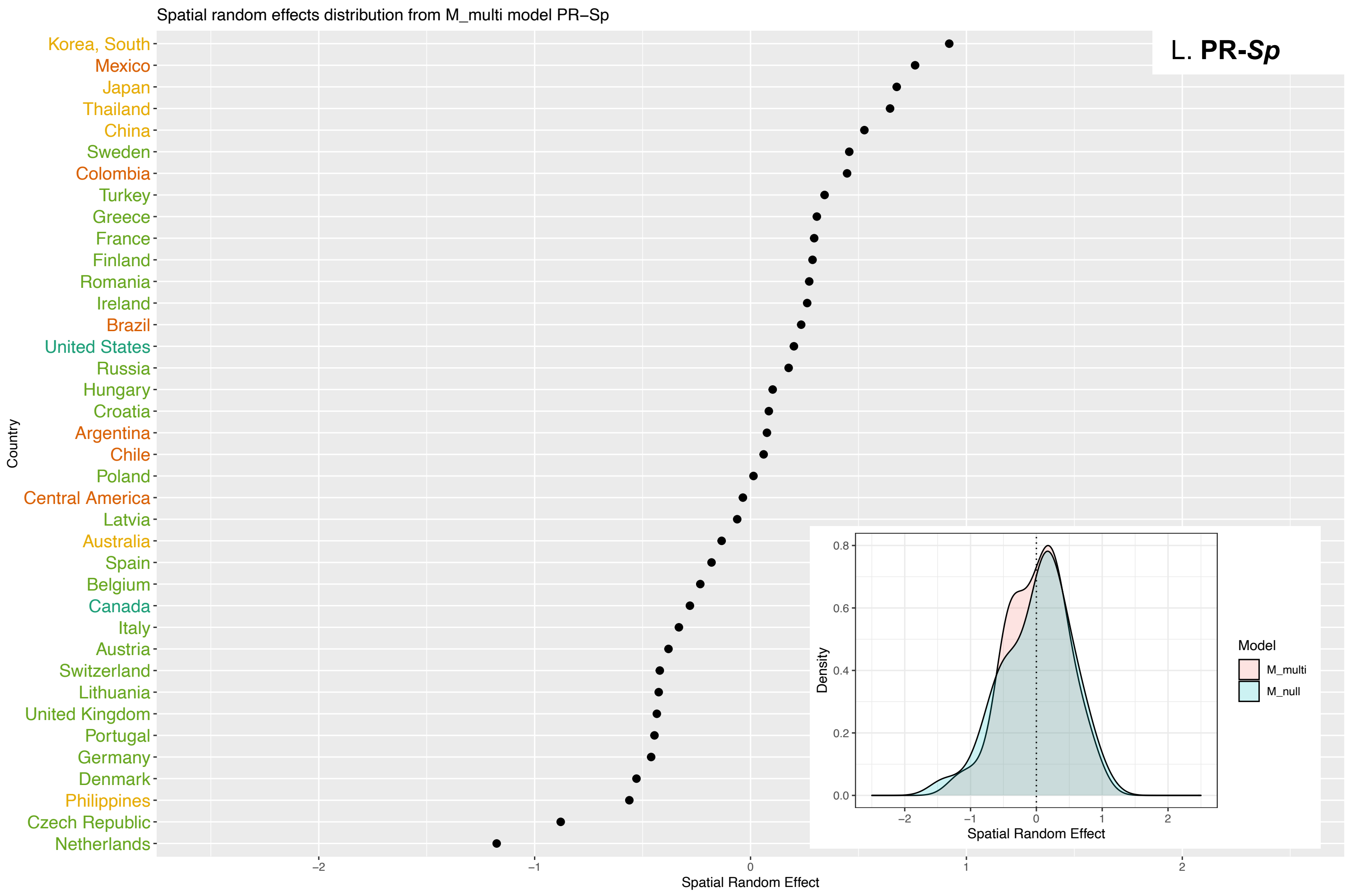
Spatial random effects distribution from M_multi model PR−Sp
L. **PR-*Sp***
Korea, South
Mexico
Japan
Thailand
China
Sweden
Colombia
Turkey
Greece
France
Finland
Romania
Ireland
Brazil
United States
Russia
Hungary
Croatia
Argentina
Chile
Poland
Central America
Latvia
Australia
Spain
Belgium
Canada
Italy
Austria
Switzerland
Lithuania
United Kingdom
Portugal
Germany
Denmark
Philippines
Czech Republic
Netherlands
Country
Spatial Random Effect
Density
Model
M_multi
M_null

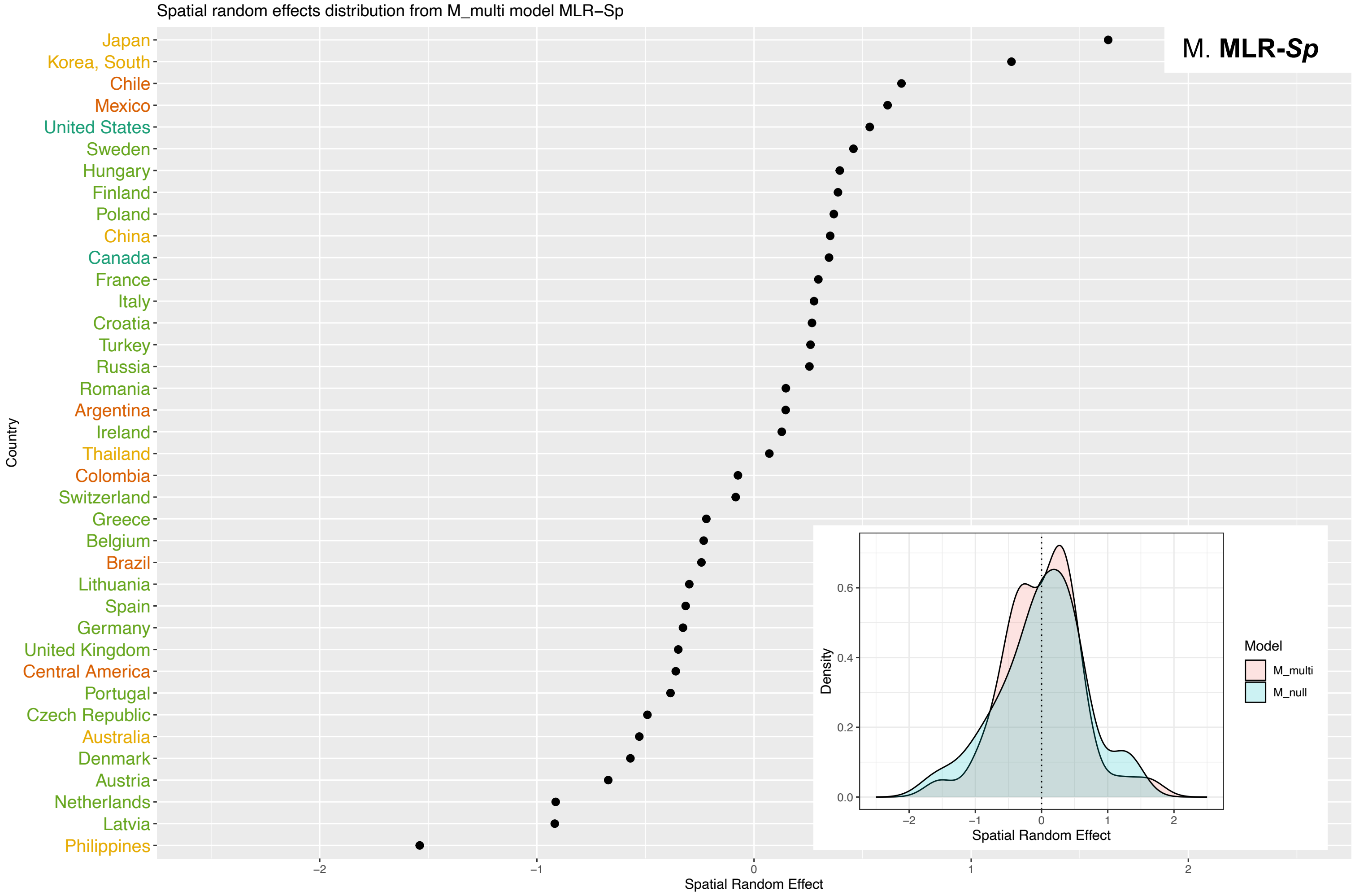
Spatial random effects distribution from M_multi model MLR−Sp
M. MLR-Sp
Japan
Korea, South
Chile
Mexico
United States
Sweden
Hungary
Finland
Poland
China
Canada
France
Italy
Croatia
Turkey
Russia
Romania
Argentina
Ireland
Thailand
Colombia
Switzerland
Greece
Belgium
Brazil
Lithuania
Spain
Germany
United Kingdom
Central America
Portugal
Czech Republic
Australia
Denmark
Austria
Netherlands
Latvia
Philippines
Country
Spatial Random Effect
Density
Model
M_multi
M_null
Spatial Random Effect

#### Supplementary analyses

***Supplementary Figure S12.* Comparison of ATLAS data and EARS-Net data (ECDC) by drug-bug pair for European countries.**

Comparisons for:

1. **FR-Ec**: fluoroquinolone-resistant *E. coli*
2. **APR-Ec**: aminopenicillin-resistant *E. coli*
3. **3GCR-Ec**: third generation cephalosporin-resistant *E. coli*
4. **FR-Kp**: fluoroquinolone-resistant *K. pneumoniae*
5. **3GCR-Kp**: third generation cephalosporin-resistant *K. pneumoniae*
6. **CR-Kp**: carbapenem-resistant *K. pneumoniae*
7. **FR-Pa**: fluoroquinolone-resistant *P. aeruginosa*
8. **CR-Pa**: carbapenem-resistant *P. aeruginosa*
9. **FR-Ab**: fluoroquinolone-resistant *A. baumannii*
10. **CR-Ab**: carbapenem-resistant *A. baumannii*
11. **VR-E**: vancomycin-resistant Enterococci
12. **PR-Sp**: penicillin-non-susceptible *S. pneumoniae*
13. **MLR-Sp**: macrolide-resistant *S. pneumoniae*
14. **MRSA**: Methicillin-resistant *Staphylococcus aureus*

We compared ATLAS data to data from another surveillance system, here the EARS-Net system (ECDC, data available on their website).

*Methods for visual comparison by country*

We plotted 2006-2019 trends for ABR rates collected by ATLAS and by EARS-Net for European countries. For ATLAS, we plotted ABR rates from blood isolates (to compare to ECDC data which collects data from blood samples) and from all sources isolates (data used in the main analyses).

*Methods for statistical comparison by country*

We compared average ABR rates in blood samples for 2006-2019 between ECDC and ATLAS using a two-proportion z-test. We compared weighted linear regression slopes between ECDC and ATLAS using an interaction term between year and the source of data (ATLAS or ECDC). We used ATLAS data from blood isolates to be comparable to ECDC data (the describe ABR rates in blood samples). Summary statistics are reported in the tables below the graphs, with average proportions by data source, regression slopes by data source, and p-values associated with statistical tests. Highlighted in bold are average ABR rates that are not significantly different between ATLAS and ECDC; and regression slopes that are not significantly different between ATLAS and ECDC (using a threshold of 0.05 for p-value). P-value indicated as “0” are < 1e-05.

*Description of results*

Visually, methicillin-resistant *Staphylococcus aureus* (MRSA) trends in ATLAS present an unusual pattern with a sharp decrease between year 2017 and 2018, not in concordance with ECDC data. For this reason, MRSA was excluded from further analyses. For all other pairs, uncertainty is greater for ATLAS data, as 95% CI around proportions are wider than those observed for ECDC data. This is due to the smaller sample size by drug-bug-country-year combination. Uncertainty is also greater for ATLAS data from blood isolates compared to all sources isolates, as sample size is further decreased.

Statistically, we observed strong concordance for trends across 2006-2019 between ATLAS and ECDC. By opposition, average proportions were less similar, with more often a higher estimation of ABR rates by ATLAS.

For FR-*Pa*, average ABR rates were significantly different between ATLAS and ECDC, with only 22% of countries not having statistically significant different average proportions over 2006-2019. ABR rates estimated by ATLAS were systematically higher than rates estimated by ECDC. However, trends were similar between ATLAS and ECDC, with 78% of countries not having significantly different regression slopes.

For the rest of the drug-bug pairs, more than 40% of countries showed statistically significant similar average ABR rates and more than 70% of countries showed statistically significant similar trends.

These supplementary analyses suggest relatively good adequacy for the drug-bug pairs chosen between ECDC and ATLAS data, especially for ABR rates trends.

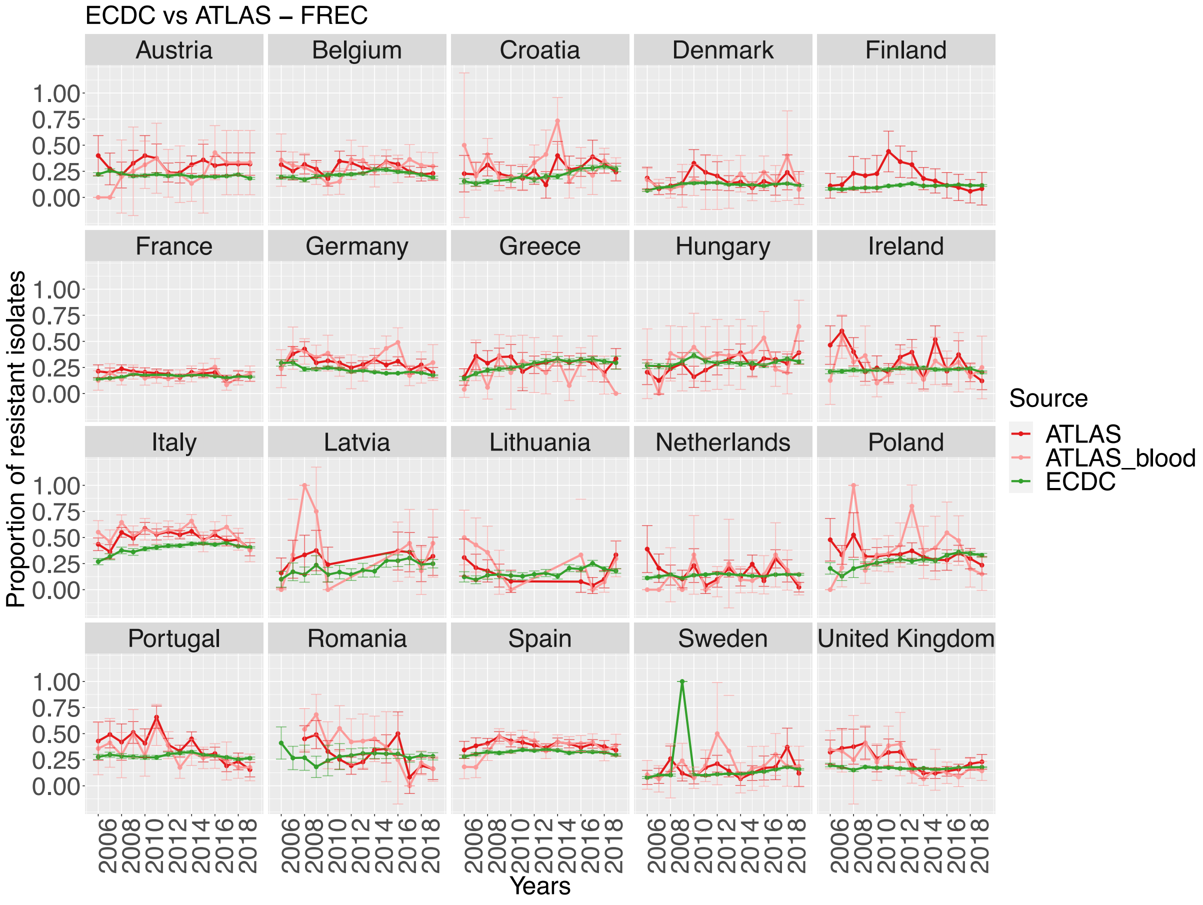

| "Country" | "ABR_prop_ATLAS" | "ABR_prop_ECDC" | "z_test" | "slope_ATLAS" | "slope_ECDC" | "anova_interaction" |
|---|---|---|---|---|---|---|
| "Austria" | **0.266** | **0.209** | **0.14558** | **0.00792** | **-0.00273** | **0.61674** |
| "Belgium" | 0.285 | 0.223 | 0.00039 | **0.00741** | **0.00205** | **0.64772** |
| "Croatia" | 0.286 | 0.218 | 8e-04 | **0.00507** | **0.01262** | **0.35713** |
| "Denmark" | **0.144** | **0.121** | **0.24612** | **0.00308** | **0.0021** | **0.93523** |
| "France" | **0.172** | **0.166** | **0.58823** | **-0.00023** | **0.00018** | **0.95248** |
| "Germany" | 0.325 | 0.202 | 0 | **-0.00626** | **-0.00593** | **0.97544** |
| "Greece" | **0.224** | **0.269** | **0.13175** | **0.01604** | **0.01351** | **0.78656** |
| "Hungary" | 0.366 | 0.298 | 0.07337 | **6e-05** | **0.002** | **0.91454** |
| "Ireland" | **0.246** | **0.229** | **0.47409** | **-0.00744** | **0.00049** | **0.39096** |
| "Italy" | 0.558 | 0.417 | 0 | **-0.00553** | **0.00376** | **0.38856** |
| "Latvia" | 0.385 | 0.223 | 0.00981 | **-0.02472** | **0.01119** | **0.13384** |
| "Lithuania" | 0.25 | 0.176 | 0.04192 | -0.02024 | 0.00762 | 0.03317 |
| "Netherlands" | **0.126** | **0.14** | **0.72244** | **-0.00458** | **0.00134** | **0.73755** |
| "Poland" | **0.337** | **0.308** | **0.35751** | **0.00167** | **0.0123** | **0.43934** |
| "Portugal" | **0.295** | **0.284** | **0.63712** | **-0.01931** | **-0.00228** | **0.17565** |
| "Romania" | 0.38 | 0.291 | 0.00463 | -0.03939 | -0.00033 | 1e-05 |
| "Spain" | 0.394 | 0.323 | 0 | **0.00712** | **-0.00011** | **0.3778** |
| "Sweden" | 0.174 | 0.127 | 0.04886 | **0.0089** | **0.00604** | **0.79684** |
| "United Kingdom" | **0.196** | **0.173** | **0.21573** | **-0.01731** | **0.00055** | **0.06593** |

Percentage of countries with no significantly different ABR proportions over 2006-2019: 9/19 = **47%**
Percentage of countries with no significantly different ABR trends (2006-2019): 17/19 = **89%**

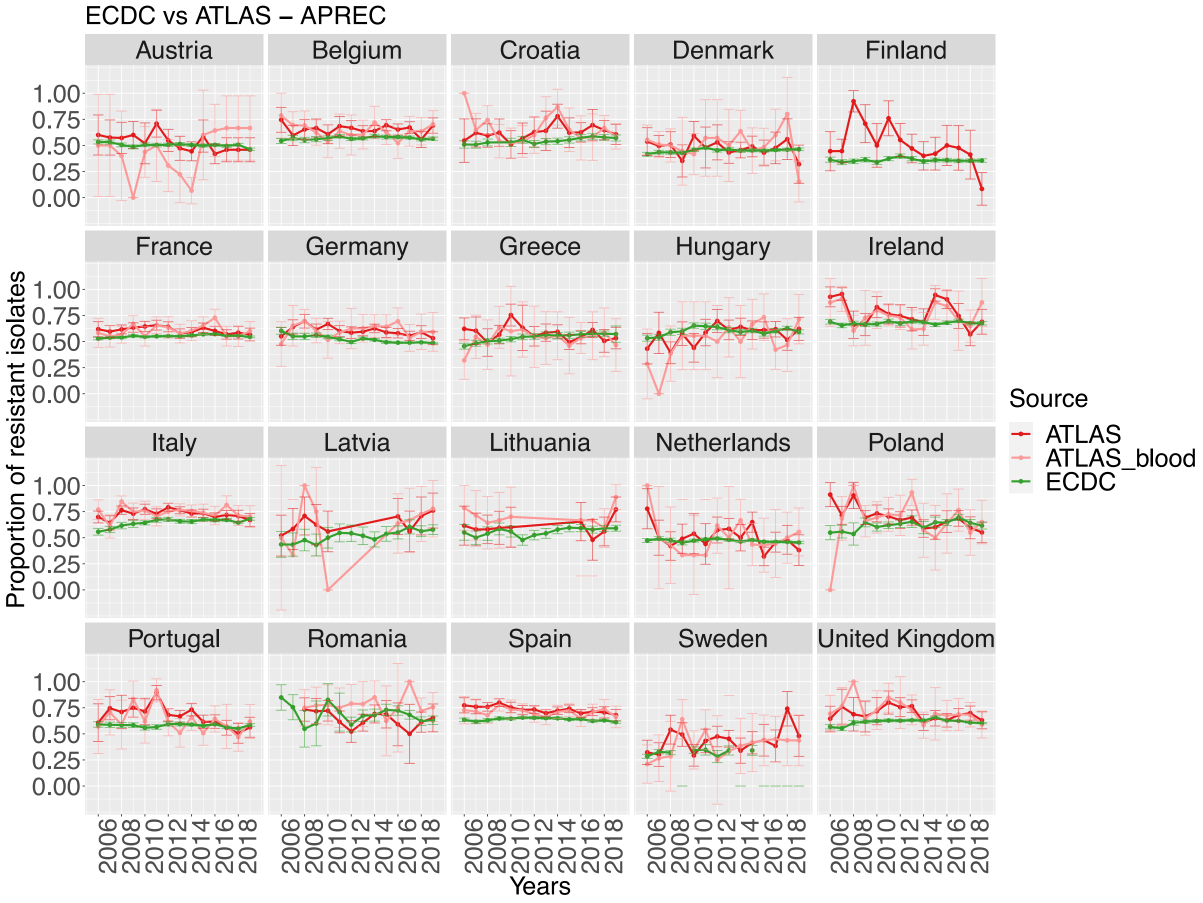

| "Country" | "ABR_prop_ATLAS" | "ABR_prop_ECDC" | "z_test" | "slope_ATLAS" | "slope_ECDC" | "anova_interaction" |
|---|---|---|---|---|---|---|
| "Austria" | **0.435** | **0.501** | **0.16953** | **0.01999** | **-0.00274** | **0.33045** |
| "Belgium" | 0.637 | 0.57 | 0.00129 | **-0.00275** | **0.00059** | **0.55309** |
| "Croatia" | 0.622 | 0.548 | 0.00248 | **4e-05** | **0.00571** | **0.35476** |
| "Denmark" | 0.505 | 0.452 | 0.07003 | **-0.00797** | **0.00226** | **0.25865** |
| "France" | 0.609 | 0.553 | 1e-05 | **0.00181** | **0.0015** | **0.95452** |
| "Germany" | 0.615 | 0.499 | 0 | **-0.00033** | **-0.00591** | **0.48291** |
| "Greece" | **0.524** | **0.536** | **0.75256** | **0.00647** | **0.00896** | **0.68923** |
| "Hungary" | **0.534** | **0.601** | **0.10052** | **0.01525** | **0.00195** | **0.50768** |
| "Ireland" | **0.726** | **0.679** | **0.05135** | **-0.00517** | **0.00068** | **0.47652** |
| "Italy" | 0.751 | 0.656 | 0 | **-0.00391** | **0.00423** | **0.17627** |
| "Latvia" | **0.673** | **0.537** | **0.07139** | **0.01806** | **0.01047** | **0.51089** |
| "Lithuania" | 0.718 | 0.569 | 0.00127 | **0.00822** | **0.00594** | **0.77822** |
| "Netherlands" | **0.51** | **0.47** | **0.37336** | **0.00541** | **-0.00209** | **0.63064** |
| "Poland" | **0.679** | **0.632** | **0.15212** | **-0.00361** | **0.00506** | **0.47005** |
| "Portugal" | **0.604** | **0.578** | **0.2601** | **-0.01991** | **-0.00119** | **0.0973** |
| "Romania" | 0.759 | 0.667 | 0.00447 | **-0.00301** | **-0.00779** | **0.63357** |
| "Spain" | 0.707 | 0.638 | 0 | **-0.00385** | **-0.00138** | **0.62937** |
| "Sweden" | 0.417 | 0.318 | 0.00252 | **0.01028** | **0.00552** | **0.62419** |
| "United Kingdom" | 0.68 | 0.617 | 0.00738 | **-0.00873** | **-0.00027** | **0.57509** |

Percentage of countries with no significantly different ABR proportions over 2006-2019: 8/19 = **42%**
Percentage of countries with no significantly different ABR trends (2006-2019): 19/19 = **100%**

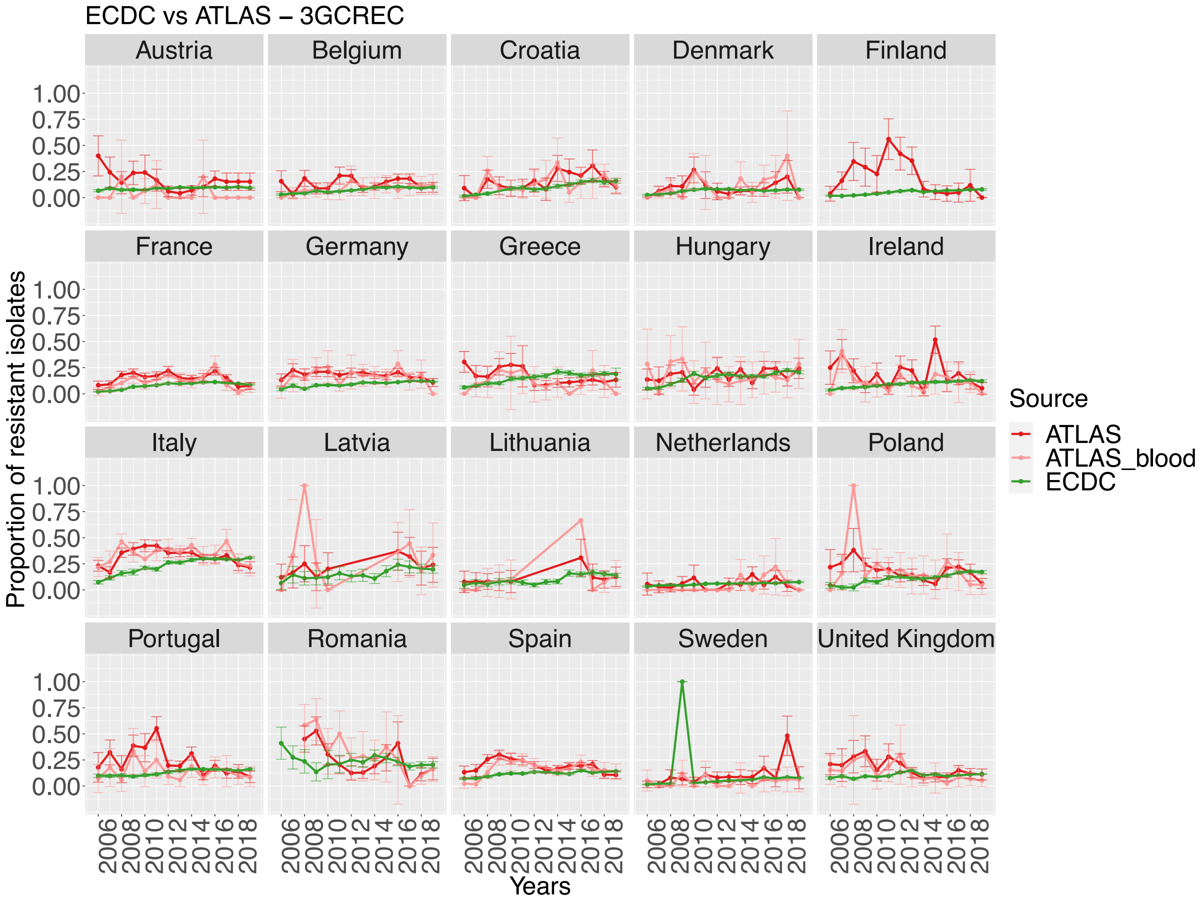

| "Country" | "ABR_prop_ATLAS" | "ABR_prop_ECDC" | "z_test" | "slope_ATLAS" | "slope_ECDC" | "anova_interaction" |
|---|---|---|---|---|---|---|
| "Austria" | 0.032 | 0.091 | 0.03437 | **0.01326** | **0.00217** | **0.81867** |
| "Belgium" | **0.09** | **0.08** | **0.43216** | **0.00623** | **0.00567** | **0.89276** |
| "Croatia" | **0.133** | **0.103** | **0.05479** | **0.00304** | **0.01192** | **0.11949** |
| "Denmark" | **0.092** | **0.068** | **0.12836** | **0.00921** | **0.00327** | **0.64897** |
| "France" | 0.122 | 0.084 | 0 | **-0.00447** | **0.00685** | **0.12755** |
| "Germany" | 0.164 | 0.11 | 1e-05 | **0.00509** | **0.00502** | **0.99342** |
| "Greece" | **0.102** | **0.148** | **0.05363** | **-0.00299** | **0.0118** | **0.13754** |
| "Hungary" | **0.186** | **0.166** | **0.56143** | **-0.00525** | **0.01236** | **0.39669** |
| "Ireland" | **0.115** | **0.097** | **0.28017** | **-0.00177** | **0.00694** | **0.26653** |
| "Italy" | 0.358 | 0.281 | 0 | **-0.00121** | **0.01382** | **0.16657** |
| "Latvia" | 0.327 | 0.174 | 0.00728 | **-0.00169** | **0.00965** | **0.56198** |
| "Lithuania" | **0.089** | **0.118** | **0.39479** | **0.00302** | **0.00919** | **0.61732** |
| "Netherlands" | **0.056** | **0.061** | **0.94448** | **-0.01875** | **0.00274** | **0.43957** |
| "Poland" | **0.159** | **0.139** | **0.41563** | -0.01113 | 0.01113 | 0.00641 |
| "Portugal" | **0.127** | **0.145** | **0.28857** | **-0.00074** | **0.00573** | **0.40025** |
| "Romania" | 0.308 | 0.225 | 0.00437 | -0.04144 | -0.00541 | 0.00053 |
| "Spain" | 0.182 | 0.123 | 0 | **0.0115** | **0.00469** | **0.21358** |
| "Sweden" | **0.055** | **0.056** | **1** | **-0.00181** | **0.00543** | **0.31501** |
| "United Kingdom" | **0.106** | **0.106** | **1** | **-0.01042** | **0.00161** | **0.32662** |

Percentage of countries with no significantly different ABR proportions over 2006-2019: 12/19 = **63%**
Percentage of countries with no significantly different ABR trends (2006-2019): 17/19 = **89%**

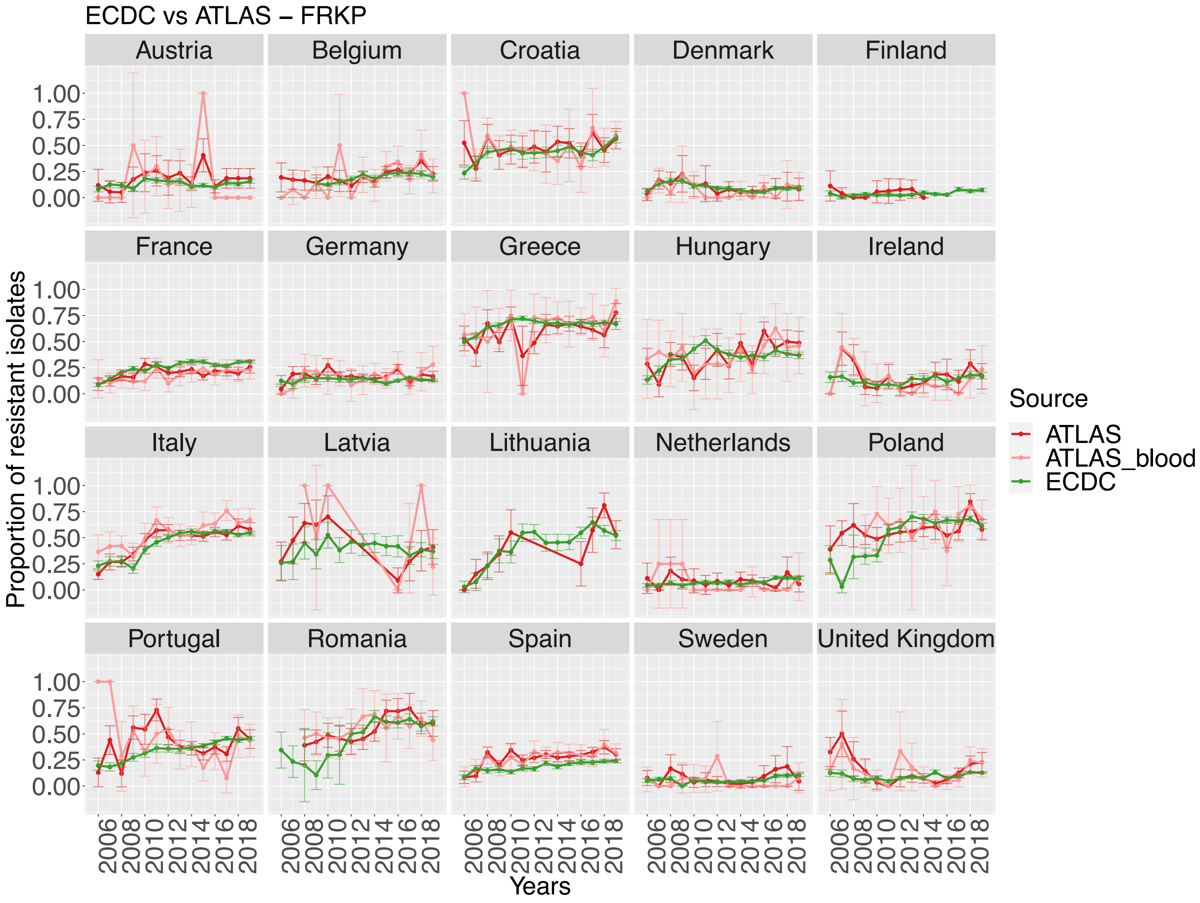

| "Country" | "ABR_prop_ATLAS" | "ABR_prop_ECDC" | "z_test" | "slope_ATLAS" | "slope_ECDC" | "anova_interaction" |
|---|---|---|---|---|---|---|
| "Austria" | **0.155** | **0.132** | **0.74738** | **-0.04353** | **0.00182** | **0.63338** |
| "Belgium" | **0.209** | **0.204** | **0.89705** | **0.01466** | **0.00865** | **0.54037** |
| "Croatia" | **0.469** | **0.439** | **0.30095** | **0.0096** | **0.01382** | **0.68735** |
| "Denmark" | **0.089** | **0.095** | **0.83852** | **-0.00071** | **-0.00322** | **0.86189** |
| "France" | 0.183 | 0.267 | 0 | **0.01054** | **0.01344** | **0.7725** |
| "Germany" | 0.171 | 0.134 | 0.03203 | **0.00276** | **0.00033** | **0.75438** |
| "Greece" | **0.688** | **0.665** | **0.47248** | **0.018** | **0.00633** | **0.53046** |
| "Hungary" | **0.393** | **0.368** | **0.60094** | **0.01325** | **0.01002** | **0.90705** |
| "Ireland" | **0.134** | **0.135** | **1** | **-0.00732** | **0.005** | **0.37986** |
| "Italy" | **0.551** | **0.522** | **0.08835** | **0.02676** | **0.01841** | **0.56418** |
| "Latvia" | **0.391** | **0.399** | **1** | **-0.02958** | **-0.0018** | **0.52998** |
| "Netherlands" | **0.078** | **0.08** | **1** | **-0.0132** | **0.00505** | **0.50813** |
| "Poland" | **0.648** | **0.628** | **0.67997** | **0.01406** | **0.03016** | **0.68994** |
| "Portugal" | **0.377** | **0.395** | **0.6056** | -0.01301 | 0.02034 | 0.03839 |
| "Romania" | **0.546** | **0.588** | **0.23891** | **0.01029** | **0.02438** | **0.40288** |
| "Spain" | 0.27 | 0.199 | 0 | **0.01477** | **0.01033** | **0.36117** |
| "Sweden" | **0.051** | **0.065** | **0.62018** | **0.00144** | **0.00321** | **0.92519** |
| "United Kingdom" | **0.127** | **0.102** | **0.18594** | **-0.00039** | **0.00549** | **0.66982** |

Percentage of countries with no significantly different ABR proportions over 2006-2019: 15/18 = **83%**
Percentage of countries with no significantly different ABR trends (2006-2019): 17/18 = **94%**

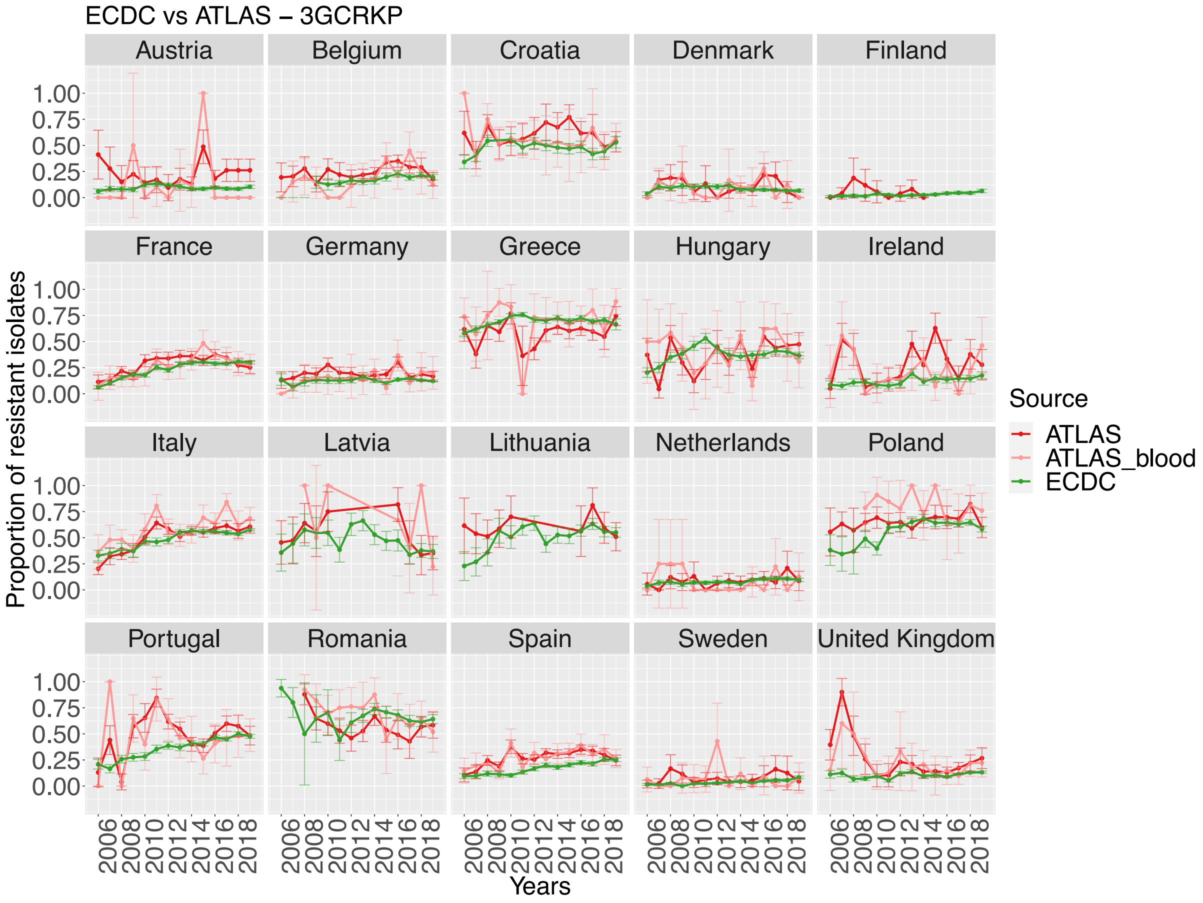

| "Country" | "ABR_prop_ATLAS" | "ABR_prop_ECDC" | "z_test" | "slope_ATLAS" | "slope_ECDC" | "anova_interaction" |
|---|---|---|---|---|---|---|
| "Austria" | **0.086** | **0.096** | **0.98646** | **-0.01293** | **0.00058** | **0.86112** |
| "Belgium" | **0.228** | **0.183** | **0.07827** | **0.00673** | **0.00867** | **0.81186** |
| "Croatia" | **0.534** | **0.479** | **0.054** | **0.00126** | **0.00194** | **0.95207** |
| "Denmark" | **0.111** | **0.083** | **0.16851** | **0.00547** | **-0.00116** | **0.62881** |
| "France" | **0.274** | **0.255** | **0.22977** | **0.01894** | **0.01824** | **0.94562** |
| "Germany" | **0.159** | **0.13** | **0.09317** | **0.00527** | **0.00088** | **0.55964** |
| "Greece" | **0.741** | **0.699** | **0.15915** | **0.00223** | **0.00413** | **0.90175** |
| "Hungary" | **0.429** | **0.383** | **0.31718** | **-0.00911** | **0.00551** | **0.5834** |
| "Ireland" | 0.228 | 0.127 | 1e-05 | **-0.00281** | **0.00667** | **0.45125** |
| "Italy" | 0.588 | 0.541 | 0.00455 | **0.02373** | **0.01322** | **0.296** |
| "Latvia" | **0.478** | **0.458** | **1** | **-0.03737** | **-0.01616** | **0.7436** |
| "Netherlands" | **0.109** | **0.085** | **0.63949** | **-0.00995** | **0.00453** | **0.46992** |
| "Poland" | 0.786 | 0.617 | 5e-05 | **-0.00932** | **0.00936** | **0.36133** |
| "Portugal" | **0.463** | **0.419** | **0.17651** | -0.02077 | 0.02286 | 0.0203 |
| "Romania" | **0.697** | **0.661** | **0.29031** | **-0.02637** | **-0.01664** | **0.39042** |
| "Spain" | 0.265 | 0.189 | 0 | **0.01435** | **0.01381** | **0.9131** |
| "Sweden" | 0.08 | 0.042 | 0.04969 | **0.0025** | **0.00408** | **0.84375** |
| "United Kingdom" | 0.19 | 0.112 | 4e-05 | **-0.00451** | **0.00466** | **0.45408** |

Percentage of countries with no significantly different ABR proportions over 2006-2019: 12/18 = **67%**
Percentage of countries with no significantly different ABR trends (2006-2019): 17/18 = **94%**

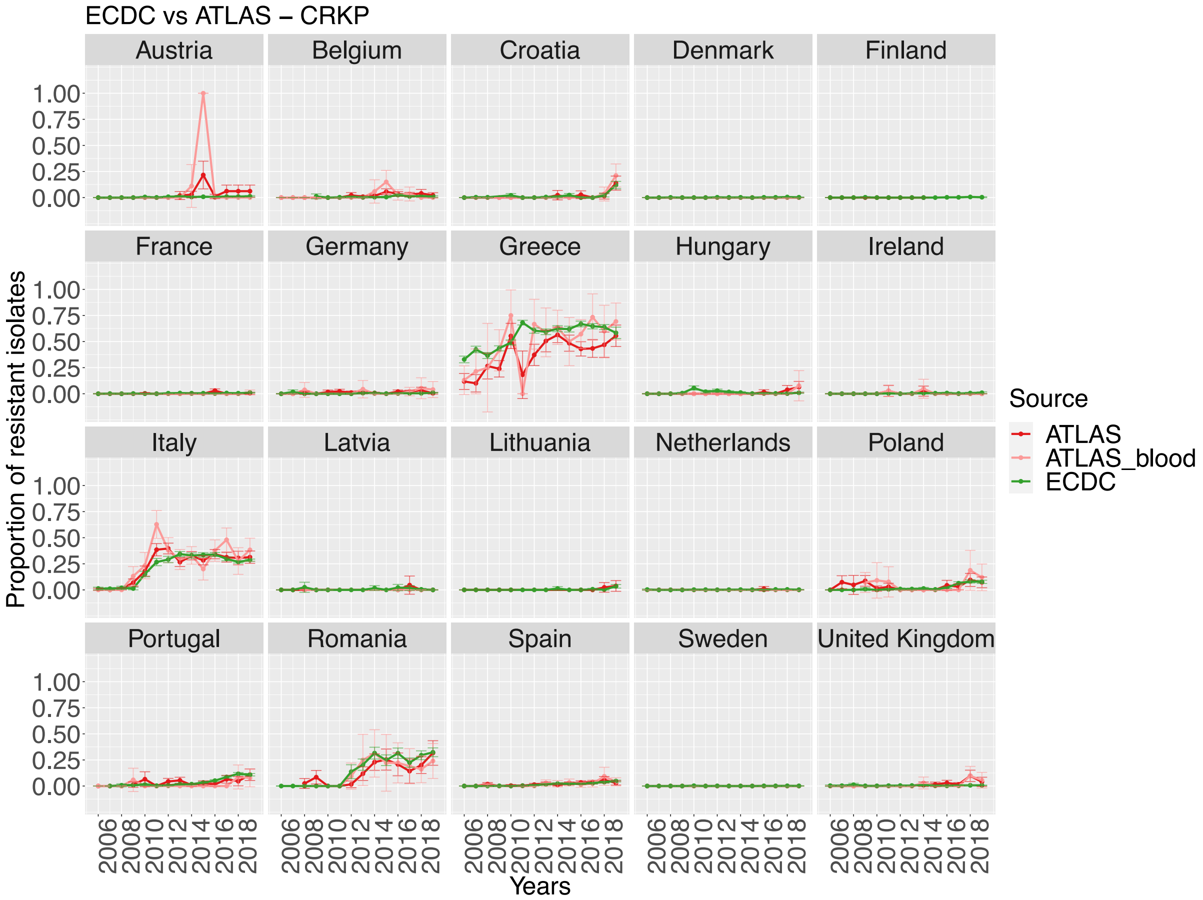

| "Country" | "ABR_prop_ATLAS" | "ABR_prop_ECDC" | "z_test" | "slope_ATLAS" | "slope_ECDC" | "anova_interaction" |
|---|---|---|---|---|---|---|
| "Austria" | **0.034** | **0.007** | **0.10871** | NA | 0.00096 | NA |
| "Belgium" | 0.034 | 0.01 | 0.00063 | **-0.02097** | **0.00125** | **0.37278** |
| "Croatia" | 0.034 | 0.017 | 0.0319 | **0.17706** | **0.00196** | **0.32882** |
| "Denmark" | **0** | **0.002** | **1** | NA | 0.00015 | NA |
| "France" | **0.001** | **0.005** | **0.24424** | NA | 0.00058 | NA |
| "Germany" | **0.012** | **0.005** | **0.12033** | **-0.00075** | **0.00076** | **0.81595** |
| "Greece" | **0.534** | **0.558** | **0.47074** | **0.03905** | **0.02404** | **0.55075** |
| "Hungary" | **0.007** | **0.011** | **0.99226** | NA | -0.00097 | NA |
| "Ireland" | **0.009** | **0.004** | **0.54643** | **0.00635** | **0.00029** | **0.66849** |
| "Italy" | **0.275** | **0.279** | **0.81776** | **0.01434** | **0.02503** | **0.76283** |
| "Latvia" | **0** | **0.006** | **1** | NA | -0.00261 | NA |
| "Netherlands" | **0** | **0.002** | **1** | NA | 4e-05 | NA |
| "Poland" | **0.069** | **0.042** | **0.15478** | **0.00679** | **0.00588** | **0.9703** |
| "Portugal" | 0.023 | 0.062 | 0.01509 | **0.00212** | **0.00992** | **0.78883** |
| "Romania" | 0.13 | 0.266 | 1e-05 | **0.00955** | **0.01639** | **0.71661** |
| "Spain" | **0.017** | **0.021** | **0.4588** | **0.00211** | **0.00402** | **0.26815** |
| "Sweden" | **0** | **0.001** | **1** | NA | -4e-05 | NA |
| "United Kingdom" | 0.023 | 0.005 | 2e-04 | **0.00826** | **0.00038** | **0.44699** |

Percentage of countries with no significantly different ABR proportions over 2006-2019: 13/18 = **72%**
Percentage of countries with no significantly different ABR trends (2006-2019): 11/11 = **100%**

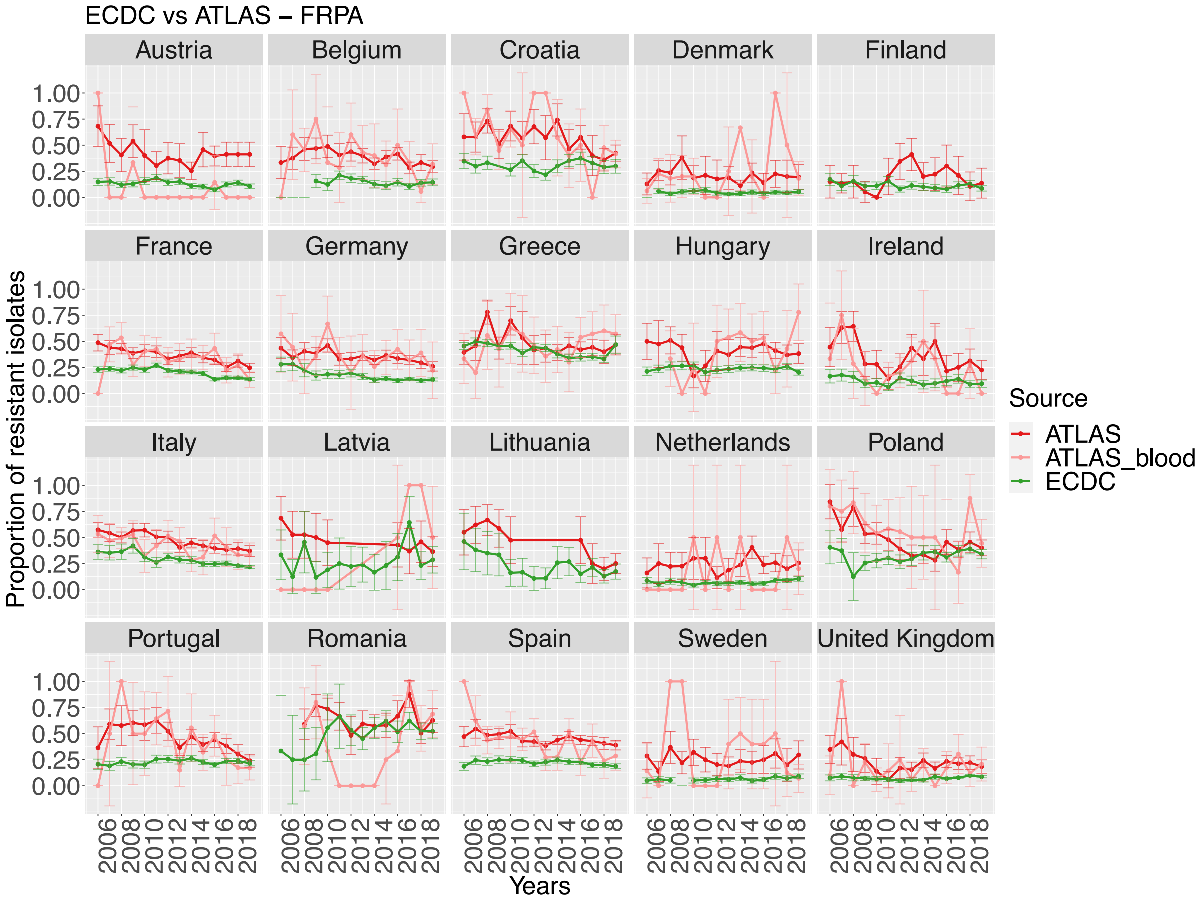

| "Country" | "ABR_prop_ATLAS" | "ABR_prop_ECDC" | "z_test" | "slope_ATLAS" | "slope_ECDC" | "anova_interaction" |
|---|---|---|---|---|---|---|
| "Austria" | **0.091** | **0.128** | **0.70694** | **-0.02721** | **-0.00365** | **0.78785** |
| "Belgium" | 0.366 | 0.149 | 0 | -0.03736 | -0.00558 | 0.0168 |
| "Croatia" | 0.541 | 0.309 | 0 | -0.02234 | 0.00055 | 0.03954 |
| "Denmark" | 0.189 | 0.048 | 0 | **0.00785** | **-0.00026** | **0.29712** |
| "France" | 0.327 | 0.196 | 0 | **-0.02254** | **-0.00965** | **0.1393** |
| "Germany" | 0.359 | 0.146 | 0 | **-0.00651** | **-0.00763** | **0.91378** |
| "Greece" | **0.48** | **0.417** | **0.08179** | 0.01818 | -0.01253 | 0.01123 |
| "Hungary" | 0.447 | 0.238 | 0 | **0.01565** | **-0.00093** | **0.37762** |
| "Ireland" | 0.229 | 0.116 | 0.00018 | **-0.00035** | **-0.00243** | **0.92349** |
| "Italy" | 0.424 | 0.252 | 0 | **-0.01402** | **-0.01163** | **0.65139** |
| "Latvia" | **0.385** | **0.261** | **0.50355** | **0** | **0.00899** | **0.95852** |
| "Netherlands" | 0.2 | 0.072 | 0.0192 | **-0.03795** | **0.00238** | **0.37212** |
| "Poland" | 0.577 | 0.333 | 0 | **-0.01683** | **0.0077** | **0.05648** |
| "Portugal" | 0.332 | 0.229 | 0.00105 | -0.03593 | 0.00079 | 0.02271 |
| "Romania" | **0.479** | **0.538** | **0.40335** | **-0.00019** | **0.01005** | **0.58026** |
| "Spain" | 0.406 | 0.222 | 0 | **-0.01916** | **-0.0031** | **0.05399** |
| "Sweden" | 0.208 | 0.067 | 0 | **-0.01109** | **0.00242** | **0.26519** |
| "United Kingdom" | 0.169 | 0.078 | 0 | **0.00788** | **0.00219** | **0.53679** |

Percentage of countries with no significantly different ABR proportions over 2006-2019: 4/18 = **22%**
Percentage of countries with no significantly different ABR trends (2006-2019): 14/18 = **78%**

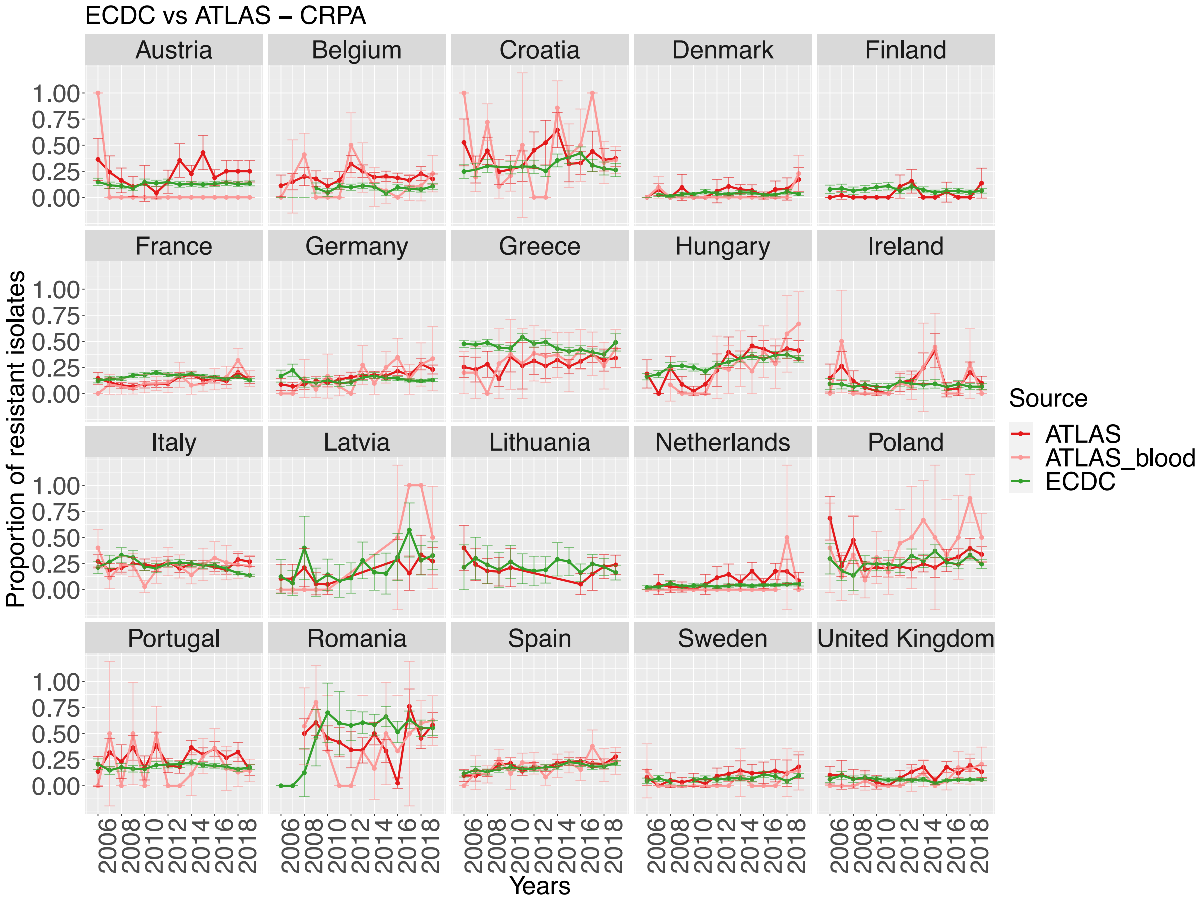

| "Country" | "ABR_prop_ATLAS" | "ABR_prop_ECDC" | "z_test" | "slope_ATLAS" | "slope_ECDC" | "anova_interaction" |
|---|---|---|---|---|---|---|
| "Austria" | **0.03** | **0.128** | **0.15637** | NA | 0.00129 | NA |
| "Belgium" | 0.186 | 0.091 | 9e-05 | **-0.02074** | **-0.00047** | **0.19469** |
| "Croatia" | **0.35** | **0.309** | **0.22849** | **0.0126** | **0.00446** | **0.56402** |
| "Denmark" | **0.056** | **0.034** | **0.24396** | **0.01087** | **0.00129** | **0.44384** |
| "France" | 0.125 | 0.161 | 0.01335 | **0.01019** | **-0.00067** | **0.1028** |
| "Germany" | **0.168** | **0.134** | **0.12869** | 0.02123 | -0.00062 | 0.04053 |
| "Greece" | 0.332 | 0.451 | 0.00093 | **0.01376** | **-0.00699** | **0.1234** |
| "Hungary" | **0.289** | **0.296** | **0.9665** | **0.03829** | **0.01481** | **0.176** |
| "Ireland" | **0.099** | **0.079** | **0.5077** | **0.01368** | **-0.00069** | **0.55483** |
| "Italy" | **0.218** | **0.193** | **0.12869** | 0.00717 | -0.01288 | 0.0263 |
| "Latvia" | **0.385** | **0.235** | **0.36714** | **0** | **0.02049** | **0.89681** |
| "Netherlands" | **0.033** | **0.041** | **1** | NA | 0.00182 | NA |
| "Poland" | 0.387 | 0.273 | 0.01099 | 0.04683 | 0.00375 | 0.00343 |
| "Portugal" | **0.202** | **0.187** | **0.66606** | **-0.03358** | **-1e-04** | **0.0628** |
| "Romania" | **0.507** | **0.574** | **0.32296** | **0.00328** | **0.00512** | **0.93188** |
| "Spain" | **0.202** | **0.182** | **0.21446** | **0.00686** | **0.00597** | **0.88344** |
| "Sweden" | **0.065** | **0.072** | **0.97627** | **-0.00026** | **0.00238** | **0.87786** |
| "United Kingdom" | **0.092** | **0.058** | **0.05866** | **0.01077** | **-0.00067** | **0.26536** |

Percentage of countries with no significantly different ABR proportions over 2006-2019: 14/18 = **78%**
Percentage of countries with no significantly different ABR trends (2006-2019): 13/16 = **81%**

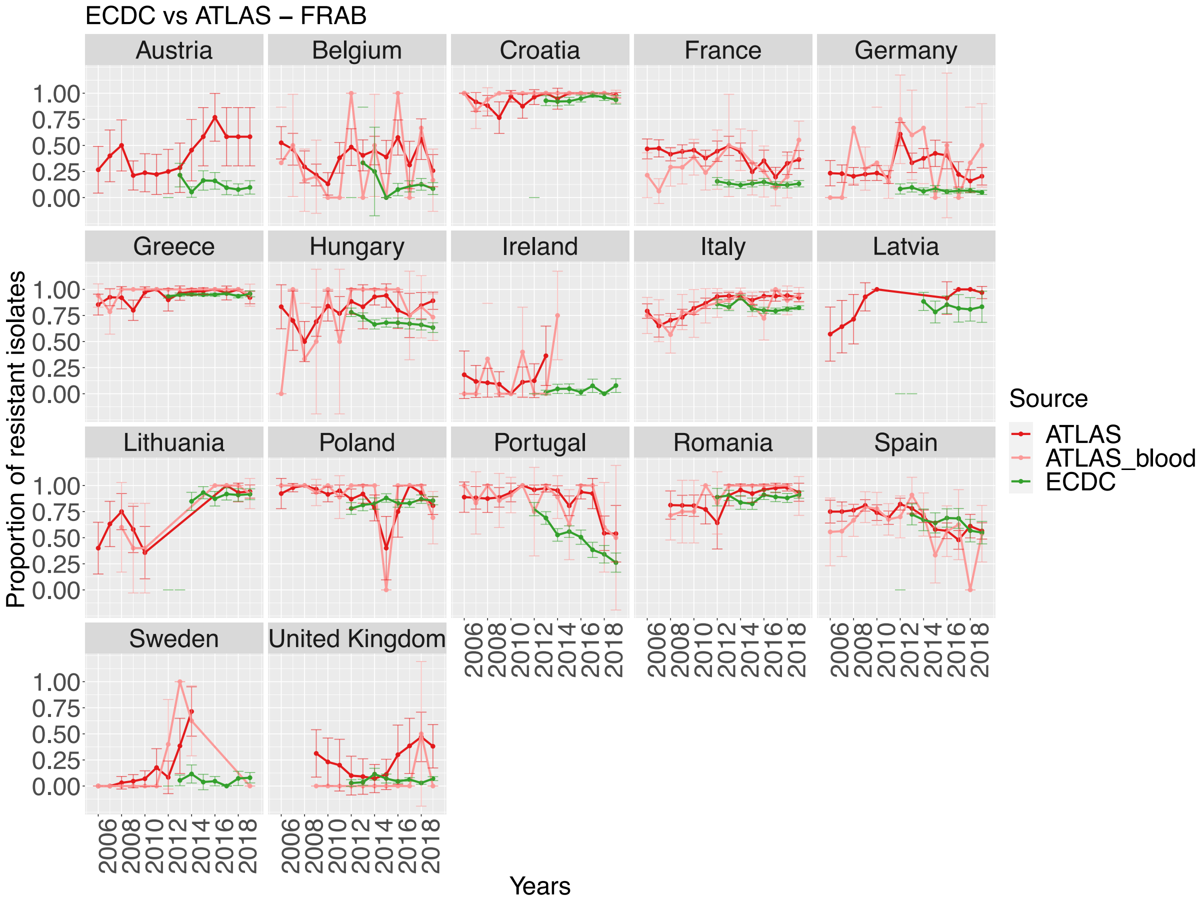

| "Country" | "ABR_prop_ATLAS" | "ABR_prop_ECDC" | "z_test" | "slope_ATLAS" | "slope_ECDC" | "anova_interaction" |
|---|---|---|---|---|---|---|
| "Belgium" | 0.295 | 0.1 | 0.00032 | **0.00206** | **-0.00016** | **0.90603** |
| "Croatia" | **0.968** | **0.944** | **0.30175** | **0.00551** | **0.00806** | **0.77525** |
| "France" | 0.316 | 0.134 | 0 | **0.01354** | **-0.00236** | **0.0817** |
| "Germany" | 0.286 | 0.067 | 0 | **0.02363** | **-0.00433** | **0.18918** |
| "Greece" | **0.97** | **0.946** | **0.25166** | **0.0025** | **0.00176** | **0.93025** |
| "Hungary" | **0.769** | **0.689** | **0.27749** | 0.03493 | -0.01732 | 0.02765 |
| "Ireland" | 0.231 | 0.039 | 6e-05 | **0.07355** | **0.0079** | **0.33366** |
| "Italy" | **0.842** | **0.823** | **0.37536** | 0.01735 | -0.00927 | 0.02116 |
| "Lithuania" | 0.757 | 0.902 | 0.01377 | 0.04435 | 0.00542 | 0.02973 |
| "Poland" | **0.916** | **0.838** | **0.07994** | **-0.0241** | **0.00771** | **0.08606** |
| "Portugal" | 0.851 | 0.535 | 0 | -0.0216 | -0.06909 | 0.01901 |
| "Romania" | **0.875** | **0.883** | **0.97389** | **0.01608** | **0.00455** | **0.40552** |
| "Spain" | **0.665** | **0.648** | **0.68427** | **-0.00873** | **-0.02357** | **0.37087** |
| "Sweden" | 0.229 | 0.061 | 0.00082 | **0.1125** | **0.00291** | **0.39117** |
| "United Kingdom" | **0.03** | **0.054** | **0.83378** | NA | 0.0011 | NA |

Percentage of countries with no significantly different ABR proportions over 2006-2019: 8/15 = **53%**
Percentage of countries with no significantly different ABR trends (2006-2019): 10/14 = **71%**

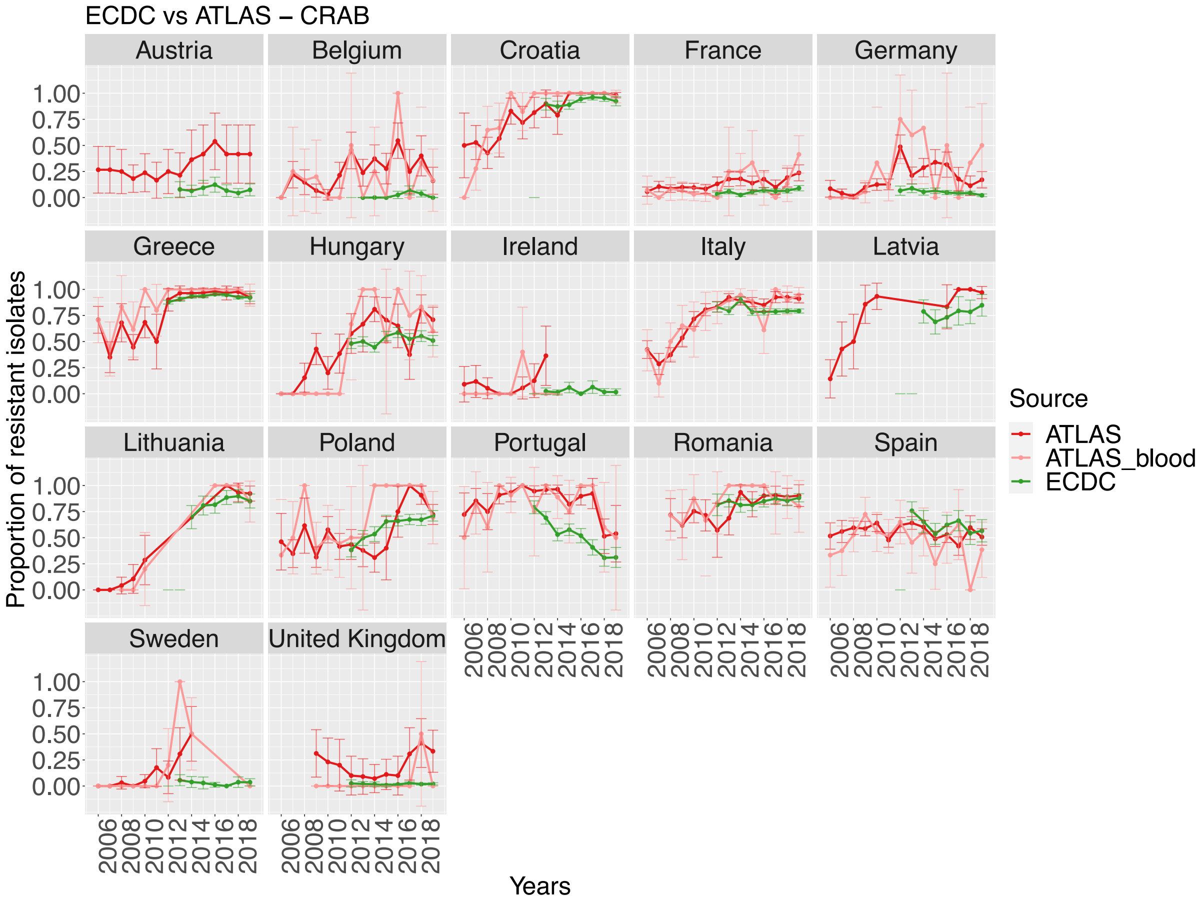

| "Country" | "ABR_prop_ATLAS" | "ABR_prop_ECDC" | "z_test" | "slope_ATLAS" | "slope_ECDC" | "anova_interaction" |
|---|---|---|---|---|---|---|
| "Belgium" | 0.182 | 0.034 | 5e-05 | **0.00054** | **0.00548** | **0.74478** |
| "Croatia" | 0.813 | 0.922 | 2e-05 | **0.03792** | **0.01245** | **0.05623** |
| "France" | 0.125 | 0.059 | 5e-05 | **0.01797** | **0.00704** | **0.18542** |
| "Germany" | 0.202 | 0.048 | 0 | 0.05485 | -0.00792 | 0.00354 |
| "Greece" | 0.867 | 0.923 | 0.011 | **0.02493** | **0.00765** | **0.24209** |
| "Hungary" | **0.577** | **0.517** | **0.47698** | **-0.00073** | **0.00869** | **0.88161** |
| "Ireland" | **0.077** | **0.028** | **0.40761** | NA | -0.00021 | NA |
| "Italy" | **0.761** | **0.8** | **0.07616** | 0.04604 | -0.00868 | 0.00084 |
| "Lithuania" | 0.568 | 0.833 | 0.00015 | **0.07179** | **0.02611** | **0.19152** |
| "Poland" | **0.614** | **0.618** | **1** | **0.02339** | **0.04057** | **0.36113** |
| "Portugal" | 0.824 | 0.545 | 0 | -0.00472 | -0.06882 | 0.00624 |
| "Romania" | **0.825** | **0.851** | **0.64169** | **0.01033** | **0.00851** | **0.82097** |
| "Spain" | 0.52 | 0.622 | 0.00629 | **-0.00737** | **-0.02595** | **0.2736** |
| "Sweden" | 0.171 | 0.03 | 0.00026 | **0.15** | **-0.00067** | **0.25849** |
| "United Kingdom" | **0.03** | **0.02** | **1** | NA | 0.001 | NA |

Percentage of countries with no significantly different ABR proportions over 2006-2019: 6/15 = **40%**
Percentage of countries with no significantly different ABR trends (2006-2019): 10/13 = **77%**

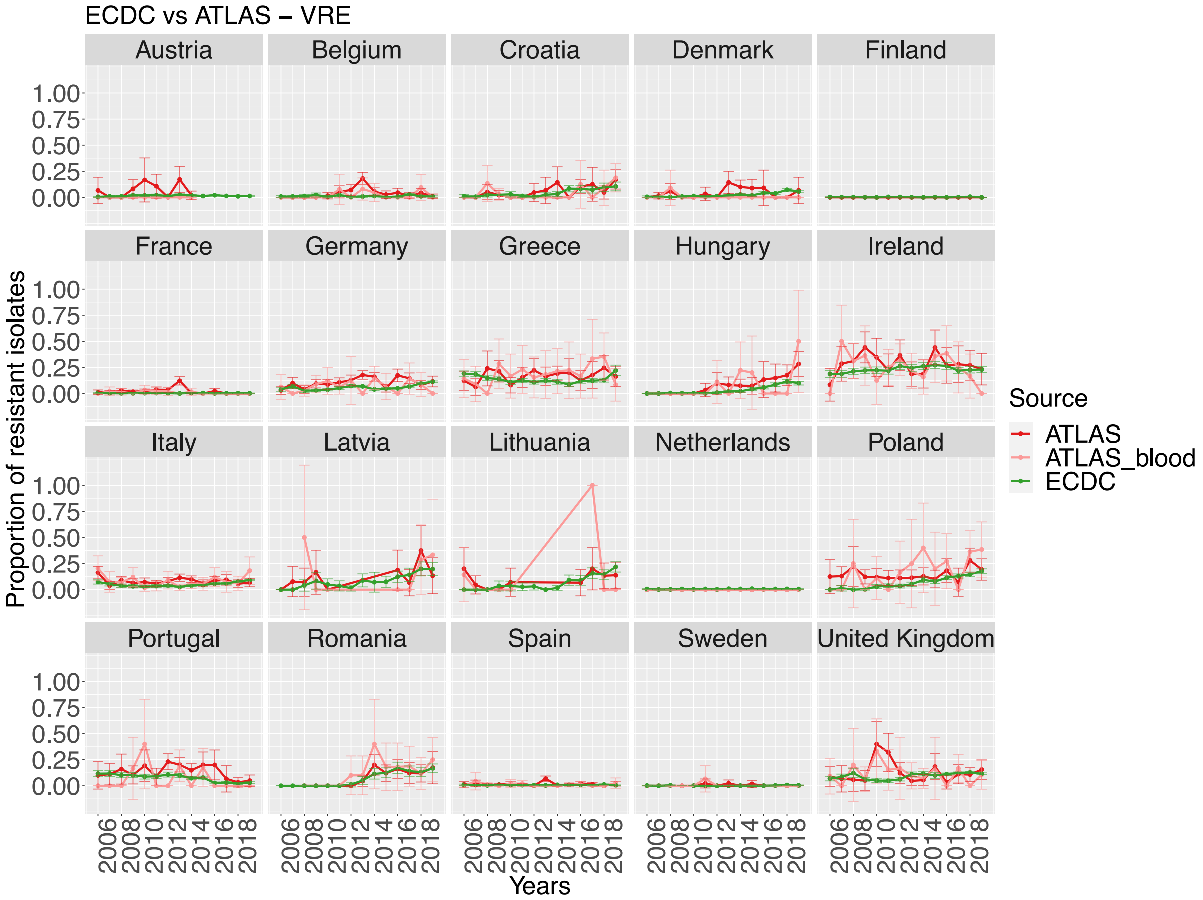

| "Country" | "ABR_prop_ATLAS" | "ABR_prop_ECDC" | "z_test" | "slope_ATLAS" | "slope_ECDC" | "anova_interaction" |
|---|---|---|---|---|---|---|
| "Austria" | **0** | **0.015** | **1** | NA | 0.00036 | NA |
| "Belgium" | **0.028** | **0.012** | **0.07983** | **0.00298** | **3e-05** | **0.87608** |
| "Croatia" | **0.045** | **0.048** | **0.97219** | **0.01133** | **0.00482** | **0.59444** |
| "Denmark" | **0.007** | **0.028** | **0.24253** | NA | 0.00366 | NA |
| "France" | 0.011 | 0.003 | 0.00043 | **1e-05** | **2e-05** | **0.99924** |
| "Germany" | **0.074** | **0.071** | **0.95011** | **0.00696** | **0.00507** | **0.90037** |
| "Greece" | 0.196 | 0.133 | 0.02385 | **0.00292** | **-0.00357** | **0.61579** |
| "Hungary" | **0.08** | **0.045** | **0.24767** | **0.05122** | **0.00758** | **0.71568** |
| "Ireland" | **0.273** | **0.232** | **0.18164** | **-0.00983** | **0.00345** | **0.2776** |
| "Italy" | **0.075** | **0.065** | **0.27818** | **0.00094** | **0.00481** | **0.55967** |
| "Latvia" | **0.16** | **0.105** | **0.57301** | **-0.01865** | **0.01562** | **0.50633** |
| "Lithuania" | **0.073** | **0.099** | **0.78003** | NA | 0.01674 | NA |
| "Netherlands" | **0** | **0.006** | **1** | NA | 0.00033 | NA |
| "Poland" | 0.167 | 0.106 | 0.03057 | **0.02497** | **0.01524** | **0.54222** |
| "Portugal" | **0.077** | **0.065** | **0.6788** | **-0.0091** | **-0.0087** | **0.99434** |
| "Romania" | **0.117** | **0.118** | **1** | **0.0106** | **0.02233** | **0.51408** |
| "Spain" | **0.009** | **0.008** | **0.86561** | **0.00029** | **1e-04** | **0.90091** |
| "Sweden" | **0.008** | **0.002** | **0.64458** | NA | 1e-04 | NA |
| "United Kingdom" | **0.103** | **0.111** | **0.80971** | **0.00207** | **0.006** | **0.74428** |

Percentage of countries with no significantly different ABR proportions over 2006-2019: 16/19 = **84%**
Percentage of countries with no significantly different ABR trends (2006-2019): 14/14 = **100%**

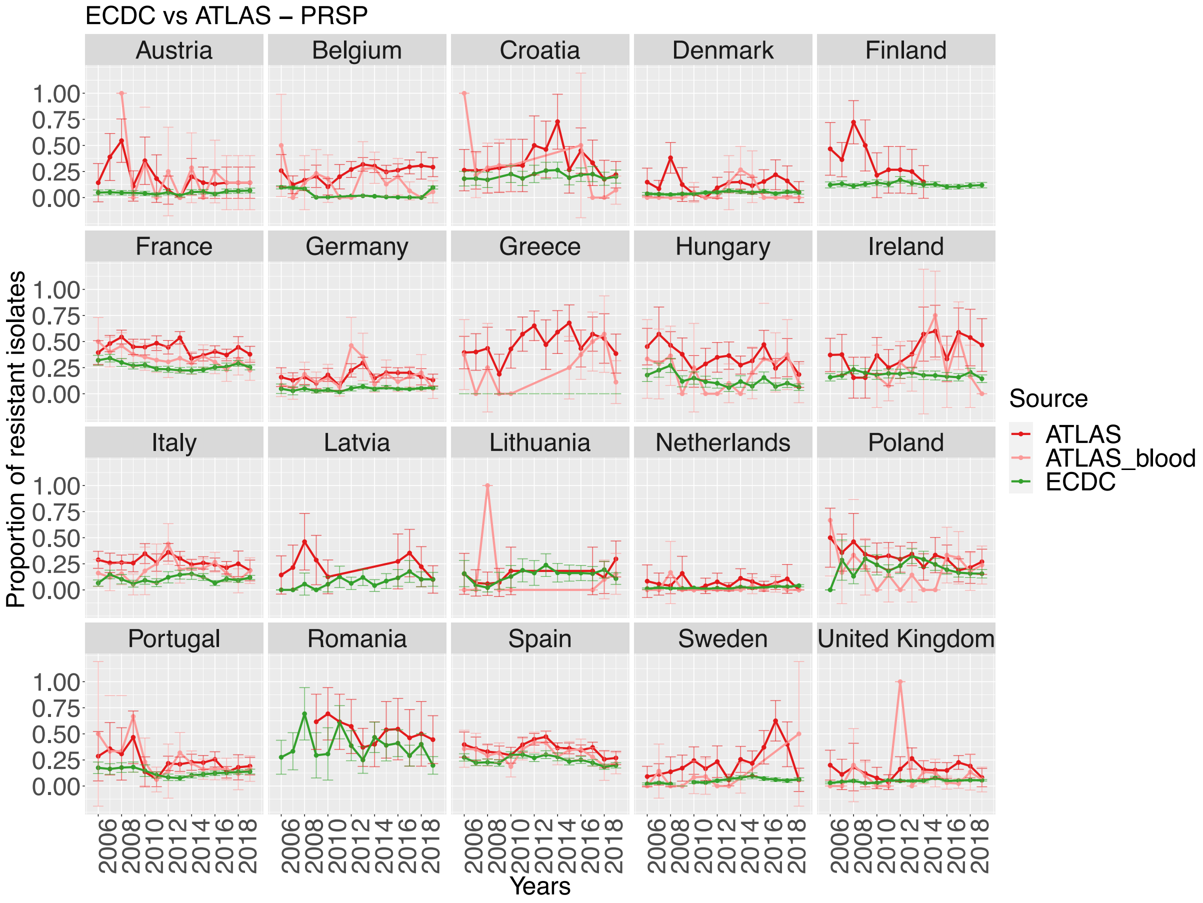

| "Country" | "ABR_prop_ATLAS" | "ABR_prop_ECDC" | "z_test" | "slope_ATLAS" | "slope_ECDC" | "anova_interaction" |
|---|---|---|---|---|---|---|
| "Austria" | 0.183 | 0.048 | 1e-05 | **-0.02311** | **0.0012** | **0.41165** |
| "Belgium" | 0.147 | 0.032 | 0 | **-0.02015** | **-9e-04** | **0.65969** |
| "Croatia" | **0.237** | **0.208** | **0.64518** | -0.01712 | 0.00187 | 0.01526 |
| "Denmark" | **0.065** | **0.045** | **0.39011** | **0.03892** | **0.00182** | **0.13436** |
| "France" | 0.333 | 0.26 | 1e-05 | **-0.01494** | **-0.00301** | **0.10108** |
| "Germany" | 0.146 | 0.049 | 0 | **0.00116** | **0.00231** | **0.82069** |
| "Hungary" | 0.196 | 0.124 | 0.04852 | **-0.01089** | **-0.00916** | **0.90789** |
| "Ireland" | **0.256** | **0.183** | **0.10563** | **0.03182** | **-0.00154** | **0.136** |
| "Italy" | 0.2 | 0.105 | 0 | **0.0043** | **0.00145** | **0.65512** |
| "Lithuania" | **0.133** | **0.143** | **1** | **0.05385** | **0.0097** | **0.8249** |
| "Netherlands" | **0.028** | **0.022** | **0.87211** | **-0.00659** | **0.00155** | **0.74723** |
| "Poland" | **0.198** | **0.202** | **1** | **-0.00084** | **-0.00668** | **0.68136** |
| "Portugal" | 0.186 | 0.125 | 0.00164 | **-0.00437** | **-0.00013** | **0.72424** |
| "Spain" | 0.318 | 0.245 | 0 | **-0.00457** | **-0.00415** | **0.95717** |
| "Sweden" | **0.072** | **0.049** | **0.52196** | **0.00216** | **0.00413** | **0.94533** |
| "United Kingdom" | **0.078** | **0.05** | **0.07472** | **-0.01053** | **0.00187** | **0.24979** |

Percentage of countries with no significantly different ABR proportions over 2006-2019: 8/16 = **50%**
Percentage of countries with no significantly different ABR trends (2006-2019): 15/16 = **94%**

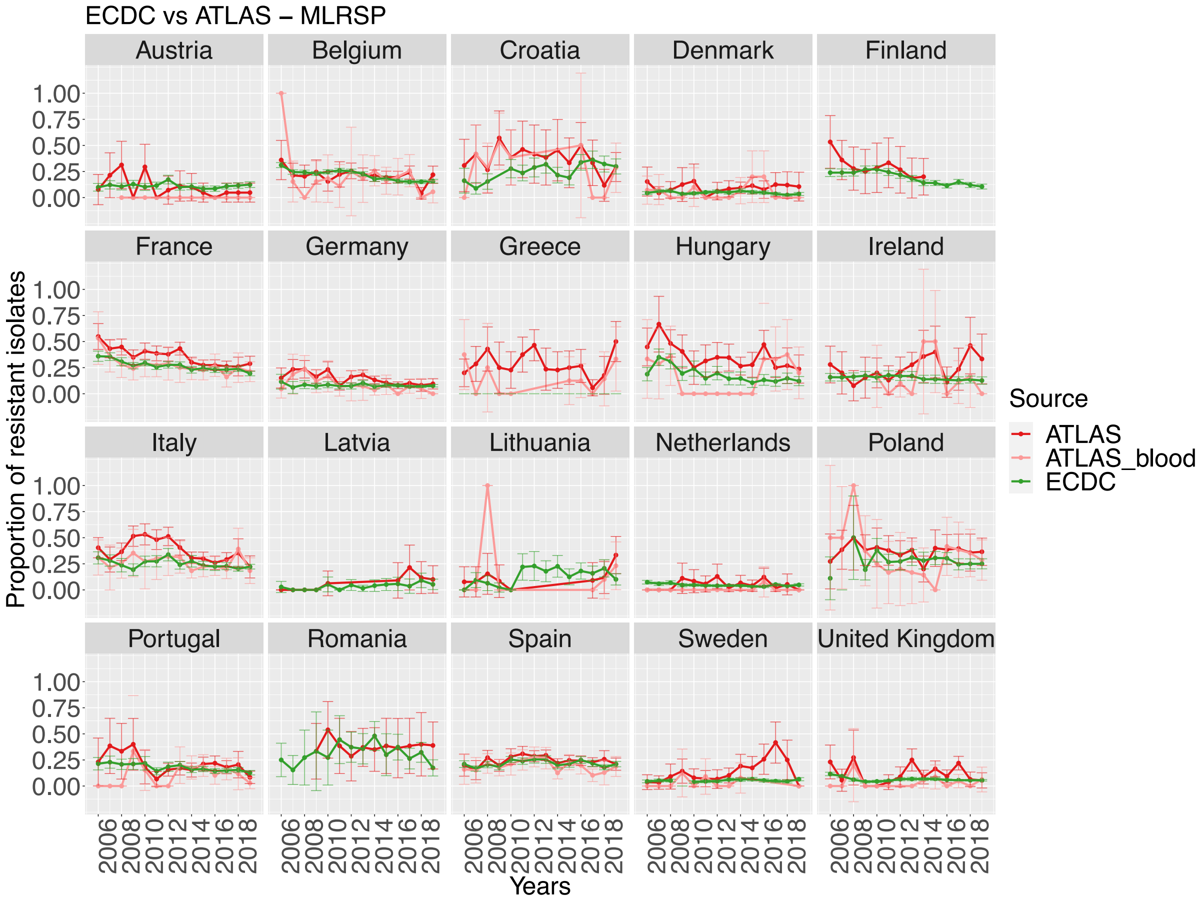

| "Country" | "ABR_prop_ATLAS" | "ABR_prop_ECDC" | "z_test" | "slope_ATLAS" | "slope_ECDC" | "anova_interaction" |
|---|---|---|---|---|---|---|
| "Austria" | 0 | 0.111 | 0.0123 | NA | -0.00036 | NA |
| "Belgium" | **0.185** | **0.217** | **0.29848** | **-0.00572** | **-0.01067** | **0.68355** |
| "Croatia" | **0.351** | **0.262** | **0.1195** | **-0.00822** | **0.01667** | **0.17202** |
| "Denmark" | **0.072** | **0.048** | **0.33608** | **0.00595** | **-0.00136** | **0.57192** |
| "France" | **0.257** | **0.26** | **0.89646** | **-0.01021** | **-0.00947** | **0.87932** |
| "Germany" | **0.085** | **0.076** | **0.5444** | **-0.0052** | **-0.00094** | **0.25013** |
| "Hungary" | **0.168** | **0.177** | **0.92786** | **-0.00491** | **-0.01127** | **0.67408** |
| "Ireland" | **0.092** | **0.151** | **0.16609** | **0.00506** | **-0.00312** | **0.52995** |
| "Italy" | **0.239** | **0.238** | **0.99348** | **-0.00696** | **-0.00544** | **0.79721** |
| "Lithuania" | **0.167** | **0.139** | **0.87168** | **0.13077** | **0.00874** | **0.63386** |
| "Netherlands" | 0.006 | 0.045 | 0.0356 | NA | -0.00143 | NA |
| "Poland" | **0.306** | **0.274** | **0.52281** | **0.00317** | **-0.00218** | **0.66464** |
| "Portugal" | **0.133** | **0.162** | **0.19289** | -0.02476 | -0.0062 | 0.02199 |
| "Spain" | **0.194** | **0.215** | **0.13231** | **-0.00511** | **0.00058** | **0.34172** |
| "Sweden" | **0.047** | **0.052** | **1** | **-0.00827** | **0.00087** | **0.7564** |
| "United Kingdom" | **0.037** | **0.062** | **0.17405** | **-0.00514** | **-0.0012** | **0.88028** |

Percentage of countries with no significantly different ABR proportions over 2006-2019: 14/16 = **87%**

Percentage of countries with no significantly different ABR trends (2006-2019): 13/14 = **93%**

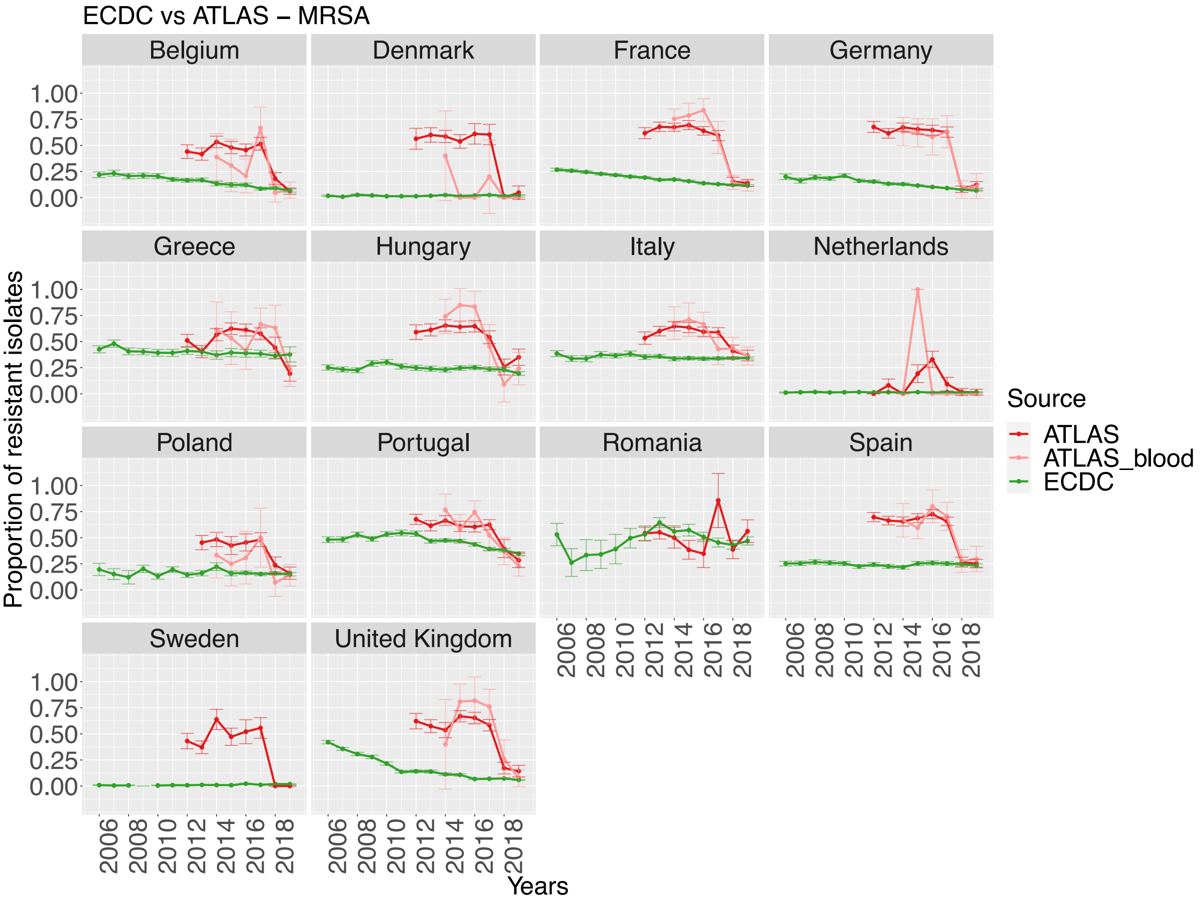

ECDC vs ATLAS – MRSA
Belgium
Denmark
France
Germany
Greece
Hungary
Italy
Netherlands
Poland
Portugal
Romania
Spain
Sweden
United Kingdom
1.00
0.75
0.50
0.25
0.00
2006
2008
2010
2012
2014
2016
2018
Proportion of resistant isolates
Years
Source
ATLAS
ATLAS_blood
ECDC
